# Critically-ill COVID-19 susceptibility gene *CCR3* shows natural selection in sub-Saharan Africans

**DOI:** 10.1101/2024.01.12.24301202

**Authors:** Zewen Sun, Lin Pan, Aowen Tian, Peng Chen

## Abstract

The prevalence of COVID-19 critical illness varies across ethnicities, with recent studies suggesting that genetic factors may contribute to this variation. The aim of this study was to investigate natural selection signals of genes associated with critically-ill COVID-19 in sub-Saharan Africans. Severe COVID-19 SNPs were obtained from the HGI website. Selection signals were assessed in 661 sub-Sahara Africans from 1000 Genomes Project using integrated haplotype score (iHS), cross-population extended haplotype homozygosity (xpEHH), and fixation index (Fst). Allele frequency trajectory analysis of ancient DNA samples were used to validate the existing of selection in sub-Sahara Africans. We also used Mendelian randomization to decipher the correlation between natural selection and critically-ill COVID-19. We identified that *CCR3* exhibited significant natural selection signals in sub-Sahara Africans. Within the *CCR3* gene, rs17217831-A showed both high iHS (Standardized iHS = 2) and high XP-EHH (Standardized XP-EHH = 2.5) in sub-Sahara Africans. Allele frequency trajectory of *CCR3* rs17217831-A revealed natural selection occurring in the recent 1,500 years. Natural selection resulted in increased *CCR3* expression in sub-Sahara Africans. Mendelian Randomization provided evidence that increased blood *CCR3* expression and eosinophil counts lowered the risk of critically ill COVID-19. Our findings suggest that sub-Saharan Africans are less vulnerable to critically ill COVID-19 due to natural selection and identify *CCR3* as a potential novel therapeutic target.

## Introduction

The pandemic of COVID-19 has caused unprecedented and devastating influence around the world^1^. Most patients are asymptomatic or present with mild disease, while others proceed to severe hypoxic pneumonia or other diseases, which could ultimately lead to death ^2^.

African ancestry is considered to be associated with the outcome of COVID-19, but the conclusion has not been made. The risk of hospitalization or in-hospital mortality of people with African ancestry is usually confounded by socioeconomic status^3^. After carefully controlling of these factors, African people manifested equal^4^ or even lower risk of critical illness^5,6^. At the same time, the vaccination rate and death rate in Africa are both the lowest.

Studies have shown that several genetic variants associated with severe COVID-19 were apparently unique in African population. Huffman et al. found that *OAS1* rs10774671 G, which protects against hospitalization for COVID-19, had a higher allele frequency (58%) in Africans than in European population (32%)^7^. It has also been suggested that severe COVID-19 genes were expressed differentially in monocytes between Europeans and Africans ^8^. And an average of 69% of COVID severity-associated genes across conditions in monocytes showed increased expression with greater European ancestry. Additionally, higher proportion of European ancestry has been linked to a more robust type I IFN (Interferon) response linked to lower viral titers at later stages, which have significant clinical implications for SARS-CoV-2 infection. All these evidence gives birth to the hypothesis that African people may be genetically invulnerable to critically-ill COVID-19.

Pathogenic viruses are among the strongest sources of selection pressure in human evolution^9^. Previous selection analysis shows several genes related to the susceptibility of SARS-CoV-2 infection, including *ACE2*, *TMPRSS2*, *DPP4* and *LY6E* are under natural selection in Africans^10^. However, whether natural selection involved in the allele frequency difference or expression regulation of critically-ill COVID-19 genes remains unknown. In this study, we investigate the recent natural selection of the risk alleles associated with critically-ill COVID-19 in sub-Saharan African aiming to explain the population-specific risk of critically-ill COVID-19. Our results could inform the development of novel therapeutic targets for critically-ill COVID-19.

## Materials and Methods

### SNPs associated with Critically-ill COVID-19

We obtained COVID-19 genome-wide association results from HGI website (http://www.covid19hg.org/results/). The round 7 summary statistics without 23andme samples was used in this study^11^. At the significance level of p<1×10^-5^, 8,305 SNPs were associated with critically-ill COVID-19. The details of GWAS meta-analysis of COVID-19 can be found in HGI website.

### Population differentiation, Natural selection analysis

Natural selection signals in the critical-ill SNPs were tested in the genotype data from the 1000 Genomes Project. In total, we selected 5 superpopulations, including Europeans (N=503), sub-Sahara Africans (N=661), East Asians (N=504), Ad Mixed American (N=347) and South Asian (N=489). The integrated haplotype Score (iHS) is a measure of the amount of extended haplotype homozygosity (EHH) at a given SNP and was used as the screening statistic for natural selection^12^. We tested the selection signals at genome-wide level in each superpopulation. The minor allele frequency threshold was set to 5%. SNPs with standardized iHS |exceeded 99% quantile (2.25) or under 1% (−2.73) of genome-wide standardized iHS was recognized as significant selection signals. In addition, cross population extended haplotype homozygosity (XP-EHH) was used as a secondary signal to reinforce the evidence of natural selection. We selected European as the reference population when calculating XP-EHH, as most studies on eQTL and critically-ill COVID-19 are based on European ancestry. Hapbin^13^ was used to calculate standardized iHS and XP-EHH. WRIGHT’s Fst statists was used to estimate the degree of differentiation at single SNP level within five superpopulations^14^ using PLINK v1.9. Vcftools^15^ was used to perform region Fst within five superpopulations.

### SNP allele frequency in ancient Human genomes

The genotype data of present and ancient DNA obtained from Allen Ancient DNA Resource (AADR) 1240K dataset (https://reich.hms.harvard.edu/allenancient-dna-resource-aadr-downloadable-genotypes-present-day-and-ancient-dnadata. Accessed 1 January 2022.) was downloaded and treated by EIGENSOFT. PLINK v1.9^16^ were used to exclude duplicated samples, high correlated samples (correlation cutoff 0.5) and samples with high missing geneotype rate (>5%). After quality control, 4741 samples with rs17217831 genotyping were used for allele frequency analysis (Supplementary table5). Archaeological ages were indicated in years before 1950 CE.

### Infer ancient allele frequency from the genotype data of 1000 Genome Project

Since the allele frequency calculated from ancient DNA can be inflated because of limited sample size in certain ages, we applied CLUES^17^, an importance sampling approach for approximating the full likelihood function for the selection coefficient, to the genotype data of African to infer the allele frequency trajectory. To validate selection at SNP of interest, CLUES relies on Markov Chain Monte Carlo (MCMC) samples of the gene tree at the SNP. Genotype data is processed as follow steps: (1) We converted the vcf format genotype data into Relate’s required haps/sample input file types. SNPs that are not biallelic were removed. Relate v1.1^18^ was used to get a tree with genome-wide genealogies. (2) We also estimated the population size, the branch length and the average mutation rate of the African genome. Given the estimated coalescence rate, the MCMC method was used to sample branch lengths. CLUES was used to perform inference and visualize results (plot trajectories). Time bins in the frequency trajectory were set in 4 different epochs: 0-1500, 1500-3000 and 3000-4500 years.

### Association analysis of critically-ill COVID-19 SNPs in Africans

The UK Biobank is consisted of data from over 500,000 individuals aged 37–73 years (99.5% were between 40 and 69 years of age) who were recruited between 2006 and 2010 from across the UK^19^. 3,460 participants reported their ethnic background as African (AFR). We recoded the critically-ill COVID-19 in UK biobank African population. Critically-ill COVID-19 was determined by COVID-related death, Intensive care unit (ICU) admission or ventilation machine use 7 days before and up to 30 days after a patient’s first positive test for SARS-CoV-2 and Diagnoses of ICD10. The association between critically-ill COVID-19 and natural selected genes was calculated using PLINK. Age, BMI, sex, the first 5 genetic principal components were used as covariates.

### Colocalization, mendelian Randomization Analysis and instrumental variable selection

We further estimated the causal association between *CCR3* blood expression level, eosinophil counts and critically-ill COVID-19. We first performed colocalization test using eQTL data from Genotype-Tissue Expression (GTEx, https://gtexportal.org/) database^20^ and HGI critically-ill COVID-19 GWAS result with R package ‘coloc 2.0’^21^. We selected 575 significant eQTL (p < 1e-5) in the colocalization test. To further discover the causal role of *CCR3* and critically-ill COVID-19, two-sample Mendelian randomization analysis was conducted using R package ‘MR-PRESSO’ ^22^which was used to correct for the pleiotropy effect. The weighted median estimator and MR-egger were used as robust tests against MR-PRESSO using the R package ‘TwoSampleMR 0.5.6’^23^. Another causal relationship between eosinophil counts and critically-ill COVID-19 was tested using previous reported eosinophil count GWAS summary^24^ and HGI critically-ill COVID-19 GWAS result. After clumping, 1,193 SNPs were selected as independent instrumental variables. Two-sample Mendelian randomization analysis was performed using same methods. Mendelian randomization power was calculated mRnd^25^.

## Results

### SNPs at *CCR3* showed recent natural selection evidence

We investigated the natural selection in Sub-Sahara African population at 8,305 critically-ill COVID-19 SNPs, which were reported by recent Host Genetics Initiative (HGI) GWAS, as of date April 2022 (version 7). We evaluated the genome-wide integrated haplotype score (iHS) in Sub-Sahara Africans.

The most significant selection signal was rs13086063, located in upstream of *CCR3*, with its standardized iHS = 6.31 exceeding 99.97% quantile of whole genome standardized iHS (Fig.1 A, Table 1). The natural selection signal on *CCR3* was also validated by XP-EHH. The top standardized XP-EHH of *CCR3* SNP rs3091311 was 2.57, which is greater than the 99% quantile of whole genome standardized XP-EHH, 2.04 (Fig.1 B, Table 1). We also identified a *CCR3* SNP rs17217831 showed natural selection validated by two index (standardized iHS = 2.49 & standardized XP-EHH = 2.00).

**Figure 1.**
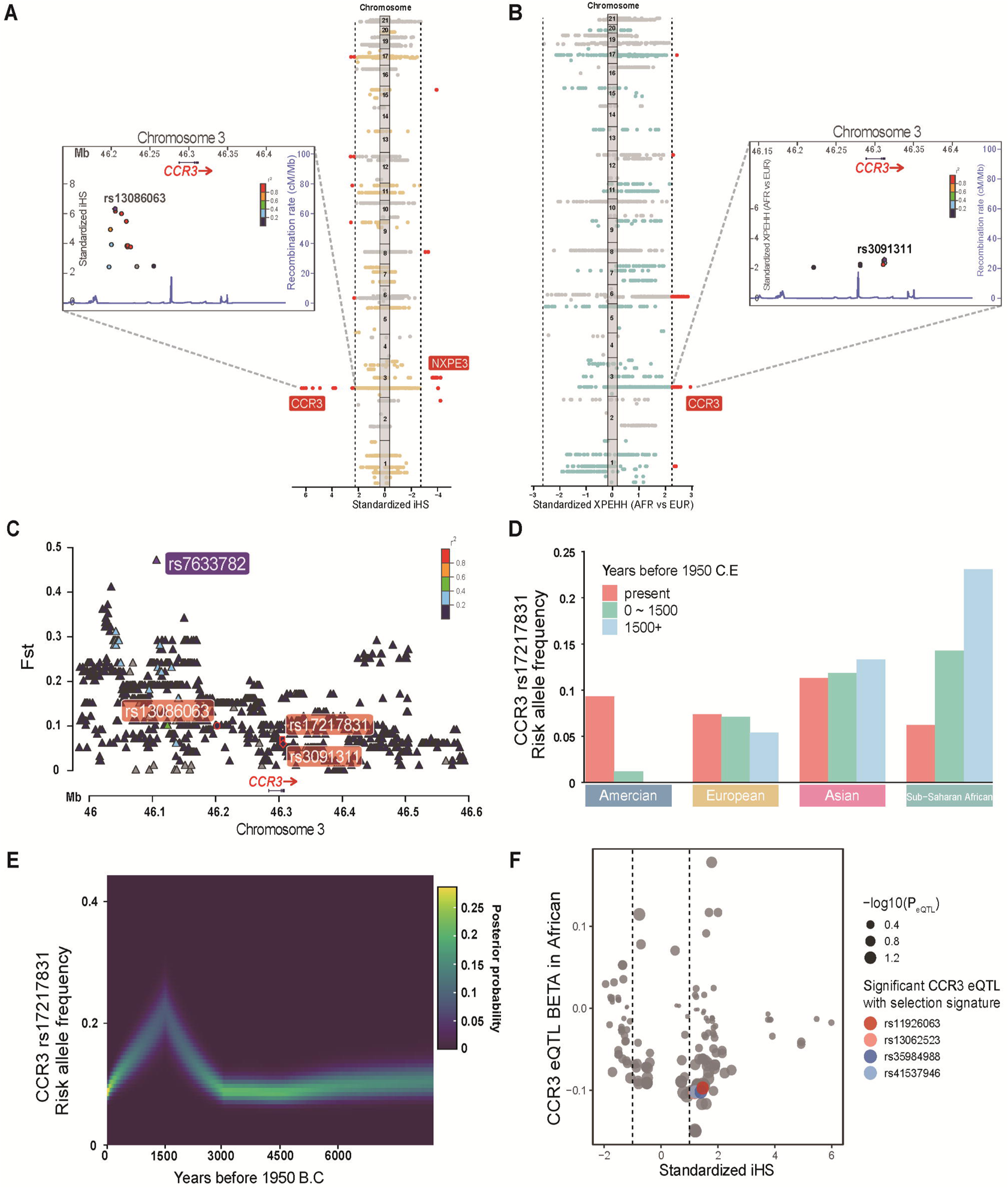
Natural selection signals of critically-ill COVID-19 SNPs. (A&B). Manhattan plot of critically-ill COVID-19 SNPs standardized iHS and standardized XP-EHH calculated between African and European. The x-axis is the chromosomal position and y-axis is the significance of nature selection. Red lines indicate genome-wide significance level (standardized iHS |& standardized XP-EHH > 99% quantile statics). Red points indicate the significant variants. (C). Locus plot for plotting Fst of variants in CCR. Linkage disequilibrium information is from 1000 Genomes African. (D). Frequency trajectory of CCR3 rs17217831 from ancient to present. (E). Frequency trajectory of CCR3 rs17217831 inferred from 1000 Genomes African genotype. The x-axis is generation before present. Yellow linear shows high probability of such frequency trajectory happened. (F) BETA, effect size of eQTL.

**Table 1.** Natural selection signatures for *CCR3* SNPs

| GENE | SNP | CHR:POS | Ancestry | Derived | AFR<br>standardized<br>XPEHH | AFR<br>standardized<br>iHS | EUR<br>standardized<br>iHS | Fst | AFR<br>Freq |
| --- | --- | --- | --- | --- | --- | --- | --- | --- | --- |
| *CCR3* | rs13086063 | 3:46200860 | T | C | 0.81 | 6.31 | 0.41 | 0.10 | 0.22 |
| *CCR3* | rs3091311 | 3:46306476 | T | C | 2.57 | 1.52 | 0.11 | 0.06 | 0.38 |
| *CCR3* | rs17217831 | 3:46305441 | C | A | 2.49 | 2.00 | 0.55 | 0.07 | 0.06 |

Ancestry, ancestry allele; Derived, derived allele; standardized XPEHH, standardized XP-EHH value
calculated in African population; AFR standardized iHS, iHS calculated in African population; EUR
standardized iHS, iHS calculated in European population; Fst, measure of population differentiation in

*CCR3* showed population differentiation as evidenced both by single SNP Fst (Fig 1.C, Supplementary table 1) and regional Fst (weighted Fst = 0.18 for CCR3 upstream chr3: 45900001-46200000 and weighted Fst = 0.10 for CCR3 chr3: 46200001-46500000) within five superpopulations in 1000 genome data.

### Allele frequency trajectories across ancient time

By looking through the frequency trajectory from ancient time to present, we found the evidence of positive selection that induced the descent of the critically-ill COVID-19 risk allele *CCR3* rs17217831-A in Sub-Saharan Africans. Using genotypes of ancient DNA^26^, we calculated the allele frequency of *CCR3* rs17217831-A in three archaeological ages starting from 1950 CE (present, 0-1500, 1500+ years).

We detected a declining trend of *CCR3* rs17217831-A frequency in Africans. The frequency showed a trend of descent from 23% at 1500+ years to 6% at present (Figure 1.D & Supplementary table 2). However, the trajectory was sparse due to the limited number of samples in certain ages. We further inferred the frequency trajectory from the genotype data at present based on likelihood^17^. The inferred frequency trajectory coincided well with the frequencies calculated from ancient DNA. We observed an inflating trend from 3000 years ago, but a sharp decline in roughly 1500 years, indicating recent unfavored selection on the critically-ill allele (Figure 1.E).

### Natural selection on *CCR3* eQTLs in African

We identified four eQTLs (rs11926063, rs13062523, rs35984988 and rs41537946) showed suggestive-level (iHS > 1 and XP-EHH > 1) natural selection signals. Importantly, these eQTLs exhibited negative correlations (BETAs < -0.1) with CCR3 expression in African (Figure 1F) showing the expression of CCR3 may increase following natural selection in African.

### Association between SNPs under natural selection and critically-ill COVID-19 in African ancestry

We next investigated the association between critically-ill COVID-19 and *CCR3* rs17217831 in cohorts of African ancestry. As of May 2021, 504 participants of African ancestry in the UK Biobank were tested positive for SARS-CoV-2 infection, of which 36 presented with critical illness. *CCR3* rs17217831 was not associated with critical illness (p= 0.381). The meta-analysis in HGI African GWAS results (version 7) also did not show significant association (p = 0.56). These insignificant associations could be attributed to the low frequency of the risk allele in modern African population (risk allele frequency *CCR3* rs17217831 =0.05).

### Causal relationship between *CCR3* expression and critically-ill COVID-19

The instrumental variables for *CCR3* expression were obtained from GTEx database. We obtained 575 *CCR3* eQTLs in blood samples of European ancestry and 5 remained for further analysis after LD-clumping (P < 5 × 10^-7^, Supplementary Table 3). Using these eQTLs, we found the genetic susceptibility of *CCR3* expression and critically-ill COVID-19 were colocalized. The probability of the causal variants of these phenotypes sharing the same locus was 1 (Fig.2 A). A significant horizontal pleiotropic effect of these eQTL was estimated (MR-PRESSO global test p < 0.001), which also indicated the shared genetic basis between *CCR3* expression level and critical illness. We further conducted a Mendelian Randomization analysis which corrected for this pleiotropy effect. The result revealed that *CCR3* expression level (the exposure) was causally associated with decreased risk of critical illness (OR = 0.89 [0.87, 0.92], p = 0.015, Fig.2 B). The robust test with weighted median model obtained the consistent result (Supplementary Table 4). Our results indicated *CCR3* as another important therapeutic target for critically-ill COVID-19.

**Figure 2.**
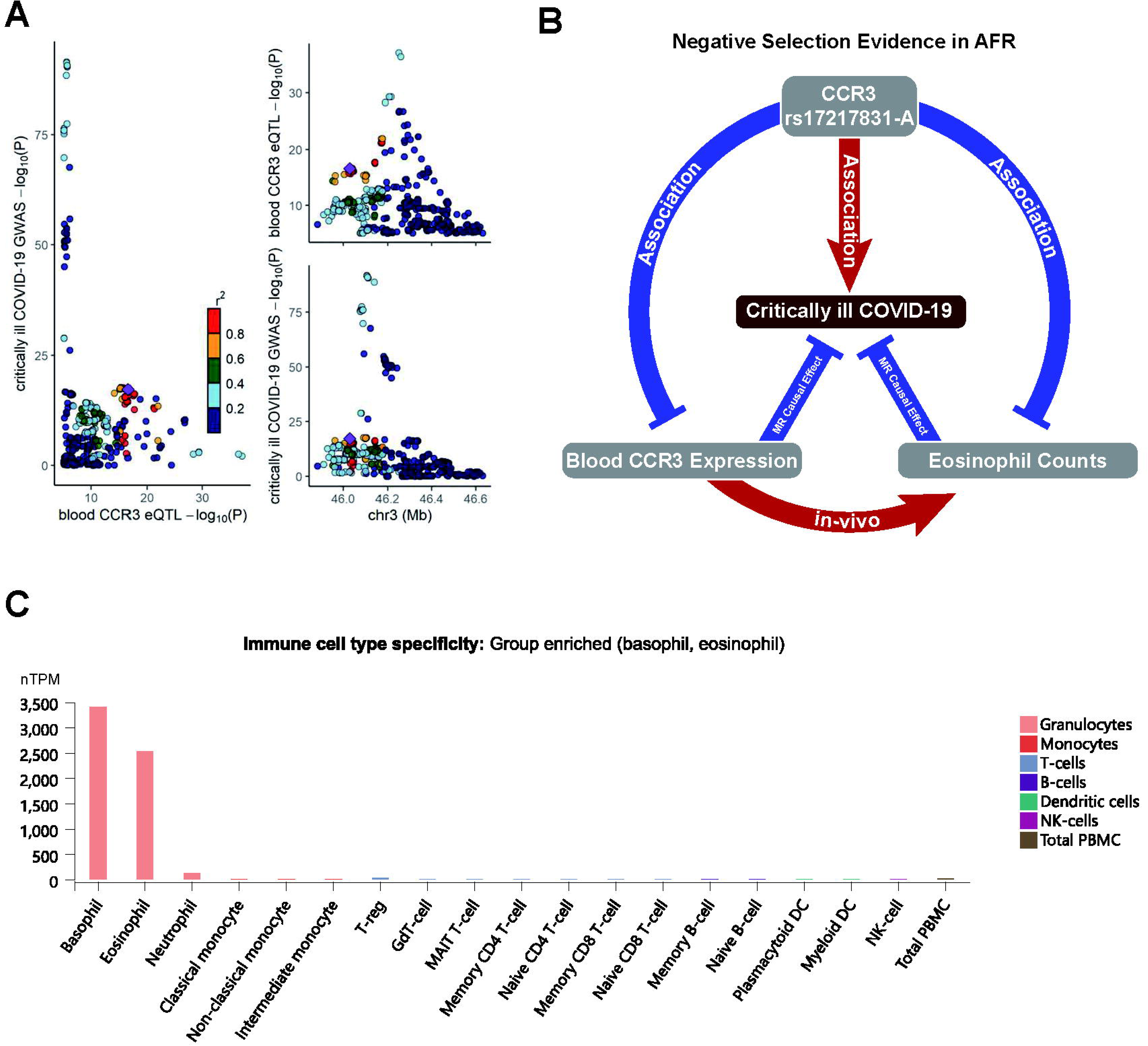
Colocalization and mendelian randomization analysis for CCR3 and critically-ill COVID-19. (A) Comparing the significant signals of *CCR3* eQTL and critically-ill COVID-19 in a same locus. r^2^ indicates the linkage disequilibrium of these variants. (B)Relationship between *CCR3* rs17217831, blood *CCR3* expression, critically-ill COVID-19 and eosinophil counts. Red arrows mean they are positively correlated. Blue blunts mean they are negatively correlated. (C) Expression levels of *CCR3* in different immune cell

## Discussion

Coronavirus disease (COVID-19) presents a wide range of clinical manifestations, from asymptomatic to severe clinical course. Recent studies have shown that ethnicity is relevant to severe COVID-19 outcomes^27,28^. Genes have undergone natural selection in sub-Saharan Africa. For example, studies have shown that ADH gene regions, which includes the ADH1B*48His allele, has experienced recent positive selection in multiple agriculturalist populations from sub-Saharan Africa over the past 2,000 years or so^29^. In this study, we confirmed that the protective alleles of critically-ill COVID-19, at least in part, were more prevalent in African people due to natural selection. We identified significant selection signal of *CCR3* rs17217831-A in Sub-Saharan African. We further revealed that *CCR3* expression level was causally associated with decreased risk of critical illness.

A study identified that a chromosome 3 haplotype which was inherited from Neanderthals, increases the risk of severe COVID-19 ^30^. Close to the locus where this haplotype is identified, locates two chemokine receptor genes, *CCR2* which has been associated with severe COVID-19 ^31^, and *CCR3*. *CCR2* is a monocyte/macrophage specific chemokine receptor, while *CCR3* is expressed mainly in eosinophils and basophils, but also a subset of Th2 lymphocytes, mast cells and myeloid-derived suppressor cells (MDSC) (Figure 2.B and C) ^32–34^. Recent studies have confirmed that these CCR3+ cells involved in a variety of diseases including asthma ^35^, tumors ^36^ and parasitic infection ^37^. Over the past few years, extensive research has confirmed Chemokine signaling through CCR3 is a key regulatory pathway for eosinophil associated with allergic inflammation and asthma ^38,39^. Furthermore, investigations have indicated that the activation of CCR3 on Th-2 cells and basophils can stimulate the release of inflammatory factors such as IL-4, thereby facilitating the activation of eosinophils ^40,41^. Notably, in the context of chronic Staphylococcus aureus infection, MDSCs exhibit eosinophilic granules and express variable levels of CCR3 ^34^. These specialized cells, known as Eo-MDSCs, accumulate at the infection site, playing a substantial role in exacerbating the infection. These findings collectively underscore the indispensable role of eosinophils, orchestrated through the CCR3 surface receptor, in the pathogenesis of various diseases. Of note, the *CCR3* rs17217831-A under selection in Africans was associated with lower eosinophil counts^42^ which was also causally associated with higher risk of critically-ill COVID-19 in our Mendelian randomization analysis. As a result, our study indicated that *CCR3* rs17217831-A may increase the risk of critically-ill COVID-19 either by reducing blood *CCR3* expression level or eosinophil counts. In support for this notion, recent *in vivo* studies showed that *CCR3* blockage or knockout could inhibit the recruitment and migration of eosinophils to the blood, airway and lung airspace^38,43^. Multiple studies of the immune profile of COVID-19 patients have also demonstrate that decreased eosinophil count is associated with a worse disease outcome^44–46^.

Nevertheless, the role of eosinophils in the progression of COVID-19 remains controversial and the underlying reasons deserve further investigations. Lucas et. al. reported that increased eosinophil counts was associated with severe COVID-19 in 253 US residents^47^. To be noted, the participants in this study achieved a relatively high percentage (29.2%) of black people as compared with those in other reports, while ethnicity was not adjusted in their analysis. On the other hand, excessive eosinophils in lung could also lead to a specific type of pneumonia^48^. However, this may represent a distinct etiological pathway as from the pneumonia induced by SARS-cov-2 infection. From a physiology point of view, eosinophils are both pro-inflammatory and anti-viral. It is plausible to believe that eosinophils help reducing the viral load in the early stages of COVID-19, while cause respiratory failure in the late stages by a synergetic action with the cytokine storm. As a result, observed association between eosinophil level and disease severity may vary by the time when samples were taken. In addition, most of the studies investigated the eosinophil counts in peripheral blood, while the status of blood eosinophils could be different from their lung-resident counterparts. Using blood eosinophil as a proxy of the severity of lung disease may lead to a confounded conclusion. Furthermore, the limited sample size and the difference of ethnic composition among cohorts could also contribute to the controversial results^49^.

*IFNAR2* showed a suggestive selection signal in African population with a missense variant rs2229207 standardized iHS = 1.85, which exceeded 94.3% quantile of African genome-wide standardized iHS. *IFNAR2* encodes the type I interferon receptor (IFN-α/β) ^50^. It was associated with critically-ill COVID-19^51^ and confirmed by a Mendelian randomization analysis aimed for drug repurposing^52^ which validate the rationale of using natural selection to detect drug target.

Our study was limited in the genomic analysis of 1000 Genomes Project data. No biological experiments were undertaken to ascertain the involvement of CCR3 or eosinophils in severe COVID-19 disease. Future studies should validate the therapeutic efficacy of CCR3 and eosinophils in the context of critically-ill COVID-19.

Taken together, the *CCR3* risk allele of critical ill COVID-19 was under recent natural selection in Africans, which may make Africans less vulnerable to the critical illness. A possible mechanism of this protective effect could originate from a higher eosinophil count. However, the mechanism and efficacy of regulating eosinophil in the treatment of critically-ill COVID-19 shall be further investigated.

## Conclusion

From a population genetics point-of-view, our study indicated *CCR3* as a potential therapeutic target for critically-ill COVID-19. The indicated therapeutic mechanism builds a bridge between the natural selection of the critical illness allele and the development of novel critical illness treatments.

## Data Availability

All data produced in the present work are contained in the manuscript.

## Acknowledgments

This study used the UK Biobank resources under the application number 53562. We thank the David Reich Lab for providing Ancient DNA genotype data. We would also acknowledge the participants and investigators of the FinnGen study.

## Author Contributions

P.C conceptualized and supervised the study. P.C collected the data. Z.S and A.T analyzed the data. P.C, Z.S and L.P interpreted the results. Z.S and P.C wrote the original manuscript. P.C revised the manuscript.

All authors approved the submission.

## Data Availability Statement

The individual-level genetic data are available on 1000 genome project website (https://www.internationalgenome.org/) and David Reich Lab website (https://reich.hms.harvard.edu/allen-ancient-dna-resource-aadr-downloadable-genotypes-present-day-and-ancient-dna-data). IHS, XPEHH, Fst values will be available through application after publication of this study.

