## Supplementary Files for "Critically-ill COVID-19 susceptibility gene *CCR3* shows natural selection in sub-Saharan Africans"

Title

**Supplementary table**

Supplementary Table1. Natural selection result for critically ill COVID-19 SNPs

Supplementary Table 2. LD information for five eQTLs

Supplementary Table 3. Blood CCR3 expression from GTEx used for mendelian randomization analysis & colocalization

Supplementary Table 4. Mendelian randomization result

Supplementary Table 5. Ancient and present samples from AADR database

**Supplementary Table1. Natural selection result for critically ill COVID-19 SNPs**

| gene_ids | gene_names | ID | CHROM | POS | Freq_AFR | Freq_HGI_GWAS | REF | ALT | standardized XPEHH | AFR \|standardized iHS\| | EUR \|standardized iHS\| | FST | HGI_GWAS_beta | HGI_GWAS_se | HGI_GWAS_p |
|---|---|---|---|---|---|---|---|---|---|---|---|---|---|---|---|
| ENSG00000183625 | CCR3 | rs13086063 | 3 | 46200860 | 0.22 | 0.11 | T | C | 0.81 | 6.31 | 0.41 | 0.10 | 0.31 | 0.02 | 9.99E-50 |
| ENSG00000183625 | CCR3 | rs3091311 | 3 | 46306476 | 0.38 | 0.35 | T | C | 2.57 | 1.52 | 0.11 | 0.06 | 0.11 | 0.01 | 1.25E-15 |
| ENSG00000183625 | CCR3 | rs17217831 | 3 | 46305441 | 0.06 | 0.09 | C | A | 2.49 | 2.00 | 0.55 | 0.07 | 0.35 | 0.02 | 4.86E-47 |
| ENSG00000144815 | NXPE3 | rs34648996 | 3 | 101511202 | 0.28 | 0.34 | A | G | 0.23 | 4.22 | 0.73 | 0.01 | -0.06 | 0.01 | 6.19E-06 |
| ENSG00000162944 | RFTN2 | rs61054961 | 2 | 198494065 | 0.30 | 0.45 | A | C | -1.86 | 4.22 | 0.29 | 0.15 | 0.07 | 0.01 | 2.12E-06 |
| ENSG00000211456 | SACM1L | rs9311378 | 3 | 45763135 | 0.34 | 0.52 | C | T | 0.97 | 4.04 | 1.12 | 0.12 | -0.07 | 0.01 | 4.96E-08 |
| ENSG00000 | ARID3B | rs76738358 | 15 | 74846276 | 0.19 | 0.10 | T | G | -0.50 | 3.93 | 1.30 | 0.09 | 0.12 | 0.03 | 2.73E-06 |

| | | | | | | | | | | | | | | | |
|---|---|---|---|---|---|---|---|---|---|---|---|---|---|---|---|
| 179361 | | | | | | | | | | | | | | | |
| ENSG00000251396 | LINC01301 | rs146987590 | 8 | 61363597 | 0.41 | 0.39 | CAGAA | C | 0.01 | 3.29 | 0.20 | 0.03 | 0.07 | 0.01 | 2.53E-08 |
| ENSG00000159110 | IFNAR2 | rs17860143 | 21 | 34608104 | 0.13 | 0.09 | G | A | 0.29 | 2.73 | 1.71 | 0.04 | -0.12 | 0.02 | 1.59E-07 |
| ENSG00000159110 | IFNAR2 | rs2229207 | 21 | 34614250 | 0.08 | 0.08 | T | C | 1.17 | 1.85 | 1.39 | 0.02 | 0.16 | 0.02 | 1.73E-13 |
| ENSG00000137478 | FCHSD2 | rs7946861 | 11 | 72557986 | 0.30 | 0.24 | A | G | 0.49 | 2.66 | 3.19 | 0.05 | 0.07 | 0.01 | 1.30E-06 |
| ENSG00000163818 | LZTFL1 | rs1488369 | 3 | 45885360 | 0.53 | 0.54 | C | T | -0.07 | 2.64 | 0.93 | 0.03 | -0.13 | 0.01 | 1.02E-23 |
| ENSG00000112787 | FBRSL1 | rs4883552 | 12 | 133145089 | 0.24 | 0.31 | G | A | 1.62 | 2.59 | 1.74 | 0.02 | -0.07 | 0.01 | 1.12E-07 |
| ENSG00000175164 | ABO | rs543968 | 9 | 136143121 | 0.45 | 0.61 | C | T | 0.19 | 2.57 | 1.74 | 0.03 | -0.11 | 0.02 | 1.11E-06 |
| ENSG00000 | MAPT-AS1 | rs56040418 | 17 | 43958079 | 0.19 | 0.18 | C | T | 1.11 | 2.57 | 0.50 | 0.08 | -0.12 | 0.02 | 1.60E-14 |

| | | | | | | | | | | | | | | | |
|---|---|---|---|---|---|---|---|---|---|---|---|---|---|---|---|
| 264589 | | | | | | | | | | | | | | | |
| ENSG00000163820,ENSG00000172215 | FYCO1,CXCR6 | rs6785091 | 3 | 45985347 | 0.33 | 0.14 | C | G | 0.83 | 2.50 | 1.50 | 0.23 | 0.40 | 0.02 | 1.89E-100 |
| ENSG00000225190 | PLEKHM1 | rs62065442 | 17 | 43563894 | 0.12 | 0.14 | T | C | -0.06 | 2.45 | 0.20 | 0.08 | -0.10 | 0.02 | 2.76E-06 |
| ENSG00000173578 | XCR1 | rs35657218 | 3 | 46094462 | 0.42 | 0.32 | A | G | 0.24 | 2.33 | 0.95 | 0.05 | 0.18 | 0.01 | 1.58E-41 |
| ENSG00000285647 | ENSG00000285647 | rs2923000 | 6 | 31339632 | 0.55 | 0.61 | G | A | 2.63 | 2.33 | 1.38 | 0.04 | -0.08 | 0.01 | 1.03E-08 |
| ENSG00000108344 | PSMD3 | rs57968500 | 17 | 38145828 | 0.32 | 0.38 | A | G | 0.30 | 2.30 | 0.96 | 0.03 | -0.06 | 0.01 | 1.21E-06 |
| ENSG00000231402 | WASF5P | rs28506073 | 6 | 31255809 | 0.20 | 0.12 | A | G | 0.84 | 2.27 | 1.69 | 0.07 | 0.10 | 0.02 | 1.27E-07 |
| ENSG00000 | SFTPA1 | rs145783872 | 10 | 81372660 | 0.49 | 0.57 | CTTG | C | 0.20 | 2.24 | 1.44 | 0.06 | -0.11 | 0.02 | 2.94E-08 |

| | | | | | | | | | | | | | | | |
|---|---|---|---|---|---|---|---|---|---|---|---|---|---|---|---|
| 122852 | | | | | | | | | | | | | | | |
| ENSG00000206077 | ZDHHC11B | rs4956967 | 5 | 732478 | 0.11 | 0.19 | C | T | 1.85 | 2.21 | 0.96 | 0.03 | -0.12 | 0.03 | 4.07E-06 |
| ENSG00000163818 | LZTFL1 | rs74586549 | 3 | 45926043 | 0.28 | 0.14 | C | T | 0.19 | 2.18 | 0.18 | 0.18 | 0.41 | 0.02 | 7.17E-106 |
| ENSG00000167914 | GSDMA | rs56030650 | 17 | 38131187 | 0.38 | 0.45 | C | A | 0.06 | 2.18 | 0.09 | 0.03 | -0.06 | 0.01 | 7.67E-06 |
| ENSG00000163827 | LRRC2 | rs34667937 | 3 | 46576351 | 0.23 | 0.38 | T | G | 0.66 | 2.17 | 0.13 | 0.09 | 0.07 | 0.01 | 1.95E-07 |
| ENSG00000225484 | NUTM2B-AS1 | rs61860601 | 10 | 81551568 | 0.35 | 0.33 | G | A | -1.14 | 2.15 | 2.44 | 0.03 | 0.11 | 0.02 | 1.03E-06 |
| ENSG00000131408 | NR1H2 | rs17373080 | 19 | 50879524 | 0.25 | 0.32 | C | G | -1.88 | 2.15 | 0.30 | 0.04 | 0.09 | 0.01 | 9.31E-11 |
| ENSG00000259803 | SLC22A31 | rs7188277 | 16 | 89263454 | 0.14 | 0.19 | T | C | 1.11 | 2.13 | 0.56 | 0.18 | -0.08 | 0.02 | 8.57E-07 |
| ENSG00000 | CSF3 | rs2227322 | 17 | 38171668 | 0.33 | 0.39 | C | G | 0.17 | 2.12 | 1.09 | 0.02 | -0.06 | 0.01 | 3.17E-06 |

| | | | | | | | | | | | | | | | |
|---|---|---|---|---|---|---|---|---|---|---|---|---|---|---|---|
| 108342 | | | | | | | | | | | | | | | |
| ENSG00000141027 | NCOR1 | rs178812 | 17 | 16097136 | 0.50 | 0.56 | G | T | -1.58 | 2.06 | 0.64 | 0.09 | 0.06 | 0.01 | 4.29E-06 |
| ENSG00000289349 | ENSG00000289349 | rs13024151 | 2 | 176581836 | 0.12 | 0.26 | A | G | 0.91 | 2.01 | 0.52 | 0.13 | 0.07 | 0.02 | 8.89E-06 |
| ENSG00000105538 | RASIP1 | rs11882796 | 19 | 49225766 | 0.27 | 0.48 | A | T | -0.74 | 2.01 | 3.01 | 0.18 | -0.07 | 0.02 | 5.75E-06 |
| ENSG00000159314 | ARHGAP27 | rs55793500 | 17 | 43479748 | 0.13 | 0.14 | T | C | 1.24 | 2.01 | 1.14 | 0.07 | -0.11 | 0.02 | 2.13E-10 |
| ENSG00000237945 | LINC00649 | rs2154568 | 21 | 35325805 | 0.40 | 0.51 | C | G | 0.62 | 2.00 | 0.62 | 0.04 | -0.07 | 0.01 | 2.07E-09 |
| ENSG00000065989 | PDE4A | rs10416143 | 19 | 10567862 | 0.23 | 0.18 | G | A | -0.77 | 1.98 | 1.07 | 0.07 | 0.07 | 0.02 | 3.31E-06 |
| ENSG00000142002 | DPP9 | rs56945978 | 19 | 4721927 | 0.39 | 0.32 | G | C | 0.04 | 1.98 | 0.55 | 0.02 | -0.12 | 0.01 | 3.20E-17 |
| ENSG00000 | ELF5 | rs11032727 | 11 | 34513571 | 0.54 | 0.66 | A | G | 1.51 | 1.97 | 0.29 | 0.02 | 0.06 | 0.01 | 8.18E-06 |

| | | | | | | | | | | | | | | | |
|---|---|---|---|---|---|---|---|---|---|---|---|---|---|---|---|
| 135374 | | | | | | | | | | | | | | | |
| ENSG00000288703 | ENSG00000288703 | rs1491951 | 3 | 46141844 | 0.29 | 0.13 | G | A | 1.46 | 1.95 | 0.50 | 0.16 | 0.40 | 0.02 | 5.15E-98 |
| ENSG00000163820 | FYCO1 | rs11130078 | 3 | 45964168 | 0.55 | 0.61 | T | C | 1.17 | 1.92 | 0.40 | 0.16 | -0.09 | 0.02 | 1.51E-07 |
| ENSG00000288692 | ENSG00000288692 | rs499905 | 4 | 111911023 | 0.67 | NA | T | C | -2.43 | 1.91 | 1.50 | 0.16 | 0.40 | 0.07 | 6.41E-08 |
| ENSG00000263715 | LINC02210-CRHR1 | rs9906974 | 17 | 43822772 | 0.19 | 0.21 | T | C | 1.03 | 1.88 | 1.75 | 0.08 | -0.10 | 0.02 | 8.09E-06 |
| ENSG00000125347 | IRF1 | rs4504381 | 5 | 131785770 | 0.59 | 0.33 | T | G | 1.61 | 1.83 | 0.51 | 0.16 | 0.07 | 0.01 | 1.94E-07 |
| ENSG00000185294 | SPPL2C | rs12373168 | 17 | 43924337 | 0.23 | 0.18 | A | C | 0.85 | 1.81 | 0.36 | 0.09 | -0.11 | 0.02 | 1.26E-12 |
| ENSG00000117500 | TMED5 | rs7553465 | 1 | 93644410 | 0.36 | 0.65 | C | T | 2.34 | 1.81 | 0.82 | 0.25 | -0.06 | 0.01 | 3.41E-06 |
| ENSG00000 | SFTPD | rs10887254 | 10 | 81722862 | 0.15 | 0.10 | T | G | -0.54 | 1.79 | 0.72 | 0.09 | 0.10 | 0.02 | 4.87E-07 |

| | | | | | | | | | | | | | | | |
|---|---|---|---|---|---|---|---|---|---|---|---|---|---|---|---|
| 133661 | | | | | | | | | | | | | | | |
| ENSG00000288724 | ENSG00000288724 | rs6441954 | 3 | 46332840 | 0.54 | 0.45 | T | C | 1.84 | 1.78 | 0.41 | 0.17 | -0.11 | 0.01 | 8.42E-17 |
| ENSG00000173585 | CCR9 | rs3774641 | 3 | 45937833 | 0.12 | 0.20 | G | T | 0.98 | 1.75 | 0.27 | 0.13 | 0.28 | 0.02 | 7.31E-70 |
| ENSG00000068650 | ATP11A | rs1278769 | 13 | 113536627 | 0.78 | 0.76 | A | G | -0.72 | 1.75 | 0.46 | 0.01 | -0.11 | 0.01 | 5.65E-15 |
| ENSG00000142453 | CARM1 | rs2053064 | 19 | 11029669 | 0.24 | 0.28 | T | C | 1.74 | 1.74 | 0.08 | 0.02 | 0.06 | 0.01 | 7.16E-06 |
| ENSG00000111335 | OAS2 | rs1293762 | 12 | 113430836 | 0.75 | 0.59 | T | G | 0.93 | 1.72 | 0.42 | 0.15 | 0.06 | 0.01 | 4.11E-06 |
| ENSG00000186868 | MAPT | rs62063276 | 17 | 44036408 | 0.19 | 0.17 | T | G | 0.80 | 1.72 | 0.12 | 0.08 | -0.12 | 0.02 | 1.77E-13 |
| ENSG00000120088 | CRHR1 | rs17689966 | 17 | 43910455 | 0.50 | 0.61 | G | A | -0.14 | 1.71 | 0.32 | 0.10 | 0.07 | 0.01 | 1.33E-07 |
| ENSG00000 | FOXP4-AS1 | rs2496647 | 6 | 41486990 | 0.43 | 0.68 | G | A | -1.28 | 1.71 | 1.53 | 0.14 | -0.07 | 0.01 | 4.58E-07 |

| | | | | | | | | | | | | | | | |
|---|---|---|---|---|---|---|---|---|---|---|---|---|---|---|---|
| 234753 | | | | | | | | | | | | | | | |
| ENSG00000108379 | WNT3 | rs199498 | 17 | 44865603 | 0.15 | 0.20 | T | C | 0.34 | 1.68 | 0.72 | 0.19 | -0.10 | 0.02 | 4.06E-10 |
| ENSG00000176715 | ACSF3 | rs12598652 | 16 | 89230316 | 0.32 | 0.26 | T | C | 0.31 | 1.68 | 2.72 | 0.12 | 0.08 | 0.02 | 1.94E-08 |
| ENSG00000129910 | CDH15 | rs2287356 | 16 | 89251798 | 0.26 | 0.23 | C | T | 1.23 | 1.67 | 1.04 | 0.01 | 0.07 | 0.02 | 9.99E-07 |
| ENSG00000162434 | JAK1 | rs10889509 | 1 | 65400572 | 0.26 | 0.18 | C | T | -1.65 | 1.66 | 0.09 | 0.13 | 0.09 | 0.02 | 4.39E-07 |
| ENSG00000073969 | NSF | rs199461 | 17 | 44762589 | 0.34 | 0.25 | G | A | -0.11 | 1.66 | 1.59 | 0.13 | -0.09 | 0.02 | 3.21E-06 |
| ENSG00000198765 | SYCP1 | rs369718574 | 1 | 115530699 | 0.05 | 0.04 | G | T | 0.06 | 1.63 | NA | 0.02 | 0.27 | 0.06 | 7.68E-06 |
| ENSG00000226676 | LINC02679 | rs1713328 | 10 | 81390377 | 0.47 | 0.38 | C | T | 0.58 | 1.62 | 2.10 | 0.07 | 0.08 | 0.01 | 2.23E-09 |
| ENSG00000 | TIMM29 | rs4804548 | 19 | 11041538 | 0.19 | 0.28 | T | G | 1.67 | 1.60 | 0.03 | 0.03 | 0.07 | 0.01 | 1.17E-06 |

| | | | | | | | | | | | | | | | |
|---|---|---|---|---|---|---|---|---|---|---|---|---|---|---|---|
| 142444 | | | | | | | | | | | | | | | |
| ENSG00000288720 | ENSG00000288720 | rs1488370 | 3 | 45862837 | 0.58 | 0.61 | T | C | 0.25 | 1.56 | 0.45 | 0.13 | -0.15 | 0.01 | 2.70E-28 |
| ENSG00000072682 | P4HA2 | rs13357124 | 5 | 131612287 | 0.12 | 0.04 | G | A | -1.40 | 1.53 | 0.39 | 0.08 | 0.14 | 0.03 | 6.37E-07 |
| ENSG00000173578 | XCR1 | rs9877748 | 3 | 46111081 | 0.35 | 0.13 | A | G | 0.80 | 1.51 | 0.55 | 0.22 | 0.39 | 0.02 | 5.00E-95 |
| ENSG00000104388 | RAB2A | rs34775678 | 8 | 61510495 | 0.68 | 0.41 | ACT | A | -1.55 | 1.51 | 3.07 | 0.18 | 0.07 | 0.01 | 3.92E-08 |
| ENSG00000072682,ENSG00000131435 | P4HA2,PDLIM4 | rs10479001 | 5 | 131607721 | 0.12 | 0.04 | C | T | -1.09 | 1.51 | 0.29 | 0.08 | 0.14 | 0.03 | 6.27E-07 |
| ENSG00000236736 | UQCRC2P1 | rs6441962 | 3 | 46352759 | 0.80 | 0.69 | G | C | 2.15 | 1.50 | 0.65 | 0.05 | -0.09 | 0.02 | 1.51E-06 |
| ENSG00000 | DNM2 | rs11881315 | 19 | 10909953 | 0.18 | 0.18 | C | T | 0.61 | 1.50 | 0.11 | 0.01 | 0.08 | 0.02 | 2.09E-06 |

| | | | | | | | | | | | | | | | |
|---|---|---|---|---|---|---|---|---|---|---|---|---|---|---|---|
| 079805 | | | | | | | | | | | | | | | |
| ENSG00000176715 | ACSF3 | rs12595941 | 16 | 89208680 | 0.30 | 0.25 | T | C | 1.25 | 1.46 | 1.66 | 0.13 | 0.09 | 0.02 | 1.21E-08 |
| ENSG00000204650 | LINC02210 | rs2942166 | 17 | 43715427 | 0.20 | 0.18 | T | C | -0.30 | 1.46 | 1.38 | 0.08 | -0.10 | 0.02 | 4.56E-07 |
| ENSG00000106261 | ZKSCAN1 | rs2897075 | 7 | 99630342 | 0.22 | 0.37 | C | T | 1.93 | 1.45 | 0.66 | 0.10 | 0.08 | 0.01 | 3.80E-10 |
| ENSG00000085276 | MECOM | rs12636139 | 3 | 168896833 | 0.20 | 0.17 | A | G | -0.15 | 1.45 | 1.09 | 0.08 | 0.07 | 0.02 | 9.09E-06 |
| ENSG00000180422 | LINC00304 | rs4581700 | 16 | 89230589 | 0.28 | 0.19 | G | C | 0.18 | 1.45 | 2.09 | 0.07 | 0.10 | 0.02 | 9.92E-07 |
| ENSG00000012223 | LTF | rs3828477 | 3 | 46483717 | 0.25 | 0.29 | T | G | 1.27 | 1.45 | 0.97 | 0.09 | 0.10 | 0.02 | 1.28E-11 |
| ENSG00000214313 | AZGP1P1 | rs11761882 | 7 | 99581469 | 0.26 | 0.39 | T | C | 1.20 | 1.41 | 0.97 | 0.16 | 0.06 | 0.01 | 5.84E-06 |
| ENSG00000 | CCDC18 | rs693737 | 1 | 93724924 | 0.64 | 0.35 | G | A | 1.03 | 1.41 | 1.66 | 0.25 | 0.06 | 0.01 | 3.94E-06 |

| | | | | | | | | | | | | | | | |
|---|---|---|---|---|---|---|---|---|---|---|---|---|---|---|---|
| 122483 | | | | | | | | | | | | | | | |
| ENSG00000214212 | C19orf38 | rs17001002 | 19 | 10948031 | 0.13 | 0.16 | G | A | 0.69 | 1.39 | 0.95 | 0.00 | 0.08 | 0.02 | 2.14E-06 |
| ENSG00000206503 | HLA-A | rs9260081 | 6 | 29909153 | 0.69 | 0.59 | G | T | -0.27 | 1.38 | 2.61 | 0.02 | -0.09 | 0.02 | 1.01E-08 |
| ENSG00000223745 | CCDC18-AS1 | rs11164902 | 1 | 93778794 | 0.36 | 0.65 | A | G | -0.90 | 1.37 | 1.01 | 0.25 | -0.06 | 0.01 | 7.31E-06 |
| ENSG00000143537 | ADAM15 | rs78266397 | 1 | 155023634 | 0.14 | 0.24 | G | A | 0.70 | 1.36 | 0.86 | 0.41 | 0.08 | 0.02 | 1.66E-06 |
| ENSG00000227939 | RPL3P2 | rs17198685 | 6 | 31248648 | 0.20 | 0.12 | C | T | 1.05 | 1.35 | 1.34 | 0.03 | 0.09 | 0.02 | 3.49E-06 |
| ENSG00000127616 | SMARCA4 | rs73013169 | 19 | 11127069 | 0.16 | 0.32 | G | T | 1.60 | 1.34 | 0.39 | 0.10 | 0.06 | 0.01 | 6.73E-06 |
| ENSG00000204542 | C6orf15 | rs2233977 | 6 | 31079894 | 0.12 | 0.17 | A | G | 1.39 | 1.34 | 0.14 | 0.08 | -0.09 | 0.02 | 3.32E-07 |
| ENSG00000 | TMED1 | rs73007599 | 19 | 10946416 | 0.13 | 0.16 | C | T | 0.52 | 1.34 | 0.97 | 0.00 | 0.09 | 0.02 | 2.86E-07 |

| | | | | | | | | | | | | | | | |
|---|---|---|---|---|---|---|---|---|---|---|---|---|---|---|---|
| 099203 | | | | | | | | | | | | | | | |
| ENSG00000163817 | SLC6A20 | rs7653640 | 3 | 45832958 | 0.12 | 0.15 | T | C | 1.76 | 1.33 | 0.61 | 0.02 | -0.08 | 0.02 | 4.91E-06 |
| ENSG00000163825 | RTP3 | rs13353428 | 3 | 46540310 | 0.21 | 0.11 | C | T | 1.01 | 1.30 | 0.21 | 0.07 | 0.16 | 0.02 | 2.14E-14 |
| ENSG00000287390 | ENSG00000287390 | rs13357247 | 5 | 130137357 | 0.14 | 0.04 | C | T | 0.07 | 1.30 | 1.35 | 0.12 | 0.13 | 0.03 | 2.21E-06 |
| ENSG00000243646 | IL10RB | rs2834168 | 21 | 34650791 | 0.21 | 0.30 | A | G | 1.43 | 1.29 | 0.20 | 0.02 | 0.07 | 0.01 | 1.53E-07 |
| ENSG00000205015 | LINC02138 | rs12597281 | 16 | 89234676 | 0.33 | 0.28 | A | G | -0.06 | 1.28 | 2.32 | 0.12 | 0.09 | 0.01 | 4.34E-09 |
| ENSG00000089127 | OAS1 | rs1131454 | 12 | 113348870 | 0.33 | 0.58 | G | A | 0.45 | 1.26 | 0.21 | 0.18 | 0.08 | 0.01 | 2.86E-09 |
| ENSG00000288717 | ENSG00000288717 | rs9869458 | 3 | 46038575 | 0.82 | 0.76 | G | A | 0.83 | 1.25 | 0.36 | 0.06 | 0.11 | 0.02 | 8.69E-13 |
| ENSG00000 | PLSCR1 | rs343316 | 3 | 146238938 | 0.09 | 0.07 | A | G | 0.91 | 1.24 | 0.65 | 0.03 | 0.13 | 0.02 | 6.92E-08 |

| | | | | | | | | | | | | | | |
|---|---|---|---|---|---|---|---|---|---|---|---|---|---|---|
| 188313 | | | | | | | | | | | | | | |
| ENSG00000178951 | ZBTB7A | rs57904377 | 19 | 4050832 | 0.24 | 0.18 | C | T | -1.88 | 1.24 | 0.35 | 0.08 | -0.08 | 0.02 | 6.61E-07 |
| ENSG00000126217 | MCF2L | rs9603802 | 13 | 113565428 | 0.17 | 0.20 | C | G | 1.67 | 1.23 | 0.27 | 0.01 | 0.10 | 0.02 | 1.89E-09 |
| ENSG00000121691 | CAT | rs554576 | 11 | 34483894 | 0.52 | 0.35 | A | T | 0.75 | 1.23 | 0.09 | 0.03 | -0.06 | 0.01 | 4.15E-06 |
| ENSG00000111331 | OAS3 | rs7980275 | 12 | 113380529 | 0.74 | 0.63 | T | A | 0.83 | 1.22 | 0.81 | 0.03 | 0.08 | 0.02 | 4.91E-06 |
| ENSG00000288587 | ENSG00000288587 | rs3093960 | 6 | 31403518 | 0.89 | 0.86 | T | C | 1.68 | 1.20 | 3.20 | 0.01 | -0.10 | 0.02 | 8.16E-07 |
| ENSG00000286940 | MICB-DT | rs9267293 | 6 | 31458508 | 0.87 | 0.82 | G | C | 2.09 | 1.20 | 2.84 | 0.01 | -0.09 | 0.02 | 5.53E-07 |
| ENSG00000186868,ENSG000 | MAPT,MAPT-IT1 | rs4792891 | 17 | 43973498 | 0.34 | 0.29 | T | G | 1.43 | 1.20 | 0.28 | 0.13 | -0.08 | 0.01 | 4.55E-08 |

| | | | | | | | | | | | | | | | |
|---|---|---|---|---|---|---|---|---|---|---|---|---|---|---|---|
| 00279685 | | | | | | | | | | | | | | | |
| ENSG00000228432 | DHFRP2 | rs2844575 | 6 | 31334945 | 0.47 | 0.45 | T | C | 2.28 | 1.18 | 0.75 | 0.01 | -0.07 | 0.01 | 1.79E-07 |
| ENSG00000171634 | BPTF | rs7223643 | 17 | 65830282 | 0.38 | 0.21 | A | G | 0.14 | 1.18 | 0.54 | 0.15 | 0.07 | 0.02 | 8.62E-06 |
| ENSG00000177628 | GBA | rs28678003 | 1 | 155207733 | 0.09 | 0.04 | C | T | 0.41 | 1.18 | 0.63 | 0.04 | -0.22 | 0.03 | 5.88E-12 |
| ENSG00000131400 | NAPSA | rs34541118 | 19 | 50864721 | 0.24 | 0.32 | C | T | -0.70 | 1.18 | 0.21 | 0.05 | 0.09 | 0.02 | 5.26E-07 |
| ENSG00000228862 | ENSG00000228862 | rs670644 | 8 | 61551106 | 0.74 | 0.50 | C | T | 0.35 | 1.18 | 1.10 | 0.18 | 0.06 | 0.01 | 8.95E-06 |
| ENSG00000204540 | PSORS1C1 | rs3131004 | 6 | 31095294 | 0.53 | 0.52 | A | G | 1.96 | 1.18 | 0.06 | 0.01 | -0.08 | 0.01 | 1.12E-09 |
| ENSG00000105376 | ICAM5 | rs885743 | 19 | 10406635 | 0.25 | 0.43 | T | A | 1.64 | 1.16 | 0.45 | 0.16 | -0.09 | 0.01 | 8.13E-13 |
| ENSG00000 | USP8P1 | rs73392806 | 6 | 31244865 | 0.20 | 0.12 | C | T | 1.08 | 1.16 | 1.36 | 0.03 | 0.09 | 0.02 | 4.25E-06 |

| | | | | | | | | | | | | | | | |
|---|---|---|---|---|---|---|---|---|---|---|---|---|---|---|---|
| 214892 | | | | | | | | | | | | | | | |
| ENSG00000204511 | MCCD1 | rs3130055 | 6 | 31497399 | 0.79 | 0.71 | C | T | 2.11 | 1.14 | 0.78 | 0.02 | -0.07 | 0.01 | 2.92E-06 |
| ENSG00000236699 | ARHGEF38 | rs4699192 | 4 | 106578151 | 0.45 | 0.13 | G | A | 0.20 | 1.13 | 0.38 | 0.38 | 0.08 | 0.02 | 7.26E-06 |
| ENSG00000226301 | PDCL3P1 | rs10957133 | 8 | 61300510 | 0.69 | 0.40 | T | C | 1.01 | 1.13 | 0.46 | 0.21 | 0.06 | 0.01 | 2.68E-06 |
| ENSG00000112787 | FBRSL1 | rs10781621 | 12 | 133142935 | 0.51 | 0.45 | A | G | 0.45 | 1.13 | 0.48 | 0.10 | -0.08 | 0.01 | 1.37E-09 |
| ENSG00000156958 | GALK2 | rs139009095 | 15 | 49460908 | 0.12 | 0.03 | C | T | -0.13 | 1.13 | NA | 0.07 | 0.16 | 0.04 | 7.44E-06 |
| ENSG00000267673 | FDX2 | rs78295726 | 19 | 10426512 | 0.12 | 0.16 | C | T | 1.71 | 1.12 | 2.28 | 0.04 | 0.10 | 0.02 | 1.08E-08 |
| ENSG00000090339 | ICAM1 | rs5498 | 19 | 10395683 | 0.28 | 0.44 | A | G | 1.92 | 1.11 | 0.79 | 0.14 | -0.08 | 0.01 | 9.23E-11 |
| ENSG00000 | ZNF3 | rs12673441 | 7 | 99670328 | 0.16 | 0.25 | A | T | 1.39 | 1.11 | 0.04 | 0.09 | 0.07 | 0.01 | 6.30E-07 |

| | | | | | | | | | | | | | | | |
|---|---|---|---|---|---|---|---|---|---|---|---|---|---|---|---|
| 166526 | | | | | | | | | | | | | | | |
| ENSG00000115464 | USP34 | rs921153 | 2 | 61563408 | 0.11 | 0.16 | G | A | 1.13 | 1.10 | 0.92 | 0.09 | 0.07 | 0.02 | 6.31E-06 |
| ENSG00000157601 | MX1 | rs8128997 | 21 | 42832796 | 0.29 | 0.19 | T | C | 0.41 | 1.09 | 0.29 | 0.06 | -0.08 | 0.02 | 6.54E-06 |
| ENSG00000198563 | DDX39B | rs3093978 | 6 | 31498497 | 0.86 | 0.82 | C | A | 1.48 | 1.08 | 1.18 | 0.01 | -0.08 | 0.02 | 3.28E-06 |
| ENSG00000225484 | NUTM2B-AS1 | rs2559675 | 10 | 81434293 | 0.60 | 0.41 | G | A | -0.39 | 1.06 | 3.73 | 0.08 | 0.12 | 0.02 | 3.65E-09 |
| ENSG00000184012 | TMPRSS2 | rs430915 | 21 | 42864743 | 0.72 | 0.55 | A | G | 1.68 | 1.06 | 0.96 | 0.13 | -0.06 | 0.01 | 8.86E-07 |
| ENSG00000180739 | S1PR5 | rs10416073 | 19 | 10625796 | 0.92 | 0.94 | A | C | 0.78 | 1.06 | NA | 0.03 | -0.15 | 0.03 | 2.71E-08 |
| ENSG00000182264 | IZUMO1 | rs12462324 | 19 | 49247086 | 0.35 | 0.44 | A | G | -0.43 | 1.05 | 1.05 | 0.18 | 0.06 | 0.01 | 3.10E-07 |
| ENSG00000 | ENSG00000 | rs2758558 | 10 | 81683578 | 0.77 | 0.59 | G | C | 1.18 | 1.04 | 0.95 | 0.08 | 0.07 | 0.01 | 1.31E-07 |

| | | | | | | | | | | | | | | | |
|---|---|---|---|---|---|---|---|---|---|---|---|---|---|---|---|
| 283913 | 283913 | | | | | | | | | | | | | | |
| ENSG00000163462 | TRIM46 | rs3814316 | 1 | 155149718 | 0.28 | 0.40 | G | A | 1.22 | 1.04 | 0.06 | 0.18 | 0.10 | 0.01 | 9.46E-14 |
| ENSG00000166508 | MCM7 | rs12267 | 7 | 99693078 | 0.15 | 0.24 | G | A | 1.88 | 1.04 | 0.06 | 0.10 | 0.07 | 0.01 | 1.74E-06 |
| ENSG00000206344 | HCG27 | rs9263870 | 6 | 31170514 | 0.13 | 0.12 | A | G | 1.48 | 1.03 | 0.05 | 0.01 | 0.10 | 0.02 | 1.06E-07 |
| ENSG00000242600 | MBL1P | rs1054053 | 10 | 81682645 | 0.77 | 0.60 | A | G | 1.19 | 1.01 | 1.02 | 0.08 | 0.07 | 0.01 | 1.02E-07 |
| ENSG00000120071 | KANSL1 | rs2066899 | 17 | 44155732 | 0.29 | 0.18 | C | T | 0.48 | 1.01 | 0.13 | 0.13 | -0.12 | 0.02 | 3.90E-13 |
| ENSG00000138785 | INTS12 | rs59462153 | 4 | 106815088 | 0.20 | 0.06 | T | C | -0.74 | 1.01 | 1.34 | 0.18 | 0.10 | 0.02 | 8.84E-06 |
| ENSG00000143761 | ARF1 | rs1188975 | 1 | 228277690 | 0.24 | 0.10 | C | T | -0.30 | 1.01 | 0.23 | 0.11 | -0.10 | 0.02 | 2.96E-06 |
| ENSG00000 | KEAP1 | rs11085735 | 19 | 10602180 | 0.90 | 0.93 | A | C | 0.91 | 1.00 | 1.54 | 0.04 | -0.17 | 0.03 | 6.92E-11 |

| | | | | | | | | | | | | | | | |
|---|---|---|---|---|---|---|---|---|---|---|---|---|---|---|---|
| 079999 | | | | | | | | | | | | | | | |
| ENSG00000138629 | UBL7 | rs2415243 | 15 | 74746771 | 0.24 | 0.10 | C | T | -1.02 | 0.99 | 1.59 | 0.09 | 0.09 | 0.02 | 3.08E-06 |
| ENSG00000154027 | AK5 | rs6697255 | 1 | 77960140 | 0.34 | 0.40 | T | C | 0.39 | 0.98 | 0.73 | 0.04 | -0.06 | 0.01 | 9.54E-06 |
| ENSG00000103005 | USB1 | rs4784882 | 16 | 58041338 | 0.83 | 0.75 | C | T | -1.21 | 0.98 | 1.06 | 0.09 | -0.09 | 0.02 | 2.24E-06 |
| ENSG00000223784 | LINP1 | rs12764794 | 10 | 6771594 | 0.22 | 0.05 | C | T | 1.47 | 0.98 | 0.10 | 0.08 | -0.12 | 0.02 | 1.73E-06 |
| ENSG00000204536 | CCHCR1 | rs138598988 | 6 | 31115032 | 0.08 | 0.10 | C | T | 1.25 | 0.98 | 2.10 | 0.02 | 0.13 | 0.02 | 4.66E-09 |
| ENSG00000163463 | KRTCAP2 | rs4971093 | 1 | 155144300 | 0.26 | 0.41 | G | A | 1.57 | 0.97 | 0.01 | 0.21 | 0.09 | 0.02 | 5.69E-08 |
| ENSG00000230994 | FGFR3P1 | rs2853976 | 6 | 31345270 | 0.40 | 0.53 | C | T | 2.69 | 0.97 | 0.09 | 0.05 | -0.06 | 0.01 | 6.85E-06 |
| ENSG00000 | ENSG00000 | rs3130468 | 6 | 31190795 | 0.92 | 0.83 | C | G | 1.76 | 0.97 | 2.56 | 0.02 | -0.09 | 0.02 | 6.27E-06 |

| | | | | | | | | | | | | | | | |
|---|---|---|---|---|---|---|---|---|---|---|---|---|---|---|---|
| 255726 | 255726 | | | | | | | | | | | | | | |
| ENSG00000137310 | TCF19 | rs2106074 | 6 | 31133509 | 0.74 | 0.65 | G | A | 1.58 | 0.96 | 0.53 | 0.04 | -0.08 | 0.01 | 6.44E-10 |
| ENSG00000286923 | ENSG00000286923 | rs56173013 | 7 | 99600405 | 0.29 | 0.40 | A | T | 0.79 | 0.96 | 1.49 | 0.12 | 0.07 | 0.01 | 8.87E-07 |
| ENSG00000143761 | ARF1 | rs2070960 | 1 | 228284991 | 0.06 | 0.08 | C | T | -0.23 | 0.95 | 0.39 | 0.03 | -0.13 | 0.02 | 3.92E-07 |
| ENSG00000244040 | IL12A-AS1 | rs9834418 | 3 | 159729622 | 0.14 | 0.02 | A | T | -0.57 | 0.95 | NA | 0.11 | 0.21 | 0.05 | 4.85E-06 |
| ENSG00000138780 | GSTCD | rs7676975 | 4 | 106763811 | 0.16 | 0.07 | A | T | -0.37 | 0.94 | 1.13 | 0.11 | 0.10 | 0.02 | 9.13E-06 |
| ENSG00000122483 | CCDC18 | rs7541687 | 1 | 93656588 | 0.36 | 0.65 | A | G | -0.48 | 0.94 | 0.90 | 0.25 | -0.06 | 0.01 | 4.33E-06 |
| ENSG00000105397 | TYK2 | rs280519 | 19 | 10472933 | 0.54 | 0.52 | A | G | 0.54 | 0.93 | 0.77 | 0.03 | 0.09 | 0.01 | 5.07E-13 |
| ENSG00000 | NSF | rs199455 | 17 | 44799216 | 0.51 | 0.77 | C | T | 0.15 | 0.92 | 1.80 | 0.20 | 0.09 | 0.01 | 5.09E-10 |

| | | | | | | | | | | | | | | | |
|---|---|---|---|---|---|---|---|---|---|---|---|---|---|---|---|
| 073969 | | | | | | | | | | | | | | | |
| ENSG000001791511 | EDC3 | rs6495119 | 15 | 74934440 | 0.20 | 0.10 | T | C | -1.30 | 0.92 | 1.70 | 0.09 | 0.10 | 0.02 | 1.20E-06 |
| ENSG000001607899 | LMNA | rs74116489 | 1 | 156109262 | 0.09 | 0.03 | C | T | -1.36 | 0.92 | NA | 0.06 | 0.20 | 0.04 | 2.74E-07 |
| ENSG000002343233 | LINC01505 | rs996856 | 9 | 109487910 | 0.24 | 0.37 | G | A | -0.54 | 0.92 | 1.10 | 0.07 | 0.07 | 0.01 | 4.87E-07 |
| ENSG000002867033 | ENSG000002867033 | rs10253651 | 7 | 22870183 | 0.27 | 0.27 | A | G | 0.80 | 0.91 | 1.00 | 0.02 | 0.06 | 0.01 | 5.82E-06 |
| ENSG000001224833 | CCDC18 | rs535760 | 1 | 93728758 | 0.64 | 0.34 | T | A | -0.70 | 0.91 | 2.12 | 0.25 | 0.06 | 0.01 | 9.38E-06 |
| ENSG000001450122 | LPP | rs4686945 | 3 | 188006256 | 0.75 | 0.57 | T | C | 0.64 | 0.90 | 0.15 | 0.19 | 0.06 | 0.01 | 3.82E-06 |
| ENSG000002836466 | LINC02009 | rs35439446 | 3 | 46462170 | 0.48 | 0.16 | A | G | -1.22 | 0.90 | 0.19 | 0.26 | 0.14 | 0.02 | 3.66E-15 |
| ENSG00000 | MED24 | rs11078934 | 17 | 38189793 | 0.68 | 0.62 | T | C | 0.02 | 0.90 | 0.02 | 0.02 | 0.06 | 0.01 | 8.58E-06 |

| | | | | | | | | | | | | | | | |
|---|---|---|---|---|---|---|---|---|---|---|---|---|---|---|---|
| 008838 | | | | | | | | | | | | | | | |
| ENSG000002202 04 | ENSG000002202 04 | rs10836215 | 11 | 34426285 | 0.21 | 0.37 | C | T | 1.40 | 0.89 | 0.33 | 0.10 | 0.07 | 0.01 | 8.24E-07 |
| ENSG000001840 12 | TMPRSS2 | rs61459778 | 21 | 42838423 | 0.21 | 0.17 | C | G | 0.03 | 0.89 | 0.14 | 0.08 | -0.09 | 0.02 | 9.57E-06 |
| ENSG000001198 66 | BCL11A | rs7560588 | 2 | 60727416 | 0.17 | 0.25 | G | A | 1.54 | 0.88 | 0.17 | 0.08 | -0.12 | 0.02 | 5.67E-14 |
| ENSG000002725 01 | ENSG000002725 01 | rs28362340 | 6 | 31165359 | 0.13 | 0.12 | G | C | 1.55 | 0.88 | 0.82 | 0.01 | 0.10 | 0.02 | 6.77E-08 |
| ENSG000001769 20 | FUT2 | rs516246 | 19 | 49206172 | 0.47 | 0.45 | C | T | 0.32 | 0.87 | 1.81 | 0.17 | -0.09 | 0.01 | 5.74E-12 |
| ENSG000001279 46 | HIP1 | rs4731232 | 7 | 75264765 | 0.13 | 0.23 | G | A | 0.80 | 0.86 | 1.65 | 0.06 | 0.08 | 0.02 | 5.55E-08 |
| ENSG000002063 37 | HCP5 | rs55886855 | 6 | 31433690 | 0.09 | 0.06 | T | G | 1.94 | 0.85 | 0.37 | 0.00 | -0.12 | 0.02 | 9.52E-07 |
| ENSG00000 | LIMD1 | rs7648681 | 3 | 45677398 | 0.27 | 0.37 | G | A | 0.65 | 0.85 | 0.07 | 0.04 | -0.06 | 0.01 | 7.93E-06 |

| | | | | | | | | | | | | | | | |
|---|---|---|---|---|---|---|---|---|---|---|---|---|---|---|---|
| 144791 | | | | | | | | | | | | | | | |
| ENSG00000204516 | MICB | rs3094003 | 6 | 31469251 | 0.81 | 0.74 | C | G | 1.53 | 0.85 | 2.20 | 0.02 | -0.10 | 0.02 | 4.80E-10 |
| ENSG00000204520 | MICA | rs2596540 | 6 | 31367882 | 0.92 | 0.86 | A | G | 1.52 | 0.85 | 2.80 | 0.01 | -0.10 | 0.02 | 1.05E-06 |
| ENSG00000206503 | HLA-A | rs9260115 | 6 | 29910027 | 0.67 | 0.59 | C | G | 0.10 | 0.83 | 2.52 | 0.01 | -0.08 | 0.01 | 2.40E-09 |
| ENSG00000206344 | HCG27 | rs28362352 | 6 | 31168387 | 0.13 | 0.12 | G | A | 1.57 | 0.82 | 0.18 | 0.01 | 0.10 | 0.02 | 9.61E-08 |
| ENSG00000241383 | LINC01997 | rs2421077 | 3 | 168723285 | 0.66 | 0.79 | A | C | -0.66 | 0.82 | 0.56 | 0.08 | -0.07 | 0.02 | 2.86E-06 |
| ENSG00000126214 | KLC1 | rs12889731 | 14 | 104087482 | 0.17 | 0.31 | C | T | 1.05 | 0.81 | 0.64 | 0.08 | 0.06 | 0.01 | 6.83E-06 |
| ENSG00000204528 | PSORS1C3 | rs2269711 | 6 | 31141612 | 0.77 | 0.62 | A | T | 1.64 | 0.80 | 0.31 | 0.07 | -0.07 | 0.01 | 3.42E-07 |
| ENSG00000 | ENSG00000 | rs2070803 | 1 | 155157715 | 0.38 | 0.55 | G | A | 0.40 | 0.79 | 0.02 | 0.10 | -0.08 | 0.01 | 1.58E-09 |

| | | | | | | | | | | | | | | | |
|---|---|---|---|---|---|---|---|---|---|---|---|---|---|---|---|
| 273088 | 273088 | | | | | | | | | | | | | | |
| ENSG00000176788 | BASP1 | rs80191700 | 5 | 17184351 | 0.05 | 0.01 | T | C | 0.25 | 0.79 | NA | 0.04 | 0.28 | 0.06 | 9.72E-06 |
| ENSG00000220201 | ZGLP1 | rs73510898 | 19 | 10416444 | 0.09 | 0.08 | G | A | 2.06 | 0.79 | 1.04 | 0.01 | 0.17 | 0.02 | 7.50E-13 |
| ENSG00000127603 | MACF1 | rs687848 | 1 | 39919234 | 0.59 | 0.57 | A | G | -0.26 | 0.78 | 1.00 | 0.07 | 0.06 | 0.01 | 3.19E-06 |
| ENSG00000182504 | CEP97 | rs13096323 | 3 | 101461177 | 0.43 | 0.35 | T | C | 0.13 | 0.78 | 0.33 | 0.06 | -0.07 | 0.01 | 3.05E-06 |
| ENSG00000235040 | MTCO3P1 | rs9275518 | 6 | 32674700 | 0.60 | 0.58 | G | A | 1.19 | 0.77 | 1.06 | 0.02 | -0.07 | 0.01 | 6.58E-07 |
| ENSG00000228432 | DHFRP2 | rs2523560 | 6 | 31330844 | 0.53 | 0.57 | C | T | 1.96 | 0.76 | 0.21 | 0.02 | -0.07 | 0.01 | 3.64E-08 |
| ENSG00000142233 | NTN5 | rs12462111 | 19 | 49171306 | 0.24 | 0.40 | C | T | 0.89 | 0.75 | 1.84 | 0.16 | -0.08 | 0.01 | 1.60E-09 |
| ENSG00000 | KLF13 | rs1465778 | 15 | 31621321 | 0.31 | 0.32 | T | C | 0.86 | 0.75 | 0.28 | 0.03 | -0.06 | 0.01 | 8.33E-06 |

| | | | | | | | | | | | | | | | |
|---|---|---|---|---|---|---|---|---|---|---|---|---|---|---|---|
| 169926 | | | | | | | | | | | | | | | |
| ENSG00000132676 | DAP3 | rs10908480 | 1 | 155699762 | 0.24 | 0.31 | T | G | -1.07 | 0.75 | 0.92 | 0.16 | 0.06 | 0.01 | 9.71E-06 |
| ENSG00000229876 | CASC20 | rs2225353 | 20 | 6499312 | 0.40 | 0.44 | G | T | 0.57 | 0.74 | 1.46 | 0.10 | -0.06 | 0.01 | 7.99E-06 |
| ENSG00000247240 | UBL7-DT | rs6495104 | 15 | 74768101 | 0.20 | 0.10 | G | C | 0.18 | 0.74 | 2.23 | 0.09 | 0.09 | 0.02 | 4.13E-06 |
| ENSG00000142583 | SLC2A5 | rs2505973 | 1 | 9122669 | 0.70 | 0.63 | A | G | 0.86 | 0.72 | 1.24 | 0.06 | -0.07 | 0.01 | 8.16E-08 |
| ENSG00000167680 | SEMA6B | rs9304911 | 19 | 4571589 | 0.76 | 0.62 | C | T | -0.20 | 0.71 | 0.30 | 0.06 | -0.07 | 0.01 | 1.73E-06 |
| ENSG00000196683 | TOMM7 | rs2286498 | 7 | 22857504 | 0.57 | 0.39 | T | C | 1.49 | 0.71 | 0.99 | 0.13 | -0.06 | 0.01 | 2.77E-06 |
| ENSG00000074842 | MYDGF | rs8108253 | 19 | 4656466 | 0.52 | 0.24 | T | C | 0.21 | 0.69 | 1.64 | 0.20 | -0.08 | 0.01 | 2.11E-07 |
| ENSG00000 | UBL7-DT | rs2099593 | 15 | 74771488 | 0.23 | 0.11 | G | A | 0.11 | 0.69 | 1.51 | 0.09 | 0.10 | 0.02 | 1.49E-06 |

| | | | | | | | | | | | | | | | |
|---|---|---|---|---|---|---|---|---|---|---|---|---|---|---|---|
| 247240 | | | | | | | | | | | | | | | |
| ENSG00000105401 | CDC37 | rs11879191 | 19 | 10512911 | 0.15 | 0.19 | G | A | 0.15 | 0.69 | 0.33 | 0.13 | 0.08 | 0.02 | 1.36E-06 |
| ENSG00000229390 | MICD | rs2523966 | 6 | 29939240 | 0.64 | 0.57 | G | A | 0.37 | 0.68 | 0.80 | 0.01 | -0.06 | 0.01 | 7.69E-06 |
| ENSG00000166997 | CNPY4 | rs7799946 | 7 | 99719753 | 0.14 | 0.23 | G | A | 0.29 | 0.68 | 0.51 | 0.11 | 0.08 | 0.01 | 7.34E-08 |
| ENSG00000162434 | JAK1 | rs3014983 | 1 | 65460691 | 0.45 | 0.72 | C | T | 0.01 | 0.68 | 1.41 | 0.24 | -0.09 | 0.02 | 1.79E-08 |
| ENSG00000216708 | CIR1P3 | rs34533645 | 11 | 34452724 | 0.23 | 0.37 | G | A | 0.62 | 0.67 | 0.24 | 0.09 | 0.08 | 0.01 | 2.06E-08 |
| ENSG00000241837 | ATP5PO | rs2834294 | 21 | 35280630 | 0.38 | 0.37 | A | G | 0.64 | 0.67 | 0.01 | 0.03 | 0.07 | 0.01 | 1.28E-07 |
| ENSG00000131408 | NR1H2 | rs11879501 | 19 | 50872118 | 0.35 | 0.34 | A | G | -0.55 | 0.66 | 0.28 | 0.03 | 0.08 | 0.01 | 1.33E-09 |
| ENSG00000 | ACSL6 | rs407204 | 5 | 131327465 | 0.42 | 0.77 | C | G | -2.42 | 0.66 | 1.48 | 0.28 | -0.08 | 0.02 | 2.45E-06 |

| | | | | | | | | | | | | | | |
|---|---|---|---|---|---|---|---|---|---|---|---|---|---|---|
| 164398 | | | | | | | | | | | | | | |
| ENSG00000206344 | HCG27 | rs74568298 | 6 | 31165597 | 0.13 | 0.12 | AT | A | 1.84 | 0.64 | 0.72 | 0.01 | 0.11 | 0.02 | 7.93E-08 |
| ENSG00000251246 | ENSG00000251246 | rs6427137 | 1 | 155048369 | 0.19 | 0.05 | A | G | 0.92 | 0.64 | 0.54 | 0.16 | -0.13 | 0.03 | 1.57E-06 |
| ENSG00000204520 | MICA | rs2857283 | 6 | 31381162 | 0.22 | 0.13 | C | T | 1.57 | 0.64 | 0.25 | 0.02 | -0.08 | 0.02 | 5.16E-06 |
| ENSG00000168010 | ATG16L2 | rs3862794 | 11 | 72538600 | 0.26 | 0.23 | T | C | 0.91 | 0.63 | 0.00 | 0.05 | 0.07 | 0.01 | 7.04E-06 |
| ENSG00000179335 | CLK3 | rs74925675 | 15 | 74905756 | 0.19 | 0.10 | A | C | -1.20 | 0.62 | 1.75 | 0.09 | 0.10 | 0.02 | 5.55E-07 |
| ENSG00000247240 | UBL7-DT | rs6495107 | 15 | 74788191 | 0.19 | 0.10 | C | T | 0.03 | 0.62 | 0.67 | 0.09 | 0.10 | 0.02 | 2.89E-06 |
| ENSG00000131401 | NAPSB | rs35522855 | 19 | 50846561 | 0.24 | 0.30 | A | G | -0.16 | 0.61 | 0.34 | 0.03 | 0.08 | 0.01 | 3.38E-08 |
| ENSG00000 | KAT7 | rs9898183 | 17 | 47907383 | 0.10 | 0.05 | G | C | -0.10 | 0.60 | 0.81 | 0.02 | 0.22 | 0.03 | 4.82E-13 |

| | | | | | | | | | | | | | | | |
|---|---|---|---|---|---|---|---|---|---|---|---|---|---|---|---|
| 136504 | | | | | | | | | | | | | | | |
| ENSG00000076662 | ICAM3 | rs3176766 | 19 | 10449778 | 0.18 | 0.22 | G | A | 1.64 | 0.60 | 0.24 | 0.02 | -0.08 | 0.02 | 4.12E-07 |
| ENSG00000150048 | CLEC1A | rs7974909 | 12 | 10242689 | 0.78 | 0.85 | C | T | -0.06 | 0.59 | 0.10 | 0.16 | -0.12 | 0.03 | 3.09E-06 |
| ENSG00000116580 | GON4L | rs12024187 | 1 | 155777840 | 0.26 | 0.29 | T | A | -1.24 | 0.57 | 1.10 | 0.09 | 0.06 | 0.01 | 9.12E-06 |
| ENSG00000137310 | TCF19 | rs114881571 | 6 | 31128605 | 0.09 | 0.10 | C | T | 1.59 | 0.56 | 2.32 | 0.02 | 0.14 | 0.03 | 3.60E-07 |
| ENSG00000161847 | RAVER1 | rs73019227 | 19 | 10432052 | 0.15 | 0.18 | C | G | 1.68 | 0.56 | 2.11 | 0.02 | -0.09 | 0.02 | 2.58E-08 |
| ENSG00000185361 | TNFAIP8L1 | rs16992471 | 19 | 4640295 | 0.34 | 0.46 | T | G | 1.59 | 0.56 | 1.18 | 0.05 | 0.09 | 0.01 | 8.15E-11 |
| ENSG00000206337 | HCP5 | rs2518030 | 6 | 31431082 | 0.90 | 0.84 | A | G | 1.70 | 0.55 | 1.45 | 0.04 | -0.11 | 0.02 | 8.22E-09 |
| ENSG00000 | MAMSTR | rs204248 | 19 | 49216224 | 0.34 | 0.36 | G | C | -0.48 | 0.55 | 0.86 | 0.18 | 0.08 | 0.01 | 1.68E-08 |

| | | | | | | | | | | | | | | | |
|---|---|---|---|---|---|---|---|---|---|---|---|---|---|---|---|
| 176909 | | | | | | | | | | | | | | | |
| ENSG00000160766 | GBAP1 | rs1057941 | 1 | 155186742 | 0.27 | 0.43 | G | A | 1.62 | 0.54 | 0.45 | 0.20 | 0.09 | 0.02 | 1.16E-06 |
| ENSG00000214584 | PGGT1BP2 | rs927452 | 10 | 81652266 | 0.78 | 0.60 | G | A | 0.94 | 0.54 | 2.03 | 0.09 | 0.07 | 0.01 | 2.64E-07 |
| ENSG00000205726 | ITSN1 | rs3746861 | 21 | 35271200 | 0.11 | 0.08 | C | T | 0.37 | 0.53 | 0.10 | 0.03 | 0.16 | 0.02 | 1.73E-13 |
| ENSG00000285082 | ENSG00000285082 | rs2806091 | 9 | 120818067 | 0.18 | 0.27 | A | G | 0.73 | 0.53 | 0.42 | 0.08 | 0.09 | 0.02 | 6.39E-06 |
| ENSG00000127946 | HIP1 | rs1179625 | 7 | 75255491 | 0.55 | 0.41 | A | G | 0.17 | 0.53 | 0.26 | 0.07 | 0.07 | 0.01 | 1.75E-07 |
| ENSG00000112787 | FBRSL1 | rs2875247 | 12 | 133129836 | 0.36 | 0.53 | C | T | -0.15 | 0.50 | 0.14 | 0.11 | 0.08 | 0.01 | 2.80E-09 |
| ENSG00000066422 | ZBTB11 | rs11712748 | 3 | 101383562 | 0.41 | 0.35 | G | A | 0.75 | 0.50 | 0.90 | 0.04 | -0.06 | 0.01 | 9.63E-06 |
| ENSG00000 | CLK3 | rs4073149 | 15 | 74898419 | 0.24 | 0.11 | G | A | -1.12 | 0.48 | 2.03 | 0.09 | 0.10 | 0.02 | 9.76E-07 |

| | | | | | | | | | | | | | | | |
|---|---|---|---|---|---|---|---|---|---|---|---|---|---|---|---|
| 179335 | | | | | | | | | | | | | | | |
| ENSG00000250751 | ENSG00000250751 | rs78997619 | 17 | 47787161 | 0.07 | 0.06 | C | T | 0.83 | 0.48 | 0.73 | 0.00 | 0.19 | 0.03 | 2.01E-10 |
| ENSG00000011376 | LARS2 | rs140846169 | 3 | 45541785 | 0.07 | 0.07 | C | T | 2.26 | 0.48 | 2.59 | 0.04 | -0.13 | 0.03 | 2.38E-06 |
| ENSG00000142002 | DPP9 | rs8105807 | 19 | 4684642 | 0.58 | 0.27 | A | G | -1.12 | 0.47 | 0.38 | 0.22 | -0.09 | 0.01 | 6.75E-11 |
| ENSG00000255899 | ENSG00000255899 | rs6904669 | 6 | 31192796 | 0.52 | 0.42 | A | G | 1.72 | 0.47 | 0.48 | 0.06 | -0.07 | 0.01 | 1.04E-08 |
| ENSG00000231389 | HLA-DPA1 | rs67035222 | 6 | 33038024 | 0.41 | 0.17 | AT | A | -0.59 | 0.46 | 2.38 | 0.16 | 0.08 | 0.02 | 9.09E-06 |
| ENSG00000170608 | FOXA3 | rs11669442 | 19 | 46374557 | 0.34 | 0.29 | T | C | 0.87 | 0.45 | 0.08 | 0.01 | 0.07 | 0.01 | 1.60E-06 |
| ENSG00000166170 | BAG5 | rs34896432 | 14 | 104027995 | 0.18 | 0.28 | C | T | 0.09 | 0.44 | 0.29 | 0.08 | 0.07 | 0.01 | 4.38E-06 |

| | | | | | | | | | | | | | | | |
|---|---|---|---|---|---|---|---|---|---|---|---|---|---|---|---|
| ENSG00000213104 | NPM1P46 | rs150345524 | 2 | 198245297 | 0.27 | 0.42 | G | A | -0.09 | 0.44 | 0.02 | 0.14 | 0.06 | 0.01 | 7.03E-06 |
| ENSG00000286921 | ENSG00000286921 | rs2283017 | 7 | 99574758 | 0.28 | 0.39 | A | G | 0.17 | 0.43 | 1.19 | 0.13 | 0.06 | 0.01 | 2.28E-06 |
| ENSG00000234745 | HLA-B | rs2596478 | 6 | 31327861 | 0.86 | 0.87 | C | T | 1.67 | 0.43 | 2.18 | 0.02 | -0.10 | 0.02 | 1.06E-06 |
| ENSG00000169231 | THBS3 | rs41264915 | 1 | 155167786 | 0.21 | 0.09 | A | G | 0.33 | 0.42 | 0.48 | 0.08 | -0.19 | 0.02 | 1.54E-20 |
| ENSG00000096696 | DSP | rs2076295 | 6 | 7563232 | 0.47 | 0.43 | T | G | -0.19 | 0.41 | 0.40 | 0.02 | 0.07 | 0.01 | 1.47E-07 |
| ENSG00000204539 | CDSN | rs3132555 | 6 | 31082910 | 0.84 | 0.70 | C | G | 1.98 | 0.41 | 0.68 | 0.04 | -0.11 | 0.02 | 3.97E-13 |
| ENSG00000134070 | IRAK2 | rs182205328 | 3 | 10270708 | 0.05 | 0.04 | C | G | -0.03 | 0.40 | NA | 0.07 | 0.50 | 0.11 | 9.20E-06 |
| ENSG00000126214 | KLC1 | rs12889403 | 14 | 104034746 | 0.17 | 0.28 | C | T | -0.32 | 0.40 | 0.17 | 0.08 | 0.06 | 0.01 | 9.65E-06 |

| | | | | | | | | | | | | | | | |
|---|---|---|---|---|---|---|---|---|---|---|---|---|---|---|---|
| ENSG00000237945 | LINC00649 | rs12482569 | 21 | 35293087 | 0.13 | 0.17 | G | A | 0.67 | 0.38 | 0.48 | 0.03 | 0.10 | 0.02 | 4.90E-09 |
| ENSG00000228570 | NUTM2E | rs10749144 | 10 | 81594285 | 0.38 | 0.58 | C | T | -1.18 | 0.38 | 2.12 | 0.09 | -0.11 | 0.02 | 9.98E-09 |
| ENSG00000173209 | AHSA2P | rs6741734 | 2 | 61407245 | 0.21 | 0.14 | G | T | 1.08 | 0.38 | 0.92 | 0.02 | 0.07 | 0.02 | 8.86E-06 |
| ENSG00000167281 | RBFOX3 | rs11867923 | 17 | 77121313 | 0.26 | 0.35 | T | A | -0.92 | 0.37 | 1.24 | 0.08 | 0.07 | 0.01 | 4.15E-06 |
| ENSG00000289406 | ENSG00000289406 | rs915654 | 6 | 31538497 | 0.44 | 0.35 | T | A | 0.89 | 0.37 | 0.36 | 0.03 | -0.06 | 0.01 | 3.66E-06 |
| ENSG00000074842,ENSG00000185361 | MYDGF,TNFAIP8L1 | rs10417957 | 19 | 4647231 | 0.38 | 0.25 | T | C | -1.38 | 0.36 | 1.66 | 0.09 | -0.08 | 0.02 | 1.53E-07 |
| ENSG00000143590 | EFNA3 | rs1462854 | 1 | 155053682 | 0.26 | 0.06 | T | A | 0.36 | 0.36 | 0.51 | 0.26 | -0.14 | 0.02 | 2.68E-08 |

| | | | | | | | | | | | | | | | |
|---|---|---|---|---|---|---|---|---|---|---|---|---|---|---|---|
| ENSG00000204623,ENSG00000204625 | ZNRD1ASP, HCG9 | rs2844803 | 6 | 29944314 | 0.57 | 0.51 | T | C | 0.27 | 0.35 | 0.34 | 0.02 | -0.07 | 0.01 | 5.52E-08 |
| ENSG00000142166 | IFNAR1 | rs2843996 | 21 | 34711448 | 0.84 | 0.74 | G | A | 1.17 | 0.34 | 1.10 | 0.09 | -0.07 | 0.01 | 4.75E-07 |
| ENSG00000243364 | EFNA4, | rs6675468 | 1 | 155040654 | 0.10 | 0.09 | C | T | 0.08 | 0.33 | 0.04 | 0.03 | -0.16 | 0.02 | 4.53E-13 |
| ENSG00000160862 | AZGP1 | rs2074567 | 7 | 99569225 | 0.28 | 0.39 | T | C | 0.50 | 0.33 | 1.51 | 0.13 | 0.06 | 0.01 | 5.95E-06 |
| ENSG00000121104 | FAM117A | rs77522818 | 17 | 47817373 | 0.07 | 0.07 | A | T | 0.00 | 0.32 | 0.37 | 0.01 | 0.19 | 0.03 | 4.46E-10 |
| ENSG00000120088 | CRHR1 | rs12944712 | 17 | 43871147 | 0.32 | 0.40 | G | A | 1.25 | 0.32 | 0.41 | 0.10 | -0.08 | 0.01 | 1.17E-08 |
| ENSG00000196526 | AFAP1 | rs34072304 | 4 | 7860885 | 0.14 | 0.17 | C | G | -0.36 | 0.31 | 0.33 | 0.01 | 0.08 | 0.02 | 3.04E-06 |

| | | | | | | | | | | | | | | |
|---|---|---|---|---|---|---|---|---|---|---|---|---|---|---|
| ENSG00000196301 | HLA-DRB9 | rs12215261 | 6 | 32441150 | 0.25 | 0.32 | C | T | 1.36 | 0.31 | 0.33 | 0.03 | -0.09 | 0.02 | 3.08E-07 |
| ENSG00000228649 | SNHG26 | rs1608553 | 7 | 22896231 | 0.75 | 0.60 | G | C | 0.87 | 0.30 | 2.65 | 0.05 | -0.07 | 0.01 | 2.87E-07 |
| ENSG00000121073 | SLC35B1 | rs8186 | 17 | 47778793 | 0.10 | 0.06 | C | T | 0.14 | 0.30 | 0.43 | 0.01 | 0.18 | 0.03 | 2.32E-10 |
| ENSG00000121104 | FAM117A | rs34759387 | 17 | 47788755 | 0.07 | 0.06 | G | A | 0.61 | 0.30 | 0.62 | 0.01 | 0.19 | 0.03 | 2.29E-10 |
| ENSG00000204498 | NFKBIL1 | rs2523500 | 6 | 31518354 | 0.70 | 0.68 | G | A | 1.21 | 0.29 | 0.84 | 0.02 | -0.06 | 0.01 | 2.33E-06 |
| ENSG00000288524 | ENSG00000288524 | rs10190523 | 2 | 127400081 | 0.07 | 0.02 | G | T | -0.23 | 0.29 | NA | 0.05 | 0.18 | 0.04 | 6.39E-06 |
| ENSG00000137478 | FCHSD2 | rs6592500 | 11 | 72651705 | 0.55 | 0.77 | G | A | 0.78 | 0.28 | 1.17 | 0.19 | -0.07 | 0.01 | 7.59E-07 |
| ENSG00000104388 | RAB2A | rs11774081 | 8 | 61464732 | 0.54 | 0.39 | C | T | 0.21 | 0.28 | 0.73 | 0.06 | 0.07 | 0.01 | 2.10E-08 |

| | | | | | | | | | | | | | | | |
|---|---|---|---|---|---|---|---|---|---|---|---|---|---|---|---|
| ENSG00000288813 | ENSG00000288813 | rs2524094 | 6 | 31240041 | 0.52 | 0.57 | C | T | 1.41 | 0.28 | 0.98 | 0.05 | -0.08 | 0.02 | 4.90E-06 |
| ENSG00000223865 | HLA-DPB1 | rs928976 | 6 | 33049211 | 0.53 | 0.30 | C | T | -0.60 | 0.27 | 2.42 | 0.11 | 0.06 | 0.01 | 8.35E-06 |
| ENSG00000144802 | NFKBIZ | rs17347644 | 3 | 101547733 | 0.28 | 0.34 | C | T | 1.19 | 0.26 | 1.19 | 0.01 | -0.06 | 0.01 | 3.91E-06 |
| ENSG00000179151 | EDC3 | rs7497906 | 15 | 74952207 | 0.19 | 0.10 | G | A | -0.11 | 0.26 | 0.78 | 0.09 | 0.11 | 0.02 | 2.60E-06 |
| ENSG00000196189 | SEMA4A | rs12239871 | 1 | 156126617 | 0.15 | 0.03 | C | T | -0.27 | 0.25 | NA | 0.09 | 0.14 | 0.03 | 7.08E-06 |
| ENSG00000076662,ENSG00000274425 | ICAM3,ENSG00000274425 | rs2304237 | 19 | 10446568 | 0.18 | 0.22 | T | C | 0.85 | 0.24 | 0.13 | 0.02 | -0.08 | 0.02 | 1.36E-07 |
| ENSG00000204538,ENSG000 | PSORS1C2,PSORS1C1 | rs1265097 | 6 | 31106459 | 0.13 | 0.11 | C | A | 1.56 | 0.24 | 0.92 | 0.03 | 0.11 | 0.02 | 3.20E-07 |

| | | | | | | | | | | | | | | | |
|---|---|---|---|---|---|---|---|---|---|---|---|---|---|---|---|
| 00204540 | | | | | | | | | | | | | | | |
| ENSG00000256628 | ZBTB11-AS1 | rs3806654 | 3 | 101396529 | 0.41 | 0.35 | G | C | 0.06 | 0.22 | 0.47 | 0.05 | -0.06 | 0.01 | 8.34E-06 |
| ENSG00000088827 | SIGLEC1 | rs611847 | 20 | 3684022 | 0.61 | 0.66 | A | G | -0.05 | 0.19 | 0.15 | 0.01 | 0.06 | 0.01 | 1.80E-06 |
| ENSG00000204031 | LCN1P2 | rs72449624 | 9 | 136184771 | 0.10 | 0.11 | ACTG | A | 1.79 | 0.18 | 1.45 | 0.02 | 0.14 | 0.03 | 8.00E-06 |
| ENSG00000087903 | RFX2 | rs11085157 | 19 | 6128869 | 0.74 | 0.71 | T | C | -2.60 | 0.13 | 0.95 | 0.02 | 0.06 | 0.01 | 6.92E-06 |
| ENSG00000234261 | ENSG00000234261 | rs57696775 | 6 | 14714722 | 0.39 | 0.82 | C | CCTT | -1.58 | 0.10 | 0.75 | 0.48 | 0.14 | 0.02 | 1.47E-08 |
| ENSG00000169242 | EFNA1 | rs66474937 | 1 | 155101171 | 0.13 | 0.06 | C | T | 0.56 | 0.09 | 0.21 | 0.07 | -0.13 | 0.03 | 6.27E-07 |
| ENSG00000119866 | BCL11A | rs7565301 | 2 | 60723266 | 0.26 | 0.26 | G | A | 1.62 | 0.08 | 0.75 | 0.07 | -0.12 | 0.01 | 8.49E-15 |
| ENSG00000 | ENSG00000 | rs10774679 | 12 | 113374748 | 0.49 | 0.66 | C | T | 0.16 | 0.06 | 0.92 | 0.11 | 0.08 | 0.01 | 3.25E-09 |

| | | | | | | | | | | | | | | | |
|---|---|---|---|---|---|---|---|---|---|---|---|---|---|---|---|
| 257452 | 257452 | | | | | | | | | | | | | | |
| ENSG000001262144 | KLC1 | rs7148456 | 14 | 104028270 | 0.25 | 0.28 | T | C | -0.08 | 0.05 | 0.27 | 0.04 | 0.07 | 0.01 | 2.68E-06 |
| ENSG000001749511 | FUT1 | rs838136 | 19 | 49256388 | 0.50 | 0.31 | T | C | 0.22 | 0.05 | 0.95 | 0.07 | 0.07 | 0.01 | 1.29E-06 |
| ENSG000001633354 | DCST2 | rs4845703 | 1 | 154993375 | 0.68 | 0.69 | C | T | 0.27 | 0.05 | 0.98 | 0.09 | -0.07 | 0.01 | 4.54E-06 |
| ENSG000001404665 | CYP1A1 | rs4646421 | 15 | 75016192 | 0.25 | 0.12 | G | A | -0.30 | 0.03 | 1.48 | 0.08 | 0.08 | 0.02 | 5.93E-06 |
| ENSG000001644000 | CSF2 | rs25882 | 5 | 131411460 | 0.29 | 0.24 | T | C | -0.98 | 0.02 | 0.51 | 0.10 | 0.07 | 0.02 | 1.87E-06 |
| ENSG000001857600 | KCNQ5 | rs9351958 | 6 | 73661010 | 0.14 | 0.20 | G | C | -0.06 | 0.02 | 0.45 | 0.03 | 0.07 | 0.02 | 7.58E-06 |
| ENSG000001967355 | HLA-DQA1 | rs2133035 | 6 | 32596475 | 0.25 | 0.32 | C | A | 1.52 | 0.00 | 2.57 | 0.02 | -0.09 | 0.02 | 7.77E-07 |

**Supplementary Table 2. LD information for five eQTLs**

**LD in non-African populations (R2)**

| rsid | rs9832730 | rs3091310 | rs17283712 | rs17217831 | rs4987053 |
|---|---|---|---|---|---|
| rs9832730 | 1.00 | 0.98 | 0.98 | 0.99 | 0.94 |
| rs3091310 | 0.98 | 1.00 | 1.00 | 0.98 | 0.93 |
| rs17283712 | 0.98 | 1.00 | 1.00 | 0.98 | 0.93 |
| rs17217831 | 0.99 | 0.98 | 0.98 | 1.00 | 0.94 |
| rs4987053 | 0.94 | 0.93 | 0.93 | 0.94 | 1.00 |

**LD in African population (R2)**

| rsid | rs9832730 | rs3091310 | rs17283712 | rs17217831 | rs4987053 |
|---|---|---|---|---|---|
| rs9832730 | 1.00 | 0.05 | 0.05 | 0.96 | 0.27 |
| rs3091310 | 0.05 | 1.00 | 1.00 | 0.06 | 0.02 |
| rs17283712 | 0.05 | 1.00 | 1.00 | 0.06 | 0.02 |
| rs17217831 | 0.96 | 0.06 | 0.06 | 1.00 | 0.29 |
| rs4987053 | 0.27 | 0.02 | 0.02 | 0.29 | 1.00 |

**Supplementary Table 3. Blood *CCR3* expression from GTEx used for mendelian randomization analysis & colocalization**

| SNP | CHROM | POS | REF | ALT | version | tss_distance | N | maf | pval | beta | se |
|---|---|---|---|---|---|---|---|---|---|---|---|
| rs4339140 | 3 | 46053270 | G | A | b38 | -110334 | 199 | 0.16 | 7.33E-07 | -0.23 | 0.05 |
| rs13086717 | 3 | 46098007 | A | G | b38 | -65597 | 226 | 0.19 | 8.27E-07 | 0.21 | 0.04 |
| rs2201150 | 3 | 46217411 | T | C | b38 | 53807 | 407 | 0.37 | 3.33E-37 | 0.42 | 0.03 |
| rs9822457 | 3 | 46562461 | C | T | b38 | 398857 | 384 | 0.34 | 2.68E-07 | 0.19 | 0.04 |
| rs9853223 | 3 | 46242602 | G | A | b38 | 78998 | 455 | 0.43 | 1.97E-15 | 0.26 | 0.03 |

**Supplementary Table 4. Mendelian randomization result**

| outcome | exposure | nsnp | method | beta | se | pvalue | OR | lower | upper | R2 | F-statics | power |
|---|---|---|---|---|---|---|---|---|---|---|---|---|
| critically ill COVID-19 | blood CCR3 eQTL_GTEx | 5 | MR-PRESSO | -0.11 | 0.01 | 1.51E-02 | 0.89 | 0.87 | 0.92 | 0.16 | 61.25 | 0.81 |
| critically ill COVID-19 | blood CCR3 eQTL_GTEx | 5 | Weight median | -0.09 | 0.04 | 1.17E-02 | 0.91 | 0.85 | 0.98 | | | |
| critically ill COVID-19 | eosinophil counts | 1193 | MR-PRESSO | -0.06 | 0.03 | 4.94E-02 | 0.95 | 0.90 | 1.00 | 0.19 | 60936.24 | 0.99 |
| critically ill COVID-19 | eosinophil counts | 1193 | MR Egger | -0.15 | 0.08 | 4.47E-02 | 0.86 | 0.74 | 1.00 | | | |

**Supplementary Table 5. Ancient and present samples from AADR database**

| Version ID | Country | years before 1950 | ethnic |
|---|---|---|---|
| I0944 | Russia | 3820 | European |
| I3807 | Spain | 400 | European |
| I3981 | Spain | 1450 | European |
| I4055_published | Spain | 1650 | European |
| I4246_published | Spain | 4209 | European |
| I7186 | Czech Republic | 5713 | European |
| I7187 | Czech Republic | 6044 | European |
| I7188 | Czech Republic | 5610 | European |
| I7189 | Czech Republic | 5526 | European |
| I7191 | Czech Republic | 5657 | European |
| I7192 | Czech Republic | 5552 | European |
| I7193 | Czech Republic | 5621 | European |
| I7194 | Czech Republic | 5527 | European |
| I7207 | Czech Republic | 4370 | European |
| I7208 | Czech Republic | 4324 | European |
| I7209 | Czech Republic | 4269 | European |
| I7579 | United Kingdom | 3349 | European |
| I7580 | United Kingdom | 3087 | European |
| I7271 | Czech Republic | 4200 | European |
| I7278 | Czech Republic | 4333 | European |
| I7041 | Hungary | 4000 | European |
| I7043 | Hungary | 4011 | European |
| I7282 | Czech Republic | 4200 | European |
| I7283 | Czech Republic | 4200 | European |
| I7289 | Czech Republic | 4289 | European |
| I7210 | Czech Republic | 4071 | European |

| | | | |
|---|---|---|---|
| I7212 | Czech Republic | 4247 | European |
| I7214 | Czech Republic | 4200 | European |
| I6712 | Russia | 4625 | European |
| I6711 | Russia | 4750 | European |
| I6714 | Russia | 4479 | European |
| I6718 | Russia | 3493 | European |
| I3023 | United Kingdom | 5300 | European |
| I6695 | Czech Republic | 4575 | European |
| I6696 | Czech Republic | 4575 | European |
| I6713 | Russia | 4450 | European |
| I6715 | Russia | 4850 | European |
| I6716 | Russia | 3571 | European |
| I6717 | Russia | 3391 | European |
| I7040 | Hungary | 4300 | European |
| I7042 | Hungary | 6086 | European |
| I7044 | Hungary | 4300 | European |
| I7045 | Hungary | 4231 | European |
| I7195 | Czech Republic | 4250 | European |
| I7196 | Czech Republic | 4250 | European |
| I7197 | Czech Republic | 6438 | European |
| I7198 | Czech Republic | 4026 | European |
| I7199 | Czech Republic | 4039 | European |
| I7200 | Czech Republic | 4250 | European |
| I7201 | Czech Republic | 4250 | European |
| I7202 | Czech Republic | 4250 | European |
| I7203 | Czech Republic | 4250 | European |
| I7205 | Czech Republic | 4200 | European |
| I7211 | Czech Republic | 4183 | European |
| I7213 | Czech Republic | 4231 | European |

| | | | |
|---|---|---|---|
| I7249 | Czech Republic | 4011 | European |
| I7251 | Czech Republic | 4200 | European |
| I7250 | Czech Republic | 4200 | European |
| I7269 | Czech Republic | 4200 | European |
| I7272 | Czech Republic | 5417 | European |
| I7270 | Czech Republic | 4200 | European |
| I7275 | Czech Republic | 4200 | European |
| I7276 | Czech Republic | 4200 | European |
| I7279 | Czech Republic | 4200 | European |
| I7280 | Czech Republic | 4200 | European |
| I7281 | Czech Republic | 4194 | European |
| I7286 | Czech Republic | 4221 | European |
| I7287 | Czech Republic | 4200 | European |
| I7288 | Czech Republic | 4200 | European |
| I7290 | Czech Republic | 4200 | European |
| I7554 | United Kingdom | 5182 | European |
| I7568 | United Kingdom | 3400 | European |
| I7569 | United Kingdom | 3484 | European |
| I7570 | United Kingdom | 3569 | European |
| I7571 | United Kingdom | 3306 | European |
| I7572 | United Kingdom | 3380 | European |
| I7573 | United Kingdom | 3520 | European |
| I7574 | United Kingdom | 3271 | European |
| I7575 | United Kingdom | 3107 | European |
| I7576 | United Kingdom | 3039 | European |
| I7577 | United Kingdom | 3198 | European |
| I7578 | United Kingdom | 3080 | European |
| I7626 | United Kingdom | 3231 | European |
| I7627 | United Kingdom | 3268 | European |

| | | | |
|---|---|---|---|
| I7628 | United Kingdom | 3033 | European |
| I7630 | United Kingdom | 3734 | European |
| I7635 | United Kingdom | 4030 | European |
| I7638 | United Kingdom | 3995 | European |
| I7639 | United Kingdom | 3797 | European |
| I7640 | United Kingdom | 3307 | European |
| MA826.RISE00.SG | Estonia | 4442 | European |
| MA968.MA976.SG | Estonia | 4652 | European |
| RISE507.508.merge.SG | Russia | 5052 | European |
| Bon001.SG | Turkey | 10032 | European |
| Bon002.SG | Turkey | 10078 | European |
| Bon004.SG | Turkey | 10075 | European |
| Bon005.SG | Turkey | 10220 | European |
| Tep001.SG | Turkey | 8518 | European |
| Tep002.SG | Turkey | 8487 | European |
| Tep003.SG | Turkey | 8419 | European |
| Tep004.SG | Turkey | 8237 | European |
| Tep006.SG | Turkey | 8099 | European |
| A_Sardinian-4.DG | Italy | 0 | European |
| A_French-4.DG | France | 0 | European |
| S_Yakut-1.DG | Russia | 0 | European |
| S_Altaian-1.DG | Russia | 0 | European |
| S_Spanish-1.DG | Spain | 0 | European |
| S_Adygei-1.DG | Russia | 0 | European |
| S_French-1.DG | France | 0 | European |
| S_Bergamo-1.DG | Italy | 0 | European |
| S_Polish-1.DG | Poland | 0 | European |
| S_Itelman-1.DG | Russia | 0 | European |
| S_Tlingit-2.DG | Russia | 0 | European |

| | | | |
|---|---|---|---|
| S_Yakut-2.DG | Russia | 0 | European |
| S_Eskimo_Chaplin-1.DG | Russia | 0 | European |
| S_Tuscan-1.DG | Italy | 0 | European |
| S_Russian-2.DG | Russia | 0 | European |
| S_Hungarian-1.DG | Hungary | 0 | European |
| S_Abkhasian-1.DG | Russia | 0 | European |
| S_Eskimo_Sireniki-1.DG | Russia | 0 | European |
| S_Even-3.DG | Russia | 0 | European |
| S_Icelandic-2.DG | Iceland | 0 | European |
| S_Finnish-3.DG | Finland | 0 | European |
| B_Sardinian-3.DG | Italy | 0 | European |
| S_Mansi-2.DG | Russia | 0 | European |
| S_Eskimo_Naukan-2.DG | Russia | 0 | European |
| S_Even-1.DG | Russia | 0 | European |
| S_Czech-2.DG | Czech Republic | 0 | European |
| B_Crete-1.DG | Greece | 0 | European |
| S_Lezgin-2.DG | Russia | 0 | European |
| S_Greek-1.DG | Greece | 0 | European |
| S_Armenian-1.DG | Armenia | 0 | European |
| S_Estonian-2.DG | Estonia | 0 | European |
| S_English-1.DG | United Kingdom | 0 | European |
| S_Tubalar-2.DG | Russia | 0 | European |
| B_Crete-2.DG | Greece | 0 | European |
| S_Finnish-1.DG | Finland | 0 | European |
| S_Aleut-2.DG | Russia | 0 | European |
| S_Eskimo_Sireniki-2.DG | Russia | 0 | European |
| S_Sardinian-1.DG | Italy | 0 | European |
| S_Russian-1.DG | Russia | 0 | European |
| S_Tuscan-2.DG | Italy | 0 | European |

| | | | |
|---|---|---|---|
| S_Basque-2.DG | France | 0 | European |
| S_Orcadian-2.DG | United Kingdom | 0 | European |
| S_Finnish-2.DG | Finland | 0 | European |
| B_French-3.DG | France | 0 | European |
| S_Armenian-2.DG | Armenia | 0 | European |
| S_Mansi-1.DG | Russia | 0 | European |
| S_Eskimo_Naukan-1.DG | Russia | 0 | European |
| S_Turkish-2.DG | Turkey | 0 | European |
| S_Turkish-1.DG | Turkey | 0 | European |
| S_Spanish-2.DG | Spain | 0 | European |
| S_Aleut-1.DG | Russia | 0 | European |
| S_Greek-2.DG | Greece | 0 | European |
| S_North_Ossetian-1.DG | Russia | 0 | European |
| S_Orcadian-1.DG | United Kingdom | 0 | European |
| S_Basque-1.DG | France | 0 | European |
| S_Norwegian-1.DG | Norway | 0 | European |
| S_Ulchi-2.DG | Russia | 0 | European |
| S_Georgian-2.DG | Georgia | 0 | European |
| S_Sardinian-2.DG | Italy | 0 | European |
| S_Chechen-1.DG | Russia | 0 | European |
| S_Abkhasian-2.DG | Russia | 0 | European |
| S_Icelandic-1.DG | Iceland | 0 | European |
| S_Ulchi-1.DG | Russia | 0 | European |
| S_Saami-1.DG | Finland | 0 | European |
| S_Tubalar-1.DG | Russia | 0 | European |
| S_English-2.DG | United Kingdom | 0 | European |
| S_Saami-2.DG | Finland | 0 | European |
| S_Hungarian-2.DG | Hungary | 0 | European |
| S_Bulgarian-1.DG | Bulgaria | 0 | European |

| | | | |
|---|---|---|---|
| S_Tlingit-1.DG | Russia | 0 | European |
| S_North_Ossetian-2.DG | Russia | 0 | European |
| S_Adygei-2.DG | Russia | 0 | European |
| S_Bergamo-2.DG | Italy | 0 | European |
| S_French-2.DG | France | 0 | European |
| S_Bulgarian-2.DG | Bulgaria | 0 | European |
| S_Georgian-1.DG | Georgia | 0 | European |
| S_Chukchi-1.DG | Russia | 0 | European |
| S_Even-2.DG | Russia | 0 | European |
| S_Albanian-1.DG | Albania | 0 | European |
| Altai_snpAD.DG | Russia | 110450 | European |
| Denisova_snpAD.DG | Russia | 63900 | European |
| Loschbour_snpAD.DG | Luxembourg | 8025 | European |
| UstIshim_snpAD.DG | Russia | 44366 | European |
| Vindija_snpAD.DG | Croatia | 50000 | European |
| Altai_published.DG | Russia | 110450 | European |
| Denisova_published.DG | Russia | 63900 | European |
| Loschbour_published.DG | Luxembourg | 8025 | European |
| Stuttgart_published.DG | Germany | 7168 | European |
| Ust_Ishim_published.DG | Russia | 44366 | European |
| S_Estonian-1.DG | Estonia | 0 | European |
| S_Lezgin-1.DG | Russia | 0 | European |
| Avar.SG | Russia | 0 | European |
| Bur1.SG | Russia | 0 | European |
| Bur2.SG | Russia | 0 | European |
| Kor1.SG | Russia | 0 | European |
| Kor2.SG | Russia | 0 | European |
| Mari.SG | Russia | 0 | European |
| Bar31.SG | Turkey | 8272 | European |

| | | | |
|---|---|---|---|
| Bar8.SG | Turkey | 8064 | European |
| Klei10.SG | Greece | 6348 | European |
| Pal7.SG | Greece | 6348 | European |
| Rev5.SG | Greece | 8316 | European |
| BerryAuBac_published | France | 7237 | European |
| GoyetQ116-1_published | Belgium | 35208 | European |
| Paglicci108_published_d | Italy | 27900 | European |
| GoyetQ53-1_published_d | Belgium | 28051 | European |
| GoyetQ56-16_published_d | Belgium | 26300 | European |
| Paglicci133_published | Italy | 33150 | European |
| Rigney1_published_d | France | 15458 | European |
| Burkhardtshohle_published_d | Germany | 14620 | European |
| Rochedane_published | France | 13027 | European |
| Iboussieres39_published_d | France | 11741 | European |
| I0156.SG | United Kingdom | 1981 | European |
| I0157.SG | United Kingdom | 1227 | European |
| I0159.SG | United Kingdom | 1234 | European |
| I0160.SG | United Kingdom | 1973 | European |
| I0161.SG | United Kingdom | 1157 | European |
| I0769.SG | United Kingdom | 1449 | European |
| I0773.SG | United Kingdom | 1479 | European |
| I0774.SG | United Kingdom | 1469 | European |
| I0777.SG | United Kingdom | 1465 | European |
| I0789.SG | United Kingdom | 2163 | European |
| I2634 | United Kingdom | 5567 | European |
| I2636 | United Kingdom | 5397 | European |
| Falkenstein | Germany | 9161 | European |
| GoyetQ-2 | Belgium | 15077 | European |
| Iboussieres25-1 | France | 11675 | European |

| | | | |
|---|---|---|---|
| Iboussieres31-2 | France | 11675 | European |
| Rochedane | France | 13027 | European |
| BerryAuBac | France | 7237 | European |
| I0070 | Greece | 4000 | European |
| I0071 | Greece | 4000 | European |
| I0073 | Greece | 4000 | European |
| I0074 | Greece | 4000 | European |
| I9010 | Greece | 3250 | European |
| I9005 | Greece | 4000 | European |
| I9006 | Greece | 3283 | European |
| I9033 | Greece | 3296 | European |
| I9041 | Greece | 3250 | European |
| I9123 | Greece | 3305 | European |
| I9127 | Greece | 3895 | European |
| I9128 | Greece | 3895 | European |
| I9129 | Greece | 3895 | European |
| I9130 | Greece | 3895 | European |
| I9131 | Greece | 3895 | European |
| ANI152 | Bulgaria | 6462 | European |
| ANI153 | Bulgaria | 6434 | European |
| ANI160 | Bulgaria | 6533 | European |
| ANI163 | Bulgaria | 6569 | European |
| Bul10 | Bulgaria | 4951 | European |
| Bul4 | Bulgaria | 4895 | European |
| Bul6 | Bulgaria | 4450 | European |
| Bul8 | Bulgaria | 4450 | European |
| ANI159-ANI181 | Bulgaria | 6566 | European |
| I0001 | Luxembourg | 8025 | European |
| I2199 | Spain | 7069 | European |

| | | | |
|---|---|---|---|
| I2405_published | Poland | 5132 | European |
| I2416 | United Kingdom | 4239 | European |
| I2418 | United Kingdom | 4237 | European |
| I2421 | United Kingdom | 3790 | European |
| I2427 | Bulgaria | 6326 | European |
| I2431 | Bulgaria | 6624 | European |
| I2433 | Poland | 4950 | European |
| I2434 | Poland | 5050 | European |
| I2435_published | Poland | 4950 | European |
| I2440 | Poland | 4950 | European |
| I2441 | Poland | 5050 | European |
| I2457 | United Kingdom | 4056 | European |
| I2459 | United Kingdom | 4231 | European |
| I2460 | United Kingdom | 3875 | European |
| I2463 | United Kingdom | 3774 | European |
| I2467 | Spain | 4327 | European |
| I2472 | Spain | 3428 | European |
| I2478 | Italy | 4006 | European |
| I0234 | Russia | 3675 | European |
| I0126 | Russia | 4610 | European |
| I0440_published | Russia | 4720 | European |
| I0432 | Russia | 4695 | European |
| I0442 | Russia | 4350 | European |
| I0418_published | Russia | 3884 | European |
| I0434 | Russia | 6925 | European |
| I0433 | Russia | 6561 | European |
| I0122 | Russia | 6788 | European |
| I0124 | Russia | 7544 | European |
| I0233 | Russia | 3475 | European |

| | | | |
|---|---|---|---|
| I0370 | Russia | 4850 | European |
| I0441 | Russia | 4752 | European |
| I0444 | Russia | 5003 | European |
| I0439 | Russia | 5021 | European |
| I0357 | Russia | 4942 | European |
| I0429 | Russia | 5074 | European |
| I0438_published | Russia | 4785 | European |
| I0576 | Russia | 2750 | European |
| I0574 | Russia | 2250 | European |
| I0575 | Russia | 2250 | European |
| I1310 | Spain | 3650 | European |
| I1282 | Spain | 4550 | European |
| I1302 | Spain | 4550 | European |
| I1276 | Spain | 4550 | European |
| I1284 | Spain | 4550 | European |
| I1274 | Spain | 4550 | European |
| I1280 | Spain | 4550 | European |
| I1314 | Spain | 4550 | European |
| I1277 | Spain | 4403 | European |
| I1272 | Spain | 4595 | European |
| I1281 | Spain | 4653 | European |
| I1300 | Spain | 4550 | European |
| I0843 | Spain | 7858 | European |
| I0585 | Spain | 7811 | European |
| Oase1_d | Romania | 38033 | European |
| I0807 | Germany | 5799 | European |
| I0559 | Germany | 5530 | European |
| I0560 | Germany | 5471 | European |
| I0785 | Bulgaria | 6353 | European |

| | | | |
|---|---|---|---|
| I0781 | Bulgaria | 6404 | European |
| I1546 | Germany | 4225 | European |
| I0806 | Germany | 4217 | European |
| I0805 | Germany | 4252 | European |
| I0113 | Germany | 4146 | European |
| I0112 | Germany | 4224 | European |
| I1530 | Germany | 4222 | European |
| I0111 | Germany | 4303 | European |
| I0108 | Germany | 4399 | European |
| I0462 | Spain | 4405 | European |
| I0460 | Spain | 4287 | European |
| I0459 | Spain | 4350 | European |
| I0457 | Spain | 4500 | European |
| I0456 | Spain | 4500 | European |
| I0453 | Spain | 4239 | European |
| I0455 | Spain | 4737 | European |
| I0263 | Spain | 4500 | European |
| I1553 | Spain | 4500 | European |
| I0825 | Spain | 4346 | European |
| I0260 | Spain | 4500 | European |
| I0261 | Spain | 4500 | European |
| I0823 | Spain | 4500 | European |
| I0258 | Spain | 4500 | European |
| I0262 | Spain | 4500 | European |
| I0257 | Spain | 4439 | European |
| I0826 | Spain | 4522 | European |
| I0458 | Spain | 4270 | European |
| I0171 | Germany | 4118 | European |
| I0059 | Germany | 4179 | European |

| | | | |
|---|---|---|---|
| I1542 | Germany | 4225 | European |
| I1536 | Germany | 4225 | European |
| I1544 | Germany | 4225 | European |
| I1538 | Germany | 4225 | European |
| I1539 | Germany | 4420 | European |
| I1534_published | Germany | 4225 | European |
| I0106 | Germany | 4297 | European |
| I1540 | Germany | 4225 | European |
| I1541 | Germany | 4225 | European |
| I1532 | Germany | 4225 | European |
| I0049 | Germany | 4295 | European |
| I0211 | Russia | 8450 | European |
| I0061 | Russia | 8450 | European |
| I0550 | Germany | 4469 | European |
| I0797 | Germany | 7125 | European |
| I0795 | Germany | 7071 | European |
| I0176 | Hungary | 7002 | European |
| I0963 | Russia | 6950 | European |
| I0802 | Germany | 5163 | European |
| I0800 | Germany | 5164 | European |
| I0551 | Germany | 5163 | European |
| I0409 | Spain | 7210 | European |
| I0411 | Spain | 7107 | European |
| I0405 | Spain | 5700 | European |
| I0174 | Hungary | 7558 | European |
| I0115 | Germany | 3815 | European |
| I0117 | Germany | 4092 | European |
| I0804 | Germany | 4010 | European |
| I0803 | Germany | 3977 | European |

| | | | |
|---|---|---|---|
| I0164 | Germany | 3906 | European |
| I0114 | Germany | 4002 | European |
| I1060 | Russia | 3825 | European |
| I1062 | Russia | 3825 | European |
| I1065 | Russia | 3825 | European |
| I1022 | Russia | 3825 | European |
| I1088 | Russia | 3825 | European |
| I1084 | Russia | 3825 | European |
| I1082 | Russia | 3825 | European |
| I1055 | Russia | 3825 | European |
| I1018 | Russia | 3825 | European |
| I1017 | Russia | 3787 | European |
| I1019 | Russia | 3825 | European |
| I1086 | Russia | 3825 | European |
| I1090 | Russia | 3825 | European |
| I1024 | Russia | 3825 | European |
| I1063 | Russia | 3825 | European |
| I1058 | Russia | 3769 | European |
| I1029 | Russia | 3830 | European |
| I1020 | Russia | 3841 | European |
| I1028 | Russia | 3704 | European |
| I1064 | Russia | 3739 | European |
| I1089 | Russia | 3825 | European |
| I1054 | Russia | 3765 | European |
| I1053 | Russia | 3785 | European |
| I1027 | Russia | 3833 | European |
| I1056 | Russia | 3750 | European |
| I1057 | Russia | 3814 | European |
| I0022 | Germany | 7125 | European |

| | | | |
|---|---|---|---|
| I0026 | Germany | 7125 | European |
| I0013 | Sweden | 7722 | European |
| I0011 | Sweden | 7564 | European |
| I0015 | Sweden | 7710 | European |
| I0012 | Sweden | 7598 | European |
| I0014 | Sweden | 7686 | European |
| I0017 | Sweden | 7624 | European |
| I0447_published | Hungary | 6700 | European |
| I0449 | Hungary | 6700 | European |
| I1388 | France | 4198 | European |
| I1382 | France | 4195 | European |
| I1381 | France | 4250 | European |
| I1391_published | France | 4179 | European |
| I1389_published | France | 4361 | European |
| I1390 | France | 4331 | European |
| I1508 | Hungary | 7591 | European |
| I1500 | Hungary | 7058 | European |
| I1100 | Turkey | 8300 | European |
| I1102 | Turkey | 8300 | European |
| I1099 | Turkey | 8300 | European |
| I1103 | Turkey | 8300 | European |
| I1101 | Turkey | 8300 | European |
| I1097 | Turkey | 8282 | European |
| I0744 | Turkey | 8270 | European |
| I1096 | Turkey | 8300 | European |
| I1098 | Turkey | 8282 | European |
| I0708 | Turkey | 8101 | European |
| I0745 | Turkey | 8243 | European |
| I0746 | Turkey | 7928 | European |

| | | | |
|---|---|---|---|
| I0707 | Turkey | 8096 | European |
| I0709 | Turkey | 8085 | European |
| I0725 | Turkey | 7950 | European |
| I0736 | Turkey | 8300 | European |
| I1634 | Armenia | 6156 | European |
| I1632 | Armenia | 6076 | European |
| I1631 | Armenia | 6099 | European |
| I1635 | Armenia | 4476 | European |
| I1633 | Armenia | 4465 | European |
| I1658 | Armenia | 5162 | European |
| I1656 | Armenia | 3397 | European |
| I0633 | Serbia | 7424 | European |
| I0634 | Serbia | 6560 | European |
| I1131 | Serbia | 6482 | European |
| I1499 | Hungary | 7077 | European |
| I1409 | Armenia | 6047 | European |
| I1407 | Armenia | 5875 | European |
| I1502 | Hungary | 4026 | European |
| I1497 | Hungary | 5019 | European |
| I1495 | Hungary | 6367 | European |
| I1498 | Hungary | 7094 | European |
| I1507 | Hungary | 7661 | European |
| I1504 | Hungary | 2858 | European |
| I1506 | Hungary | 7146 | European |
| I1496 | Hungary | 7040 | European |
| I0676 | North Macedonia | 7792 | European |
| I1505 | Hungary | 7057 | European |
| I1736 | Ukraine | 8110 | European |
| I1734 | Ukraine | 8100 | European |

| | | | |
|---|---|---|---|
| I1738 | Ukraine | 7348 | European |
| I1732 | Ukraine | 7218 | European |
| I1193 | Russia | 6832 | European |
| I1190 | Russia | 6675 | European |
| I1196 | Russia | 7058 | European |
| I1197_published | Russia | 6804 | European |
| I1198 | Russia | 6872 | European |
| I1202 | Russia | 2850 | European |
| MA1.SG | Russia | 24320 | European |
| AfontovaGora2.SG | Russia | 16767 | European |
| Inuk.SG | Greenland | 3885 | European |
| Kostenki14.SG | Russia | 38052 | European |
| BR1.SG | Hungary | 4026 | European |
| BR2.SG | Hungary | 2858 | European |
| CO1.SG | Hungary | 5019 | European |
| IR1.SG | Hungary | 2849 | European |
| KO1_published.SG | Hungary | 7661 | European |
| KO2.SG | Hungary | 7591 | European |
| NE1.SG | Hungary | 7146 | European |
| NE2.SG | Hungary | 7094 | European |
| NE3.SG | Hungary | 7077 | European |
| NE4.SG | Hungary | 7057 | European |
| NE5.SG | Hungary | 7058 | European |
| NE7.SG | Hungary | 6367 | European |
| StoraForvar11.SG | Sweden | 7365 | European |
| Ajvide52.SG | Sweden | 4700 | European |
| Ajvide53.SG | Sweden | 4700 | European |
| Ajvide58.SG | Sweden | 4700 | European |
| Ajvide59.SG | Sweden | 4700 | European |

| | | | |
|---|---|---|---|
| Ajvide70.SG | Sweden | 4700 | European |
| Gokhem2.SG | Sweden | 4928 | European |
| Gokhem4.SG | Sweden | 4924 | European |
| Gokhem5.SG | Sweden | 5084 | European |
| Gokhem7.SG | Sweden | 4755 | European |
| Ire8.SG | Sweden | 4575 | European |
| Iceman.SG | Italy | 5199 | European |
| LaBrana1_published.SG | Spain | 7811 | European |
| I0061.SG | Russia | 8450 | European |
| RISE1.SG | Poland | 4663 | European |
| RISE21.SG | Denmark | 3301 | European |
| RISE42.SG | Denmark | 4021 | European |
| RISE47.SG | Denmark | 3380 | European |
| RISE61.SG | Denmark | 4569 | European |
| RISE71.SG | Denmark | 4040 | European |
| RISE94.SG | Sweden | 4487 | European |
| RISE97.SG | Sweden | 3895 | European |
| RISE98.SG | Sweden | 4082 | European |
| RISE109.SG | Poland | 3821 | European |
| RISE139.SG | Poland | 3970 | European |
| RISE145.SG | Poland | 4015 | European |
| RISE150.SG | Poland | 3744 | European |
| RISE154.SG | Poland | 3786 | European |
| RISE174.SG | Sweden | 1405 | European |
| RISE175.SG | Sweden | 3233 | European |
| RISE179.SG | Sweden | 3843 | European |
| RISE207.SG | Sweden | 3344 | European |
| RISE210.SG | Sweden | 3312 | European |
| RISE240.SG | Russia | 4705 | European |

| | | | |
|---|---|---|---|
| RISE247.SG | Hungary | 3607 | European |
| RISE254.SG | Hungary | 3948 | European |
| RISE276.SG | Denmark | 2612 | European |
| RISE349.SG | Hungary | 3893 | European |
| RISE371.SG | Hungary | 3982 | European |
| RISE373.SG | Hungary | 3751 | European |
| RISE374.SG | Hungary | 3643 | European |
| RISE386.SG | Russia | 4147 | European |
| RISE392.SG | Russia | 3943 | European |
| RISE394.SG | Russia | 3803 | European |
| RISE395.SG | Russia | 3815 | European |
| RISE396.SG | Armenia | 3009 | European |
| RISE397.SG | Armenia | 2908 | European |
| RISE407.SG | Armenia | 2935 | European |
| RISE408.SG | Armenia | 3050 | European |
| RISE412.SG | Armenia | 3018 | European |
| RISE413.SG | Armenia | 3764 | European |
| RISE416.SG | Armenia | 3477 | European |
| RISE423.SG | Armenia | 3251 | European |
| RISE431.SG | Poland | 4125 | European |
| RISE434.SG | Germany | 4703 | European |
| RISE435.SG | Germany | 4627 | European |
| RISE436.SG | Germany | 4671 | European |
| RISE446.SG | Germany | 4486 | European |
| RISE471.SG | Germany | 3543 | European |
| RISE479.SG | Hungary | 3700 | European |
| RISE480.SG | Hungary | 3550 | European |
| RISE483.SG | Hungary | 3700 | European |
| RISE484.SG | Hungary | 3700 | European |

| | | | |
|---|---|---|---|
| RISE486.SG | Italy | 3902 | European |
| RISE487.SG | Italy | 5207 | European |
| RISE489.SG | Italy | 4702 | European |
| RISE492.SG | Russia | 2248 | European |
| RISE493.SG | Russia | 3423 | European |
| RISE494.SG | Russia | 3293 | European |
| RISE495.SG | Russia | 3100 | European |
| RISE496.SG | Russia | 3285 | European |
| RISE497.SG | Russia | 3100 | European |
| RISE499.SG | Russia | 3100 | European |
| RISE500.SG | Russia | 3550 | European |
| RISE502.SG | Russia | 3360 | European |
| RISE503.SG | Russia | 3547 | European |
| RISE504.SG | Russia | 1126 | European |
| RISE505.SG | Russia | 3628 | European |
| RISE509.SG | Russia | 4726 | European |
| RISE510.SG | Russia | 4531 | European |
| RISE511.SG | Russia | 4757 | European |
| RISE512.SG | Russia | 3328 | European |
| RISE523.SG | Russia | 3411 | European |
| RISE524.SG | Russia | 3000 | European |
| RISE525.SG | Russia | 3000 | European |
| RISE546.SG | Russia | 4850 | European |
| RISE547.SG | Russia | 4713 | European |
| RISE548.SG | Russia | 4850 | European |
| RISE550.SG | Russia | 4916 | European |
| RISE552.SG | Russia | 4382 | European |
| RISE553.SG | Russia | 2819 | European |
| RISE554.SG | Russia | 2878 | European |

| | | | |
|---|---|---|---|
| RISE555.SG | Russia | 4598 | European |
| RISE559.SG | Germany | 4278 | European |
| RISE560.SG | Germany | 4200 | European |
| RISE562.SG | Germany | 4267 | European |
| RISE563.SG | Germany | 4411 | European |
| RISE564.SG | Germany | 4200 | European |
| RISE566.SG | Czech Republic | 4089 | European |
| RISE567.SG | Czech Republic | 4250 | European |
| RISE568.SG | Czech Republic | 1200 | European |
| RISE569.SG | Czech Republic | 1229 | European |
| RISE577.SG | Czech Republic | 4150 | European |
| RISE586.SG | Czech Republic | 4250 | European |
| RISE595.SG | Montenegro | 2950 | European |
| RISE596.SG | Montenegro | 2550 | European |
| RISE598.SG | Lithuania | 2653 | European |
| RISE600.SG | Russia | 2451 | European |
| RISE601.SG | Russia | 2451 | European |
| RISE602.SG | Russia | 2451 | European |
| I1206 | Russia | 6954 | European |
| I1271 | Spain | 4550 | European |
| I1303 | Spain | 4550 | European |
| I1549 | Germany | 4225 | European |
| I0407 | Spain | 5700 | European |
| I1560_d | Germany | 5350 | European |
| I0025 | Germany | 7125 | European |
| I0247 | Russia | 2229 | European |
| I0423 | Russia | 3475 | European |
| I0443 | Russia | 4850 | European |
| I0726 | Turkey | 7950 | European |

| | | | |
|---|---|---|---|
| I0103 | Germany | 4472 | European |
| I0172 | Germany | 5155 | European |
| I0408 | Spain | 5686 | European |
| I0412 | Spain | 7177 | European |
| I0054 | Germany | 7074 | European |
| I0104 | Germany | 4358 | European |
| I0116 | Germany | 3978 | European |
| I0118 | Germany | 4325 | European |
| I0406 | Spain | 5700 | European |
| I0410 | Spain | 7106 | European |
| I0413 | Spain | 7143 | European |
| NE6.SG | Hungary | 7040 | European |
| Villabruna | Italy | 14021 | European |
| Kostenki12 | Russia | 32650 | European |
| Vestonice16 | Czech Republic | 29800 | European |
| Kostenki14 | Russia | 38052 | European |
| Pavlov1_d | Czech Republic | 30350 | European |
| Muierii2_d | Romania | 33646 | European |
| Vestonice13_d | Czech Republic | 30800 | European |
| Vestonice15_d | Czech Republic | 30800 | European |
| Vestonice43_d | Czech Republic | 29800 | European |
| Ostuni2_d | Italy | 29116 | European |
| Ostuni1_d | Italy | 27614 | European |
| Continenza_d | Italy | 10875 | European |
| Vestonice14_d | Czech Republic | 30800 | European |
| ElMiron_d | Spain | 18775 | European |
| AfontovaGora3_d | Russia | 18036 | European |
| I1577 | Austria | 30950 | European |
| I1893 | Hungary | 6895 | European |

| | | | |
|---|---|---|---|
| I1904 | Hungary | 7010 | European |
| I1593 | Germany | 5527 | European |
| I1563 | Germany | 5448 | European |
| I1565 | Germany | 5674 | European |
| CB13.SG | Spain | 7345 | European |
| ATP3.SG | Spain | 5397 | European |
| ATP7.SG | Spain | 5122 | European |
| ATP9.SG | Spain | 3628 | European |
| ATP17.SG | Spain | 4849 | European |
| ATP20.SG | Spain | 4138 | European |
| I1876 | Hungary | 7541 | European |
| I1882 | Hungary | 7050 | European |
| I1883 | Hungary | 7146 | European |
| I1885 | Hungary | 7228 | European |
| I1917 | Ukraine | 4955 | European |
| I1926 | Ukraine | 5649 | European |
| I1977 | Spain | 3475 | European |
| I1981 | Spain | 4891 | European |
| I2069 | Russia | 5061 | European |
| I2071 | Russia | 4940 | European |
| I2105 | Ukraine | 4850 | European |
| I2110 | Ukraine | 5697 | European |
| I2111_published | Ukraine | 5622 | European |
| I2158 | Italy | 12000 | European |
| I2163 | Bulgaria | 3639 | European |
| I2165 | Bulgaria | 4910 | European |
| I2351 | Hungary | 6050 | European |
| I2352 | Hungary | 6550 | European |
| I2353 | Hungary | 6200 | European |

| | | | |
|---|---|---|---|
| I2354 | Hungary | 6200 | European |
| I2355 | Hungary | 7050 | European |
| I2356 | Hungary | 6200 | European |
| I2357 | Hungary | 7050 | European |
| I2358 | Hungary | 6700 | European |
| I2359 | Hungary | 6700 | European |
| I2364 | Hungary | 4220 | European |
| I2368 | Hungary | 5025 | European |
| I2369 | Hungary | 5186 | European |
| I2370 | Hungary | 5122 | European |
| I2371 | Hungary | 5169 | European |
| I2374 | Hungary | 7700 | European |
| I2375 | Hungary | 7050 | European |
| I2376 | Hungary | 7050 | European |
| I2377 | Hungary | 7001 | European |
| I2378 | Hungary | 7050 | European |
| I2379 | Hungary | 6984 | European |
| I2380 | Hungary | 7350 | European |
| I2382 | Hungary | 7350 | European |
| I2383 | Hungary | 7050 | European |
| I2387 | Hungary | 6700 | European |
| I2394 | Hungary | 6231 | European |
| I2395 | Hungary | 6200 | European |
| I0056 | Germany | 7059 | European |
| I0100 | Germany | 6944 | European |
| Bichon.SG | Switzerland | 13698 | European |
| KK1.SG | Georgia | 9678 | European |
| SATP.SG | Georgia | 13282 | European |
| kum4.SG | Turkey | 6550 | European |

| | | | |
|---|---|---|---|
| kum6.SG | Turkey | 5615 | European |
| I0017.SG | Sweden | 7624 | European |
| 3DT16.SG | United Kingdom | 1750 | European |
| 3DT26.SG | United Kingdom | 1750 | European |
| 6DT18.SG | United Kingdom | 1750 | European |
| 6DT21.SG | United Kingdom | 1750 | European |
| 6DT22.SG | United Kingdom | 1750 | European |
| 6DT23.SG | United Kingdom | 1750 | European |
| 6DT3.SG | United Kingdom | 1750 | European |
| M1489.SG | United Kingdom | 2035 | European |
| NO3423.SG | United Kingdom | 1200 | European |
| I0461 | Spain | 4241 | European |
| I2445 | United Kingdom | 4048 | European |
| I2452 | United Kingdom | 4052 | European |
| I2453 | United Kingdom | 4122 | European |
| I2454 | United Kingdom | 4042 | European |
| I2455 | United Kingdom | 3948 | European |
| I2373_all | Hungary | 7700 | European |
| I1877_all | Hungary | 7567 | European |
| I2483 | Austria | 30850 | European |
| I3354 | Russia | 7237 | European |
| I3358 | Russia | 769 | European |
| I3356 | Russia | 5605 | European |
| I3355 | Russia | 6821 | European |
| I3499 | Croatia | 4718 | European |
| I2014 | Germany | 6946 | European |
| I2020 | Germany | 7125 | European |
| I2022 | Germany | 7125 | European |
| I2980 | United Kingdom | 5173 | European |

| | | | |
|---|---|---|---|
| I2981 | United Kingdom | 3711 | European |
| I2988 | United Kingdom | 5397 | European |
| I3136 | United Kingdom | 5397 | European |
| I3137_published | United Kingdom | 5350 | European |
| I3138 | United Kingdom | 4998 | European |
| I3141_published | Ukraine | 4850 | European |
| I3151_published | Ukraine | 5675 | European |
| I3255 | United Kingdom | 3994 | European |
| I3270 | Spain | 4850 | European |
| I3271 | Spain | 4850 | European |
| I3272 | Spain | 4959 | European |
| I3313 | Croatia | 2722 | European |
| I3322 | Spain | 2500 | European |
| I3324 | Spain | 2223 | European |
| I3388_published | Russia | 4650 | European |
| I3390 | Russia | 3600 | European |
| I3395 | Russia | 3600 | European |
| I3433 | Croatia | 7800 | European |
| I3485 | Spain | 4050 | European |
| I3486 | Spain | 3575 | European |
| I3487 | Spain | 3603 | European |
| I3488 | Spain | 3575 | European |
| I3490 | Spain | 3450 | European |
| I3491 | Spain | 3450 | European |
| I3493 | Spain | 3301 | European |
| I3498 | Croatia | 7681 | European |
| I3528 | Hungary | 4364 | European |
| I3529 | Hungary | 4300 | European |
| I3574 | Spain | 1450 | European |

| | | | |
|---|---|---|---|
| I3575 | Spain | 1450 | European |
| I3577 | Spain | 1450 | European |
| I3578 | Spain | 1450 | European |
| I3579 | Spain | 1450 | European |
| I3581 | Spain | 1450 | European |
| I3599_published | Germany | 4175 | European |
| I3600 | Germany | 4175 | European |
| I3601 | Germany | 4175 | European |
| I3602_published | Germany | 4175 | European |
| I3604_published | Germany | 4200 | European |
| I3607_published | Germany | 4200 | European |
| I3642 | Italy | 2994 | European |
| I3775 | Spain | 1300 | European |
| I3776 | Spain | 1300 | European |
| I3777 | Spain | 1300 | European |
| I3778 | Spain | 1300 | European |
| I3808 | Spain | 400 | European |
| I3810 | Spain | 400 | European |
| I3866 | Spain | 1300 | European |
| I3874 | France | 4065 | European |
| I3875 | France | 3985 | European |
| I3879 | Bulgaria | 7550 | European |
| I3951 | Russia | 4711 | European |
| I3269 | Spain | 4850 | European |
| I2977 | United Kingdom | 4843 | European |
| I2978 | United Kingdom | 5140 | European |
| I3133 | United Kingdom | 5446 | European |
| I3256 | United Kingdom | 4063 | European |
| I2979 | United Kingdom | 5104 | European |

| | | | |
|---|---|---|---|
| I3134 | United Kingdom | 5461 | European |
| I3135 | United Kingdom | 5517 | European |
| I3132 | United Kingdom | 3927 | European |
| I3389 | Russia | 3600 | European |
| I3392 | Russia | 3600 | European |
| I3394 | Russia | 3600 | European |
| I3396 | Russia | 3600 | European |
| I3576 | Spain | 1468 | European |
| I3949 | Russia | 4569 | European |
| I3947 | Croatia | 7834 | European |
| I3948 | Croatia | 7863 | European |
| I3950 | Russia | 4707 | European |
| I3315_published | Spain | 2809 | European |
| I3876 | Italy | 2928 | European |
| I3391 | Russia | 3600 | European |
| I3393 | Russia | 3600 | European |
| I3494 | Spain | 3781 | European |
| I3068 | United Kingdom | 5533 | European |
| I2365 | Hungary | 4299 | European |
| I2403 | Poland | 4667 | European |
| I3323 | Spain | 2226 | European |
| I3535 | Hungary | 7068 | European |
| I3536 | Hungary | 7050 | European |
| I3537 | Hungary | 7366 | European |
| I3582 | Spain | 1450 | European |
| I3583_published | Spain | 1450 | European |
| I3585 | Spain | 1195 | European |
| I3587_published | Germany | 4175 | European |
| I3588 | Germany | 4175 | European |

| | | | |
|---|---|---|---|
| I3589 | Germany | 4175 | European |
| I3590 | Germany | 4186 | European |
| I3594 | Germany | 4175 | European |
| I3714 | Ukraine | 7100 | European |
| I3715 | Ukraine | 7526 | European |
| I3717 | Ukraine | 7242 | European |
| I3741 | Italy | 3088 | European |
| I3743 | Italy | 3985 | European |
| I3756 | Spain | 3860 | European |
| I3759 | Spain | 2222 | European |
| I3954 | Russia | 4690 | European |
| I3980 | Spain | 1387 | European |
| I3983 | Spain | 1550 | European |
| I4054 | Spain | 1650 | European |
| I4067_published | Netherlands | 3770 | European |
| I4068 | Netherlands | 3985 | European |
| I4069 | Netherlands | 3966 | European |
| I4070 | Netherlands | 3703 | European |
| I4071 | Netherlands | 3717 | European |
| I4073 | Netherlands | 3992 | European |
| I4075 | Netherlands | 3950 | European |
| I4076 | Netherlands | 3763 | European |
| I4088 | Romania | 6191 | European |
| I4089 | Romania | 5636 | European |
| I4109 | Italy | 3516 | European |
| I4110 | Ukraine | 5394 | European |
| I4111 | Ukraine | 6593 | European |
| I4112_published | Ukraine | 6950 | European |
| I4114 | Ukraine | 7350 | European |

| | | | |
|---|---|---|---|
| I4167 | Croatia | 6635 | European |
| I4168 | Croatia | 6588 | European |
| I4175 | Croatia | 4800 | European |
| I4178 | Hungary | 4300 | European |
| I4181 | Hungary | 7050 | European |
| I4183 | Hungary | 6743 | European |
| I4184 | Hungary | 6754 | European |
| I4185 | Hungary | 6873 | European |
| I4186 | Hungary | 7050 | European |
| I4187 | Hungary | 7050 | European |
| I4188_published | Hungary | 7050 | European |
| I4189 | Hungary | 6050 | European |
| I4196 | Hungary | 7050 | European |
| I4199 | Hungary | 7050 | European |
| I4247 | Spain | 4302 | European |
| I4248_published | Germany | 4200 | European |
| I4249_published | Germany | 4186 | European |
| I4250_published | Germany | 4220 | European |
| I4251_published | Poland | 4219 | European |
| I4252_published | Poland | 4155 | European |
| I4253 | Poland | 4266 | European |
| I0577 | Russia | 2750 | European |
| I1594 | Germany | 5140 | European |
| I1763 | Ukraine | 10081 | European |
| I1767 | United Kingdom | 4045 | European |
| I1770 | United Kingdom | 3950 | European |
| I1775 | United Kingdom | 3563 | European |
| I1819 | Ukraine | 10658 | European |
| I1821 | Russia | 3490 | European |

| | | | |
|---|---|---|---|
| I1828 | Russia | 3484 | European |
| I1829 | Russia | 4879 | European |
| I1836 | Spain | 3527 | European |
| I1838 | Spain | 5126 | European |
| I1840 | Spain | 3475 | European |
| I1843 | Spain | 4904 | European |
| I1851 | Russia | 3463 | European |
| I1852 | Russia | 3512 | European |
| I1853 | Russia | 3484 | European |
| I1856 | Russia | 3540 | European |
| I1887 | Hungary | 7150 | European |
| I1889 | Hungary | 6943 | European |
| I1890 | Hungary | 6875 | European |
| I1891 | Hungary | 6913 | European |
| I1894 | Hungary | 7150 | European |
| I1896 | Hungary | 7190 | European |
| I1899 | Hungary | 6550 | European |
| I1900 | Hungary | 6686 | European |
| I1901 | Hungary | 6550 | European |
| I1903 | Hungary | 6550 | European |
| I1906 | Hungary | 6550 | European |
| I1908 | Hungary | 6050 | European |
| I1909 | Hungary | 6086 | European |
| I1958 | Russia | 6606 | European |
| I1960 | Russia | 8116 | European |
| I1972 | Spain | 6703 | European |
| I1975 | Spain | 4927 | European |
| I2464 | United Kingdom | 3615 | European |
| I2509 | Bulgaria | 6350 | European |

| | | | |
|---|---|---|---|
| I2510 | Bulgaria | 4794 | European |
| I2519 | Bulgaria | 6237 | European |
| I2520 | Bulgaria | 5153 | European |
| I2532 | Romania | 7614 | European |
| I2533 | Romania | 7371 | European |
| I2534 | Romania | 7670 | European |
| I2565_published | United Kingdom | 4237 | European |
| I2566 | United Kingdom | 4078 | European |
| I2567 | United Kingdom | 4018 | European |
| I2568 | United Kingdom | 4113 | European |
| I2569_published | United Kingdom | 3978 | European |
| I2573 | United Kingdom | 3361 | European |
| I2574_published | United Kingdom | 3277 | European |
| I2575_published | France | 4323 | European |
| I2596 | United Kingdom | 4086 | European |
| I2598_published | United Kingdom | 3998 | European |
| I2600 | United Kingdom | 3970 | European |
| I2601 | United Kingdom | 3808 | European |
| I2602 | United Kingdom | 3762 | European |
| I2605 | United Kingdom | 5397 | European |
| I2609 | United Kingdom | 3848 | European |
| I2610 | United Kingdom | 3783 | European |
| I2612 | United Kingdom | 4293 | European |
| I2618 | United Kingdom | 3993 | European |
| I2629_published | United Kingdom | 4800 | European |
| I2630 | United Kingdom | 4471 | European |
| I2631 | United Kingdom | 4957 | European |
| I2633 | United Kingdom | 5639 | European |
| I2635 | United Kingdom | 5523 | European |

| | | | |
|---|---|---|---|
| I2637 | United Kingdom | 5375 | European |
| I2639 | United Kingdom | 3430 | European |
| I2650 | United Kingdom | 5380 | European |
| I2651 | United Kingdom | 5160 | European |
| I2653 | United Kingdom | 3224 | European |
| I2654 | United Kingdom | 3287 | European |
| I2655 | United Kingdom | 3311 | European |
| I2659 | United Kingdom | 5637 | European |
| I2660 | United Kingdom | 5394 | European |
| I2691 | United Kingdom | 5613 | European |
| I2741 | Hungary | 4245 | European |
| I2743 | Hungary | 7050 | European |
| I2744 | Hungary | 7050 | European |
| I2745 | Hungary | 7050 | European |
| I2746 | Hungary | 6700 | European |
| I2752 | Hungary | 5175 | European |
| I2753 | Hungary | 5085 | European |
| I2754 | Hungary | 5140 | European |
| I2755 | Hungary | 5175 | European |
| I2763 | Hungary | 5172 | European |
| I2783 | Hungary | 6010 | European |
| I2785 | Hungary | 5175 | European |
| I2787 | Hungary | 4254 | European |
| I2788 | Hungary | 5685 | European |
| I2789 | Hungary | 5650 | European |
| I2796_published | United Kingdom | 5571 | European |
| I2859 | United Kingdom | 2810 | European |
| I2860 | United Kingdom | 2824 | European |
| I2861 | United Kingdom | 2843 | European |

| | | | |
|---|---|---|---|
| I2932 | United Kingdom | 4433 | European |
| I2933 | United Kingdom | 4877 | European |
| I2934 | United Kingdom | 5138 | European |
| I2935 | United Kingdom | 5115 | European |
| I3005 | United Kingdom | 5657 | European |
| I3033 | United Kingdom | 5533 | European |
| I3039 | United Kingdom | 5525 | European |
| I3040 | United Kingdom | 5525 | European |
| I3041 | United Kingdom | 5440 | European |
| I3065 | United Kingdom | 6290 | European |
| I3085 | United Kingdom | 5149 | European |
| I3130 | United Kingdom | 2844 | European |
| I1765_d | United Kingdom | 4156 | European |
| I2656_published_d | United Kingdom | 3081 | European |
| I3025 | United Kingdom | 9257 | European |
| I3592 | Germany | 4260 | European |
| I3593_published | Germany | 4207 | European |
| I3596_published | Germany | 4175 | European |
| I3597 | Germany | 4175 | European |
| I3712_published | Ukraine | 7377 | European |
| I3713_published | Ukraine | 6942 | European |
| I3716_published | Ukraine | 7347 | European |
| DevilsGate2.SG | Russia | 7620 | European |
| DevilsGate1.SG | Russia | 7620 | European |
| Ukraine_HG1.SG | Ukraine | 10658 | European |
| Ukraine_N1.SG | Ukraine | 7300 | European |
| Latvia_HG2.SG | Latvia | 7677 | European |
| Latvia_MN1.SG | Latvia | 6071 | European |
| Latvia_HG3.SG | Latvia | 7033 | European |

| | | | |
|---|---|---|---|
| Latvia_LN1.SG | Latvia | 4842 | European |
| Latvia_MN2.SG | Latvia | 5997 | European |
| Latvia_HG1.SG | Latvia | 8316 | European |
| I3952 | Russia | 4667 | European |
| I4074 | Netherlands | 4030 | European |
| I4081_published | Romania | 9361 | European |
| I4303 | France | 6624 | European |
| I4331 | Croatia | 3519 | European |
| I4332 | Croatia | 3508 | European |
| I4432 | Latvia | 7958 | European |
| I4434 | Latvia | 7441 | European |
| I4435 | Latvia | 5972 | European |
| I4436 | Latvia | 6109 | European |
| I4437 | Latvia | 6164 | European |
| I4438 | Latvia | 7248 | European |
| I4439 | Latvia | 7646 | European |
| I4440 | Latvia | 7074 | European |
| I4441 | Latvia | 6718 | European |
| I4550 | Latvia | 8489 | European |
| I4551 | Latvia | 7670 | European |
| I4552 | Latvia | 7369 | European |
| I4553 | Latvia | 7370 | European |
| I4554 | Latvia | 5946 | European |
| I4562 | Spain | 3614 | European |
| I4563 | Spain | 3550 | European |
| I4565 | Spain | 4915 | European |
| I4626 | Latvia | 7677 | European |
| I4627 | Latvia | 6071 | European |
| I4628 | Latvia | 7033 | European |

| | | | |
|---|---|---|---|
| I4629 | Latvia | 4842 | European |
| I4655_published | Romania | 8764 | European |
| I4657_published | Serbia | 11465 | European |
| I4665_published | Serbia | 8006 | European |
| I4666_published | Serbia | 8017 | European |
| HG00096.SG | United Kingdom | 0 | European |
| HG00097.SG | United Kingdom | 0 | European |
| HG00099.SG | United Kingdom | 0 | European |
| HG00100.SG | United Kingdom | 0 | European |
| HG00101.SG | United Kingdom | 0 | European |
| HG00102.SG | United Kingdom | 0 | European |
| HG00103.SG | United Kingdom | 0 | European |
| HG00105.SG | United Kingdom | 0 | European |
| HG00106.SG | United Kingdom | 0 | European |
| HG00107.SG | United Kingdom | 0 | European |
| HG00108.SG | United Kingdom | 0 | European |
| HG00109.SG | United Kingdom | 0 | European |
| HG00110.SG | United Kingdom | 0 | European |
| HG00111.SG | United Kingdom | 0 | European |
| HG00112.SG | United Kingdom | 0 | European |
| HG00113.SG | United Kingdom | 0 | European |
| HG00114.SG | United Kingdom | 0 | European |
| HG00115.SG | United Kingdom | 0 | European |
| HG00116.SG | United Kingdom | 0 | European |
| HG00117.SG | United Kingdom | 0 | European |
| HG00118.SG | United Kingdom | 0 | European |
| HG00119.SG | United Kingdom | 0 | European |
| HG00120.SG | United Kingdom | 0 | European |
| HG00121.SG | United Kingdom | 0 | European |

| | | | |
|---|---|---|---|
| HG00122.SG | United Kingdom | 0 | European |
| HG00123.SG | United Kingdom | 0 | European |
| HG00124.SG | United Kingdom | 0 | European |
| HG00125.SG | United Kingdom | 0 | European |
| HG00126.SG | United Kingdom | 0 | European |
| HG00127.SG | United Kingdom | 0 | European |
| HG00128.SG | United Kingdom | 0 | European |
| HG00129.SG | United Kingdom | 0 | European |
| HG00130.SG | United Kingdom | 0 | European |
| HG00131.SG | United Kingdom | 0 | European |
| HG00132.SG | United Kingdom | 0 | European |
| HG00133.SG | United Kingdom | 0 | European |
| HG00136.SG | United Kingdom | 0 | European |
| HG00137.SG | United Kingdom | 0 | European |
| HG00138.SG | United Kingdom | 0 | European |
| HG00139.SG | United Kingdom | 0 | European |
| HG00140.SG | United Kingdom | 0 | European |
| HG00141.SG | United Kingdom | 0 | European |
| HG00142.SG | United Kingdom | 0 | European |
| HG00143.SG | United Kingdom | 0 | European |
| HG00145.SG | United Kingdom | 0 | European |
| HG00146.SG | United Kingdom | 0 | European |
| HG00148.SG | United Kingdom | 0 | European |
| HG00149.SG | United Kingdom | 0 | European |
| HG00150.SG | United Kingdom | 0 | European |
| HG00151.SG | United Kingdom | 0 | European |
| HG00154.SG | United Kingdom | 0 | European |
| HG00155.SG | United Kingdom | 0 | European |
| HG00157.SG | United Kingdom | 0 | European |

| | | | |
|---|---|---|---|
| HG00158.SG | United Kingdom | 0 | European |
| HG00159.SG | United Kingdom | 0 | European |
| HG00160.SG | United Kingdom | 0 | European |
| HG00171.SG | Finland | 0 | European |
| HG00173.SG | Finland | 0 | European |
| HG00174.SG | Finland | 0 | European |
| HG00176.SG | Finland | 0 | European |
| HG00177.SG | Finland | 0 | European |
| HG00178.SG | Finland | 0 | European |
| HG00179.SG | Finland | 0 | European |
| HG00180.SG | Finland | 0 | European |
| HG00181.SG | Finland | 0 | European |
| HG00182.SG | Finland | 0 | European |
| HG00183.SG | Finland | 0 | European |
| HG00185.SG | Finland | 0 | European |
| HG00186.SG | Finland | 0 | European |
| HG00187.SG | Finland | 0 | European |
| HG00188.SG | Finland | 0 | European |
| HG00189.SG | Finland | 0 | European |
| HG00190.SG | Finland | 0 | European |
| HG00231.SG | United Kingdom | 0 | European |
| HG00232.SG | United Kingdom | 0 | European |
| HG00233.SG | United Kingdom | 0 | European |
| HG00234.SG | United Kingdom | 0 | European |
| HG00235.SG | United Kingdom | 0 | European |
| HG00236.SG | United Kingdom | 0 | European |
| HG00237.SG | United Kingdom | 0 | European |
| HG00238.SG | United Kingdom | 0 | European |
| HG00239.SG | United Kingdom | 0 | European |

| | | | |
|---|---|---|---|
| HG00240.SG | United Kingdom | 0 | European |
| HG00242.SG | United Kingdom | 0 | European |
| HG00243.SG | United Kingdom | 0 | European |
| HG00244.SG | United Kingdom | 0 | European |
| HG00245.SG | United Kingdom | 0 | European |
| HG00246.SG | United Kingdom | 0 | European |
| HG00250.SG | United Kingdom | 0 | European |
| HG00251.SG | United Kingdom | 0 | European |
| HG00252.SG | United Kingdom | 0 | European |
| HG00253.SG | United Kingdom | 0 | European |
| HG00254.SG | United Kingdom | 0 | European |
| HG00255.SG | United Kingdom | 0 | European |
| HG00256.SG | United Kingdom | 0 | European |
| HG00257.SG | United Kingdom | 0 | European |
| HG00258.SG | United Kingdom | 0 | European |
| HG00259.SG | United Kingdom | 0 | European |
| HG00260.SG | United Kingdom | 0 | European |
| HG00261.SG | United Kingdom | 0 | European |
| HG00262.SG | United Kingdom | 0 | European |
| HG00263.SG | United Kingdom | 0 | European |
| HG00264.SG | United Kingdom | 0 | European |
| HG00265.SG | United Kingdom | 0 | European |
| HG00266.SG | Finland | 0 | European |
| HG00267.SG | Finland | 0 | European |
| HG00268.SG | Finland | 0 | European |
| HG00269.SG | Finland | 0 | European |
| HG00271.SG | Finland | 0 | European |
| HG00272.SG | Finland | 0 | European |
| HG00273.SG | Finland | 0 | European |

| | | | |
|---|---|---|---|
| HG00274.SG | Finland | 0 | European |
| HG00275.SG | Finland | 0 | European |
| HG00276.SG | Finland | 0 | European |
| HG00277.SG | Finland | 0 | European |
| HG00278.SG | Finland | 0 | European |
| HG00280.SG | Finland | 0 | European |
| HG00281.SG | Finland | 0 | European |
| HG00282.SG | Finland | 0 | European |
| HG00284.SG | Finland | 0 | European |
| HG00285.SG | Finland | 0 | European |
| HG00288.SG | Finland | 0 | European |
| HG00290.SG | Finland | 0 | European |
| HG00304.SG | Finland | 0 | European |
| HG00306.SG | Finland | 0 | European |
| HG00308.SG | Finland | 0 | European |
| HG00309.SG | Finland | 0 | European |
| HG00310.SG | Finland | 0 | European |
| HG00311.SG | Finland | 0 | European |
| HG00313.SG | Finland | 0 | European |
| HG00315.SG | Finland | 0 | European |
| HG00318.SG | Finland | 0 | European |
| HG00319.SG | Finland | 0 | European |
| HG00320.SG | Finland | 0 | European |
| HG00321.SG | Finland | 0 | European |
| HG00323.SG | Finland | 0 | European |
| HG00324.SG | Finland | 0 | European |
| HG00325.SG | Finland | 0 | European |
| HG00326.SG | Finland | 0 | European |
| HG00327.SG | Finland | 0 | European |

| | | | |
|---|---|---|---|
| HG00328.SG | Finland | 0 | European |
| HG00329.SG | Finland | 0 | European |
| HG00330.SG | Finland | 0 | European |
| HG00331.SG | Finland | 0 | European |
| HG00332.SG | Finland | 0 | European |
| HG00334.SG | Finland | 0 | European |
| HG00335.SG | Finland | 0 | European |
| HG00336.SG | Finland | 0 | European |
| HG00337.SG | Finland | 0 | European |
| HG00338.SG | Finland | 0 | European |
| HG00339.SG | Finland | 0 | European |
| HG00341.SG | Finland | 0 | European |
| HG00342.SG | Finland | 0 | European |
| HG00343.SG | Finland | 0 | European |
| HG00344.SG | Finland | 0 | European |
| HG00345.SG | Finland | 0 | European |
| HG00346.SG | Finland | 0 | European |
| HG00349.SG | Finland | 0 | European |
| HG00350.SG | Finland | 0 | European |
| HG00351.SG | Finland | 0 | European |
| HG00353.SG | Finland | 0 | European |
| HG00355.SG | Finland | 0 | European |
| HG00356.SG | Finland | 0 | European |
| HG00357.SG | Finland | 0 | European |
| HG00358.SG | Finland | 0 | European |
| HG00360.SG | Finland | 0 | European |
| HG00361.SG | Finland | 0 | European |
| HG00362.SG | Finland | 0 | European |
| HG00364.SG | Finland | 0 | European |

| | | | |
|---|---|---|---|
| HG00365.SG | Finland | 0 | European |
| HG00366.SG | Finland | 0 | European |
| HG00367.SG | Finland | 0 | European |
| HG00368.SG | Finland | 0 | European |
| HG00369.SG | Finland | 0 | European |
| HG00371.SG | Finland | 0 | European |
| HG00372.SG | Finland | 0 | European |
| HG00373.SG | Finland | 0 | European |
| HG00375.SG | Finland | 0 | European |
| HG00376.SG | Finland | 0 | European |
| HG00378.SG | Finland | 0 | European |
| HG00379.SG | Finland | 0 | European |
| HG00380.SG | Finland | 0 | European |
| HG00381.SG | Finland | 0 | European |
| HG00382.SG | Finland | 0 | European |
| HG00383.SG | Finland | 0 | European |
| HG00384.SG | Finland | 0 | European |
| HG01334.SG | United Kingdom | 0 | European |
| HG01500.SG | Spain | 0 | European |
| HG01501.SG | Spain | 0 | European |
| HG01503.SG | Spain | 0 | European |
| HG01504.SG | Spain | 0 | European |
| HG01506.SG | Spain | 0 | European |
| HG01507.SG | Spain | 0 | European |
| HG01509.SG | Spain | 0 | European |
| HG01510.SG | Spain | 0 | European |
| HG01512.SG | Spain | 0 | European |
| HG01513.SG | Spain | 0 | European |
| HG01515.SG | Spain | 0 | European |

| | | | |
|---|---|---|---|
| HG01516.SG | Spain | 0 | European |
| HG01518.SG | Spain | 0 | European |
| HG01519.SG | Spain | 0 | European |
| HG01521.SG | Spain | 0 | European |
| HG01522.SG | Spain | 0 | European |
| HG01524.SG | Spain | 0 | European |
| HG01525.SG | Spain | 0 | European |
| HG01527.SG | Spain | 0 | European |
| HG01528.SG | Spain | 0 | European |
| HG01530.SG | Spain | 0 | European |
| HG01531.SG | Spain | 0 | European |
| HG01536.SG | Spain | 0 | European |
| HG01537.SG | Spain | 0 | European |
| HG01602.SG | Spain | 0 | European |
| HG01603.SG | Spain | 0 | European |
| HG01605.SG | Spain | 0 | European |
| HG01606.SG | Spain | 0 | European |
| HG01607.SG | Spain | 0 | European |
| HG01608.SG | Spain | 0 | European |
| HG01610.SG | Spain | 0 | European |
| HG01612.SG | Spain | 0 | European |
| HG01613.SG | Spain | 0 | European |
| HG01615.SG | Spain | 0 | European |
| HG01617.SG | Spain | 0 | European |
| HG01618.SG | Spain | 0 | European |
| HG01619.SG | Spain | 0 | European |
| HG01620.SG | Spain | 0 | European |
| HG01623.SG | Spain | 0 | European |
| HG01624.SG | Spain | 0 | European |

| | | | |
|---|---|---|---|
| HG01625.SG | Spain | 0 | European |
| HG01626.SG | Spain | 0 | European |
| HG01628.SG | Spain | 0 | European |
| HG01630.SG | Spain | 0 | European |
| HG01631.SG | Spain | 0 | European |
| HG01632.SG | Spain | 0 | European |
| HG01668.SG | Spain | 0 | European |
| HG01669.SG | Spain | 0 | European |
| HG01670.SG | Spain | 0 | European |
| HG01672.SG | Spain | 0 | European |
| HG01673.SG | Spain | 0 | European |
| HG01675.SG | Spain | 0 | European |
| HG01676.SG | Spain | 0 | European |
| HG01678.SG | Spain | 0 | European |
| HG01679.SG | Spain | 0 | European |
| HG01680.SG | Spain | 0 | European |
| HG01682.SG | Spain | 0 | European |
| HG01684.SG | Spain | 0 | European |
| HG01685.SG | Spain | 0 | European |
| HG01686.SG | Spain | 0 | European |
| HG01694.SG | Spain | 0 | European |
| HG01695.SG | Spain | 0 | European |
| HG01697.SG | Spain | 0 | European |
| HG01699.SG | Spain | 0 | European |
| HG01700.SG | Spain | 0 | European |
| HG01702.SG | Spain | 0 | European |
| HG01704.SG | Spain | 0 | European |
| HG01705.SG | Spain | 0 | European |
| HG01707.SG | Spain | 0 | European |

| | | | |
|---|---|---|---|
| HG01708.SG | Spain | 0 | European |
| HG01709.SG | Spain | 0 | European |
| HG01710.SG | Spain | 0 | European |
| HG01746.SG | Spain | 0 | European |
| HG01747.SG | Spain | 0 | European |
| HG01756.SG | Spain | 0 | European |
| HG01757.SG | Spain | 0 | European |
| HG01761.SG | Spain | 0 | European |
| HG01762.SG | Spain | 0 | European |
| HG01765.SG | Spain | 0 | European |
| HG01766.SG | Spain | 0 | European |
| HG01767.SG | Spain | 0 | European |
| HG01768.SG | Spain | 0 | European |
| HG01770.SG | Spain | 0 | European |
| HG01771.SG | Spain | 0 | European |
| HG01773.SG | Spain | 0 | European |
| HG01775.SG | Spain | 0 | European |
| HG01776.SG | Spain | 0 | European |
| HG01777.SG | Spain | 0 | European |
| HG01779.SG | Spain | 0 | European |
| HG01781.SG | Spain | 0 | European |
| HG01783.SG | Spain | 0 | European |
| HG01784.SG | Spain | 0 | European |
| HG01785.SG | Spain | 0 | European |
| HG01786.SG | Spain | 0 | European |
| HG01789.SG | United Kingdom | 0 | European |
| HG01790.SG | United Kingdom | 0 | European |
| HG01791.SG | United Kingdom | 0 | European |
| HG02215.SG | United Kingdom | 0 | European |

| | | | |
|---|---|---|---|
| HG02219.SG | Spain | 0 | European |
| HG02220.SG | Spain | 0 | European |
| HG02221.SG | Spain | 0 | European |
| HG02223.SG | Spain | 0 | European |
| HG02224.SG | Spain | 0 | European |
| HG02230.SG | Spain | 0 | European |
| HG02231.SG | Spain | 0 | European |
| HG02232.SG | Spain | 0 | European |
| HG02233.SG | Spain | 0 | European |
| HG02235.SG | Spain | 0 | European |
| HG02236.SG | Spain | 0 | European |
| HG02238.SG | Spain | 0 | European |
| HG02239.SG | Spain | 0 | European |
| NA20502.SG | Italy | 0 | European |
| NA20503.SG | Italy | 0 | European |
| NA20504.SG | Italy | 0 | European |
| NA20505.SG | Italy | 0 | European |
| NA20506.SG | Italy | 0 | European |
| NA20507.SG | Italy | 0 | European |
| NA20508.SG | Italy | 0 | European |
| NA20509.SG | Italy | 0 | European |
| NA20510.SG | Italy | 0 | European |
| NA20511.SG | Italy | 0 | European |
| NA20512.SG | Italy | 0 | European |
| NA20513.SG | Italy | 0 | European |
| NA20514.SG | Italy | 0 | European |
| NA20515.SG | Italy | 0 | European |
| NA20516.SG | Italy | 0 | European |
| NA20517.SG | Italy | 0 | European |

| | | | |
|---|---|---|---|
| NA20518.SG | Italy | 0 | European |
| NA20519.SG | Italy | 0 | European |
| NA20520.SG | Italy | 0 | European |
| NA20521.SG | Italy | 0 | European |
| NA20522.SG | Italy | 0 | European |
| NA20524.SG | Italy | 0 | European |
| NA20525.SG | Italy | 0 | European |
| NA20526.SG | Italy | 0 | European |
| NA20527.SG | Italy | 0 | European |
| NA20528.SG | Italy | 0 | European |
| NA20529.SG | Italy | 0 | European |
| NA20530.SG | Italy | 0 | European |
| NA20531.SG | Italy | 0 | European |
| NA20532.SG | Italy | 0 | European |
| NA20533.SG | Italy | 0 | European |
| NA20534.SG | Italy | 0 | European |
| NA20535.SG | Italy | 0 | European |
| NA20536.SG | Italy | 0 | European |
| NA20538.SG | Italy | 0 | European |
| NA20539.SG | Italy | 0 | European |
| NA20540.SG | Italy | 0 | European |
| NA20541.SG | Italy | 0 | European |
| NA20542.SG | Italy | 0 | European |
| NA20543.SG | Italy | 0 | European |
| NA20544.SG | Italy | 0 | European |
| NA20581.SG | Italy | 0 | European |
| NA20582.SG | Italy | 0 | European |
| NA20585.SG | Italy | 0 | European |
| NA20586.SG | Italy | 0 | European |

| | | | |
|---|---|---|---|
| NA20587.SG | Italy | 0 | European |
| NA20588.SG | Italy | 0 | European |
| NA20589.SG | Italy | 0 | European |
| NA20752.SG | Italy | 0 | European |
| NA20753.SG | Italy | 0 | European |
| NA20754.SG | Italy | 0 | European |
| NA20755.SG | Italy | 0 | European |
| NA20756.SG | Italy | 0 | European |
| NA20757.SG | Italy | 0 | European |
| NA20758.SG | Italy | 0 | European |
| NA20759.SG | Italy | 0 | European |
| NA20760.SG | Italy | 0 | European |
| NA20761.SG | Italy | 0 | European |
| NA20762.SG | Italy | 0 | European |
| NA20763.SG | Italy | 0 | European |
| NA20764.SG | Italy | 0 | European |
| NA20765.SG | Italy | 0 | European |
| NA20766.SG | Italy | 0 | European |
| NA20767.SG | Italy | 0 | European |
| NA20768.SG | Italy | 0 | European |
| NA20769.SG | Italy | 0 | European |
| NA20770.SG | Italy | 0 | European |
| NA20771.SG | Italy | 0 | European |
| NA20772.SG | Italy | 0 | European |
| NA20773.SG | Italy | 0 | European |
| NA20774.SG | Italy | 0 | European |
| NA20775.SG | Italy | 0 | European |
| NA20778.SG | Italy | 0 | European |
| NA20783.SG | Italy | 0 | European |

| | | | |
|---|---|---|---|
| NA20785.SG | Italy | 0 | European |
| NA20786.SG | Italy | 0 | European |
| NA20787.SG | Italy | 0 | European |
| NA20790.SG | Italy | 0 | European |
| NA20792.SG | Italy | 0 | European |
| NA20795.SG | Italy | 0 | European |
| NA20796.SG | Italy | 0 | European |
| NA20797.SG | Italy | 0 | European |
| NA20798.SG | Italy | 0 | European |
| NA20799.SG | Italy | 0 | European |
| NA20800.SG | Italy | 0 | European |
| NA20801.SG | Italy | 0 | European |
| NA20802.SG | Italy | 0 | European |
| NA20803.SG | Italy | 0 | European |
| NA20804.SG | Italy | 0 | European |
| NA20805.SG | Italy | 0 | European |
| NA20806.SG | Italy | 0 | European |
| NA20807.SG | Italy | 0 | European |
| NA20808.SG | Italy | 0 | European |
| NA20809.SG | Italy | 0 | European |
| NA20810.SG | Italy | 0 | European |
| NA20811.SG | Italy | 0 | European |
| NA20812.SG | Italy | 0 | European |
| NA20813.SG | Italy | 0 | European |
| NA20814.SG | Italy | 0 | European |
| NA20815.SG | Italy | 0 | European |
| NA20818.SG | Italy | 0 | European |
| NA20819.SG | Italy | 0 | European |
| NA20821.SG | Italy | 0 | European |

| | | | |
|---|---|---|---|
| NA20822.SG | Italy | 0 | European |
| NA20826.SG | Italy | 0 | European |
| NA20827.SG | Italy | 0 | European |
| NA20828.SG | Italy | 0 | European |
| NA20832.SG | Italy | 0 | European |
| I5014 | Germany | 4200 | European |
| I0700 | Bulgaria | 7550 | European |
| I0704_published | Bulgaria | 7892 | European |
| I0706 | Bulgaria | 7899 | European |
| I1108 | Bulgaria | 7550 | European |
| I1109 | Bulgaria | 7550 | European |
| I1113 | Bulgaria | 7550 | European |
| I1295 | Bulgaria | 7550 | European |
| I1296_published | Bulgaria | 7550 | European |
| I1297 | Bulgaria | 7550 | European |
| I1733 | Ukraine | 10200 | European |
| I1737_published | Ukraine | 10358 | European |
| I2175 | Bulgaria | 5097 | European |
| I2176_published | Bulgaria | 5146 | European |
| I2181_published | Bulgaria | 6458 | European |
| I2215_published | Bulgaria | 7550 | European |
| I2216_published | Bulgaria | 7550 | European |
| I2423 | Bulgaria | 6381 | European |
| I2424 | Bulgaria | 6288 | European |
| I2425 | Bulgaria | 6475 | European |
| I2426_published | Bulgaria | 6302 | European |
| I2430 | Bulgaria | 6448 | European |
| I2526 | Bulgaria | 7293 | European |
| I4131 | Hungary | 4305 | European |

| | | | |
|---|---|---|---|
| I4124 | Germany | 4278 | European |
| I5019 | Germany | 4200 | European |
| I4143 | Germany | 4267 | European |
| I4144 | Germany | 4411 | European |
| I4134 | Germany | 4250 | European |
| I5023 | Germany | 4200 | European |
| I4145 | Czech Republic | 4089 | European |
| I4136 | Czech Republic | 4250 | European |
| I4139 | Czech Republic | 4150 | European |
| I5037 | Czech Republic | 4100 | European |
| I5042 | Czech Republic | 4100 | European |
| I4141 | Czech Republic | 4250 | European |
| I4130 | Czech Republic | 4250 | European |
| I5116 | Hungary | 5100 | European |
| I5117 | Hungary | 5100 | European |
| I5118 | Hungary | 5100 | European |
| I5119 | Hungary | 5097 | European |
| I0018 | Germany | 7168 | European |
| ILK001 | Ukraine | 4773 | European |
| ILK002 | Ukraine | 4734 | European |
| ILK003 | Ukraine | 4781 | European |
| I1875 | Croatia | 9056 | European |
| I4304 | France | 6637 | European |
| I4305 | France | 6680 | European |
| I4308 | France | 5450 | European |
| I4595 | Latvia | 7637 | European |
| I4596 | Latvia | 7970 | European |
| I4630 | Latvia | 9218 | European |
| I4632 | Latvia | 8316 | European |

| | | | |
|---|---|---|---|
| I4871 | Serbia | 8450 | European |
| I4872 | Serbia | 8450 | European |
| I4873 | Serbia | 7867 | European |
| I4874 | Serbia | 8499 | European |
| I4875 | Serbia | 8458 | European |
| I4876 | Serbia | 8455 | European |
| I4877 | Serbia | 8505 | European |
| I4878 | Serbia | 7803 | European |
| I4880 | Serbia | 7813 | European |
| I4881_published | Serbia | 8363 | European |
| I4882 | Serbia | 8000 | European |
| I4914_published | Serbia | 8088 | European |
| I4915 | Serbia | 8115 | European |
| I4916 | Serbia | 8763 | European |
| I4917 | Serbia | 8058 | European |
| I4918 | Serbia | 7600 | European |
| I4945 | Czech Republic | 4177 | European |
| I4946 | Czech Republic | 4188 | European |
| I4947 | Czech Republic | 4300 | European |
| I4949_published | United Kingdom | 5417 | European |
| I4950 | United Kingdom | 4250 | European |
| I4951 | United Kingdom | 4250 | European |
| I4971 | Hungary | 7599 | European |
| I4972 | Hungary | 7700 | European |
| I5068 | Austria | 7088 | European |
| I5069 | Austria | 6961 | European |
| I5070 | Austria | 7048 | European |
| I5071 | Croatia | 7582 | European |
| I5072 | Croatia | 7545 | European |

| | | | |
|---|---|---|---|
| I5077 | Croatia | 7001 | European |
| I5078 | Croatia | 6571 | European |
| I5079 | Croatia | 5421 | European |
| I5232 | Serbia | 7901 | European |
| I5233 | Serbia | 8001 | European |
| I5234 | Serbia | 9500 | European |
| I5235 | Serbia | 10835 | European |
| I5236 | Serbia | 10038 | European |
| I5237 | Serbia | 9500 | European |
| I5238 | Serbia | 9993 | European |
| I5239 | Serbia | 10333 | European |
| I5240 | Serbia | 10805 | European |
| I5241 | Serbia | 11198 | European |
| I5242 | Serbia | 10530 | European |
| I5244 | Serbia | 10785 | European |
| I5269 | Russia | 4902 | European |
| I5270 | Russia | 5058 | European |
| I5271 | Russia | 4897 | European |
| I5272 | Russia | 4861 | European |
| I5273 | Russia | 4872 | European |
| I5277 | Russia | 4952 | European |
| I5278 | Russia | 4928 | European |
| I5279 | Russia | 4891 | European |
| I4884 | Czech Republic | 3758 | European |
| I4885 | Czech Republic | 4170 | European |
| I4886 | Czech Republic | 4088 | European |
| I4887 | Czech Republic | 4071 | European |
| I4888 | Czech Republic | 4038 | European |
| I4889 | Czech Republic | 4131 | European |

| | | | |
|---|---|---|---|
| I4890 | Czech Republic | 4250 | European |
| I4891 | Czech Republic | 4131 | European |
| I4892 | Czech Republic | 3754 | European |
| I4893 | Czech Republic | 6345 | European |
| I4894 | Czech Republic | 6371 | European |
| I4895 | Czech Republic | 4106 | European |
| I4896 | Czech Republic | 4163 | European |
| I5204 | Austria | 6950 | European |
| I5205_published | Austria | 6950 | European |
| I5206_published | Austria | 6950 | European |
| I5512 | United Kingdom | 4250 | European |
| I5513 | United Kingdom | 4250 | European |
| I5514 | Czech Republic | 4155 | European |
| I5515 | United Kingdom | 3882 | European |
| I5516 | United Kingdom | 3641 | European |
| I5519 | Germany | 4200 | European |
| I5520 | Germany | 4250 | European |
| I5521 | Germany | 4250 | European |
| I5522 | Germany | 4250 | European |
| I5523 | Germany | 4250 | European |
| I5524 | Germany | 4250 | European |
| I5525 | Germany | 4250 | European |
| I5526 | Germany | 4250 | European |
| I5527 | Germany | 4250 | European |
| I5529 | Germany | 4250 | European |
| I5530 | Germany | 4250 | European |
| I5531 | Germany | 4250 | European |
| I5748 | Netherlands | 4383 | European |
| I5750 | Netherlands | 4050 | European |

| | | | |
|---|---|---|---|
| I5755 | Switzerland | 4175 | European |
| I5757 | Switzerland | 4175 | European |
| I5759 | Switzerland | 4175 | European |
| I5766 | Russia | 6049 | European |
| I5769 | Bulgaria | 2592 | European |
| I5771 | Serbia | 8325 | European |
| I5772 | Serbia | 8450 | European |
| I5773_published | Serbia | 10040 | European |
| I5833 | Germany | 4200 | European |
| I5834 | Germany | 4200 | European |
| I5835 | Germany | 4200 | European |
| I5836 | Germany | 4250 | European |
| I5838 | Spain | 4550 | European |
| I5868_published | Ukraine | 7313 | European |
| I5869 | Ukraine | 7465 | European |
| I5870 | Ukraine | 7363 | European |
| I5872_published | Ukraine | 7374 | European |
| I5873_published | Ukraine | 7334 | European |
| I5875 | Ukraine | 7095 | European |
| I5876 | Ukraine | 8832 | European |
| I5878_published | Ukraine | 7139 | European |
| I5879 | Ukraine | 7208 | European |
| I5881_published | Ukraine | 7077 | European |
| I5882 | Ukraine | 5005 | European |
| I5883 | Ukraine | 7048 | European |
| I5885_published | Ukraine | 8099 | European |
| I5886_published | Ukraine | 7212 | European |
| I5888_published | Ukraine | 6950 | European |
| I5889_published | Ukraine | 6988 | European |

| | | | |
|---|---|---|---|
| I5890 | Ukraine | 7092 | European |
| I5891_published | Ukraine | 7297 | European |
| I5892 | Ukraine | 7069 | European |
| I5893_published | Ukraine | 7249 | European |
| I0679_d | Bulgaria | 7617 | European |
| I2417 | United Kingdom | 4250 | European |
| I2443 | United Kingdom | 4091 | European |
| I2446_published | United Kingdom | 4215 | European |
| I2447 | United Kingdom | 3938 | European |
| I2450 | United Kingdom | 4248 | European |
| I2458 | United Kingdom | 3331 | European |
| I2461 | United Kingdom | 3975 | European |
| I2462 | United Kingdom | 3942 | European |
| I2469_published | Spain | 2826 | European |
| I2477 | Italy | 4015 | European |
| I2484_all | Austria | 30850 | European |
| I2521 | Bulgaria | 7503 | European |
| I2529_published | Bulgaria | 7610 | European |
| I2794 | Hungary | 7564 | European |
| I2937_all | Greece | 7360 | European |
| I3238 | Spain | 4300 | European |
| I4930 | Italy | 4709 | European |
| I4933 | Italy | 4150 | European |
| I5207_published | Austria | 6950 | European |
| I5208 | Austria | 6950 | European |
| I5358 | United Kingdom | 4918 | European |
| I5359 | United Kingdom | 5600 | European |
| I5364 | United Kingdom | 2644 | European |
| I5366 | United Kingdom | 5516 | European |

| | | | |
|---|---|---|---|
| I5367 | United Kingdom | 4312 | European |
| I5370 | United Kingdom | 5600 | European |
| I5371 | United Kingdom | 5600 | European |
| I5373 | United Kingdom | 4034 | European |
| I5374 | United Kingdom | 4480 | European |
| I5376 | United Kingdom | 4216 | European |
| I5379 | United Kingdom | 4333 | European |
| I5382 | United Kingdom | 4070 | European |
| I5385 | United Kingdom | 4230 | European |
| I5387 | United Kingdom | 4850 | European |
| I5401 | Serbia | 8838 | European |
| I5402 | Serbia | 8156 | European |
| I5405 | Serbia | 7684 | European |
| I5407 | Serbia | 8000 | European |
| I5408 | Romania | 8704 | European |
| I5409_published | Serbia | 9500 | European |
| I5411 | Romania | 8600 | European |
| I5427 | Greece | 7883 | European |
| I5441 | United Kingdom | 3750 | European |
| I5469 | United Kingdom | 3517 | European |
| I5470_published | United Kingdom | 3477 | European |
| I5471_published | United Kingdom | 4063 | European |
| I5473_published | United Kingdom | 4200 | European |
| I5655_published | Germany | 4200 | European |
| I5656 | Germany | 4200 | European |
| I5657_published | Germany | 4200 | European |
| I5658 | Germany | 4200 | European |
| I5659 | Germany | 4200 | European |
| I5660_published | Germany | 4200 | European |

| | | | |
|---|---|---|---|
| I5661_published | Germany | 4200 | European |
| I5663_published | Germany | 4200 | European |
| I5665 | Spain | 4077 | European |
| I5666 | Czech Republic | 4250 | European |
| I5957_published | Ukraine | 7287 | European |
| I6047 | Russia | 3234 | European |
| I6133_published | Ukraine | 7200 | European |
| I6468 | Czech Republic | 4150 | European |
| I6470 | Spain | 3609 | European |
| I6471_published | Spain | 4200 | European |
| I6475 | Spain | 4200 | European |
| I6476 | Czech Republic | 4100 | European |
| I6480 | Czech Republic | 4150 | European |
| I6481 | Germany | 4250 | European |
| I6482 | Germany | 4250 | European |
| I6531 | Poland | 4113 | European |
| I6534 | Poland | 4237 | European |
| I6535 | Poland | 4284 | European |
| I6537 | Poland | 4130 | European |
| I6538 | Poland | 3823 | European |
| I6539 | Spain | 4200 | European |
| I6542_published | Spain | 4100 | European |
| I6543 | Spain | 4100 | European |
| I6579 | Poland | 4155 | European |
| I6580 | Poland | 4175 | European |
| I6581 | Poland | 4228 | European |
| I6582 | Poland | 4171 | European |
| I6583 | Poland | 4138 | European |
| I6584 | Spain | 4200 | European |

| | | | |
|---|---|---|---|
| I6585 | Spain | 4181 | European |
| I6587 | Spain | 4200 | European |
| I6588 | Spain | 4200 | European |
| I6589_published | Spain | 4200 | European |
| I6590 | Germany | 4200 | European |
| I6591 | Germany | 4200 | European |
| I6596_published | Spain | 4550 | European |
| I6604 | Spain | 3947 | European |
| I6605 | Spain | 4335 | European |
| I6608_published | Spain | 3840 | European |
| I6610 | Spain | 3803 | European |
| I6612 | Spain | 4100 | European |
| I6613 | Spain | 4100 | European |
| I6617 | Spain | 4550 | European |
| I6618 | Spain | 3735 | European |
| I6623 | Spain | 4100 | European |
| I6624 | Germany | 4200 | European |
| I6628 | Spain | 4550 | European |
| I6629 | Spain | 4550 | European |
| I6630 | Spain | 4550 | European |
| I3082 | United Kingdom | 3386 | European |
| I3209 | Spain | 7815 | European |
| I4132 | Germany | 4200 | European |
| I4245 | Spain | 4316 | European |
| I6472 | Spain | 4200 | European |
| I0698_published | Bulgaria | 7900 | European |
| Canes.SG | Spain | 7089 | European |
| Chan.SG | Spain | 9109 | European |
| GB.SG | Romania | 5389 | European |

| | | | |
|---|---|---|---|
| M95.SG | Romania | 8814 | European |
| M96.SG | Romania | 8825 | European |
| OC.SG | Romania | 8704 | European |
| I6744 | United Kingdom | 10393 | European |
| I6745 | United Kingdom | 10546 | European |
| I1311 | Spain | 3650 | European |
| I1378 | Ukraine | 7300 | European |
| I3484 | Spain | 4074 | European |
| I6490 | Spain | 1600 | European |
| I6492 | Spain | 1600 | European |
| I6746 | United Kingdom | 5526 | European |
| I6747 | United Kingdom | 5528 | European |
| I6750 | United Kingdom | 5650 | European |
| I6751 | United Kingdom | 5650 | European |
| I6753 | United Kingdom | 5305 | European |
| I6754 | United Kingdom | 9580 | European |
| I6755 | United Kingdom | 5528 | European |
| I6757 | United Kingdom | 5493 | European |
| I6759 | United Kingdom | 5527 | European |
| I6760 | United Kingdom | 5781 | European |
| I6761 | United Kingdom | 5525 | European |
| I6762 | United Kingdom | 5700 | European |
| I6764 | United Kingdom | 5100 | European |
| I6766 | United Kingdom | 5100 | European |
| I6774 | United Kingdom | 4121 | European |
| I6775 | United Kingdom | 4150 | European |
| I6777 | United Kingdom | 4050 | European |
| I6778 | United Kingdom | 4150 | European |
| I2597 | United Kingdom | 4080 | European |

| | | | |
|---|---|---|---|
| I2604 | United Kingdom | 4073 | European |
| I2606 | United Kingdom | 5065 | European |
| I6561 | Ukraine | 3985 | European |
| I6609 | Spain | 4100 | European |
| I0519 | United Kingdom | 5180 | European |
| I0520 | United Kingdom | 5180 | European |
| I2792 | Croatia | 4676 | European |
| I2683 | Turkey | 4479 | European |
| I3122 | Italy | 4073 | European |
| I3123 | Italy | 4121 | European |
| I3124 | Italy | 3824 | European |
| I3125 | Italy | 3490 | European |
| I4062 | Italy | 6815 | European |
| I4065 | Italy | 6815 | European |
| I6294 | Russia | 4709 | European |
| I0518 | United Kingdom | 5180 | European |
| I2786 | Hungary | 4270 | European |
| I3708 | Greece | 6500 | European |
| I3718 | Ukraine | 7217 | European |
| GoyetQ-2_udg | Belgium | 15077 | European |
| GoyetQ116-1_udg_published | Belgium | 35208 | European |
| I5383 | United Kingdom | 2945 | European |
| I6767 | United Kingdom | 10280 | European |
| I5377 | United Kingdom | 3758 | European |
| I3387 | Russia | 4725 | European |
| I2495 | Turkey | 4355 | European |
| I3709 | Greece | 5860 | European |
| I3243 | Spain | 4300 | European |
| I1392 | France | 4516 | European |

| | | | |
|---|---|---|---|
| I1192 | Russia | 6711 | European |
| I1194 | Russia | 6652 | European |
| I2484 | Austria | 30850 | European |
| I4582 | Romania | 8618 | European |
| I4063 | Italy | 6832 | European |
| I2318 | Greece | 5935 | European |
| I2937 | Greece | 7360 | European |
| I4607 | Romania | 8927 | European |
| I2499 | Turkey | 4514 | European |
| I3920 | Greece | 5734 | European |
| I3997 | Spain | 3642 | European |
| I4660 | Serbia | 9713 | European |
| I4870 | Serbia | 8740 | European |
| I3982 | Spain | 1650 | European |
| I6679 | United Kingdom | 3936 | European |
| I6680 | United Kingdom | 3671 | European |
| I1842 | Spain | 4850 | European |
| I1845 | Spain | 4825 | European |
| I1846 | Spain | 4825 | European |
| I1978 | Spain | 4825 | European |
| I6677 | Czech Republic | 5550 | European |
| I1979 | Italy | 4015 | European |
| I1982 | Spain | 3558 | European |
| I6048 | Russia | 3573 | European |
| MA969.SG | Estonia | 4652 | European |
| MA971.SG | Estonia | 4400 | European |
| MA973.SG | Estonia | 4416 | European |
| MA974.SG | Estonia | 5586 | European |
| MA975.SG | Estonia | 5488 | European |

| | | | |
|---|---|---|---|
| Ofnet_published_d | Germany | 8277 | European |
| LesCloseaux13_published_d | France | 9933 | European |
| HohleFels79_published_d | Germany | 14691 | European |
| Cioclovina1_published_d | Romania | 32669 | European |
| Brillenhohle_published_d | Germany | 14817 | European |
| Bockstein_published_d | Germany | 8205 | European |
| GoyetQ376-19_published_d | Belgium | 27505 | European |
| HohleFels49_published_d | Germany | 15491 | European |
| Falkenstein_published_d | Germany | 9161 | European |
| Chaudardes1_published_d | France | 8218 | European |
| Ranchot88_published_d | France | 10134 | European |
| Sunghir1.SG | Russia | 33209 | European |
| Sunghir2.SG | Russia | 34629 | European |
| Sunghir3.SG | Russia | 34517 | European |
| Sunghir4.SG | Russia | 34323 | European |
| Sunghir6.SG | Russia | 813 | European |
| I7158 | Spain | 1600 | European |
| I7160_published | Spain | 5877 | European |
| I3584_published | Spain | 1400 | European |
| I0982 | Russia | 3825 | European |
| I7162_published | Spain | 3760 | European |
| I4558 | Spain | 3550 | European |
| I4559 | Spain | 3550 | European |
| I4560 | Spain | 3550 | European |
| I4561 | Spain | 3550 | European |
| I0937 | Russia | 3825 | European |
| I0938 | Russia | 3825 | European |
| I0939 | Russia | 3825 | European |
| I0940 | Russia | 3825 | European |

| | | | |
|---|---|---|---|
| I0942 | Russia | 3825 | European |
| I0943 | Russia | 3825 | European |
| I0945 | Russia | 3900 | European |
| I0946 | Russia | 3886 | European |
| I0980 | Russia | 3825 | European |
| I0983 | Russia | 3825 | European |
| I0984 | Russia | 3870 | European |
| I0985 | Russia | 3825 | European |
| I0986 | Russia | 3825 | European |
| I0989 | Russia | 3825 | European |
| I1003 | Russia | 3825 | European |
| I1006 | Russia | 3825 | European |
| I1007 | Russia | 3825 | European |
| I1008 | Russia | 3825 | European |
| I1010 | Russia | 3825 | European |
| I1011 | Russia | 3825 | European |
| I1013 | Russia | 3825 | European |
| I3809_published | Spain | 400 | European |
| I4329 | Spain | 4333 | European |
| I1525 | Russia | 1695 | European |
| I1526 | Russia | 4779 | European |
| I0244 | Russia | 4154 | European |
| I0232 | Russia | 3475 | European |
| I0246 | Russia | 4138 | European |
| I0354 | Russia | 3791 | European |
| I0358 | Russia | 3719 | European |
| I0359 | Russia | 3475 | European |
| I0360 | Russia | 3800 | European |
| I0361 | Russia | 3475 | European |

| | | | |
|---|---|---|---|
| I0419 | Russia | 4000 | European |
| I0421 | Russia | 3675 | European |
| I0422 | Russia | 3475 | European |
| I0424 | Russia | 3675 | European |
| I0430 | Russia | 3675 | European |
| I0431 | Russia | 3675 | European |
| I1061 | Russia | 3825 | European |
| I7333 | Russia | 1250 | European |
| I7347 | Russia | 725 | European |
| I7349 | Russia | 1140 | European |
| I7423_published | Spain | 400 | European |
| I7424 | Spain | 400 | European |
| I7425 | Spain | 400 | European |
| I7426_published | Spain | 400 | European |
| I7458 | Spain | 750 | European |
| I7480 | Russia | 3825 | European |
| I7489 | Russia | 5047 | European |
| I7497 | Spain | 900 | European |
| I7499 | Spain | 900 | European |
| I7500 | Spain | 950 | European |
| I7549 | Spain | 5950 | European |
| I7550 | Spain | 6100 | European |
| I7588 | Spain | 4650 | European |
| I7600 | Spain | 5625 | European |
| I7602 | Spain | 5050 | European |
| I7603 | Spain | 5050 | European |
| I7605 | Spain | 5050 | European |
| I7621 | Russia | 1355 | European |
| I7643 | Spain | 5803 | European |

| | | | |
|---|---|---|---|
| I7644 | Spain | 5521 | European |
| I7646 | Spain | 5613 | European |
| I7670 | Russia | 3958 | European |
| I7759 | Russia | 7035 | European |
| I7779 | Russia | 4750 | European |
| I7781 | Russia | 615 | European |
| I7782 | Russia | 4630 | European |
| I7647 | Spain | 5945 | European |
| I7645 | Spain | 5845 | European |
| I7547 | Spain | 5950 | European |
| I7587 | Spain | 4650 | European |
| I7601 | Spain | 5527 | European |
| I7348 | Russia | 1205 | European |
| I7606 | Spain | 5050 | European |
| I7331 | Russia | 1045 | European |
| I7339 | Russia | 1085 | European |
| I7335 | Russia | 4465 | European |
| I7498 | Spain | 900 | European |
| I7671 | Russia | 4729 | European |
| I7642 | Spain | 4809 | European |
| I7457 | Spain | 750 | European |
| I7604 | Spain | 5050 | European |
| I7340 | Russia | 1020 | European |
| I7338 | Russia | 610 | European |
| I7343 | Russia | 860 | European |
| I7344 | Russia | 1080 | European |
| I7337 | Russia | 1150 | European |
| I7760 | Russia | 5820 | European |
| I7342_d | Russia | 2062 | European |

| | | | |
|---|---|---|---|
| I7672 | Spain | 1153 | European |
| I7673 | Spain | 1153 | European |
| I7674 | Spain | 1153 | European |
| I7675 | Spain | 1153 | European |
| I7676 | Spain | 1153 | European |
| I7679_published | Spain | 5950 | European |
| Hum1.SG | Norway | 9364 | European |
| Hum2.SG | Norway | 9364 | European |
| SBj.SG | Sweden | 8771 | European |
| SF11.SG | Sweden | 8892 | European |
| SF12.SG | Sweden | 8895 | European |
| SF9.SG | Sweden | 9144 | European |
| Steigen.SG | Norway | 5857 | European |
| TAP002 | French Polynesia | 203 | European |
| TAP003 | French Polynesia | 366 | European |
| TAP004 | French Polynesia | 228 | European |
| Donkalnis1 | Lithuania | 6150 | European |
| Donkalnis4 | Lithuania | 7823 | European |
| Donkalnis6 | Lithuania | 6569 | European |
| Donkalnis7 | Lithuania | 7110 | European |
| Gyvakarai1_10bp | Lithuania | 4495 | European |
| Kivisaare3 | Estonia | 6594 | European |
| Kivutkalns153 | Latvia | 2633 | European |
| Kivutkalns164 | Latvia | 2423 | European |
| Kivutkalns194 | Latvia | 2298 | European |
| Kivutkalns19 | Latvia | 2433 | European |
| Kivutkalns207 | Latvia | 2600 | European |
| Kivutkalns209 | Latvia | 2616 | European |
| Kivutkalns215 | Latvia | 2559 | European |

| | | | |
|---|---|---|---|
| Kivutkalns222 | Latvia | 2473 | European |
| Kivutkalns25 | Latvia | 2637 | European |
| Kivutkalns42 | Latvia | 2697 | European |
| Kretuonas1 | Lithuania | 6123 | European |
| Kretuonas2 | Lithuania | 6150 | European |
| Kretuonas4 | Lithuania | 6150 | European |
| Kretuonas5 | Lithuania | 6150 | European |
| Kunila2 | Estonia | 4416 | European |
| Kvarlov5164 | Sweden | 5746 | European |
| MotalaAA | Sweden | 7599 | European |
| Olsund | Sweden | 4306 | European |
| Plinkaigalis241 | Lithuania | 4528 | European |
| Plinkaigalis242 | Lithuania | 4842 | European |
| Popovo2 | Russia | 8200 | European |
| Spiginas1 | Lithuania | 6263 | European |
| Spiginas2 | Lithuania | 3880 | European |
| Spiginas4 | Lithuania | 8276 | European |
| Tamula1 | Estonia | 5407 | European |
| Tamula3 | Estonia | 5666 | European |
| Turlojiske1932 | Lithuania | 3036 | European |
| Turlojiske1 | Lithuania | 2819 | European |
| Turlojiske3 | Lithuania | 2846 | European |
| Turlojiske5 | Lithuania | 3300 | European |
| UzOO77 | Russia | 7200 | European |
| Veibri4 | Estonia | 6760 | European |
| I0987 | Russia | 3825 | European |
| I10045 | Croatia | 6050 | European |
| I10047 | Croatia | 6050 | European |
| I10048 | Croatia | 6050 | European |

| | | | |
|---|---|---|---|
| I10049 | Croatia | 6050 | European |
| I10050 | Croatia | 6050 | European |
| I10052 | Croatia | 6095 | European |
| I10053 | Croatia | 6050 | European |
| I10055 | Croatia | 6050 | European |
| I10056 | Croatia | 6050 | European |
| I10057 | Croatia | 6050 | European |
| I10058 | Croatia | 6050 | European |
| I10061 | Croatia | 6050 | European |
| I10062 | Croatia | 6050 | European |
| I10063 | Croatia | 6050 | European |
| I10064 | Croatia | 6050 | European |
| I10065 | Croatia | 6050 | European |
| I10067 | Croatia | 6050 | European |
| I10068 | Croatia | 6050 | European |
| I10069 | Croatia | 6050 | European |
| I10071 | Croatia | 6050 | European |
| I10072 | Croatia | 6050 | European |
| I10074 | Croatia | 6050 | European |
| I10271 | Croatia | 6050 | European |
| I10272 | Croatia | 6023 | European |
| I10273 | Croatia | 6050 | European |
| I10275 | Croatia | 6050 | European |
| I10277 | Spain | 4900 | European |
| I10278 | Spain | 4900 | European |
| I10280 | Spain | 4900 | European |
| I10282_published | Spain | 4900 | European |
| I10283 | Spain | 4900 | European |
| I10284 | Spain | 4900 | European |

| | | | |
|---|---|---|---|
| I10285 | Spain | 4900 | European |
| I10286 | Spain | 4900 | European |
| I10291 | Croatia | 6050 | European |
| I10292 | Croatia | 6050 | European |
| I10293 | Croatia | 6093 | European |
| I10295 | Croatia | 6050 | European |
| I10299 | Croatia | 6050 | European |
| I10303 | Croatia | 6050 | European |
| I10364 | Italy | 2925 | European |
| I10365_published | Italy | 3404 | European |
| I10366 | Italy | 2244 | European |
| I10939 | Gibraltar | 3600 | European |
| I10940 | Gibraltar | 3600 | European |
| I10941 | Gibraltar | 3600 | European |
| I10942 | Gibraltar | 6950 | European |
| I11214 | Russia | 3497 | European |
| I11442 | Italy | 4120 | European |
| I11443 | Italy | 4623 | European |
| I4420 | Spain | 3748 | European |
| I7594 | Spain | 5396 | European |
| I7595_published | Spain | 5396 | European |
| I7597 | Spain | 5625 | European |
| I7598 | Spain | 5411 | European |
| I7796 | Italy | 3731 | European |
| I7800 | Italy | 3945 | European |
| I7807 | Italy | 4038 | European |
| I7949 | Czech Republic | 3022 | European |
| I8048 | Spain | 4050 | European |
| I8130 | Spain | 7957 | European |

| | | | |
|---|---|---|---|
| I8132 | Spain | 4450 | European |
| I8136 | Spain | 3426 | European |
| I8141 | Spain | 4050 | European |
| I8144 | Spain | 3680 | European |
| I8145 | Spain | 550 | European |
| I8146 | Spain | 550 | European |
| I8147 | Spain | 550 | European |
| I8148 | Spain | 4650 | European |
| I8149 | Spain | 4650 | European |
| I8150 | Spain | 4650 | European |
| I8153 | Spain | 4650 | European |
| I8154 | Spain | 4650 | European |
| I8155_published | Spain | 4650 | European |
| I8156 | Spain | 4650 | European |
| I8157_published | Spain | 4650 | European |
| I8158 | Spain | 4650 | European |
| I8197 | Spain | 4650 | European |
| I8198 | Spain | 4650 | European |
| I8199 | Spain | 4650 | European |
| I8338 | Spain | 1800 | European |
| I8339 | Spain | 1777 | European |
| I8340 | Spain | 2375 | European |
| I8341 | Spain | 2375 | European |
| I8343 | Spain | 1287 | European |
| I8344 | Spain | 2400 | European |
| I8364 | Spain | 4588 | European |
| I8365 | Spain | 4588 | European |
| I8474 | Spain | 1800 | European |
| I8475 | Spain | 1800 | European |

| | | | |
|---|---|---|---|
| I8561 | Italy | 4212 | European |
| I8566 | Spain | 5100 | European |
| I8567 | Spain | 5350 | European |
| I8568 | Spain | 5387 | European |
| I8569 | Spain | 4692 | European |
| I8570_published | Spain | 3100 | European |
| I8571 | Spain | 3350 | European |
| I8745 | Russia | 4300 | European |
| I4420_all | Spain | 3748 | European |
| I10046 | Croatia | 6050 | European |
| I10054 | Croatia | 6050 | European |
| I10070 | Croatia | 6050 | European |
| I10274 | Croatia | 6050 | European |
| I10294 | Croatia | 6050 | European |
| I10297 | Croatia | 6050 | European |
| I1976 | Spain | 4427 | European |
| I2471 | Spain | 3500 | European |
| I2473 | Spain | 4811 | European |
| I3214 | Spain | 5200 | European |
| I3239 | Spain | 4300 | European |
| I3277 | Spain | 4950 | European |
| I3326 | Spain | 2175 | European |
| I3327 | Spain | 2175 | European |
| I3492 | Spain | 3450 | European |
| I3757 | Spain | 2300 | European |
| I3758 | Spain | 2226 | European |
| I4665 | Serbia | 8006 | European |
| I4936_published | Italy | 4150 | European |
| I6622 | Spain | 4100 | European |

| | | | |
|---|---|---|---|
| I8131 | Spain | 4459 | European |
| I8202 | Spain | 1750 | European |
| I8203 | Spain | 2150 | European |
| I8204 | Spain | 2179 | European |
| I8205 | Spain | 2150 | European |
| I8206 | Spain | 2103 | European |
| I8208 | Spain | 2226 | European |
| I8209 | Spain | 2554 | European |
| I8210 | Spain | 2375 | European |
| I8211 | Spain | 2425 | European |
| I8212 | Spain | 2375 | European |
| I8213 | Spain | 2400 | European |
| I8214 | Spain | 2375 | European |
| I8215 | Spain | 2526 | European |
| I8216 | Spain | 1795 | European |
| I1312_d | Spain | 3650 | European |
| I1313_d | Spain | 3650 | European |
| atp002.SG | Spain | 4746 | European |
| atp005.SG | Spain | 7069 | European |
| atp016.SG | Spain | 4971 | European |
| atp12-1420.SG | Spain | 4868 | European |
| c40331.SG | Spain | 5630 | European |
| esp005.SG | Spain | 3400 | European |
| mur.SG | Spain | 7127 | European |
| pir001.SG | Spain | 3725 | European |
| por002.SG | Spain | 5111 | European |
| por004.SG | Spain | 4955 | European |
| san216.SG | Spain | 5657 | European |
| DA111.SG | Czech Republic | 2752 | European |

| | | | |
|---|---|---|---|
| DA112.SG | Czech Republic | 2507 | European |
| DA119.SG | Slovakia | 1600 | European |
| DA134.SG | Russia | 2300 | European |
| DA139.SG | Russia | 2300 | European |
| DA141.SG | Russia | 2300 | European |
| DA142.SG | Russia | 1000 | European |
| DA143.SG | Russia | 2300 | European |
| DA144.SG | Russia | 2300 | European |
| DA145.SG | Russia | 2300 | European |
| DA146.SG | Russia | 1300 | European |
| DA160.SG | Russia | 1300 | European |
| DA161.SG | Russia | 1300 | European |
| DA162.SG | Russia | 1300 | European |
| DA164.SG | Russia | 850 | European |
| DA171.SG | Lithuania | 1600 | European |
| DA188.SG | Russia | 1107 | European |
| DA189.SG | Russia | 1150 | European |
| DA190.SG | Russia | 1263 | European |
| DA191.SG | Hungary | 2472 | European |
| DA194.SG | Hungary | 2328 | European |
| DA195.SG | Hungary | 2572 | European |
| DA197.SG | Hungary | 2426 | European |
| DA198.SG | Hungary | 2441 | European |
| DA199.SG | Hungary | 703 | European |
| DA243.SG | Russia | 1050 | European |
| DA2.SG | Russia | 2817 | European |
| DA31.SG | Armenia | 3200 | European |
| DA343.SG | Russia | 4819 | European |
| DA353.SG | Russia | 4285 | European |

| | | | |
|---|---|---|---|
| DA356.SG | Russia | 4277 | European |
| DA35.SG | Armenia | 3259 | European |
| DA361.SG | Russia | 4130 | European |
| DA3.SG | Russia | 2850 | European |
| DA4.SG | Russia | 2850 | European |
| DA5.SG | Russia | 2850 | European |
| DA6.SG | Russia | 2850 | European |
| DA7.SG | Russia | 2850 | European |
| DA8.SG | Russia | 2850 | European |
| DA9.SG | Russia | 2850 | European |
| Altais1.SG | Russia | 0 | European |
| Altais2.SG | Russia | 0 | European |
| Bashkirs1.SG | Russia | 0 | European |
| Bashkirs2.SG | Russia | 0 | European |
| Bashkirs3.SG | Russia | 0 | European |
| Buryats1.SG | Russia | 0 | European |
| Buryats2.SG | Russia | 0 | European |
| DA245.SG | Russia | 7944 | European |
| DA246.SG | Russia | 7723 | European |
| DA247.SG | Russia | 7689 | European |
| DA248.SG | Russia | 7646 | European |
| DA249.SG | Russia | 7840 | European |
| DA250.SG | Russia | 7377 | European |
| DA251.SG | Russia | 7299 | European |
| DA252.SG | Russia | 7317 | European |
| DA253.SG | Russia | 7240 | European |
| DA334.SG | Russia | 4129 | European |
| DA335.SG | Russia | 4200 | European |
| DA336.SG | Russia | 4200 | European |

| | | | |
|---|---|---|---|
| DA337.SG | Russia | 4328 | European |
| DA338.SG | Russia | 4200 | European |
| DA339.SG | Russia | 4040 | European |
| DA340.SG | Russia | 7023 | European |
| DA341.SG | Russia | 7577 | European |
| DA342.SG | Russia | 5617 | European |
| DA344.SG | Russia | 5595 | European |
| DA345.SG | Russia | 5454 | European |
| DA354.SG | Russia | 4591 | European |
| DA355.SG | Russia | 5469 | European |
| DA357.SG | Russia | 7785 | European |
| DA358.SG | Russia | 4713 | European |
| DA359.SG | Russia | 7545 | European |
| DA360.SG | Russia | 4704 | European |
| DA362.SG | Russia | 7230 | European |
| IrtyshBarabinskTatars1.SG | Russia | 0 | European |
| IrtyshBarabinskTatars2.SG | Russia | 0 | European |
| Kalmyks1.SG | Russia | 0 | European |
| Kalmyks2.SG | Russia | 0 | European |
| MA2195_final.SG | Turkey | 375 | European |
| MA2196_final.SG | Turkey | 375 | European |
| MA2197_final.SG | Turkey | 2900 | European |
| MA2198_final.SG | Turkey | 2900 | European |
| MA2200_final.SG | Turkey | 3800 | European |
| MA2203_final.SG | Turkey | 3800 | European |
| MA2205_final.SG | Turkey | 3800 | European |
| MA2206_final.SG | Turkey | 3800 | European |
| MA2208_final.SG | Turkey | 3800 | European |
| MA2210_final.SG | Turkey | 4700 | European |

| | | | |
|---|---|---|---|
| MA2212_final.SG | Turkey | 4700 | European |
| MA2213_final.SG | Turkey | 4700 | European |
| RISE515.SG | Russia | 4196 | European |
| RISE516.SG | Russia | 4065 | European |
| RISE662.SG | Russia | 4067 | European |
| RISE664.SG | Russia | 4270 | European |
| RISE667.SG | Russia | 4300 | European |
| RISE670.SG | Russia | 3959 | European |
| RISE671.SG | Russia | 4300 | European |
| RISE672.SG | Russia | 4300 | European |
| RISE673.SG | Russia | 4300 | European |
| RISE674.SG | Russia | 4064 | European |
| RISE675.SG | Russia | 4517 | European |
| RISE677.SG | Russia | 4409 | European |
| RISE680.SG | Russia | 4300 | European |
| RISE681.SG | Russia | 4300 | European |
| RISE683.SG | Russia | 3973 | European |
| RISE684.SG | Russia | 4239 | European |
| RISE685.SG | Russia | 4300 | European |
| RISE718.SG | Russia | 4434 | European |
| RISE719.SG | Russia | 4300 | European |
| Sidelkino.SG | Russia | 11321 | European |
| Teleuts1.SG | Russia | 0 | European |
| Teleuts2.SG | Russia | 0 | European |
| TomskTatars1.SG | Russia | 0 | European |
| TomskTatars2.SG | Russia | 0 | European |
| VolgaTatars1.SG | Russia | 0 | European |
| VolgaTatars2.SG | Russia | 0 | European |
| HGDP01368.SDG | France | 0 | European |

| | | | |
|---|---|---|---|
| HGDP01385.SDG | Russia | 0 | European |
| HGDP00948.SDG | Russia | 0 | European |
| HGDP00945.SDG | Russia | 0 | European |
| HGDP01396.SDG | Russia | 0 | European |
| HGDP01383.SDG | Russia | 0 | European |
| HGDP00947.SDG | Russia | 0 | European |
| HGDP00958.SDG | Russia | 0 | European |
| HGDP00946.SDG | Russia | 0 | European |
| HGDP00966.SDG | Russia | 0 | European |
| HGDP00965.SDG | Russia | 0 | European |
| HGDP00954.SDG | Russia | 0 | European |
| HGDP00962.SDG | Russia | 0 | European |
| HGDP00963.SDG | Russia | 0 | European |
| HGDP00953.SDG | Russia | 0 | European |
| HGDP00950.SDG | Russia | 0 | European |
| HGDP00960.SDG | Russia | 0 | European |
| HGDP00964.SDG | Russia | 0 | European |
| HGDP00957.SDG | Russia | 0 | European |
| HGDP00968.SDG | Russia | 0 | European |
| HGDP00961.SDG | Russia | 0 | European |
| HGDP00952.SDG | Russia | 0 | European |
| HGDP00969.SDG | Russia | 0 | European |
| HGDP00955.SDG | Russia | 0 | European |
| HGDP01361.SDG | France | 0 | European |
| HGDP01358.SDG | France | 0 | European |
| HGDP01359.SDG | France | 0 | European |
| HGDP00536.SDG | France | 0 | European |
| HGDP00539.SDG | France | 0 | European |
| HGDP00879.SDG | Russia | 0 | European |

| | | | |
|---|---|---|---|
| HGDP00902.SDG | Russia | 0 | European |
| HGDP00519.SDG | France | 0 | European |
| HGDP00512.SDG | France | 0 | European |
| HGDP01357.SDG | France | 0 | European |
| HGDP00880.SDG | Russia | 0 | European |
| HGDP00513.SDG | France | 0 | European |
| HGDP00900.SDG | Russia | 0 | European |
| HGDP00898.SDG | Russia | 0 | European |
| HGDP00527.SDG | France | 0 | European |
| HGDP01363.SDG | France | 0 | European |
| HGDP00524.SDG | France | 0 | European |
| HGDP00511.SDG | France | 0 | European |
| HGDP01404.SDG | Russia | 0 | European |
| HGDP01384.SDG | Russia | 0 | European |
| HGDP00899.SDG | Russia | 0 | European |
| HGDP00901.SDG | Russia | 0 | European |
| HGDP00886.SDG | Russia | 0 | European |
| HGDP01388.SDG | Russia | 0 | European |
| HGDP01360.SDG | France | 0 | European |
| HGDP00520.SDG | France | 0 | European |
| HGDP00522.SDG | France | 0 | European |
| HGDP00535.SDG | France | 0 | European |
| HGDP00523.SDG | France | 0 | European |
| HGDP00518.SDG | France | 0 | European |
| HGDP00531.SDG | France | 0 | European |
| HGDP00537.SDG | France | 0 | European |
| HGDP00528.SDG | France | 0 | European |
| HGDP00516.SDG | France | 0 | European |
| HGDP00895.SDG | Russia | 0 | European |

| | | | |
|---|---|---|---|
| HGDP00891.SDG | Russia | 0 | European |
| HGDP01362.SDG | France | 0 | European |
| HGDP00529.SDG | France | 0 | European |
| HGDP00883.SDG | Russia | 0 | European |
| HGDP00515.SDG | France | 0 | European |
| HGDP00882.SDG | Russia | 0 | European |
| HGDP00538.SDG | France | 0 | European |
| HGDP01400.SDG | Russia | 0 | European |
| HGDP01398.SDG | Russia | 0 | European |
| HGDP01382.SDG | Russia | 0 | European |
| HGDP00884.SDG | Russia | 0 | European |
| HGDP01376.SDG | France | 0 | European |
| HGDP00889.SDG | Russia | 0 | European |
| HGDP01397.SDG | Russia | 0 | European |
| HGDP00896.SDG | Russia | 0 | European |
| HGDP01386.SDG | Russia | 0 | European |
| HGDP00525.SDG | France | 0 | European |
| HGDP00890.SDG | Russia | 0 | European |
| HGDP01403.SDG | Russia | 0 | European |
| HGDP00893.SDG | Russia | 0 | European |
| HGDP00897.SDG | Russia | 0 | European |
| HGDP00517.SDG | France | 0 | European |
| HGDP00799.SDG | United Kingdom | 0 | European |
| HGDP00885.SDG | Russia | 0 | European |
| HGDP00892.SDG | Russia | 0 | European |
| HGDP00888.SDG | Russia | 0 | European |
| HGDP01074.SDG | Italy | 0 | European |
| HGDP00881.SDG | Russia | 0 | European |
| HGDP00671.SDG | Italy | 0 | European |

| | | | |
|---|---|---|---|
| HGDP00514.SDG | France | 0 | European |
| HGDP01157.SDG | Italy | 0 | European |
| HGDP00894.SDG | Russia | 0 | European |
| HGDP01164.SDG | Italy | 0 | European |
| HGDP01156.SDG | Italy | 0 | European |
| HGDP01073.SDG | Italy | 0 | European |
| HGDP01399.SDG | Russia | 0 | European |
| HGDP00797.SDG | United Kingdom | 0 | European |
| HGDP01375.SDG | France | 0 | European |
| HGDP01065.SDG | Italy | 0 | European |
| HGDP01066.SDG | Italy | 0 | European |
| HGDP00808.SDG | United Kingdom | 0 | European |
| HGDP01166.SDG | Italy | 0 | European |
| HGDP00807.SDG | United Kingdom | 0 | European |
| HGDP01387.SDG | Russia | 0 | European |
| HGDP01367.SDG | France | 0 | European |
| HGDP00534.SDG | France | 0 | European |
| HGDP00670.SDG | Italy | 0 | European |
| HGDP01071.SDG | Italy | 0 | European |
| HGDP00668.SDG | Italy | 0 | European |
| HGDP00803.SDG | United Kingdom | 0 | European |
| HGDP00794.SDG | United Kingdom | 0 | European |
| HGDP01173.SDG | Italy | 0 | European |
| HGDP00674.SDG | Italy | 0 | European |
| HGDP01068.SDG | Italy | 0 | European |
| HGDP01075.SDG | Italy | 0 | European |
| HGDP01366.SDG | France | 0 | European |
| HGDP01070.SDG | Italy | 0 | European |
| HGDP00800.SDG | United Kingdom | 0 | European |

| | | | |
|---|---|---|---|
| HGDP01171.SDG | Italy | 0 | European |
| HGDP01169.SDG | Italy | 0 | European |
| HGDP00802.SDG | United Kingdom | 0 | European |
| HGDP01149.SDG | Italy | 0 | European |
| HGDP01155.SDG | Italy | 0 | European |
| HGDP00667.SDG | Italy | 0 | European |
| HGDP01161.SDG | Italy | 0 | European |
| HGDP01063.SDG | Italy | 0 | European |
| HGDP01067.SDG | Italy | 0 | European |
| HGDP01167.SDG | Italy | 0 | European |
| HGDP01069.SDG | Italy | 0 | European |
| HGDP01152.SDG | Italy | 0 | European |
| HGDP01369.SDG | France | 0 | European |
| HGDP00810.SDG | United Kingdom | 0 | European |
| HGDP00673.SDG | Italy | 0 | European |
| HGDP01379.SDG | France | 0 | European |
| HGDP00669.SDG | Italy | 0 | European |
| HGDP01072.SDG | Italy | 0 | European |
| HGDP01162.SDG | Italy | 0 | European |
| HGDP00672.SDG | Italy | 0 | European |
| HGDP01378.SDG | France | 0 | European |
| HGDP00949.SDG | Russia | 0 | European |
| HGDP00805.SDG | United Kingdom | 0 | European |
| HGDP01373.SDG | France | 0 | European |
| HGDP01370.SDG | France | 0 | European |
| HGDP01077.SDG | Italy | 0 | European |
| HGDP01174.SDG | Italy | 0 | European |
| HGDP01372.SDG | France | 0 | European |
| HGDP01151.SDG | Italy | 0 | European |

| | | | |
|---|---|---|---|
| HGDP01377.SDG | France | 0 | European |
| HGDP00666.SDG | Italy | 0 | European |
| HGDP00959.SDG | Russia | 0 | European |
| HGDP00806.SDG | United Kingdom | 0 | European |
| HGDP01177.SDG | Italy | 0 | European |
| HGDP00804.SDG | United Kingdom | 0 | European |
| HGDP01380.SDG | France | 0 | European |
| HGDP00795.SDG | United Kingdom | 0 | European |
| HGDP01374.SDG | France | 0 | European |
| HGDP01062.SDG | Italy | 0 | European |
| HGDP00967.SDG | Russia | 0 | European |
| HGDP01064.SDG | Italy | 0 | European |
| HGDP00521.SDG | France | 0 | European |
| HGDP00665.SDG | Italy | 0 | European |
| HGDP01079.SDG | Italy | 0 | European |
| HGDP01078.SDG | Italy | 0 | European |
| HGDP01402.SDG | Russia | 0 | European |
| HGDP00903.SDG | Russia | 0 | European |
| HGDP01364.SDG | France | 0 | European |
| HGDP01153.SDG | Italy | 0 | European |
| HGDP01076.SDG | Italy | 0 | European |
| HGDP00956.SDG | Russia | 0 | European |
| HGDP00526.SDG | France | 0 | European |
| HGDP01401.SDG | Russia | 0 | European |
| HGDP01365.SDG | France | 0 | European |
| HGDP00887.SDG | Russia | 0 | European |
| HGDP00951.SDG | Russia | 0 | European |
| HGDP01172.SDG | Italy | 0 | European |
| HGDP00533.SDG | France | 0 | European |

| | | | |
|---|---|---|---|
| HGDP01163.SDG | Italy | 0 | European |
| HGDP01168.SDG | Italy | 0 | European |
| HGDP00530.SDG | France | 0 | European |
| HGDP00798.SDG | United Kingdom | 0 | European |
| HGDP00796.SDG | United Kingdom | 0 | European |
| AED106.SG | Germany | 1455 | European |
| AED1108.SG | Germany | 1469 | European |
| AED1119.SG | Germany | 1430 | European |
| AED1135b.SG | Germany | 1445 | European |
| AED125b.SG | Germany | 1498 | European |
| AED204.SG | Germany | 1455 | European |
| AED249.SG | Germany | 1455 | European |
| AED432b.SG | Germany | 1445 | European |
| AED513.SG | Germany | 1525 | European |
| AED92b.SG | Germany | 1455 | European |
| AEHIb.SG | Germany | 1503 | European |
| Alh10.SG | Germany | 1477 | European |
| Alh1.SG | Germany | 1500 | European |
| Alh2.SG | Germany | 1529 | European |
| Alh3a.SG | Germany | 1491 | European |
| BIM33.SG | Germany | 1475 | European |
| BIM37b.SG | Germany | 1350 | European |
| FN2.SG | Germany | 1650 | European |
| Ker1.SG | Crimea | 1604 | European |
| NW255a.SG | Germany | 1467 | European |
| NW54.SG | Germany | 1482 | European |
| Pr10.SG | Russia | 2200 | European |
| Pr4.SG | Russia | 2200 | European |
| STR220c.SG | Germany | 1445 | European |

| | | | |
|---|---|---|---|
| STR228.SG | Germany | 1471 | European |
| STR241.SG | Germany | 1455 | European |
| STR248b.SG | Germany | 1405 | European |
| STR266b.SG | Germany | 1430 | European |
| STR300b.SG | Germany | 1430 | European |
| STR310.SG | Germany | 1430 | European |
| STR316b.SG | Germany | 1455 | European |
| STR328c.SG | Germany | 1497 | European |
| STR355c.SG | Germany | 1430 | European |
| STR360c.SG | Germany | 1430 | European |
| STR393b.SG | Germany | 1455 | European |
| STR480.SG | Germany | 1430 | European |
| STR486.SG | Germany | 1455 | European |
| STR491.SG | Germany | 1430 | European |
| STR502b.SG | Germany | 1364 | European |
| STR535.SG | Germany | 1483 | European |
| Vim2b.SG | Serbia | 1400 | European |
| I1583.DG | Turkey | 8273 | European |
| I1583.SG | Turkey | 8273 | European |
| I11300 | Spain | 5050 | European |
| I11301 | Spain | 5050 | European |
| I11306 | Spain | 4900 | European |
| I11304_published | Spain | 4900 | European |
| I11305 | Spain | 4900 | European |
| I11303 | Spain | 4900 | European |
| I8134 | Spain | 6100 | European |
| I11912 | Romania | 4250 | European |
| I10373 | Italy | 3200 | European |
| I10372 | Italy | 3198 | European |

| | | | |
|---|---|---|---|
| I10371 | Italy | 3100 | European |
| I4064 | Italy | 6718 | European |
| I10554 | Italy | 3122 | European |
| I10892 | Spain | 950 | European |
| I10866 | Spain | 1947 | European |
| I10895 | Spain | 1038 | European |
| I10865 | Spain | 1800 | European |
| I10552 | Italy | 3234 | European |
| I10899_published | Spain | 24450 | European |
| I10553 | Italy | 3110 | European |
| I10897_published | Spain | 828 | European |
| I10854_published | Spain | 908 | European |
| I10565 | Russia | 4726 | European |
| I10564 | Russia | 4850 | European |
| I10851_published | Spain | 1006 | European |
| I10852_published | Spain | 908 | European |
| I10853_published | Spain | 876 | European |
| I12030_published | Spain | 1400 | European |
| I12031 | Spain | 1350 | European |
| I12032_published | Spain | 1350 | European |
| I12029_published | Spain | 1350 | European |
| I12033_published | Spain | 1350 | European |
| I12034 | Spain | 1350 | European |
| DA136.SG | Russia | 2300 | European |
| I11249 | Spain | 5050 | European |
| I11248 | Spain | 5050 | European |
| I11112 | Russia | 4850 | European |
| I12171_published | Spain | 2608 | European |
| I12163 | Spain | 1350 | European |

| | | | |
|---|---|---|---|
| I12173_published | Spain | 2550 | European |
| I12164_published | Spain | 1350 | European |
| I12162 | Spain | 1350 | European |
| I11752 | Russia | 4650 | European |
| I2657 | United Kingdom | 5814 | European |
| I2407_published | Poland | 4955 | European |
| TOR.11.SG | Spain | 6896 | European |
| TOR.6.SG | Spain | 6900 | European |
| TOR.7.SG | Spain | 6900 | European |
| TOR.8.SG | Spain | 6900 | European |
| VAD001 | Spain | 3641 | European |
| VAD002 | Spain | 3650 | European |
| VAD003 | Spain | 3650 | European |
| VAD004 | Spain | 3650 | European |
| VAD005 | Spain | 3590 | European |
| EHU001 | Spain | 4121 | European |
| EHU002 | Spain | 4367 | European |
| vik_2072.SG | Sweden | 973 | European |
| vik_84001.SG | Sweden | 964 | European |
| vik_84005.SG | Sweden | 900 | European |
| vik_84035.SG | Sweden | 1061 | European |
| vik_97002.SG | Sweden | 900 | European |
| vik_97026.SG | Sweden | 900 | European |
| vik_97029.SG | Sweden | 900 | European |
| vik_bns023.SG | Sweden | 900 | European |
| vik_grt035.SG | Sweden | 994 | European |
| vik_grt036.SG | Sweden | 870 | European |
| vik_gtm021.SG | Sweden | 900 | European |
| vik_gtm127.SG | Sweden | 900 | European |

| | | | |
|---|---|---|---|
| vik_KAL006.SG | Sweden | 900 | European |
| vik_kal009.SG | Sweden | 900 | European |
| vik_kls001.SG | Sweden | 900 | European |
| vik_nuf002.SG | Sweden | 900 | European |
| vik_stg020.SG | Sweden | 900 | European |
| vik_stg021.SG | Sweden | 919 | European |
| vik_stg026.SG | Sweden | 1018 | European |
| vik_urm035.SG | Sweden | 863 | European |
| vik_urm045.SG | Sweden | 900 | European |
| vik_urm160.SG | Sweden | 798 | European |
| vik_urm161.SG | Sweden | 857 | European |
| I12410 | Spain | 2375 | European |
| I12208 | Spain | 3250 | European |
| I12209 | Spain | 3250 | European |
| I12514 | Spain | 854 | European |
| I12516 | Spain | 937 | European |
| I12515 | Spain | 750 | European |
| I12878 | Spain | 2360 | European |
| I12879 | Spain | 2429 | European |
| I12877 | Spain | 2400 | European |
| I12855 | Spain | 3775 | European |
| I12809 | Spain | 3775 | European |
| I12561 | Spain | 2550 | European |
| I12560 | Spain | 2550 | European |
| I12640 | Spain | 2550 | European |
| I12641 | Spain | 2599 | European |
| I12648 | Spain | 750 | European |
| I12646 | Spain | 750 | European |
| I12649 | Spain | 750 | European |

| | | | |
|---|---|---|---|
| I12647 | Spain | 750 | European |
| I12645 | Spain | 750 | European |
| I12642 | Spain | 2443 | European |
| I12650_published | Spain | 750 | European |
| I12644 | Spain | 750 | European |
| I12643 | Spain | 2553 | European |
| Denisova11.SG | Russia | 98700 | European |
| I11902 | Romania | 5950 | European |
| I11906 | Romania | 5950 | European |
| I11929 | Hungary | 6150 | European |
| I11933 | Hungary | 6150 | European |
| I3878 | Italy | 3216 | European |
| AV1 | Hungary | 1360 | European |
| AV2 | Hungary | 1348 | European |
| CL102 | Italy | 1345 | European |
| CL110 | Italy | 1345 | European |
| CL121 | Italy | 1345 | European |
| CL145 | Italy | 1345 | European |
| CL146 | Italy | 1345 | European |
| CL151 | Italy | 1345 | European |
| CL23 | Italy | 1345 | European |
| CL25 | Italy | 1345 | European |
| CL30 | Italy | 1345 | European |
| CL31 | Italy | 1345 | European |
| CL36 | Italy | 1345 | European |
| CL38 | Italy | 1345 | European |
| CL47 | Italy | 1345 | European |
| CL49 | Italy | 1345 | European |
| CL53 | Italy | 1345 | European |

| | | | |
|---|---|---|---|
| CL57 | Italy | 1345 | European |
| CL63 | Italy | 1345 | European |
| CL83 | Italy | 1345 | European |
| CL84 | Italy | 1345 | European |
| CL87 | Italy | 1345 | European |
| CL92 | Italy | 1345 | European |
| CL93 | Italy | 1345 | European |
| CL94 | Italy | 1345 | European |
| CL97 | Italy | 1345 | European |
| SZ12 | Hungary | 1442 | European |
| SZ13 | Hungary | 1462 | European |
| SZ14 | Hungary | 1442 | European |
| SZ16 | Hungary | 1442 | European |
| SZ18 | Hungary | 1442 | European |
| SZ19 | Hungary | 1456 | European |
| SZ20 | Hungary | 1442 | European |
| SZ22 | Hungary | 1442 | European |
| SZ23 | Hungary | 1442 | European |
| SZ24 | Hungary | 1442 | European |
| SZ25 | Hungary | 1442 | European |
| SZ27 | Hungary | 1467 | European |
| SZ28 | Hungary | 1442 | European |
| SZ30 | Hungary | 1442 | European |
| SZ31 | Hungary | 1442 | European |
| SZ32 | Hungary | 1442 | European |
| SZ35 | Hungary | 1442 | European |
| SZ37 | Hungary | 1430 | European |
| SZ38 | Hungary | 1442 | European |
| SZ40 | Hungary | 1442 | European |

| | | | |
|---|---|---|---|
| SZ41 | Hungary | 1442 | European |
| SZ42 | Hungary | 1442 | European |
| SZ44 | Hungary | 1442 | European |
| SZ6 | Hungary | 1442 | European |
| SZ7 | Hungary | 1442 | European |
| SZ8 | Hungary | 1442 | European |
| SZ9 | Hungary | 1442 | European |
| SZ11.SG | Hungary | 1442 | European |
| SZ15.SG | Hungary | 1442 | European |
| SZ1.SG | Hungary | 3850 | European |
| SZ2.SG | Hungary | 1442 | European |
| SZ36.SG | Hungary | 1442 | European |
| SZ3.SG | Hungary | 1442 | European |
| SZ43.SG | Hungary | 1393 | European |
| SZ45.SG | Hungary | 1442 | European |
| SZ4.SG | Hungary | 1442 | European |
| SZ5.SG | Hungary | 1442 | European |
| chy001.SG | Russia | 1829 | European |
| chy002.SG | Russia | 1786 | European |
| cim357.SG | Moldova | 2808 | European |
| cim358.SG | Moldova | 2822 | European |
| cim359.SG | Moldova | 2875 | European |
| kzb001.SG | Russia | 3590 | European |
| kzb002.SG | Russia | 3704 | European |
| kzb003.SG | Russia | 3653 | European |
| kzb004.SG | Russia | 3661 | European |
| kzb005.SG | Russia | 3734 | European |
| kzb006.SG | Russia | 3648 | European |
| kzb007.SG | Russia | 3672 | European |

| | | | |
|---|---|---|---|
| kzb008.SG | Russia | 3754 | European |
| kzb009.SG | Russia | 3640 | European |
| mur001.SG | Russia | 3590 | European |
| mur002.SG | Russia | 3533 | European |
| mur003.SG | Russia | 3721 | European |
| mur004.SG | Russia | 3755 | European |
| scy006.WGC | Ukraine | 2550 | European |
| scy009.SG | Ukraine | 2568 | European |
| scy010.SG | Ukraine | 2606 | European |
| scy011.SG | Ukraine | 2176 | European |
| scy192.SG | Moldova | 2225 | European |
| scy193.SG | Moldova | 2225 | European |
| scy197.SG | Moldova | 2225 | European |
| scy300.SG | Moldova | 2250 | European |
| scy301.SG | Moldova | 2237 | European |
| scy303.SG | Moldova | 2231 | European |
| scy304.SG | Moldova | 2213 | European |
| scy305.SG | Moldova | 2254 | European |
| scy311.SG | Moldova | 2235 | European |
| scy332.SG | Moldova | 1616 | European |
| tem001.SG | Russia | 1725 | European |
| tem002.SG | Russia | 1761 | European |
| tem003.SG | Russia | 1698 | European |
| I3320_published | Spain | 2150 | European |
| I3321_published | Spain | 2150 | European |
| I7427_published | Spain | 1000 | European |
| N17.SG | Poland | 3850 | European |
| N18.SG | Poland | 5459 | European |
| N19.SG | Poland | 5490 | European |

| | | | |
|---|---|---|---|
| N20.SG | Poland | 5467 | European |
| N22.SG | Poland | 6263 | European |
| N25.SG | Poland | 6250 | European |
| N26.SG | Poland | 6177 | European |
| N27.SG | Poland | 6250 | European |
| N28.SG | Poland | 6074 | European |
| N31.SG | Poland | 6130 | European |
| N36.SG | Poland | 6250 | European |
| N38.SG | Poland | 4959 | European |
| N42.SG | Poland | 6049 | European |
| N44.SG | Poland | 4400 | European |
| N45.SG | Poland | 4375 | European |
| N47.SG | Poland | 4375 | European |
| N49.SG | Poland | 4375 | European |
| I14173 | Czech Republic | 5850 | European |
| I14171 | Czech Republic | 5850 | European |
| I14172 | Czech Republic | 5850 | European |
| I14176 | Czech Republic | 5550 | European |
| I14167 | Czech Republic | 5850 | European |
| I14175 | Czech Republic | 5850 | European |
| I14170 | Czech Republic | 5850 | European |
| I14169 | Czech Republic | 5850 | European |
| I14168 | Czech Republic | 5850 | European |
| I14174 | Czech Republic | 5850 | European |
| I1584 | Turkey | 5770 | European |
| I14678 | Italy | 4247 | European |
| I14308 | Russia | 6714 | European |
| I14676_published | Italy | 4053 | European |
| I14307 | Russia | 6725 | European |

| | | | |
|---|---|---|---|
| I14677 | Italy | 4329 | European |
| I14675 | Italy | 4021 | European |
| I14772 | Russia | 5625 | European |
| I14773 | Russia | 5625 | European |
| I14774 | Russia | 5625 | European |
| I14771 | Russia | 5625 | European |
| I7805 | Italy | 3860 | European |
| I16122 | Czech Republic | 5850 | European |
| I16170_published | Italy | 3337 | European |
| I16165 | Italy | 5671 | European |
| I16163 | Italy | 2576 | European |
| I16183 | Italy | 5164 | European |
| I16168 | Italy | 5807 | European |
| I16166 | Italy | 5746 | European |
| I16169 | Italy | 4177 | European |
| I16121 | Czech Republic | 5850 | European |
| I16164 | Italy | 4415 | European |
| I15943_published | Italy | 5723 | European |
| I15941 | Italy | 4196 | European |
| I15947 | Italy | 5575 | European |
| I15942 | Italy | 4297 | European |
| I15945 | Italy | 5825 | European |
| I15946 | Italy | 5968 | European |
| I15939 | Italy | 3500 | European |
| I15940_published | Italy | 4203 | European |
| MS10614.SG | Spain | 2217 | European |
| BAL003.A0101 | Spain | 11908 | European |
| BAL051_merged | Spain | 13015 | European |
| CHA001_merged | Spain | 7127 | European |

| | | | |
|---|---|---|---|
| CHA002_merged | Spain | 7127 | European |
| CHA003_merged | Spain | 7075 | European |
| CHA004.A0102 | Spain | 6420 | European |
| ELT002_merged | Spain | 5753 | European |
| ELT006_merged | Spain | 5809 | European |
| FUC003_merged | Spain | 7019 | European |
| ZBC_IPB001.B-C0101_Luk2-Pinarbasi | Turkey | 15411 | European |
| ZHAG_BON004.A0101_Luk10 | Turkey | 9900 | European |
| ZHAJ_BON034.A0101_Luk9 | Turkey | 9900 | European |
| ZHJ_BON024.A0101_Luk84 | Turkey | 10190 | European |
| ZKO_BON001.A0101_Luk7 | Turkey | 9900 | European |
| ZMOJ_BON014.A0101_Luk21 | Turkey | 9900 | European |
| Goyet_final_provisional.SG | Belgium | 42436 | European |
| Les_Cottes_final_provisional.SG | France | 42846 | European |
| Mezmaiskaya1_final_provisional.SG | Russia | 65000 | European |
| Mezmaiskaya2_final_provisional.SG | Russia | 43449 | European |
| Spy_final_provisional.SG | Belgium | 40922 | European |
| VindijaG1_final_provisional.SG | Croatia | 50000 | European |
| Chagyrskaya.SG | Russia | 80000 | European |
| I3388.DG | Russia | 4650 | European |
| I3388.SG | Russia | 4650 | European |
| I3949.DG | Russia | 4569 | European |
| I3949.SG | Russia | 4569 | European |
| I3950.DG | Russia | 4707 | European |
| I3950.SG | Russia | 4707 | European |
| I6714.DG | Russia | 4479 | European |
| I6714.SG | Russia | 4479 | European |
| I14819 | Russia | 5625 | European |

| | | | |
|---|---|---|---|
| DAV-A-8_38.SG | Iceland | 1000 | European |
| DAV-A-9_38.SG | Iceland | 950 | European |
| DKS-A-1_38.SG | Iceland | 1025 | European |
| FOV-A-1_38.SG | Iceland | 603 | European |
| FSS-A-1_38.SG | Iceland | 1015 | European |
| GRS-A-1_38.SG | Iceland | 1015 | European |
| GTE-A-1_38.SG | Iceland | 1015 | European |
| HSJ-A-1_38.SG | Iceland | 1015 | European |
| KNS-A-1_38.SG | Iceland | 950 | European |
| KOV-A-2_38.SG | Iceland | 236 | European |
| MKR-A-1_38.SG | Iceland | 1015 | European |
| NNM-A-1_38.SG | Iceland | 1015 | European |
| NTR-A-2_38.SG | Iceland | 1015 | European |
| ORE-A-1_38.SG | Iceland | 1000 | European |
| SBT-A-1_38.SG | Iceland | 1015 | European |
| SSG-A-2_38.SG | Iceland | 1025 | European |
| SSG-A-3_38.SG | Iceland | 1025 | European |
| SSG-A-4_38.SG | Iceland | 1025 | European |
| SSJ-A-2_38.SG | Iceland | 1025 | European |
| STT-A-2_38.SG | Iceland | 1000 | European |
| SVK-A-1_38.SG | Iceland | 1015 | European |
| TGS-A-1_38.SG | Iceland | 854 | European |
| TSK-A-26_38.SG | Iceland | 825 | European |
| VDP-A-5_38.SG | Iceland | 1000 | European |
| VDP-A-6_38.SG | Iceland | 1000 | European |
| VDP-A-7_38.SG | Iceland | 1000 | European |
| YGS-B-2_38.SG | Iceland | 1015 | European |
| s19_0LS03_1.SG | Estonia | 740 | European |
| s19_0LS09_1.SG | Estonia | 2389 | European |

| | | | |
|---|---|---|---|
| s19_0LS10_1.SG | Estonia | 2609 | European |
| s19_0LS11_1.SG | Estonia | 2918 | European |
| s19_IIa_1.SG | Estonia | 685 | European |
| s19_IIf_1.SG | Estonia | 575 | European |
| s19_IIg_1.SG | Estonia | 740 | European |
| s19_IIIt_1.SG | Estonia | 600 | European |
| s19_ILS01_1.SG | Estonia | 388 | European |
| s19_IVLS09KT_1.SG | Estonia | 365 | European |
| s19_V10_2.SG | Estonia | 2573 | European |
| s19_V11_1.SG | Estonia | 2229 | European |
| s19_V12_1.SG | Estonia | 2104 | European |
| s19_V14_2.SG | Estonia | 3130 | European |
| s19_V16_1.SG | Estonia | 2434 | European |
| s19_V7_1.SG | Estonia | 2572 | European |
| s19_V8_1.SG | Estonia | 2462 | European |
| s19_V9_2.SG | Estonia | 3072 | European |
| s19_VII15_1.SG | Russia | 1911 | European |
| s19_VII3_1.SG | Estonia | 2222 | European |
| s19_VII4_1.SG | Estonia | 2493 | European |
| s19_VIII5_2.SG | Russia | 1763 | European |
| s19_VIII6_1.SG | Russia | 1763 | European |
| s19_VIII7_1.SG | Russia | 1813 | European |
| s19_VIII8_1.SG | Russia | 1813 | European |
| s19_VIII9_1.SG | Russia | 1813 | European |
| s19_X02_1.SG | Estonia | 2937 | European |
| s19_X04_1.SG | Estonia | 2345 | European |
| s19_X05_1.SG | Estonia | 3049 | European |
| s19_X06_1.SG | Estonia | 3046 | European |
| s19_X07_1.SG | Estonia | 2950 | European |

| | | | |
|---|---|---|---|
| s19_X08_1.SG | Estonia | 2819 | European |
| s19_X09_1.SG | Estonia | 2740 | European |
| s19_X10_1.SG | Estonia | 3075 | European |
| s19_X11_1.SG | Estonia | 2906 | European |
| s19_X12_1.SG | Estonia | 2767 | European |
| s19_X13_1.SG | Estonia | 2583 | European |
| s19_X14_1.SG | Estonia | 2580 | European |
| s19_X15_2.SG | Estonia | 3037 | European |
| s19_X16_1.SG | Estonia | 2936 | European |
| s19_X17_2.SG | Estonia | 2819 | European |
| s19_X19_1.SG | Estonia | 2750 | European |
| s19_X20_1.SG | Estonia | 2787 | European |
| RISE1159.SG | Poland | 4724 | European |
| RISE1160.SG | Poland | 4721 | European |
| RISE1161.SG | Poland | 4785 | European |
| RISE1162.SG | Poland | 4765 | European |
| RISE1163.SG | Poland | 4736 | European |
| RISE1164.SG | Poland | 4463 | European |
| RISE1165.SG | Poland | 4754 | European |
| RISE1166.SG | Poland | 4904 | European |
| RISE1167.SG | Poland | 4721 | European |
| RISE1168.SG | Poland | 4672 | European |
| RISE1169.SG | Poland | 4747 | European |
| RISE1170.SG | Poland | 4754 | European |
| RISE1171.SG | Poland | 4826 | European |
| RISE1172.SG | Poland | 4664 | European |
| RISE1173.SG | Poland | 4635 | European |
| RISE1241.SG | Poland | 4773 | European |
| RISE1243.SG | Poland | 4725 | European |

| | | | |
|---|---|---|---|
| RISE1246.SG | Poland | 4721 | European |
| RISE1247.SG | Poland | 4725 | European |
| RISE1248.SG | Poland | 4725 | European |
| RISE1249.SG | Poland | 4737 | European |
| RISE1250.SG | Poland | 4750 | European |
| RISE1252.SG | Poland | 4725 | European |
| RISE1254.SG | Poland | 4725 | European |
| COV20126.SG | Spain | 3632 | European |
| LD1174.SG | Spain | 4466 | European |
| BOO001.A0101 | Russia | 3740 | European |
| BOO002.A0101 | Russia | 3725 | European |
| BOO003.A0101 | Russia | 3725 | European |
| BOO004.A0101 | Russia | 3725 | European |
| BOO005.A0101 | Russia | 3725 | European |
| BOO006.A0101 | Russia | 3725 | European |
| CHV001.A0101 | Russia | 150 | European |
| CHV002.A0101 | Russia | 150 | European |
| JK1963 | Finland | 1400 | European |
| JK1967 | Finland | 1400 | European |
| JK1968 | Finland | 1400 | European |
| JK1970 | Finland | 1400 | European |
| JK2065 | Finland | 1400 | European |
| JK2066 | Finland | 1400 | European |
| JK2067 | Finland | 1400 | European |
| Saami.SG | Finland | 0 | European |
| ans003.SG | Sweden | 5282 | European |
| ans005.SG | Sweden | 5367 | European |
| ans008.SG | Sweden | 5154 | European |
| ans014.SG | Sweden | 5111 | European |

| | | | |
|---|---|---|---|
| ans016.SG | Sweden | 4705 | European |
| ans017.SG | Sweden | 5053 | European |
| bal004.SG | United Kingdom | 5186 | European |
| kol2BE.SG | Czech Republic | 6494 | European |
| kol6-ALL_DATA.SG | Czech Republic | 6775 | European |
| lai001.SG | United Kingdom | 5178 | European |
| mid001.SG | United Kingdom | 5173 | European |
| mid002.SG | United Kingdom | 5410 | European |
| ARM001 | Armenia | 5417 | European |
| ARM002 | Armenia | 5163 | European |
| ARM003 | Armenia | 5163 | European |
| AY2001 | Russia | 5397 | European |
| AY2003 | Russia | 5452 | European |
| BU2001 | Russia | 4675 | European |
| GW1001 | Russia | 4718 | European |
| I1720 | Russia | 5300 | European |
| I1722 | Russia | 6408 | European |
| I1723 | Russia | 4696 | European |
| I2051 | Russia | 3275 | European |
| I2055 | Russia | 6508 | European |
| I2056 | Russia | 6466 | European |
| I6266 | Russia | 5385 | European |
| I6267 | Russia | 5410 | European |
| I6268 | Russia | 5550 | European |
| I6272 | Russia | 5040 | European |
| IV3002 | Russia | 5201 | European |
| KBD001 | Russia | 4030 | European |
| KBD002 | Russia | 4035 | European |
| KDC001 | Russia | 3838 | European |

| | | | |
|---|---|---|---|
| KDC002 | Russia | 3750 | European |
| LYG001 | Russia | 4670 | European |
| MK3003 | Russia | 4477 | European |
| MK5001 | Russia | 5150 | European |
| MK5004 | Russia | 5150 | European |
| MK5008 | Russia | 5187 | European |
| MK5009 | Russia | 4705 | European |
| NV3001 | Russia | 3945 | European |
| OSS001 | Russia | 5582 | European |
| PG2001 | Russia | 6850 | European |
| PG2002 | Russia | 4361 | European |
| PG2004 | Russia | 6088 | European |
| RK1001 | Russia | 4717 | European |
| RK1003 | Russia | 4764 | European |
| RK1007 | Russia | 5105 | European |
| RK4001 | Russia | 4238 | European |
| RK4002 | Russia | 4510 | European |
| SA6001 | Russia | 5397 | European |
| SA6002 | Russia | 5126 | European |
| SA6003 | Russia | 4318 | European |
| SA6004 | Russia | 5159 | European |
| SA6010 | Russia | 4728 | European |
| SA6013 | Russia | 5167 | European |
| SIJ002 | Russia | 5164 | European |
| SIJ003 | Russia | 5164 | European |
| VJ1001 | Russia | 6230 | European |
| ZO2002 | Russia | 4850 | European |
| DA237.SG | Finland | 1388 | European |
| Kolyma_River.SG | Russia | 9775 | European |

| | | | |
|---|---|---|---|
| M0831.SG | Russia | 3050 | European |
| M9984.SG | Russia | 3068 | European |
| NEO229.SG | Russia | 6577 | European |
| NEO230.SG | Russia | 4875 | European |
| NEO231.SG | Russia | 4665 | European |
| NEO232.SG | Russia | 4812 | European |
| NEO233.SG | Russia | 1744 | European |
| NEO234.SG | Russia | 2818 | European |
| NEO235.SG | Russia | 7659 | European |
| NEO236.SG | Russia | 7650 | European |
| NEO237.SG | Russia | 7620 | European |
| NEO238.SG | Russia | 7551 | European |
| NEO239.SG | Russia | 7543 | European |
| NEO240.SG | Russia | 7508 | European |
| NEO241.SG | Russia | 1949 | European |
| NEO242.SG | Russia | 1757 | European |
| NEO243.SG | Russia | 2209 | European |
| NEO246.SG | Russia | 860 | European |
| NEO247.SG | Russia | 1085 | European |
| NEO248.SG | Russia | 2062 | European |
| NEO249.SG | Russia | 1100 | European |
| NEO250.SG | Russia | 2142 | European |
| NEO251.SG | Russia | 1140 | European |
| NEO253.SG | Russia | 1721 | European |
| NEO298.SG | Russia | 4630 | European |
| NEO299.SG | Russia | 605 | European |
| Yana_old.SG | Russia | 31850 | European |
| Yana_old2.SG | Russia | 31850 | European |
| Yana_Young.SG | Russia | 761 | European |

| | | | |
|---|---|---|---|
| I7774_d | Italy | 3875 | European |
| I16161 | Italy | 3434 | European |
| I1012_published | Russia | 3825 | European |
| I0231_published | Russia | 4844 | European |
| LS-13.SG | Russia | 2324 | European |
| MJ-06.SG | Ukraine | 4620 | European |
| MJ-08.SG | Ukraine | 2587 | European |
| MJ-09.SG | Ukraine | 4286 | European |
| MJ-12.SG | Ukraine | 2847 | European |
| MJ-13.SG | Ukraine | 2495 | European |
| MJ-14.SG | Ukraine | 2593 | European |
| MJ-15.SG | Ukraine | 2246 | European |
| MJ-16.SG | Ukraine | 2544 | European |
| MJ-19.SG | Ukraine | 1537 | European |
| MJ-31.SG | Ukraine | 3136 | European |
| MJ-32.SG | Ukraine | 2246 | European |
| MJ-33.SG | Ukraine | 2553 | European |
| MJ-34.SG | Ukraine | 2231 | European |
| MJ-35.SG | Ukraine | 2515 | European |
| MJ-36.SG | Ukraine | 1628 | European |
| MJ-37.SG | Ukraine | 1624 | European |
| MJ-38.SG | Russia | 2016 | European |
| MJ-39.SG | Russia | 2233 | European |
| MJ-40.SG | Russia | 2254 | European |
| MJ-41.SG | Russia | 2477 | European |
| MJ-42.SG | Russia | 2604 | European |
| MJ-43.SG | Russia | 2312 | European |
| MJ-44.SG | Russia | 2213 | European |
| MJ-46.SG | Ukraine | 2220 | European |

| | | | |
|---|---|---|---|
| MJ-47.SG | Ukraine | 2429 | European |
| MJ-56.SG | Russia | 2436 | European |
| SB337B3_lib.SG | United Kingdom | 10393 | European |
| SB338B2_lib.SG | United Kingdom | 5657 | European |
| SB343C3_lib.SG | United Kingdom | 10546 | European |
| SB379D2_lib.SG | United Kingdom | 5533 | European |
| I5374.SG | United Kingdom | 4480 | European |
| I5374_contam.SG | United Kingdom | 4480 | European |
| SB415A_lib.SG | United Kingdom | 5300 | European |
| I3025.SG | United Kingdom | 9257 | European |
| SB460A3_lib.SG | United Kingdom | 9580 | European |
| SB462A2_lib.SG | United Kingdom | 5533 | European |
| I3039.SG | United Kingdom | 5525 | European |
| I3040.SG | United Kingdom | 5525 | European |
| I3041.SG | United Kingdom | 5440 | European |
| SB514B2_lib.SG | United Kingdom | 6290 | European |
| SB527A_lib.SG | United Kingdom | 5600 | European |
| SB528A_lib.SG | United Kingdom | 5600 | European |
| SB568A_lib.SG | United Kingdom | 4776 | European |
| SB570A_lib.SG | United Kingdom | 4918 | European |
| SB573A_lib.SG | United Kingdom | 5600 | European |
| SB577A_lib.SG | United Kingdom | 5200 | European |
| SB603A_lib.SG | United Kingdom | 5516 | European |
| AITI_119 | Germany | 3747 | European |
| AITI_120 | Germany | 3662 | European |
| AITI_2 | Germany | 3753 | European |
| AITI_32_d | Germany | 3728 | European |
| AITI_33_d | Germany | 3596 | European |
| AITI_36 | Germany | 3836 | European |

| | | | |
|---|---|---|---|
| AITI_37_d | Germany | 3850 | European |
| AITI_40 | Germany | 3746 | European |
| AITI_43 | Germany | 3760 | European |
| AITI_50 | Germany | 3773 | European |
| AITI_55_d | Germany | 3772 | European |
| AITI_5_d | Germany | 3764 | European |
| AITI_65adult_d | Germany | 3728 | European |
| AITI_66_d | Germany | 3761 | European |
| AITI_70 | Germany | 3901 | European |
| AITI_72 | Germany | 3776 | European |
| AITI_77A_d | Germany | 3828 | European |
| AITI_77B_d | Germany | 3743 | European |
| AITI_78 | Germany | 3748 | European |
| AITI_86_published | Germany | 3753 | European |
| AITI_87_d | Germany | 3692 | European |
| AITI_92_d | Germany | 3761 | European |
| AITI_95_d | Germany | 3671 | European |
| AITI_98 | Germany | 3605 | European |
| ALT_3_d | Germany | 4450 | European |
| ALT_4_d | Germany | 4453 | European |
| AMP_1_d | Germany | 4356 | European |
| HUGO_168_d | Germany | 4167 | European |
| HUGO_169Sk1_d | Germany | 4385 | European |
| HUGO_171_d | Germany | 4203 | European |
| HUGO_180Sk1_d | Germany | 4289 | European |
| HUGO_180Sk2_d | Germany | 4307 | European |
| HUGO_190_d | Germany | 4103 | European |
| OBKR_117_d | Germany | 3883 | European |
| OBKR_2_d | Germany | 3906 | European |

| | | | |
|---|---|---|---|
| OBKR_47_d | Germany | 4012 | European |
| OBKR_50_d | Germany | 3935 | European |
| OBKR_66_d | Germany | 3918 | European |
| OBKR_67_d | Germany | 3906 | European |
| OBKR_73_d | Germany | 3911 | European |
| OBKR_80 | Germany | 4000 | European |
| OBKR_81_d | Germany | 3910 | European |
| OBKR_86_d | Germany | 3926 | European |
| OBKR_93_d | Germany | 3933 | European |
| OBKR_95_d | Germany | 3917 | European |
| OBKR_96_d | Germany | 3863 | European |
| OBKR_9A_d | Germany | 3902 | European |
| OTTM_141A | Germany | 3515 | European |
| OTTM_141_d | Germany | 3516 | European |
| OTTM_142_d | Germany | 3527 | European |
| OTTM_151ind1_d | Germany | 3602 | European |
| OTTM_151ind2_d | Germany | 3507 | European |
| OTTM_152_d | Germany | 3513 | European |
| OTTM_154 | Germany | 3709 | European |
| OTTM_156 | Germany | 3448 | European |
| OTTM_165_d | Germany | 3572 | European |
| OTTM_81_d | Germany | 3497 | European |
| OTTM_84_d | Germany | 3503 | European |
| OTTM_87_d | Germany | 3540 | European |
| OTTM_91_d | Germany | 3283 | European |
| POST_12_d | Germany | 4036 | European |
| POST_131_d | Germany | 3950 | European |
| POST_140_d | Germany | 3944 | European |
| POST_1_d | Germany | 3886 | European |

| | | | |
|---|---|---|---|
| POST_28 | Germany | 3917 | European |
| POST_32_d | Germany | 3930 | European |
| POST_35_d | Germany | 3932 | European |
| POST_44 | Germany | 4023 | European |
| POST_47 | Germany | 3997 | European |
| POST_50 | Germany | 4045 | European |
| POST_6 | Germany | 3873 | European |
| POST_99_d | Germany | 3962 | European |
| UNTA58_147_d | Germany | 3922 | European |
| UNTA58_152_d | Germany | 3848 | European |
| UNTA58_153 | Germany | 3839 | European |
| UNTA58_67_d | Germany | 4242 | European |
| UNTA58_68Sk1 | Germany | 4250 | European |
| UNTA58_68Sk2_d | Germany | 4347 | European |
| UNTA85_1336_d | Germany | 4334 | European |
| UNTA85_1343 | Germany | 4208 | European |
| UNTA85_1412 | Germany | 3910 | European |
| WEHR_1192SkA | Germany | 4193 | European |
| WEHR_1192SkB_d | Germany | 3785 | European |
| WEHR_1193_d | Germany | 3823 | European |
| WEHR_1414_d | Germany | 3908 | European |
| WEHR_1415adult_d | Germany | 3872 | European |
| WEHR_1415child | Germany | 3902 | European |
| WEHR_1474_d | Germany | 3924 | European |
| WEHR_1564_d | Germany | 3899 | European |
| WEHR_1586_d | Germany | 3835 | European |
| R1014.SG | Italy | 7074 | European |
| R1015.SG | Italy | 2800 | European |
| R1016.SG | Italy | 2750 | European |

| | | | |
|---|---|---|---|
| R1021.SG | Italy | 2600 | European |
| R104.SG | Italy | 1319 | European |
| R105.SG | Italy | 1450 | European |
| R106.SG | Italy | 1450 | European |
| R107.SG | Italy | 1450 | European |
| R108.SG | Italy | 1450 | European |
| R109.SG | Italy | 1450 | European |
| R10.SG | Italy | 7628 | European |
| R110.SG | Italy | 1450 | European |
| R111.SG | Italy | 1850 | European |
| R113.SG | Italy | 1850 | European |
| R114.SG | Italy | 1850 | European |
| R115.SG | Italy | 1850 | European |
| R116.SG | Italy | 1850 | European |
| R117.SG | Italy | 1450 | European |
| R118.SG | Italy | 1450 | European |
| R11.SG | Italy | 11868 | European |
| R120.SG | Italy | 1450 | European |
| R1219.SG | Italy | 508 | European |
| R121.SG | Italy | 1450 | European |
| R1220.SG | Italy | 465 | European |
| R1221.SG | Italy | 465 | European |
| R1224.SG | Italy | 465 | European |
| R122.SG | Italy | 1450 | European |
| R123.SG | Italy | 1780 | European |
| R125.SG | Italy | 1750 | European |
| R126.SG | Italy | 1650 | European |
| R1283.SG | Italy | 1095 | European |
| R1285.SG | Italy | 935 | European |

| | | | |
|---|---|---|---|
| R1286.SG | Italy | 520 | European |
| R1287.SG | Italy | 525 | European |
| R1288.SG | Italy | 525 | European |
| R1289.SG | Italy | 525 | European |
| R128.SG | Italy | 1750 | European |
| R1290.SG | Italy | 602 | European |
| R130.SG | Italy | 1550 | European |
| R131.SG | Italy | 1850 | European |
| R132.SG | Italy | 1670 | European |
| R133.SG | Italy | 1550 | European |
| R134.SG | Italy | 1550 | European |
| R136.SG | Italy | 1550 | European |
| R137.SG | Italy | 1550 | European |
| R1543.SG | Italy | 1750 | European |
| R1544.SG | Italy | 1750 | European |
| R1545.SG | Italy | 1750 | European |
| R1547.SG | Italy | 1814 | European |
| R1548.SG | Italy | 1814 | European |
| R1549.SG | Italy | 1814 | European |
| R1550.SG | Italy | 1814 | European |
| R1551.SG | Italy | 1797 | European |
| R15.SG | Italy | 9086 | European |
| R16.SG | Italy | 7202 | European |
| R17.SG | Italy | 7216 | European |
| R18.SG | Italy | 7271 | European |
| R19.SG | Italy | 7146 | European |
| R1.SG | Italy | 2832 | European |
| R22.SG | Italy | 3868 | European |
| R24.SG | Italy | 5396 | European |

| | | | |
|---|---|---|---|
| R25.SG | Italy | 4081 | European |
| R26.SG | Italy | 4347 | European |
| R27.SG | Italy | 4150 | European |
| R28.SG | Italy | 4185 | European |
| R29.SG | Italy | 4150 | European |
| R2.SG | Italy | 7981 | European |
| R30.SG | Italy | 1450 | European |
| R31.SG | Italy | 1538 | European |
| R32.SG | Italy | 1450 | European |
| R33.SG | Italy | 1450 | European |
| R34.SG | Italy | 1450 | European |
| R35.SG | Italy | 1450 | European |
| R36.SG | Italy | 1450 | European |
| R37.SG | Italy | 1750 | European |
| R38.SG | Italy | 1750 | European |
| R39.SG | Italy | 1798 | European |
| R3.SG | Italy | 7716 | European |
| R40.SG | Italy | 1750 | European |
| R41.SG | Italy | 1750 | European |
| R42.SG | Italy | 1863 | European |
| R435.SG | Italy | 2350 | European |
| R436.SG | Italy | 1850 | European |
| R437.SG | Italy | 2250 | European |
| R43.SG | Italy | 1750 | European |
| R44.SG | Italy | 1750 | European |
| R45.SG | Italy | 1750 | European |
| R473.SG | Italy | 2600 | European |
| R474.SG | Italy | 2600 | European |
| R475.SG | Italy | 2600 | European |

| | | | |
|---|---|---|---|
| R47.SG | Italy | 1655 | European |
| R49.SG | Italy | 1750 | European |
| R4.SG | Italy | 4841 | European |
| R50.SG | Italy | 1699 | European |
| R51.SG | Italy | 1750 | European |
| R52.SG | Italy | 595 | European |
| R53.SG | Italy | 595 | European |
| R54.SG | Italy | 595 | European |
| R55.SG | Italy | 595 | European |
| R56.SG | Italy | 595 | European |
| R57.SG | Italy | 595 | European |
| R58.SG | Italy | 850 | European |
| R59.SG | Italy | 1045 | European |
| R5.SG | Italy | 4832 | European |
| R60.SG | Italy | 1045 | European |
| R61.SG | Italy | 595 | European |
| R62.SG | Italy | 735 | European |
| R63.SG | Italy | 960 | European |
| R64.SG | Italy | 840 | European |
| R65.SG | Italy | 840 | European |
| R66.SG | Italy | 1750 | European |
| R67.SG | Italy | 1750 | European |
| R68.SG | Italy | 1750 | European |
| R69.SG | Italy | 1750 | European |
| R6.SG | Italy | 7202 | European |
| R70.SG | Italy | 1750 | European |
| R71.SG | Italy | 1750 | European |
| R72.SG | Italy | 1750 | European |
| R73.SG | Italy | 1750 | European |

| | | | |
|---|---|---|---|
| R75.SG | Italy | 1850 | European |
| R76.SG | Italy | 1850 | European |
| R78.SG | Italy | 1850 | European |
| R7.SG | Italy | 10706 | European |
| R80.SG | Italy | 1840 | European |
| R81.SG | Italy | 1850 | European |
| R835.SG | Italy | 1814 | European |
| R836.SG | Italy | 1814 | European |
| R850.SG | Italy | 2600 | European |
| R851.SG | Italy | 2600 | European |
| R8.SG | Italy | 7708 | European |
| R969.SG | Italy | 300 | European |
| R970.SG | Italy | 300 | European |
| R973.SG | Italy | 300 | European |
| R9.SG | Italy | 7481 | European |
| I8571_d | Spain | 3350 | European |
| I4556 | Spain | 2550 | European |
| I3025_all.SG | United Kingdom | 9257 | European |
| I3033_all.SG | United Kingdom | 5533 | European |
| I3039_all.SG | United Kingdom | 5525 | European |
| I3040_all.SG | United Kingdom | 5525 | European |
| I3041_all.SG | United Kingdom | 5440 | European |
| I3065.SG | United Kingdom | 6290 | European |
| I5387_all.SG | United Kingdom | 4850 | European |
| I5387.SG | United Kingdom | 4850 | European |
| I6746_all.SG | United Kingdom | 5526 | European |
| I6746.SG | United Kingdom | 5526 | European |
| I6747_all.SG | United Kingdom | 5528 | European |
| I6747.SG | United Kingdom | 5528 | European |

| | | | |
|---|---|---|---|
| I6750_all.SG | United Kingdom | 5650 | European |
| I6750.SG | United Kingdom | 5650 | European |
| I6751_all.SG | United Kingdom | 5650 | European |
| I6751.SG | United Kingdom | 5650 | European |
| I6753_all.SG | United Kingdom | 5305 | European |
| I6753.SG | United Kingdom | 5305 | European |
| I6755_all.SG | United Kingdom | 5528 | European |
| I6755.SG | United Kingdom | 5528 | European |
| I6757.SG | United Kingdom | 5493 | European |
| I6759.SG | United Kingdom | 5527 | European |
| I6760_all.SG | United Kingdom | 5781 | European |
| I6760.SG | United Kingdom | 5781 | European |
| I6761.SG | United Kingdom | 5525 | European |
| I6762_all.SG | United Kingdom | 5700 | European |
| I6762.SG | United Kingdom | 5700 | European |
| I6764.SG | United Kingdom | 5100 | European |
| I6766.SG | United Kingdom | 5100 | European |
| I6767_all.SG | United Kingdom | 10280 | European |
| I6767.SG | United Kingdom | 10280 | European |
| I4383 | Italy | 3950 | European |
| I6913_published | Austria | 7366 | European |
| I12221_published | Italy | 1005 | European |
| I12223 | Italy | 1597 | European |
| I12220_published | Italy | 1306 | European |
| I6912_all_published | Austria | 7088 | European |
| I10502_published | Italy | 3151 | European |
| I0099_published | Germany | 3024 | European |
| I0047_published | Germany | 3923 | European |
| I1579_published | Turkey | 8072 | European |

| | | | |
|---|---|---|---|
| I1581_published | Turkey | 8252 | European |
| I1580_published | Turkey | 8205 | European |
| I1585_published | Turkey | 8021 | European |
| I1583_published | Turkey | 8273 | European |
| I0727_published | Turkey | 7950 | European |
| I0724_published | Turkey | 7950 | European |
| I0581_published_d | Spain | 4746 | European |
| I0854_published | Turkey | 8104 | European |
| I0723_published | Turkey | 7870 | European |
| I2739_published | Hungary | 7158 | European |
| I2790_published | Hungary | 5625 | European |
| I2791_published | Hungary | 5523 | European |
| I2793_published | Hungary | 6273 | European |
| I0235 | Russia | 3675 | European |
| I0371_published | Russia | 4678 | European |
| I0060_published | Germany | 4214 | European |
| I1298_published | Bulgaria | 7883 | European |
| I1895_published | Hungary | 7195 | European |
| I1902_published | Hungary | 6550 | European |
| I1905_published | Hungary | 6664 | European |
| I1907_published | Hungary | 6183 | European |
| I1878_published | Hungary | 7687 | European |
| I2366_published | Hungary | 5118 | European |
| I2367_published | Hungary | 5085 | European |
| I2384_published | Hungary | 7114 | European |
| I0046_published | Germany | 7041 | European |
| I0659_published | Germany | 7031 | European |
| I0048_published | Germany | 7057 | European |
| I1550_published | Germany | 7125 | European |

| | | | |
|---|---|---|---|
| I0821_published | Germany | 6938 | European |
| I0057_published | Germany | 7070 | European |
| I2005_published | Germany | 7100 | European |
| I2008_published | Germany | 7041 | European |
| I2016_published | Germany | 7125 | European |
| I2017_published | Germany | 7125 | European |
| I2021_published | Germany | 7125 | European |
| I2026_published | Germany | 7125 | European |
| I2029_published | Germany | 7100 | European |
| I2030_published | Germany | 7125 | European |
| I2032_published | Germany | 7125 | European |
| I2036_published | Germany | 7125 | European |
| I2037_published | Germany | 7050 | European |
| I2038_published | Germany | 7125 | European |
| I1880 | Hungary | 7600 | European |
| I0166 | Germany | 6400 | European |
| I3273_published | Spain | 5403 | European |
| I3496_published | Spain | 2200 | European |
| I3719_published | Ukraine | 6831 | European |
| I3276_published | Spain | 4955 | European |
| I5436 | Romania | 9025 | European |
| I6626_published | Spain | 4100 | European |
| I8294_published | Russia | 795 | European |
| I8295_published | Russia | 4500 | European |
| I8296_published | Russia | 4665 | European |
| I8298_published | Russia | 5690 | European |
| I5884_published | Ukraine | 4735 | European |
| I2470_published | Spain | 3275 | European |
| I6491_published | Spain | 1600 | European |

| | | | |
|---|---|---|---|
| I10287_published | Spain | 4900 | European |
| I8140_published_d | Spain | 3895 | European |
| ajv54.SG | Sweden | 4849 | European |
| ber1M.SG | Sweden | 4483 | European |
| ber2F.SG | Sweden | 4492 | European |
| EKA1.SG | Estonia | 4195 | European |
| oll007.SG | Sweden | 4617 | European |
| oll009.SG | Sweden | 3787 | European |
| oll010.SG | Sweden | 3704 | European |
| poz44.SG | Poland | 4663 | European |
| poz81.SG | Poland | 4702 | European |
| ros005_renamed.SG | Sweden | 4955 | European |
| ros3.SG | Sweden | 5084 | European |
| AMC001 | Italy | 1772 | European |
| AMC005 | Italy | 1775 | European |
| AMC014 | Italy | 1745 | European |
| COR001 | Italy | 1017 | European |
| COR002 | Italy | 1106 | European |
| FIL004 | Italy | 5155 | European |
| S1249 | Italy | 3650 | European |
| S1250 | Italy | 3650 | European |
| S1252 | Italy | 3650 | European |
| S1253 | Italy | 3650 | European |
| ISB001 | Italy | 3735 | European |
| ISC001 | Italy | 4175 | European |
| LON001 | Italy | 3775 | European |
| LON003 | Italy | 6274 | European |
| MA110 | Italy | 3097 | European |
| MA112_renamed | Italy | 3111 | European |

| | | | |
|---|---|---|---|
| MA138 | Italy | 3096 | European |
| MA78 | Italy | 3990 | European |
| MA79 | Italy | 6045 | European |
| MA82 | Italy | 3427 | European |
| MA85 | Italy | 3881 | European |
| MA87 | Italy | 2988 | European |
| MA88 | Italy | 4177 | European |
| MA89 | Italy | 5198 | European |
| MSR002 | Italy | 2674 | European |
| MSR003 | Italy | 2500 | European |
| NOE001 | Italy | 6090 | European |
| NOE002 | Italy | 6132 | European |
| ORC001 | Italy | 3163 | European |
| ORC002 | Italy | 2249 | European |
| ORC003 | Italy | 3181 | European |
| ORC004 | Italy | 3226 | European |
| ORC005 | Italy | 3140 | European |
| ORC006 | Italy | 3123 | European |
| ORC007 | Italy | 3173 | European |
| ORC008 | Italy | 3023 | European |
| ORC009 | Italy | 3062 | European |
| PJU002 | Italy | 4145 | European |
| SEC001 | Italy | 4237 | European |
| SEC002 | Italy | 4352 | European |
| SEC004 | Italy | 4337 | European |
| SEC005 | Italy | 4348 | European |
| SEC006 | Italy | 4331 | European |
| SID005 | Italy | 6095 | European |
| SID006 | Italy | 4663 | European |

| | | | |
|---|---|---|---|
| SNN001 | Italy | 603 | European |
| SNN002 | Italy | 604 | European |
| SNN003 | Italy | 481 | European |
| SNN004 | Italy | 674 | European |
| SUA002 | Italy | 4076 | European |
| SUA003 | Italy | 3112 | European |
| SUA005 | Italy | 4170 | European |
| SUA006 | Italy | 3128 | European |
| SUA007 | Italy | 3904 | European |
| SUC001 | Italy | 4020 | European |
| SUC002 | Italy | 4081 | European |
| SUC003 | Italy | 3963 | European |
| SUC004 | Italy | 4185 | European |
| SUC005 | Italy | 4146 | European |
| SUC006 | Italy | 4347 | European |
| SUC007 | Italy | 4321 | European |
| SUC008 | Italy | 5817 | European |
| SUC009 | Italy | 3667 | European |
| VIL004 | Italy | 2381 | European |
| VIL006 | Italy | 2622 | European |
| VIL007 | Italy | 2409 | European |
| VIL009 | Italy | 2400 | European |
| VIL010 | Italy | 2741 | European |
| VIL011 | Italy | 2754 | European |
| I2357_all | Hungary | 7050 | European |
| I20767 | Hungary | 2100 | European |
| I20776 | Ukraine | 6950 | European |
| I20766 | Hungary | 2100 | European |
| I20770 | Hungary | 3050 | European |

| | | | |
|---|---|---|---|
| I20773 | Hungary | 3200 | European |
| I20748 | Hungary | 3050 | European |
| I20747 | Hungary | 2100 | European |
| NIEcap9 | Germany | 1345 | European |
| NIEcap2 | Germany | 1345 | European |
| NIEcap1 | Germany | 1345 | European |
| NIEcap10 | Germany | 1345 | European |
| NIEcap3a | Germany | 1345 | European |
| NIEcap12a | Germany | 1345 | European |
| NIEcap3c | Germany | 1345 | European |
| NIEcap5 | Germany | 1345 | European |
| NIEcap6 | Germany | 1345 | European |
| NIEcap3b | Germany | 1345 | European |
| NIEcap12b | Germany | 1345 | European |
| NIEcap12c | Germany | 1345 | European |
| I15944_d | Italy | 5575 | European |
| Aes1 | Switzerland | 4939 | European |
| Aes10 | Switzerland | 4833 | European |
| Aes11 | Switzerland | 4885 | European |
| Aes12 | Switzerland | 4864 | European |
| Aes13 | Switzerland | 4902 | European |
| Aes14 | Switzerland | 4897 | European |
| Aes15 | Switzerland | 4899 | European |
| Aes16 | Switzerland | 4942 | European |
| Aes17 | Switzerland | 4878 | European |
| Aes18 | Switzerland | 4838 | European |
| Aes19 | Switzerland | 4825 | European |
| Aes2 | Switzerland | 4885 | European |
| Aes20 | Switzerland | 4842 | European |

| | | | |
|---|---|---|---|
| Aes21 | Switzerland | 4825 | European |
| Aes22 | Switzerland | 4740 | European |
| Aes23 | Switzerland | 4722 | European |
| Aes24 | Switzerland | 4839 | European |
| Aes25 | Switzerland | 4635 | European |
| Aes3 | Switzerland | 4919 | European |
| Aes4 | Switzerland | 4970 | European |
| Aes5 | Switzerland | 4906 | European |
| Aes8 | Switzerland | 4851 | European |
| Aes9 | Switzerland | 4848 | European |
| MX150 | Switzerland | 5164 | European |
| MX182 | Switzerland | 5158 | European |
| MX183 | Switzerland | 5160 | European |
| MX184 | Switzerland | 4950 | European |
| MX187 | Switzerland | 5055 | European |
| MX188 | Switzerland | 4399 | European |
| MX189 | Switzerland | 4058 | European |
| MX190 | Switzerland | 4530 | European |
| MX191 | Switzerland | 4523 | European |
| MX192 | Switzerland | 4470 | European |
| MX193 | Switzerland | 4087 | European |
| MX195 | Switzerland | 4225 | European |
| MX196 | Switzerland | 4495 | European |
| MX197 | Switzerland | 4351 | European |
| MX198 | Switzerland | 4509 | European |
| MX199 | Switzerland | 4235 | European |
| MX203 | Switzerland | 4950 | European |
| MX204 | Switzerland | 4950 | European |
| MX209 | Switzerland | 4950 | European |

| | | | |
|---|---|---|---|
| MX210 | Switzerland | 4950 | European |
| MX211 | Switzerland | 4992 | European |
| MX212 | Switzerland | 5165 | European |
| MX213 | Switzerland | 5125 | European |
| MX219 | Switzerland | 5130 | European |
| MX251 | Germany | 4039 | European |
| MX252 | Germany | 3808 | European |
| MX254 | Germany | 3900 | European |
| MX256 | Germany | 3963 | European |
| MX257 | Germany | 3742 | European |
| MX258 | Germany | 3917 | European |
| MX259 | Germany | 4246 | European |
| MX265 | Germany | 2569 | European |
| MX270 | Germany | 3900 | European |
| MX275 | Germany | 4002 | European |
| MX277 | Germany | 3791 | European |
| MX279 | Germany | 3758 | European |
| MX280 | Germany | 3931 | European |
| MX283 | Germany | 3946 | European |
| MX286 | Germany | 3913 | European |
| MX288 | Germany | 4051 | European |
| MX298 | Switzerland | 4461 | European |
| MX299 | Switzerland | 4757 | European |
| MX304 | Switzerland | 4675 | European |
| MX310 | Switzerland | 4672 | European |
| RA42 | Switzerland | 5165 | European |
| RA43 | Switzerland | 5122 | European |
| RA44 | Switzerland | 4828 | European |
| RA45 | Switzerland | 4975 | European |

| | | | |
|---|---|---|---|
| RA54 | Switzerland | 4498 | European |
| RA57 | Switzerland | 4806 | European |
| RA58 | Switzerland | 4865 | European |
| RA59.Furtwangler | Switzerland | 4830 | European |
| RA61 | Switzerland | 4804 | European |
| RA62 | Switzerland | 4853 | European |
| RA63 | Switzerland | 3970 | European |
| RA64 | Switzerland | 4099 | European |
| TU876_SX10 | Switzerland | 4550 | European |
| TU877_SX11 | Switzerland | 6355 | European |
| TU904_SX17 | Switzerland | 4344 | European |
| TU905_SX18 | Switzerland | 2032 | European |
| TU907_SX20 | Switzerland | 3576 | European |
| TU908_SX21 | Switzerland | 4327 | European |
| TU910_SX22 | Switzerland | 4949 | European |
| TU911_SX23 | Switzerland | 3762 | European |
| TU914_SX26 | Switzerland | 4322 | European |
| TU915_SX29 | France | 6600 | European |
| TU916_SX30 | France | 6662 | European |
| TU918_SX32 | France | 4289 | European |
| TU919_SX33 | France | 6625 | European |
| TU874_SX8 | Switzerland | 4573 | European |
| TU875_SX9 | Switzerland | 6338 | European |
| pcw040.SG | Poland | 4400 | European |
| pcw041.SG | Poland | 4400 | European |
| pcw061.SG | Poland | 4321 | European |
| pcw062.SG | Poland | 4300 | European |
| pcw070.SG | Poland | 4321 | European |
| pcw110.SG | Poland | 4300 | European |

| | | | |
|---|---|---|---|
| pcw160.SG | Poland | 4299 | European |
| pcw191.SG | Poland | 4237 | European |
| pcw211.SG | Poland | 4300 | European |
| pcw212.SG | Poland | 4460 | European |
| pcw250.SG | Poland | 4300 | European |
| pcw260.SG | Poland | 4237 | European |
| pcw270.SG | Poland | 4250 | European |
| pcw280.SG | Poland | 4250 | European |
| pcw350.SG | Poland | 4293 | European |
| pcw361.SG | Poland | 4300 | European |
| pcw362.SG | Poland | 4305 | European |
| pcw420.SG | Poland | 4300 | European |
| pcw430.SG | Poland | 4300 | European |
| ALA002 | Turkey | 3388 | European |
| ALA004 | Turkey | 3773 | European |
| ALA008 | Turkey | 3752 | European |
| ALA009 | Turkey | 3699 | European |
| ALA011 | Turkey | 3620 | European |
| ALA013 | Turkey | 3734 | European |
| ALA015 | Turkey | 3861 | European |
| ALA016 | Turkey | 3503 | European |
| ALA017 | Turkey | 3478 | European |
| ALA018 | Turkey | 3382 | European |
| ALA019 | Turkey | 3514 | European |
| ALA020 | Turkey | 3395 | European |
| ALA023 | Turkey | 3785 | European |
| ALA024 | Turkey | 3889 | European |
| ALA025 | Turkey | 3711 | European |
| ALA026 | Turkey | 3627 | European |

| | | | |
|---|---|---|---|
| ALA028 | Turkey | 3705 | European |
| ALA029 | Turkey | 3743 | European |
| ALA030 | Turkey | 3467 | European |
| ALA034 | Turkey | 3697 | European |
| ALA035 | Turkey | 3820 | European |
| ALA037 | Turkey | 3754 | European |
| ALA038 | Turkey | 3473 | European |
| ALA039 | Turkey | 3337 | European |
| ALA084 | Turkey | 3844 | European |
| ALA095 | Turkey | 3780 | European |
| ART001 | Turkey | 4342 | European |
| ART004 | Turkey | 5632 | European |
| ART005 | Turkey | 5650 | European |
| ART009 | Turkey | 4553 | European |
| ART010 | Turkey | 4625 | European |
| ART011 | Turkey | 4640 | European |
| ART012 | Turkey | 5159 | European |
| ART014 | Turkey | 5255 | European |
| ART015 | Turkey | 5212 | European |
| ART017 | Turkey | 5166 | European |
| ART018 | Turkey | 5260 | European |
| ART019 | Turkey | 5393 | European |
| ART022 | Turkey | 5413 | European |
| ART023 | Turkey | 5227 | European |
| ART024 | Turkey | 5384 | European |
| ART026 | Turkey | 5163 | European |
| ART032 | Turkey | 5254 | European |
| ART038 | Turkey | 5175 | European |
| ART039 | Turkey | 5638 | European |

| | | | |
|---|---|---|---|
| ART042 | Turkey | 5767 | European |
| CBT001 | Turkey | 5446 | European |
| CBT002 | Turkey | 5529 | European |
| CBT003 | Turkey | 5450 | European |
| CBT004 | Turkey | 5526 | European |
| CBT005 | Turkey | 5415 | European |
| CBT010 | Turkey | 5480 | European |
| CBT011 | Turkey | 5480 | European |
| CBT013 | Turkey | 5526 | European |
| CBT014 | Turkey | 5518 | European |
| CBT015 | Turkey | 5525 | European |
| CBT016 | Turkey | 5538 | European |
| CBT017 | Turkey | 5480 | European |
| CBT018 | Turkey | 7516 | European |
| IKI002 | Turkey | 5165 | European |
| IKI006 | Turkey | 5250 | European |
| IKI009 | Turkey | 5206 | European |
| IKI012 | Turkey | 5218 | European |
| IKI016 | Turkey | 5396 | European |
| IKI017 | Turkey | 5284 | European |
| IKI019 | Turkey | 5250 | European |
| IKI020 | Turkey | 5250 | European |
| IKI024 | Turkey | 5821 | European |
| IKI030 | Turkey | 5397 | European |
| IKI032 | Turkey | 5250 | European |
| IKI034 | Turkey | 5392 | European |
| IKI036 | Turkey | 5405 | European |
| IKI037 | Turkey | 5496 | European |
| IKI038 | Turkey | 5480 | European |

| | | | |
|---|---|---|---|
| KRD001 | Turkey | 7629 | European |
| KRD002 | Turkey | 6889 | European |
| KRD003 | Turkey | 7602 | European |
| KRD004 | Turkey | 7620 | European |
| KRD005 | Turkey | 7661 | European |
| KRD006 | Turkey | 7550 | European |
| TIT003 | Turkey | 4100 | European |
| TIT012 | Turkey | 4100 | European |
| TIT014 | Turkey | 4100 | European |
| TIT015 | Turkey | 4100 | European |
| TIT019 | Turkey | 4100 | European |
| TIT025 | Turkey | 4100 | European |
| I20749 | Hungary | 3200 | European |
| I20750 | Hungary | 3900 | European |
| I20751 | Hungary | 3200 | European |
| I20752 | Hungary | 2201 | European |
| I20756 | Russia | 860 | European |
| ATT26.SG | France | 2200 | European |
| BERG02-2.SG | France | 5867 | European |
| BERG157-2.SG | France | 6050 | European |
| BERG157-7.SG | France | 6112 | European |
| BES1248.SG | France | 2450 | European |
| BFM265.SG | France | 2175 | European |
| BIS130.SG | France | 3850 | European |
| BIS385.SG | France | 3850 | European |
| BLP10.SG | France | 6065 | European |
| BUCH2.SG | France | 6250 | European |
| CBV95.SG | France | 4447 | European |
| COL11.SG | France | 2290 | European |

| | | | |
|---|---|---|---|
| COL153A.SG | France | 2290 | European |
| COL153i.SG | France | 2290 | European |
| CRE20D.SG | France | 6250 | European |
| ERS1164.SG | France | 2200 | European |
| ERS86.SG | France | 2200 | European |
| ERS88.SG | France | 2200 | European |
| Es97-1.SG | France | 6500 | European |
| EUG11.SG | France | 3882 | European |
| Jeb8.SG | France | 2450 | European |
| MDV248.SG | France | 6963 | European |
| Mor6.SG | France | 7019 | European |
| NIED.SG | France | 2825 | European |
| NOR2B6.SG | France | 2700 | European |
| NOR3-15.SG | France | 2700 | European |
| NOR3-6.SG | France | 2500 | European |
| NOR4.SG | France | 2500 | European |
| OBE3626-1.SG | France | 3774 | European |
| OBE3722.SG | France | 3750 | European |
| PECH5.SG | France | 2400 | European |
| PECH8.SG | France | 2400 | European |
| PEI10.SG | France | 4736 | European |
| PEI2.SG | France | 4750 | European |
| PER503.SG | France | 9050 | European |
| PER3023.SG | France | 9050 | European |
| PER3123.SG | France | 9050 | European |
| PEY163.SG | France | 2250 | European |
| PEY53.SG | France | 2250 | European |
| PIR3037AB.SG | France | 3995 | European |
| PIR3116B.SG | France | 4170 | European |

| | | | |
|---|---|---|---|
| Pir4.SG | France | 6293 | European |
| Pir6.SG | France | 3584 | European |
| PSS4170.SG | France | 4030 | European |
| PSS4693.SG | France | 5396 | European |
| PT2.SG | France | 2375 | European |
| QUIN234.SG | France | 3600 | European |
| QUIN58.SG | France | 3600 | European |
| RIX15.SG | France | 3600 | European |
| RIX2.SG | France | 3600 | European |
| RIX4.SG | France | 3664 | European |
| ROS102.SG | France | 6600 | European |
| ROS45.SG | France | 6644 | European |
| ROS78.SG | France | 6600 | European |
| ROS82.SG | France | 6619 | European |
| Schw72-15.SG | France | 7025 | European |
| Schw432.SG | France | 7031 | European |
| WET370.SG | France | 5494 | European |
| BDB001 | Germany | 8600 | European |
| BOT004 | Germany | 7550 | European |
| BOT005 | Germany | 7548 | European |
| FLR001 | France | 6414 | European |
| FLR002 | France | 5972 | European |
| FLR003 | France | 6376 | European |
| FLR004 | France | 6512 | European |
| FLR005 | France | 6301 | European |
| FLR007 | France | 6326 | European |
| FLR010 | France | 6345 | European |
| FLR013 | France | 6356 | European |
| FLR014 | France | 6426 | European |

| | | | |
|---|---|---|---|
| GRG003 | France | 6450 | European |
| GRG008 | France | 6450 | European |
| GRG015 | France | 6450 | European |
| GRG016 | France | 6407 | European |
| GRG018 | France | 6593 | European |
| GRG019 | France | 6450 | European |
| GRG021 | France | 6450 | European |
| GRG022 | France | 6450 | European |
| GRG023 | France | 6450 | European |
| GRG025 | France | 6450 | European |
| GRG027 | France | 6690 | European |
| GRG028 | France | 6671 | European |
| GRG032 | France | 6088 | European |
| GRG035 | France | 6450 | European |
| GRG041 | France | 6450 | European |
| GRG043 | France | 6450 | European |
| GRG047 | France | 6450 | European |
| GRG049 | France | 6571 | European |
| GRG050 | France | 6450 | European |
| GRG052 | France | 6450 | European |
| GRG056 | France | 6450 | European |
| GRG057 | France | 6615 | European |
| HBS002 | Germany | 6975 | European |
| HBS004 | Germany | 6975 | European |
| HBS005 | Germany | 6975 | European |
| HBS006 | Germany | 6975 | European |
| HBS007 | Germany | 6975 | European |
| HBS008 | Germany | 6975 | European |
| HBS009 | Germany | 6975 | European |

| | | | |
|---|---|---|---|
| LBR001 | France | 6740 | European |
| LBR002 | France | 6991 | European |
| LBR003 | France | 6680 | European |
| LBR004 | France | 6797 | European |
| LBR005 | France | 7051 | European |
| OBN001 | France | 6609 | European |
| OBN002 | France | 6700 | European |
| OBN003 | France | 6544 | European |
| OBN004 | France | 6528 | European |
| OBN005 | France | 6888 | European |
| OBN006 | France | 7026 | European |
| OBN007 | France | 6377 | European |
| OBN008 | France | 6543 | European |
| OBN009 | France | 6398 | European |
| OBN010 | France | 6700 | European |
| OBN011 | France | 6549 | European |
| PEN001_real1 | France | 7361 | European |
| PEN001_real2 | France | 7359 | European |
| PEN003 | France | 7363 | European |
| PRI001 | France | 6218 | European |
| PRI005 | France | 6150 | European |
| PRI006 | France | 6150 | European |
| SCH001 | Germany | 7050 | European |
| SCH004 | Germany | 7050 | European |
| SCH007 | Germany | 7050 | European |
| SCH009 | Germany | 7050 | European |
| SCH010 | Germany | 7050 | European |
| SCH011 | Germany | 7050 | European |
| SCH014 | Germany | 7050 | European |

| | | | |
|---|---|---|---|
| SCH015 | Germany | 7050 | European |
| SCH016 | Germany | 7091 | European |
| SCH018 | Germany | 7045 | European |
| SMH004 | Germany | 7116 | European |
| TGM008 | Germany | 5107 | European |
| TGM009 | Germany | 5208 | European |
| XN164 | Germany | 7110 | European |
| XN165 | Germany | 7099 | European |
| XN166 | Germany | 7100 | European |
| XN167 | Germany | 7176 | European |
| XN168 | Germany | 7093 | European |
| XN169 | Germany | 7081 | European |
| XN170 | Germany | 7024 | European |
| XN171 | Germany | 7106 | European |
| XN172 | Germany | 7101 | European |
| XN173 | Germany | 7060 | European |
| XN174 | Germany | 6800 | European |
| XN175 | Germany | 7092 | European |
| XN178 | Germany | 7100 | European |
| XN180 | Germany | 7068 | European |
| XN182 | Germany | 7069 | European |
| XN183 | Germany | 7152 | European |
| XN188 | Germany | 7130 | European |
| XN191 | Germany | 7199 | European |
| XN205 | Germany | 7100 | European |
| XN206 | Germany | 7086 | European |
| XN207 | Germany | 7100 | European |
| XN211 | Germany | 7068 | European |
| XN215 | Germany | 7010 | European |

| | | | |
|---|---|---|---|
| XN224 | Germany | 7091 | European |
| XN225 | Germany | 7100 | European |
| ANG001 | Russia | 7274 | European |
| BZK002 | Russia | 4709 | European |
| GLZ001 | Russia | 4556 | European |
| GLZ002 | Russia | 4430 | European |
| GLZ003 | Russia | 4478 | European |
| IUO001 | Russia | 6905 | European |
| KAG001 | Russia | 6856 | European |
| KAG002 | Russia | 3748 | European |
| KPT001 | Russia | 3759 | European |
| KPT002_petrous | Russia | 3931 | European |
| KPT002_tooth | Russia | 3931 | European |
| KPT003 | Russia | 3945 | European |
| KPT004 | Russia | 3773 | European |
| KPT005 | Russia | 4034 | European |
| KPT006 | Russia | 3614 | European |
| UKY001 | Russia | 13929 | European |
| STB001 | Russia | 4604 | European |
| STB002 | Russia | 6587 | European |
| ZPL001 | Russia | 3951 | European |
| ZPL002 | Russia | 4168 | European |
| VK101.SG | Iceland | 850 | European |
| VK102.SG | Iceland | 850 | European |
| VK108.SG | Sweden | 950 | European |
| VK110.SG | Iceland | 850 | European |
| VK111.SG | Iceland | 850 | European |
| VK113.SG | Norway | 750 | European |
| VK114.SG | Norway | 750 | European |

| | | | |
|---|---|---|---|
| VK116.SG | Norway | 750 | European |
| VK117.SG | Norway | 750 | European |
| VK118.SG | Norway | 750 | European |
| VK11.SG | Greenland | 950 | European |
| VK122.SG | Iceland | 850 | European |
| VK123.SG | Iceland | 850 | European |
| VK124.SG | Norway | 750 | European |
| VK125.SG | Norway | 750 | European |
| VK127.SG | Iceland | 1000 | European |
| VK128.SG | Iceland | 1000 | European |
| VK129.SG | Iceland | 1000 | European |
| VK133.SG | Denmark | 1050 | European |
| VK134.SG | Denmark | 1000 | European |
| VK135.SG | Denmark | 1000 | European |
| VK138.SG | Denmark | 1000 | European |
| VK139.SG | Denmark | 1000 | European |
| VK140.SG | Denmark | 1000 | European |
| VK141.SG | Denmark | 1000 | European |
| VK143.SG | United Kingdom | 1010 | European |
| VK144.SG | United Kingdom | 1010 | European |
| VK145.SG | United Kingdom | 1010 | European |
| VK146.SG | United Kingdom | 1010 | European |
| VK147.SG | United Kingdom | 1010 | European |
| VK148.SG | United Kingdom | 1010 | European |
| VK149.SG | United Kingdom | 1010 | European |
| VK14.SG | Russia | 900 | European |
| VK150.SG | United Kingdom | 1010 | European |
| VK151.SG | United Kingdom | 1010 | European |
| VK153.SG | Poland | 950 | European |

| | | | |
|---|---|---|---|
| VK154.SG | Poland | 950 | European |
| VK155.SG | Poland | 950 | European |
| VK156.SG | Poland | 950 | European |
| VK157.SG | Poland | 950 | European |
| VK158.SG | Russia | 950 | European |
| VK159.SG | Russia | 950 | European |
| VK15.SG | Russia | 900 | European |
| VK160.SG | Russia | 850 | European |
| VK161.SG | Russia | 850 | European |
| VK163.SG | United Kingdom | 1010 | European |
| VK164.SG | United Kingdom | 1010 | European |
| VK165.SG | United Kingdom | 1010 | European |
| VK166.SG | United Kingdom | 1010 | European |
| VK167.SG | United Kingdom | 1010 | European |
| VK168.SG | United Kingdom | 1010 | European |
| VK16.SG | Russia | 900 | European |
| VK171.SG | United Kingdom | 945 | European |
| VK172.SG | United Kingdom | 1010 | European |
| VK173.SG | United Kingdom | 1010 | European |
| VK174.SG | United Kingdom | 1010 | European |
| VK175.SG | United Kingdom | 1010 | European |
| VK176.SG | United Kingdom | 1010 | European |
| VK177.SG | United Kingdom | 1010 | European |
| VK178.SG | United Kingdom | 1010 | European |
| VK179.SG | Greenland | 900 | European |
| VK17.SG | Russia | 900 | European |
| VK180.SG | Greenland | 900 | European |
| VK182.SG | Greenland | 900 | European |
| VK183.SG | Greenland | 900 | European |

| | | | |
|---|---|---|---|
| VK184.SG | Greenland | 900 | European |
| VK185.SG | Greenland | 900 | European |
| VK186.SG | Greenland | 900 | European |
| VK187.SG | Greenland | 998 | European |
| VK189.SG | Greenland | 726 | European |
| VK18.SG | Russia | 900 | European |
| VK190.SG | Greenland | 834 | European |
| VK191.SG | Greenland | 762 | European |
| VK193.SG | Greenland | 900 | European |
| VK196.SG | Greenland | 1103 | European |
| VK19.SG | Russia | 900 | European |
| VK1.SG | Greenland | 950 | European |
| VK200.SG | Poland | 1059 | European |
| VK201.SG | United Kingdom | 1450 | European |
| VK202.SG | United Kingdom | 1000 | European |
| VK203.SG | United Kingdom | 1589 | European |
| VK204.SG | United Kingdom | 1000 | European |
| VK205.SG | United Kingdom | 1020 | European |
| VK206.SG | United Kingdom | 1000 | European |
| VK207.SG | United Kingdom | 930 | European |
| VK208.SG | United Kingdom | 1112 | European |
| VK20.SG | Russia | 900 | European |
| VK210.SG | Poland | 800 | European |
| VK211.SG | Poland | 800 | European |
| VK212.SG | Poland | 908 | European |
| VK213.SG | Denmark | 1561 | European |
| VK214.SG | Denmark | 3838 | European |
| VK215.SG | Denmark | 1100 | European |
| VK216.SG | Denmark | 1100 | European |

| | | | |
|---|---|---|---|
| VK217.SG | Sweden | 950 | European |
| VK218.SG | Russia | 900 | European |
| VK219.SG | Russia | 950 | European |
| VK21.SG | Russia | 900 | European |
| VK220.SG | Russia | 900 | European |
| VK221.SG | Russia | 1050 | European |
| VK222.SG | Russia | 950 | European |
| VK223.SG | Russia | 950 | European |
| VK224.SG | Russia | 950 | European |
| VK225.SG | Iceland | 850 | European |
| VK226.SG | Iceland | 850 | European |
| VK227.SG | Iceland | 850 | European |
| VK228.SG | Iceland | 850 | European |
| VK22.SG | Russia | 900 | European |
| VK230.SG | Iceland | 850 | European |
| VK232.SG | Sweden | 975 | European |
| VK234.SG | Faroes | 350 | European |
| VK236.SG | Faroes | 350 | European |
| VK237.SG | Faroes | 350 | European |
| VK238.SG | Faroes | 350 | European |
| VK239.SG | Faroes | 350 | European |
| VK23.SG | Russia | 900 | European |
| VK240.SG | Faroes | 350 | European |
| VK241.SG | Faroes | 350 | European |
| VK242.SG | Faroes | 350 | European |
| VK244.SG | Faroes | 350 | European |
| VK245.SG | Faroes | 350 | European |
| VK247.SG | Denmark | 1075 | European |
| VK248.SG | Faroes | 350 | European |

| | | | |
|---|---|---|---|
| VK24.SG | Faroes | 900 | European |
| VK251.SG | Sweden | 975 | European |
| VK252.SG | Russia | 950 | European |
| VK253.SG | Russia | 950 | European |
| VK254.SG | Russia | 950 | European |
| VK255.SG | Russia | 950 | European |
| VK256.SG | United Kingdom | 953 | European |
| VK257.SG | United Kingdom | 953 | European |
| VK258.SG | United Kingdom | 953 | European |
| VK259.SG | United Kingdom | 953 | European |
| VK25.SG | Faroes | 350 | European |
| VK260.SG | United Kingdom | 953 | European |
| VK261.SG | United Kingdom | 953 | European |
| VK262.SG | United Kingdom | 953 | European |
| VK263.SG | United Kingdom | 953 | European |
| VK264.SG | United Kingdom | 953 | European |
| VK265.SG | Sweden | 1000 | European |
| VK266.SG | Sweden | 1000 | European |
| VK267.SG | Sweden | 1000 | European |
| VK268.SG | Sweden | 1000 | European |
| VK269.SG | Sweden | 1000 | European |
| VK270.SG | Sweden | 1000 | European |
| VK272.SG | Russia | 950 | European |
| VK273.SG | Russia | 950 | European |
| VK274.SG | Denmark | 1000 | European |
| VK275.SG | Denmark | 1000 | European |
| VK276.SG | Denmark | 1000 | European |
| VK278.SG | Denmark | 1000 | European |
| VK279.SG | Denmark | 1000 | European |

| | | | |
|---|---|---|---|
| VK27.SG | Faroes | 350 | European |
| VK280.SG | Denmark | 1000 | European |
| VK281.SG | Denmark | 1000 | European |
| VK282.SG | Denmark | 1000 | European |
| VK284.SG | Denmark | 1000 | European |
| VK285.SG | Denmark | 1000 | European |
| VK286.SG | Denmark | 1000 | European |
| VK287.SG | Denmark | 1000 | European |
| VK288.SG | Denmark | 1000 | European |
| VK289.SG | Denmark | 1100 | European |
| VK290.SG | Denmark | 1000 | European |
| VK291.SG | Denmark | 1100 | European |
| VK292.SG | Denmark | 1000 | European |
| VK294.SG | Denmark | 1075 | European |
| VK295.SG | Denmark | 1000 | European |
| VK296.SG | Denmark | 1230 | European |
| VK297.SG | Denmark | 1200 | European |
| VK298.SG | Denmark | 1050 | European |
| VK29.SG | Sweden | 900 | European |
| VK300.SG | Denmark | 1075 | European |
| VK301.SG | Denmark | 1185 | European |
| VK303.SG | Sweden | 900 | European |
| VK304.SG | Sweden | 900 | European |
| VK306.SG | Sweden | 900 | European |
| VK308.SG | Sweden | 900 | European |
| VK309.SG | Sweden | 900 | European |
| VK30.SG | Sweden | 900 | European |
| VK312.SG | Denmark | 1075 | European |
| VK313.SG | Denmark | 1075 | European |

| | | | |
|---|---|---|---|
| VK314.SG | Denmark | 1075 | European |
| VK315.SG | Denmark | 1075 | European |
| VK316.SG | Denmark | 1000 | European |
| VK317.SG | Denmark | 1000 | European |
| VK318.SG | Denmark | 1025 | European |
| VK319.SG | Denmark | 1150 | European |
| VK31.SG | Sweden | 900 | European |
| VK320.SG | Denmark | 1000 | European |
| VK322.SG | Denmark | 934 | European |
| VK323.SG | Denmark | 1000 | European |
| VK324.SG | Denmark | 935 | European |
| VK325.SG | Denmark | 1145 | European |
| VK326.SG | Denmark | 1026 | European |
| VK327.SG | Denmark | 979 | European |
| VK328.SG | Denmark | 1104 | European |
| VK329.SG | Denmark | 1183 | European |
| VK330.SG | Denmark | 896 | European |
| VK332.SG | Sweden | 1085 | European |
| VK333.SG | Sweden | 1052 | European |
| VK334.SG | Sweden | 893 | European |
| VK335.SG | Sweden | 1000 | European |
| VK336.SG | Sweden | 1091 | European |
| VK337.SG | Sweden | 1069 | European |
| VK338.SG | Denmark | 1000 | European |
| VK339.SG | Denmark | 1075 | European |
| VK33.SG | Sweden | 900 | European |
| VK340.SG | Denmark | 1075 | European |
| VK342.SG | Sweden | 1000 | European |
| VK343.SG | Sweden | 1000 | European |

| | | | |
|---|---|---|---|
| VK344.SG | Sweden | 1000 | European |
| VK345.SG | Sweden | 1000 | European |
| VK346.SG | Sweden | 1000 | European |
| VK347.SG | Sweden | 1000 | European |
| VK348.SG | Sweden | 1000 | European |
| VK349.SG | Sweden | 1110 | European |
| VK34.SG | Sweden | 900 | European |
| VK350.SG | Sweden | 1134 | European |
| VK352.SG | Sweden | 1000 | European |
| VK353.SG | Sweden | 893 | European |
| VK354.SG | Sweden | 939 | European |
| VK355.SG | Sweden | 1097 | European |
| VK357.SG | Sweden | 887 | European |
| VK358.SG | Sweden | 1092 | European |
| VK359.SG | Sweden | 1200 | European |
| VK35.SG | Sweden | 900 | European |
| VK361.SG | Denmark | 1000 | European |
| VK362.SG | Denmark | 1000 | European |
| VK363.SG | Denmark | 1000 | European |
| VK364.SG | Denmark | 1000 | European |
| VK365.SG | Denmark | 1000 | European |
| VK366.SG | Denmark | 1000 | European |
| VK367.SG | Denmark | 1000 | European |
| VK368.SG | Denmark | 1000 | European |
| VK369.SG | Denmark | 1075 | European |
| VK370.SG | Denmark | 1000 | European |
| VK371.SG | Denmark | 1065 | European |
| VK372.SG | Denmark | 1000 | European |
| VK373.SG | Denmark | 1000 | European |

| | | | |
|---|---|---|---|
| VK379.SG | Sweden | 1200 | European |
| VK380.SG | Sweden | 1000 | European |
| VK382.SG | Sweden | 1200 | European |
| VK383.SG | Denmark | 1075 | European |
| VK384.SG | Denmark | 1075 | European |
| VK385.SG | Denmark | 1075 | European |
| VK386.SG | Norway | 1000 | European |
| VK387.SG | Norway | 950 | European |
| VK388.SG | Norway | 800 | European |
| VK389.SG | Norway | 1000 | European |
| VK390.SG | Norway | 1450 | European |
| VK391.SG | Norway | 1450 | European |
| VK392.SG | Norway | 1100 | European |
| VK393.SG | Norway | 1000 | European |
| VK394.SG | Norway | 1000 | European |
| VK395.SG | Sweden | 900 | European |
| VK396.SG | Sweden | 900 | European |
| VK397.SG | Sweden | 900 | European |
| VK398.SG | Sweden | 900 | European |
| VK399.SG | Sweden | 900 | European |
| VK39.SG | Sweden | 900 | European |
| VK400.SG | Sweden | 900 | European |
| VK401.SG | Sweden | 900 | European |
| VK402.SG | Sweden | 900 | European |
| VK403.SG | Sweden | 900 | European |
| VK404.SG | Sweden | 900 | European |
| VK405.SG | Sweden | 900 | European |
| VK406.SG | Sweden | 900 | European |
| VK407.SG | Sweden | 900 | European |

| | | | |
|---|---|---|---|
| VK408.SG | Russia | 900 | European |
| VK409.SG | Russia | 900 | European |
| VK40.SG | Sweden | 900 | European |
| VK410.SG | Russia | 850 | European |
| VK411.SG | Denmark | 1000 | European |
| VK413.SG | Russia | 950 | European |
| VK414.SG | Norway | 950 | European |
| VK415.SG | Norway | 1050 | European |
| VK417.SG | Norway | 1100 | European |
| VK418.SG | Norway | 1600 | European |
| VK419.SG | Norway | 1200 | European |
| VK420.SG | Norway | 1050 | European |
| VK421.SG | Norway | 950 | European |
| VK422.SG | Norway | 1000 | European |
| VK424.SG | Sweden | 900 | European |
| VK425.SG | Sweden | 900 | European |
| VK426.SG | Sweden | 900 | European |
| VK427.SG | Sweden | 900 | European |
| VK428.SG | Sweden | 975 | European |
| VK429.SG | Sweden | 975 | European |
| VK42.SG | Sweden | 900 | European |
| VK430.SG | Sweden | 975 | European |
| VK431.SG | Sweden | 975 | European |
| VK432.SG | Sweden | 975 | European |
| VK433.SG | Sweden | 975 | European |
| VK434.SG | Sweden | 975 | European |
| VK435.SG | Sweden | 975 | European |
| VK437.SG | Sweden | 975 | European |
| VK438.SG | Sweden | 975 | European |

| | | | |
|---|---|---|---|
| VK439.SG | Sweden | 975 | European |
| VK440.SG | Sweden | 975 | European |
| VK441.SG | Sweden | 975 | European |
| VK442.SG | Sweden | 1097 | European |
| VK443.SG | Sweden | 1000 | European |
| VK444.SG | Sweden | 1097 | European |
| VK445.SG | Denmark | 1000 | European |
| VK446.SG | Denmark | 1000 | European |
| VK448.SG | Norway | 1050 | European |
| VK449.SG | United Kingdom | 953 | European |
| VK44.SG | Faroes | 350 | European |
| VK450.SG | Sweden | 975 | European |
| VK452.SG | Sweden | 975 | European |
| VK453.SG | Sweden | 975 | European |
| VK454.SG | Sweden | 975 | European |
| VK455.SG | Sweden | 975 | European |
| VK456.SG | Sweden | 975 | European |
| VK457.SG | Sweden | 975 | European |
| VK458.SG | Sweden | 975 | European |
| VK459.SG | Sweden | 975 | European |
| VK45.SG | Faroes | 350 | European |
| VK460.SG | Sweden | 975 | European |
| VK461.SG | Sweden | 975 | European |
| VK462.SG | Sweden | 975 | European |
| VK463.SG | Sweden | 975 | European |
| VK464.SG | Sweden | 975 | European |
| VK466.SG | Russia | 950 | European |
| VK467.SG | Sweden | 975 | European |
| VK468.SG | Sweden | 975 | European |

| | | | |
|---|---|---|---|
| VK469.SG | Sweden | 975 | European |
| VK46.SG | Faroes | 350 | European |
| VK470.SG | Russia | 950 | European |
| VK471.SG | Sweden | 975 | European |
| VK472.SG | Sweden | 975 | European |
| VK473.SG | Sweden | 975 | European |
| VK474.SG | Sweden | 975 | European |
| VK475.SG | Sweden | 975 | European |
| VK476.SG | Sweden | 975 | European |
| VK477.SG | Sweden | 975 | European |
| VK478.SG | Sweden | 975 | European |
| VK479.SG | Sweden | 975 | European |
| VK480.SG | Estonia | 1200 | European |
| VK481.SG | Estonia | 1200 | European |
| VK482.SG | Estonia | 1200 | European |
| VK483.SG | Estonia | 1191 | European |
| VK484.SG | Estonia | 1200 | European |
| VK485.SG | Estonia | 1244 | European |
| VK486.SG | Estonia | 1200 | European |
| VK487.SG | Estonia | 1200 | European |
| VK488.SG | Estonia | 1200 | European |
| VK489.SG | Estonia | 1200 | European |
| VK48.SG | Sweden | 975 | European |
| VK490.SG | Estonia | 1227 | European |
| VK491.SG | Estonia | 1200 | European |
| VK492.SG | Estonia | 1200 | European |
| VK493.SG | Estonia | 1200 | European |
| VK494.SG | Poland | 950 | European |
| VK495.SG | Estonia | 1200 | European |

| | | | |
|---|---|---|---|
| VK496.SG | Estonia | 1200 | European |
| VK497.SG | Estonia | 1200 | European |
| VK498.SG | Estonia | 1200 | European |
| VK504.SG | Estonia | 1200 | European |
| VK505.SG | Estonia | 1200 | European |
| VK506.SG | Estonia | 1237 | European |
| VK507.SG | Estonia | 1121 | European |
| VK508.SG | Estonia | 1200 | European |
| VK509.SG | Estonia | 1200 | European |
| VK50.SG | Sweden | 975 | European |
| VK510.SG | Estonia | 1200 | European |
| VK511.SG | Estonia | 1200 | European |
| VK512.SG | Estonia | 1200 | European |
| VK513.SG | Greenland | 900 | European |
| VK514.SG | Norway | 1200 | European |
| VK515.SG | Norway | 1000 | European |
| VK516.SG | Norway | 1000 | European |
| VK517.SG | Sweden | 900 | European |
| VK518.SG | Norway | 1100 | European |
| VK519.SG | Norway | 1200 | European |
| VK51.SG | Sweden | 975 | European |
| VK520.SG | Norway | 1000 | European |
| VK521.SG | Denmark | 1650 | European |
| VK522.SG | Sweden | 1550 | European |
| VK523.SG | Norway | 1450 | European |
| VK524.SG | Norway | 1000 | European |
| VK525.SG | Norway | 1100 | European |
| VK526.SG | Norway | 1100 | European |
| VK527.SG | Sweden | 900 | European |

| | | | |
|---|---|---|---|
| VK528.SG | Norway | 1150 | European |
| VK529.SG | Norway | 1150 | European |
| VK530.SG | Norway | 1000 | European |
| VK531.SG | Norway | 4347 | European |
| VK532.SG | Denmark | 1850 | European |
| VK533.SG | Sweden | 1000 | European |
| VK534.SG | Italy | 766 | European |
| VK535.SG | Italy | 700 | European |
| VK536.SG | Italy | 800 | European |
| VK537.SG | Italy | 800 | European |
| VK538.SG | Italy | 800 | European |
| VK539.SG | Ukraine | 900 | European |
| VK53.SG | Sweden | 975 | European |
| VK540.SG | Ukraine | 900 | European |
| VK541.SG | Ukraine | 700 | European |
| VK542.SG | Ukraine | 900 | European |
| VK547.SG | Norway | 1050 | European |
| VK548.SG | Norway | 1100 | European |
| VK549.SG | Estonia | 1200 | European |
| VK550.SG | Estonia | 1200 | European |
| VK551.SG | Estonia | 1200 | European |
| VK552.SG | Estonia | 1200 | European |
| VK553.SG | Estonia | 1200 | European |
| VK554.SG | Estonia | 1120 | European |
| VK555.SG | Estonia | 1200 | European |
| VK56.SG | Sweden | 975 | European |
| VK579.SG | Sweden | 1650 | European |
| VK57.SG | Sweden | 975 | European |
| VK582.SG | Denmark | 1900 | European |

| | | | |
|---|---|---|---|
| VK58.SG | Sweden | 975 | European |
| VK60.SG | Sweden | 975 | European |
| VK63.SG | Sweden | 975 | European |
| VK64.SG | Sweden | 975 | European |
| VK65.SG | Denmark | 1100 | European |
| VK69.SG | Denmark | 1100 | European |
| VK6.SG | Greenland | 950 | European |
| VK70.SG | Denmark | 1100 | European |
| VK71.SG | Denmark | 1100 | European |
| VK74.SG | Greenland | 776 | European |
| VK75.SG | Greenland | 931 | European |
| VK76.SG | Greenland | 878 | European |
| VK77.SG | Greenland | 845 | European |
| VK78.SG | Greenland | 843 | European |
| VK84.SG | Denmark | 1075 | European |
| VK86.SG | Denmark | 1075 | European |
| VK87.SG | Denmark | 1075 | European |
| VK90.SG | Denmark | 1075 | European |
| VK92.SG | Denmark | 1075 | European |
| VK94.SG | Denmark | 1000 | European |
| VK95.SG | Iceland | 850 | European |
| VK98.SG | Iceland | 850 | European |
| VK99.SG | Iceland | 850 | European |
| VK9.SG | Greenland | 950 | European |
| I1524 | Russia | 905 | European |
| I20760 | Russia | 6255 | European |
| I7341 | Russia | 1100 | European |
| I7757 | Russia | 875 | European |
| I7332 | Russia | 1070 | European |

| | | | |
|---|---|---|---|
| I7346 | Russia | 1145 | European |
| I7780 | Russia | 4615 | European |
| bla001.SG | Russia | 1299 | European |
| brn001.SG | Russia | 7334 | European |
| brn002.SG | Russia | 6061 | European |
| brn003.SG | Russia | 6550 | European |
| brn008.SG | Russia | 7379 | European |
| brn012.SG | Russia | 6115 | European |
| cta016.SG | Russia | 8264 | European |
| irk007.SG | Russia | 8801 | European |
| irk008.SG | Russia | 3787 | European |
| irk00x.SG | Russia | 8419 | European |
| irk017.SG | Russia | 5132 | European |
| irk022.SG | Russia | 4251 | European |
| irk025.SG | Russia | 4350 | European |
| irk030.SG | Russia | 5969 | European |
| irk032.SG | Russia | 613 | European |
| irk033.SG | Russia | 4858 | European |
| irk034.SG | Russia | 5525 | European |
| irk036.SG | Russia | 4720 | European |
| irk040.SG | Russia | 5559 | European |
| irk050.SG | Russia | 4350 | European |
| irk051.SG | Russia | 7922 | European |
| irk057.SG | Russia | 4380 | European |
| irk061.SG | Russia | 4341 | European |
| irk068.SG | Russia | 4468 | European |
| irk071.SG | Russia | 5132 | European |
| irk075.SG | Russia | 5397 | European |
| irk076.SG | Russia | 4104 | European |

| | | | |
|---|---|---|---|
| irk078.SG | Russia | 3107 | European |
| kra001.SG | Russia | 4170 | European |
| mak026.SG | Russia | 4759 | European |
| N2a.SG | Russia | 6762 | European |
| N4a1.SG | Russia | 4505 | European |
| N4b2.SG | Russia | 4200 | European |
| N5a.SG | Russia | 6232 | European |
| yak021.SG | Russia | 3225 | European |
| yak022.SG | Russia | 3799 | European |
| yak023.SG | Russia | 3735 | European |
| yak024.SG | Russia | 3500 | European |
| yak025.SG | Russia | 16736 | European |
| yak030-b1e1l2p1.SG | Russia | 2606 | European |
| WEZ15.SG | Germany | 3200 | European |
| WEZ16.SG | Germany | 5050 | European |
| WEZ24.SG | Germany | 3200 | European |
| WEZ35.SG | Germany | 3200 | European |
| WEZ39.SG | Germany | 3200 | European |
| WEZ40.SG | Germany | 3200 | European |
| WEZ48.SG | Germany | 3200 | European |
| WEZ51.SG | Germany | 3200 | European |
| WEZ53.SG | Germany | 3200 | European |
| WEZ54.SG | Germany | 3200 | European |
| WEZ56.SG | Germany | 3200 | European |
| WEZ57.SG | Germany | 3200 | European |
| WEZ58.SG | Germany | 3200 | European |
| WEZ59.SG | Germany | 3200 | European |
| WEZ61.SG | Germany | 3200 | European |
| WEZ64.SG | Germany | 3200 | European |

| | | | |
|---|---|---|---|
| WEZ71.SG | Germany | 3200 | European |
| WEZ74.SG | Germany | 3200 | European |
| WEZ83.SG | Germany | 3200 | European |
| BLS001 | Czech Republic | 4675 | European |
| BLS002 | Czech Republic | 4692 | European |
| BNL001 | Czech Republic | 4020 | European |
| BNL002 | Czech Republic | 3995 | European |
| BNL003 | Czech Republic | 3995 | European |
| BNL004 | Czech Republic | 3995 | European |
| BNL005 | Czech Republic | 4084 | European |
| BNL006 | Czech Republic | 4070 | European |
| BNL007 | Czech Republic | 4070 | European |
| BNL008 | Czech Republic | 3926 | European |
| BNL009 | Czech Republic | 3975 | European |
| BNL010 | Czech Republic | 3772 | European |
| BRZ001 | Czech Republic | 5820 | European |
| BRZ002 | Czech Republic | 5521 | European |
| BUT002 | Czech Republic | 4350 | European |
| BUT003 | Czech Republic | 4335 | European |
| CAH005 | Czech Republic | 4349 | European |
| CAH010 | Czech Republic | 4275 | European |
| CHL001 | Czech Republic | 3905 | European |
| CHL002 | Czech Republic | 4000 | European |
| CHL003 | Czech Republic | 3937 | European |
| CHL005 | Czech Republic | 4000 | European |
| CHL006 | Czech Republic | 3897 | European |
| CHL007 | Czech Republic | 4000 | European |
| CHL008 | Czech Republic | 3960 | European |
| DRO001 | Czech Republic | 4697 | European |

| | | | |
|---|---|---|---|
| HOL001 | Czech Republic | 5613 | European |
| HOL002 | Czech Republic | 5140 | European |
| HOL004 | Czech Republic | 5154 | European |
| HOP001 | Czech Republic | 4240 | European |
| HOP002 | Czech Republic | 4177 | European |
| HOP003 | Czech Republic | 4250 | European |
| HOP004 | Czech Republic | 4222 | European |
| KNE001 | Czech Republic | 4017 | European |
| KNE002 | Czech Republic | 4043 | European |
| KNE003 | Czech Republic | 3971 | European |
| KNE004 | Czech Republic | 4042 | European |
| KNE005 | Czech Republic | 4072 | European |
| KNE006 | Czech Republic | 4065 | European |
| KNE007 | Czech Republic | 3977 | European |
| KO1001 | Czech Republic | 4761 | European |
| KO1002 | Czech Republic | 4533 | European |
| KO1003 | Czech Republic | 4160 | European |
| KO1004 | Czech Republic | 4166 | European |
| KO1005 | Czech Republic | 4331 | European |
| KO1006 | Czech Republic | 3868 | European |
| KO1007 | Czech Republic | 3737 | European |
| KO1008 | Czech Republic | 3805 | European |
| KO1009 | Czech Republic | 3894 | European |
| KO1010 | Czech Republic | 3906 | European |
| KO1011 | Czech Republic | 3993 | European |
| KO1013 | Czech Republic | 3914 | European |
| KO1014 | Czech Republic | 3915 | European |
| KO1015 | Czech Republic | 3774 | European |
| KO1016 | Czech Republic | 4750 | European |

| | | | |
|---|---|---|---|
| KO7001 | Czech Republic | 3985 | European |
| KO7002 | Czech Republic | 4000 | European |
| KO7003 | Czech Republic | 4124 | European |
| KOB001 | Czech Republic | 5650 | European |
| KOB002 | Czech Republic | 5630 | European |
| KOB003 | Czech Republic | 5600 | European |
| KOB004 | Czech Republic | 5600 | European |
| KOB005 | Czech Republic | 5704 | European |
| KOB007 | Czech Republic | 5632 | European |
| KON001 | Czech Republic | 4697 | European |
| KON003 | Czech Republic | 4650 | European |
| KON004 | Czech Republic | 4398 | European |
| KON005 | Czech Republic | 4680 | European |
| KOP001 | Czech Republic | 4250 | European |
| KOP002 | Czech Republic | 4250 | European |
| KOP003 | Czech Republic | 4342 | European |
| MIB001 | Czech Republic | 3779 | European |
| MIB002 | Czech Republic | 3765 | European |
| MIB003 | Czech Republic | 3810 | European |
| MIB004 | Czech Republic | 3769 | European |
| MIB005 | Czech Republic | 3767 | European |
| MIB006 | Czech Republic | 3808 | European |
| MIB007 | Czech Republic | 3817 | European |
| MIB008 | Czech Republic | 3911 | European |
| MIB009 | Czech Republic | 3773 | European |
| MIB010 | Czech Republic | 3846 | European |
| MIB011 | Czech Republic | 3761 | European |
| MIB012 | Czech Republic | 3774 | European |
| MIB013 | Czech Republic | 3762 | European |

| | | | |
|---|---|---|---|
| MIB014 | Czech Republic | 3789 | European |
| MIB015 | Czech Republic | 3770 | European |
| MIB016 | Czech Republic | 3738 | European |
| MIB017 | Czech Republic | 3806 | European |
| MIB018 | Czech Republic | 3775 | European |
| MIB019 | Czech Republic | 3812 | European |
| MIB020 | Czech Republic | 3848 | European |
| MIB021 | Czech Republic | 3839 | European |
| MIB022 | Czech Republic | 3866 | European |
| MIB024 | Czech Republic | 3875 | European |
| MIB026 | Czech Republic | 3793 | European |
| MIB027 | Czech Republic | 3941 | European |
| MIB028 | Czech Republic | 3855 | European |
| MIB029 | Czech Republic | 3785 | European |
| MIB030 | Czech Republic | 3932 | European |
| MIB031 | Czech Republic | 3791 | European |
| MIB032 | Czech Republic | 3791 | European |
| MIB033 | Czech Republic | 3776 | European |
| MIB034 | Czech Republic | 3659 | European |
| MIB035 | Czech Republic | 3795 | European |
| MIB036 | Czech Republic | 3837 | European |
| MIB037 | Czech Republic | 3793 | European |
| MIB038 | Czech Republic | 3693 | European |
| MIB039 | Czech Republic | 3768 | European |
| MIB040 | Czech Republic | 3829 | European |
| MIB041 | Czech Republic | 3773 | European |
| MIB042 | Czech Republic | 3776 | European |
| MIB043 | Czech Republic | 3807 | European |
| MIB044 | Czech Republic | 3840 | European |

| | | | |
|---|---|---|---|
| MIB045 | Czech Republic | 3894 | European |
| MIB046 | Czech Republic | 3872 | European |
| MIB047 | Czech Republic | 3879 | European |
| MIB048 | Czech Republic | 3825 | European |
| MIB049 | Czech Republic | 3758 | European |
| MIB051 | Czech Republic | 3792 | European |
| MIB052 | Czech Republic | 3777 | European |
| MIB053 | Czech Republic | 3761 | European |
| MIB054 | Czech Republic | 3835 | European |
| MIB055 | Czech Republic | 3773 | European |
| MIB056 | Czech Republic | 3761 | European |
| MIB057 | Czech Republic | 3843 | European |
| MIB058 | Czech Republic | 3763 | European |
| MIB059 | Czech Republic | 3781 | European |
| MIB060 | Czech Republic | 3865 | European |
| MIB061 | Czech Republic | 3765 | European |
| MIB062 | Czech Republic | 3864 | European |
| MIB063 | Czech Republic | 3813 | European |
| MIB064 | Czech Republic | 3853 | European |
| MIB065 | Czech Republic | 3744 | European |
| MIB066 | Czech Republic | 3801 | European |
| MIB067 | Czech Republic | 3782 | European |
| MIB068 | Czech Republic | 3757 | European |
| MIB069 | Czech Republic | 3931 | European |
| MIB070 | Czech Republic | 3825 | European |
| MIB071 | Czech Republic | 3790 | European |
| MIB072 | Czech Republic | 3918 | European |
| MIB073 | Czech Republic | 3921 | European |
| MIB074 | Czech Republic | 4014 | European |

| | | | |
|---|---|---|---|
| MIB075 | Czech Republic | 3866 | European |
| MIB076 | Czech Republic | 3887 | European |
| MIB077 | Czech Republic | 3961 | European |
| MIB078 | Czech Republic | 3993 | European |
| MIB079 | Czech Republic | 3871 | European |
| MIB082 | Czech Republic | 3770 | European |
| MIB083 | Czech Republic | 4176 | European |
| MIB084 | Czech Republic | 4031 | European |
| MIB086 | Czech Republic | 3906 | European |
| MIB087 | Czech Republic | 3749 | European |
| MIG008 | Czech Republic | 3872 | European |
| MIG009 | Czech Republic | 3932 | European |
| MIG010 | Czech Republic | 3992 | European |
| MIG011 | Czech Republic | 3990 | European |
| MIG012 | Czech Republic | 3800 | European |
| MIS001 | Czech Republic | 3908 | European |
| MIS002 | Czech Republic | 3883 | European |
| MIS004 | Czech Republic | 3833 | European |
| MIS005 | Czech Republic | 4054 | European |
| MIS006 | Czech Republic | 4235 | European |
| MIS007 | Czech Republic | 3927 | European |
| NER001 | Czech Republic | 6076 | European |
| OBR001 | Czech Republic | 4545 | European |
| OBR002 | Czech Republic | 4382 | European |
| OBR003 | Czech Republic | 4835 | European |
| OBR004 | Czech Republic | 4512 | European |
| OHR001 | Czech Republic | 4467 | European |
| OHR002 | Czech Republic | 4477 | European |
| PDA001 | Czech Republic | 4017 | European |

| | | | |
|---|---|---|---|
| PDA002 | Czech Republic | 4057 | European |
| PDA003 | Czech Republic | 4131 | European |
| PDA004 | Czech Republic | 4053 | European |
| PDA005 | Czech Republic | 4164 | European |
| PMI001 | Czech Republic | 4096 | European |
| PMI002 | Czech Republic | 4171 | European |
| PMI003 | Czech Republic | 4005 | European |
| PMI004 | Czech Republic | 4013 | European |
| PMI006 | Czech Republic | 4104 | European |
| PMI009 | Czech Republic | 3849 | European |
| PMI010 | Czech Republic | 3902 | European |
| PMI012 | Czech Republic | 3657 | European |
| PNL001 | Czech Republic | 4843 | European |
| PNL002 | Czech Republic | 4653 | European |
| PRE001 | Czech Republic | 4625 | European |
| PRU001 | Czech Republic | 4331 | European |
| PRU002 | Czech Republic | 4470 | European |
| PRU003 | Czech Republic | 4427 | European |
| PRU004 | Czech Republic | 4264 | European |
| PRU005 | Czech Republic | 4350 | European |
| RDV001 | Czech Republic | 4592 | European |
| ROU001 | Czech Republic | 4000 | European |
| ROU002 | Czech Republic | 3997 | European |
| ROU003 | Czech Republic | 3995 | European |
| ROU004 | Czech Republic | 3995 | European |
| ROU005 | Czech Republic | 4000 | European |
| ROU006 | Czech Republic | 4000 | European |
| ROU007 | Czech Republic | 4000 | European |
| STD001 | Czech Republic | 4500 | European |

| | | | |
|---|---|---|---|
| STD002 | Czech Republic | 4720 | European |
| STD003 | Czech Republic | 4877 | European |
| TIS001 | Czech Republic | 4326 | European |
| TIS002 | Czech Republic | 4470 | European |
| TOU001 | Czech Republic | 4712 | European |
| TRM001 | Czech Republic | 4650 | European |
| TRM003 | Czech Republic | 4620 | European |
| TRM006 | Czech Republic | 4644 | European |
| TUC001 | Czech Republic | 4940 | European |
| TUC002 | Czech Republic | 4932 | European |
| TUC003 | Czech Republic | 4940 | European |
| TUC004 | Czech Republic | 4940 | European |
| TUC005 | Czech Republic | 4940 | European |
| TUC006 | Czech Republic | 4879 | European |
| TUC007 | Czech Republic | 6080 | European |
| VLI004 | Czech Republic | 6141 | European |
| VLI006 | Czech Republic | 5661 | European |
| VLI007 | Czech Republic | 4739 | European |
| VLI008 | Czech Republic | 4759 | European |
| VLI009 | Czech Republic | 4585 | European |
| VLI010 | Czech Republic | 4483 | European |
| VLI011 | Czech Republic | 4718 | European |
| VLI015 | Czech Republic | 4725 | European |
| VLI016 | Czech Republic | 4345 | European |
| VLI017 | Czech Republic | 4336 | European |
| VLI019 | Czech Republic | 4500 | European |
| VLI020 | Czech Republic | 4446 | European |
| VLI022 | Czech Republic | 4265 | European |
| VLI024 | Czech Republic | 4150 | European |

| | | | |
|---|---|---|---|
| VLI025 | Czech Republic | 4150 | European |
| VLI026 | Czech Republic | 4150 | European |
| VLI028 | Czech Republic | 4150 | European |
| VLI029 | Czech Republic | 4150 | European |
| VLI030 | Czech Republic | 4150 | European |
| VLI031 | Czech Republic | 4510 | European |
| VLI032 | Czech Republic | 4682 | European |
| VLI033 | Czech Republic | 4682 | European |
| VLI039 | Czech Republic | 4000 | European |
| VLI042 | Czech Republic | 3774 | European |
| VLI045 | Czech Republic | 3943 | European |
| VLI046 | Czech Republic | 3809 | European |
| VLI047 | Czech Republic | 3800 | European |
| VLI050 | Czech Republic | 3796 | European |
| VLI051 | Czech Republic | 3721 | European |
| VLI053 | Czech Republic | 3550 | European |
| VLI058 | Czech Republic | 4066 | European |
| VLI060 | Czech Republic | 3930 | European |
| VLI061 | Czech Republic | 4000 | European |
| VLI064 | Czech Republic | 4018 | European |
| VLI065 | Czech Republic | 3936 | European |
| VLI067 | Czech Republic | 4718 | European |
| VLI070 | Czech Republic | 4351 | European |
| VLI071 | Czech Republic | 4715 | European |
| VLI075 | Czech Republic | 4650 | European |
| VLI076 | Czech Republic | 4906 | European |
| VLI079 | Czech Republic | 4623 | European |
| VLI081 | Czech Republic | 4650 | European |
| VLI085 | Czech Republic | 4653 | European |

| | | | |
|---|---|---|---|
| VLI088 | Czech Republic | 4736 | European |
| VLI090 | Czech Republic | 4768 | European |
| VLI092 | Czech Republic | 4719 | European |
| VPR001 | Czech Republic | 4756 | European |
| ZEL001 | Czech Republic | 4350 | European |
| TRPTN_611.SG | United Kingdom | 5418 | European |
| TRPTN_613.SG | United Kingdom | 5639 | European |
| Kou01.SG | Greece | 4250 | European |
| Kou03.SG | Greece | 4250 | European |
| Log02.SG | Greece | 4250 | European |
| Log04.SG | Greece | 4250 | European |
| Mik15.SG | Greece | 4250 | European |
| Pta08.SG | Greece | 4250 | European |
| FNO001 | Russia | 7881 | European |
| FNO003 | Russia | 7800 | European |
| FNO006 | Russia | 7800 | European |
| FNO007 | Russia | 7742 | European |
| IMA001 | Russia | 2000 | European |
| IMA002 | Russia | 2000 | European |
| IMA003 | Russia | 2062 | European |
| IMA004 | Russia | 2000 | European |
| IMA005 | Russia | 2051 | European |
| IMA006 | Russia | 1949 | European |
| IMA007 | Russia | 2000 | European |
| IMA008 | Russia | 2000 | European |
| PTO001 | Russia | 2932 | European |
| hem001.SG | Sweden | 4929 | European |
| hem004.SG | Sweden | 5145 | European |
| hem005.SG | Sweden | 5178 | European |

| | | | |
|---|---|---|---|
| vbj001.SG | Sweden | 4675 | European |
| vbj002.SG | Sweden | 4780 | European |
| vbj003.SG | Sweden | 4879 | European |
| vbj004.SG | Sweden | 4702 | European |
| vbj006.SG | Sweden | 4898 | European |
| vbj007.SG | Sweden | 4469 | European |
| vbj008.SG | Sweden | 4663 | European |
| vbj012.SG | Sweden | 4933 | European |
| vbj013.SG | Sweden | 4778 | European |
| vbj014.SG | Sweden | 4802 | European |
| vbj015.SG | Sweden | 4865 | European |
| vbj017.SG | Sweden | 4864 | European |
| vbj018.SG | Sweden | 4868 | European |
| ajv28.minus.SG | Sweden | 4829 | European |
| ajv36.minus.SG | Sweden | 4924 | European |
| hem001.minus.SG | Sweden | 4929 | European |
| hem004.minus.SG | Sweden | 5145 | European |
| hem005.minus.SG | Sweden | 5178 | European |
| vbj001.minus.SG | Sweden | 4675 | European |
| vbj002.minus.SG | Sweden | 4780 | European |
| vbj003.minus.SG | Sweden | 4879 | European |
| vbj004.minus.SG | Sweden | 4702 | European |
| vbj006.minus.SG | Sweden | 4898 | European |
| vbj007.minus.SG | Sweden | 4469 | European |
| vbj008.minus.SG | Sweden | 4663 | European |
| vbj012.minus.SG | Sweden | 4933 | European |
| vbj013.minus.SG | Sweden | 4778 | European |
| vbj014.minus.SG | Sweden | 4802 | European |
| vbj015.minus.SG | Sweden | 4865 | European |

| | | | |
|---|---|---|---|
| vbj017.minus.SG | Sweden | 4864 | European |
| vbj018.minus.SG | Sweden | 4868 | European |
| BER001.SG | Russia | 6283 | European |
| BOL001.SG | Russia | 4478 | European |
| BOL002.SG | Russia | 4306 | European |
| BOL003.SG | Russia | 4414 | European |
| GOL001.SG | Russia | 4441 | European |
| HAL001.SG | Russia | 4504 | European |
| HAN002.SG | Russia | 4605 | European |
| HAN003.SG | Russia | 4500 | European |
| HAN004.SG | Russia | 4511 | European |
| IVA001.SG | Russia | 4624 | European |
| KAR001.SG | Russia | 8321 | European |
| MIL001.SG | Russia | 4494 | European |
| MIL002.SG | Russia | 4127 | European |
| MOT001.SG | Russia | 4300 | European |
| NAU001.SG | Russia | 4526 | European |
| NAU002.SG | Russia | 4516 | European |
| NIK001.SG | Russia | 4500 | European |
| NIK002.SG | Russia | 4637 | European |
| NIK003.SG | Russia | 4430 | European |
| NIK004.SG | Russia | 4552 | European |
| NIK005.SG | Russia | 4686 | European |
| NIK006.SG | Russia | 4608 | European |
| NIK007.SG | Russia | 4555 | European |
| NIK008AB.SG | Russia | 4508 | European |
| PES001.SG | Russia | 12711 | European |
| RDT002.SG | Russia | 4364 | European |
| SOP002.SG | Estonia | 4620 | European |

| | | | |
|---|---|---|---|
| TIM001.SG | Russia | 4300 | European |
| TIM002.SG | Russia | 4300 | European |
| TIM003.SG | Russia | 4300 | European |
| TIM005.SG | Russia | 4300 | European |
| TIM006.SG | Russia | 4502 | European |
| TIM008.SG | Russia | 4502 | European |
| TIM009.SG | Russia | 4300 | European |
| TIM010.SG | Russia | 4300 | European |
| TIM011.SG | Russia | 4300 | European |
| VOD001.SG | Russia | 4383 | European |
| VORONKOVO001.SG | Russia | 4500 | European |
| VOR002.SG | Russia | 4500 | European |
| VORONKOVO003.SG | Russia | 4465 | European |
| VOR004.SG | Russia | 4698 | European |
| VOR005.SG | Russia | 4481 | European |
| RISE1283.SG | Denmark | 4383 | European |
| RISE1281.SG | Denmark | 4226 | European |
| RISE1280.SG | Denmark | 4226 | European |
| 1H04.SG | France | 4851 | European |
| 1H06.SG | France | 5115 | European |
| 1H07.SG | France | 5166 | European |
| 1H13.SG | France | 4950 | European |
| 1H14.SG | France | 5160 | European |
| 2H06.SG | France | 5164 | European |
| 2H07.SG | France | 5184 | European |
| 2H10.SG | France | 5165 | European |
| 2H11.SG | France | 5162 | European |
| 2H17.SG | France | 5157 | European |
| 2HC51961131.SG | France | 5161 | European |

| | | | |
|---|---|---|---|
| GBVPK.SG | France | 4336 | European |
| GBVPL.SG | France | 4474 | European |
| GBVPO.SG | France | 3598 | European |
| MAS15.SG | France | 4950 | European |
| ROUQCC.SG | France | 5346 | European |
| ROUQEE.SG | France | 5047 | European |
| ROUQFF.SG | France | 5166 | European |
| ROUQHH.SG | France | 5163 | European |
| ROUQV.SG | France | 5159 | European |
| ROUQW.SG | France | 5165 | European |
| TORTC.SG | France | 4476 | European |
| TORTD.SG | France | 4973 | European |
| TORTE.SG | France | 4574 | European |
| TORTF.SG | France | 3861 | European |
| AITI_62A | Germany | 3732 | European |
| AITI_65adult | Germany | 3728 | European |
| AITI_66 | Germany | 3761 | European |
| AITI_77A | Germany | 3828 | European |
| AITI_77B | Germany | 3743 | European |
| AITI_87 | Germany | 3692 | European |
| AITI_92 | Germany | 3761 | European |
| AITI_95 | Germany | 3671 | European |
| ALT_1 | Germany | 4463 | European |
| ALT_2 | Germany | 4425 | European |
| ALT_3 | Germany | 4425 | European |
| ALT_4 | Germany | 4453 | European |
| AMP_1 | Germany | 4356 | European |
| FMW | Germany | 4005 | European |
| HUGO_167 | Germany | 4089 | European |

| | | | |
|---|---|---|---|
| HUGO_168 | Germany | 4167 | European |
| HUGO_171 | Germany | 4203 | European |
| HUGO_180Sk1 | Germany | 4289 | European |
| HUGO_180Sk2 | Germany | 4307 | European |
| HUGO_190 | Germany | 4103 | European |
| OBKR_117 | Germany | 3883 | European |
| OBKR_2 | Germany | 3906 | European |
| OBKR_47 | Germany | 4012 | European |
| OBKR_50 | Germany | 3935 | European |
| OBKR_6 | Germany | 3921 | European |
| OBKR_66 | Germany | 3918 | European |
| OBKR_67 | Germany | 3906 | European |
| OBKR_73 | Germany | 3911 | European |
| OBKR_76 | Germany | 3863 | European |
| OBKR_81 | Germany | 3910 | European |
| OBKR_82 | Germany | 3908 | European |
| OBKR_84 | Germany | 3875 | European |
| OBKR_86 | Germany | 3926 | European |
| OBKR_93 | Germany | 3933 | European |
| OBKR_95 | Germany | 3917 | European |
| OBKR_96 | Germany | 3863 | European |
| OBKR_9A | Germany | 3902 | European |
| OTTM_142 | Germany | 3527 | European |
| OTTM_151ind2 | Germany | 3507 | European |
| OTTM_152 | Germany | 3513 | European |
| POST_1 | Germany | 3886 | European |
| POST_12 | Germany | 4036 | European |
| POST_131 | Germany | 3950 | European |
| POST_16 | Germany | 3922 | European |

| | | | |
|---|---|---|---|
| POST_2 | Germany | 3971 | European |
| POST_32 | Germany | 3930 | European |
| POST_35 | Germany | 3932 | European |
| POST_85 | Germany | 3946 | European |
| POST_99 | Germany | 3962 | European |
| UNTA121_FK61 | Germany | 4795 | European |
| UNTA58_147 | Germany | 3922 | European |
| UNTA58_149 | Germany | 3904 | European |
| UNTA58_152 | Germany | 3848 | European |
| UNTA58_68Sk2 | Germany | 4347 | European |
| UNTA85_1336 | Germany | 4334 | European |
| UNTA89_FK231 | Germany | 4704 | European |
| WEHR_1192SkB | Germany | 3785 | European |
| WEHR_1193 | Germany | 3823 | European |
| WEHR_1414 | Germany | 3908 | European |
| WEHR_1415adult | Germany | 3872 | European |
| WEHR_1474 | Germany | 3924 | European |
| WEHR_1564 | Germany | 3899 | European |
| WEHR_1586 | Germany | 3835 | European |
| MOK10B.SG | Serbia | 3900 | European |
| MOK12.SG | Serbia | 3900 | European |
| MOK13.SG | Serbia | 3900 | European |
| MOK14.SG | Serbia | 3900 | European |
| MOK15.SG | Serbia | 3900 | European |
| MOK16A.SG | Serbia | 3900 | European |
| MOK17A.SG | Serbia | 3900 | European |
| MOK18A.SG | Serbia | 3900 | European |
| MOK19A.SG | Serbia | 3900 | European |
| MOK20.SG | Serbia | 3900 | European |

| | | | |
|---|---|---|---|
| MOK21A.SG | Serbia | 3900 | European |
| MOK22.SG | Serbia | 3900 | European |
| MOK23.SG | Serbia | 3900 | European |
| MOK24A.SG | Serbia | 3900 | European |
| MOK25A.SG | Serbia | 3900 | European |
| MOK26A.SG | Serbia | 3900 | European |
| MOK27.SG | Serbia | 3900 | European |
| MOK28A.SG | Serbia | 3900 | European |
| MOK29A.SG | Serbia | 3900 | European |
| MOK30.SG | Serbia | 3900 | European |
| MOK31.SG | Serbia | 3900 | European |
| MOK32.SG | Serbia | 3900 | European |
| MOK33.SG | Serbia | 3900 | European |
| MOK9B.SG | Serbia | 3900 | European |
| Aes6_d | Switzerland | 4799 | European |
| Aes7_d | Switzerland | 4750 | European |
| ALA001_d | Turkey | 3378 | European |
| ALA014_d | Turkey | 3629 | European |
| ART020_d | Turkey | 5177 | European |
| ART027_d | Turkey | 5190 | European |
| I10899_published_d | Spain | 24450 | European |
| I14308_d | Russia | 6714 | European |
| I15947_d | Italy | 5575 | European |
| I2373_d | Hungary | 7700 | European |
| MX283_d | Germany | 3946 | European |
| MX286_d | Germany | 3913 | European |
| SUA001_d | Italy | 3276 | European |
| BGD001 | Russia | 2051 | European |
| BGD002 | Russia | 2051 | European |

| | | | |
|---|---|---|---|
| BGD004 | Russia | 2051 | European |
| BIY001 | Russia | 2307 | European |
| BIY002 | Russia | 2150 | European |
| BIY003 | Russia | 2230 | European |
| BIY005 | Russia | 2150 | European |
| BIY006 | Russia | 2150 | European |
| BIY007 | Russia | 2150 | European |
| BIY008 | Russia | 2150 | European |
| BIY009 | Russia | 2150 | European |
| BIY010 | Russia | 2150 | European |
| BIY011 | Russia | 2150 | European |
| BIY012 | Russia | 2260 | European |
| HUN001 | Hungary | 1566 | European |
| KOK001 | Russia | 2250 | European |
| KOK002 | Russia | 2095 | European |
| KOK003 | Russia | 2250 | European |
| SHD001 | Russia | 2300 | European |
| SHD002 | Russia | 2300 | European |
| SMV001 | Russia | 2295 | European |
| SMV002 | Russia | 2250 | European |
| VOR001 | Russia | 2250 | European |
| VOR003 | Russia | 2250 | European |
| MS20.SG | Russia | 1100 | European |
| Ash002.SG | Turkey | 9483 | European |
| Ash040.SG | Turkey | 9647 | European |
| Ash128.SG | Turkey | 10018 | European |
| Ash129.SG | Turkey | 9837 | European |
| Ash131.SG | Turkey | 9888 | European |
| Ash133.SG | Turkey | 9845 | European |

| | | | |
|---|---|---|---|
| Ash136.SG | Turkey | 9827 | European |
| CCH144.SG | Turkey | 8789 | European |
| CCH163.SG | Turkey | 8636 | European |
| CCH285.SG | Turkey | 8500 | European |
| CCH289.SG | Turkey | 8661 | European |
| CCH290.SG | Turkey | 8577 | European |
| CCH294.SG | Turkey | 8500 | European |
| CCH311.SG | Turkey | 8500 | European |
| cth006.SG | Turkey | 8517 | European |
| cth217.SG | Turkey | 8274 | European |
| cth728.SG | Turkey | 8568 | European |
| cth739.SG | Turkey | 8105 | European |
| cth747.SG | Turkey | 8506 | European |
| cth842.SG | Turkey | 8559 | European |
| pch034.SG | Turkey | 8500 | European |
| RIP001 | Italy | 16736 | European |
| I1584_all | Turkey | 5770 | European |
| PM1 | Romania | 34415 | European |
| PM1_d | Romania | 34415 | European |
| I0626_all | Vietnam | 3850 | Asian |
| I0627_all | Vietnam | 3949 | Asian |
| I1137_all_published | Vietnam | 3758 | Asian |
| I1859_all | Vietnam | 3762 | Asian |
| I2497_all | Vietnam | 1950 | Asian |
| I2731_all | Vietnam | 3684 | Asian |
| I0626 | Vietnam | 3850 | Asian |
| I0627 | Vietnam | 3949 | Asian |
| I1135 | Vietnam | 3715 | Asian |
| I1137 | Vietnam | 3758 | Asian |

| | | | |
|---|---|---|---|
| I1781 | Turkmenistan | 3832 | Asian |
| I1782 | Turkmenistan | 4163 | Asian |
| I1787 | Turkmenistan | 4023 | Asian |
| I1788 | Turkmenistan | 3947 | Asian |
| I1789 | Turkmenistan | 4082 | Asian |
| I1790 | Turkmenistan | 3994 | Asian |
| I1792 | Turkmenistan | 4254 | Asian |
| I1793 | Turkmenistan | 4038 | Asian |
| I1859 | Vietnam | 3762 | Asian |
| I2085 | Turkmenistan | 3881 | Asian |
| I2087 | Turkmenistan | 4051 | Asian |
| I2116 | Turkmenistan | 3916 | Asian |
| I2121 | Turkmenistan | 4079 | Asian |
| I2122 | Turkmenistan | 4023 | Asian |
| I2123 | Turkmenistan | 4211 | Asian |
| I2125 | Turkmenistan | 4011 | Asian |
| I2128 | Turkmenistan | 4056 | Asian |
| I2497 | Vietnam | 1950 | Asian |
| I2731 | Vietnam | 3684 | Asian |
| I2922 | Iran | 4043 | Asian |
| I2927 | Iran | 4447 | Asian |
| I2928 | Iran | 4625 | Asian |
| I2960 | Pakistan | 825 | Asian |
| I3374 | Turkmenistan | 3966 | Asian |
| I3448 | Kazakhstan | 3700 | Asian |
| I3456 | Kazakhstan | 2994 | Asian |
| I7179 | Israel | 3725 | Asian |
| I7180 | Israel | 3725 | Asian |
| I7182 | Israel | 3725 | Asian |

| | | | |
|---|---|---|---|
| I7184 | Israel | 3725 | Asian |
| I4157 | Uzbekistan | 3400 | Asian |
| I6707 | Kazakhstan | 3432 | Asian |
| I6708 | Kazakhstan | 4042 | Asian |
| I6709 | Kazakhstan | 3234 | Asian |
| I6788 | Kazakhstan | 3674 | Asian |
| I6789 | Kazakhstan | 3732 | Asian |
| I6790 | Kazakhstan | 3695 | Asian |
| I6791 | Kazakhstan | 3674 | Asian |
| I6792 | Kazakhstan | 3724 | Asian |
| I6793 | Kazakhstan | 3631 | Asian |
| I6794 | Kazakhstan | 3715 | Asian |
| I6795 | Kazakhstan | 3642 | Asian |
| I6796 | Kazakhstan | 3761 | Asian |
| I6797 | Kazakhstan | 3695 | Asian |
| I6799 | Kazakhstan | 3715 | Asian |
| I6800 | Kazakhstan | 3656 | Asian |
| I7101 | Turkmenistan | 4050 | Asian |
| I7102 | Turkmenistan | 3850 | Asian |
| I7170 | Turkmenistan | 3850 | Asian |
| I7171 | Turkmenistan | 4050 | Asian |
| I7411 | Uzbekistan | 3542 | Asian |
| I7173 | Turkmenistan | 4050 | Asian |
| I7412 | Uzbekistan | 3642 | Asian |
| I7414 | Uzbekistan | 3925 | Asian |
| I7416 | Uzbekistan | 3824 | Asian |
| I7419 | Uzbekistan | 3754 | Asian |
| I7420 | Uzbekistan | 3750 | Asian |
| I7421 | Uzbekistan | 3790 | Asian |

| | | | |
|---|---|---|---|
| I7492 | Uzbekistan | 3853 | Asian |
| I7493 | Uzbekistan | 3750 | Asian |
| I7494 | Uzbekistan | 3875 | Asian |
| I7495 | Uzbekistan | 3853 | Asian |
| I7542 | Uzbekistan | 3766 | Asian |
| I4158 | Uzbekistan | 3400 | Asian |
| I1783 | Turkmenistan | 4068 | Asian |
| Sanganji_131421-3_A1.SG | Japan | 3177 | Asian |
| Sanganji_131421-3_A2.SG | Japan | 3177 | Asian |
| Sanganji_131464_B.SG | Japan | 3280 | Asian |
| A_Han-4.DG | China | 0 | Asian |
| A_Dai-5.DG | China | 0 | Asian |
| S_Jordanian-2.DG | Jordan | 0 | Asian |
| BR_Kurumba-1.DG | India | 0 | Asian |
| S_Jordanian-3.DG | Jordan | 0 | Asian |
| S_Punjabi-2.DG | Pakistan | 0 | Asian |
| S_Naxi-2.DG | China | 0 | Asian |
| S_Irula-2.DG | India | 0 | Asian |
| S_Kalash-1.DG | Pakistan | 0 | Asian |
| S_Burusho-1.DG | Pakistan | 0 | Asian |
| S_BedouinB-1.DG | Israel | 0 | Asian |
| S_Iraqi_Jew-2.DG | Iraq | 0 | Asian |
| T_Sherpa-1.DG | Nepal | 0 | Asian |
| S_Kalash-2.DG | Pakistan | 0 | Asian |
| S_Daur-2.DG | China | 0 | Asian |
| S_Hazara-1.DG | Pakistan | 0 | Asian |
| S_BedouinB-2.DG | Israel | 0 | Asian |
| S_Burusho-2.DG | Pakistan | 0 | Asian |
| S_Mongola-2.DG | China | 0 | Asian |

| | | | |
|---|---|---|---|
| S_Oroqen-2.DG | China | 0 | Asian |
| S_Druze-1.DG | Israel | 0 | Asian |
| S_Kusunda-2.DG | Nepal | 0 | Asian |
| S_Burmese-2.DG | Myanmar | 0 | Asian |
| S_Relli-1.DG | India | 0 | Asian |
| S_Kapu-1.DG | India | 0 | Asian |
| S_Sindhi-2.DG | Pakistan | 0 | Asian |
| S_Oroqen-1.DG | China | 0 | Asian |
| S_Mongola-1.DG | China | 0 | Asian |
| S_Hazara-2.DG | Pakistan | 0 | Asian |
| S_Dusun-2.DG | Brunei | 0 | Asian |
| S_Tajik-2.DG | Tajikistan | 0 | Asian |
| S_Iranian-1.DG | Iran | 0 | Asian |
| S_Yadava-2.DG | India | 0 | Asian |
| S_Ami-1.DG | Taiwan | 0 | Asian |
| S_Cambodian-2.DG | Cambodia | 0 | Asian |
| S_Korean-1.DG | South Korea | 0 | Asian |
| S_Dai-2.DG | China | 0 | Asian |
| S_Uygur-2.DG | China | 0 | Asian |
| S_Thai-2.DG | Thailand | 0 | Asian |
| S_Brahmin-1.DG | India | 0 | Asian |
| S_Madiga-1.DG | India | 0 | Asian |
| S_Igorot-2.DG | Philippines | 0 | Asian |
| S_Uygur-1.DG | China | 0 | Asian |
| S_Punjabi-4.DG | Pakistan | 0 | Asian |
| S_Ami-2.DG | Taiwan | 0 | Asian |
| S_She-1.DG | China | 0 | Asian |
| S_Iraqi_Jew-1.DG | Iraq | 0 | Asian |
| T_Sherpa-2.DG | Nepal | 0 | Asian |

| | | | |
|---|---|---|---|
| S_Bengali-1.DG | Bangladesh | 0 | Asian |
| S_Yi-1.DG | China | 0 | Asian |
| S_Tujia-2.DG | China | 0 | Asian |
| B_Dai-4.DG | China | 0 | Asian |
| S_Atayal-1.DG | Taiwan | 0 | Asian |
| T_Tibetan-2.DG | Nepal | 0 | Asian |
| S_Irula-1.DG | India | 0 | Asian |
| S_Yadava-1.DG | India | 0 | Asian |
| S_Igorot-1.DG | Philippines | 0 | Asian |
| S_Tajik-1.DG | Tajikistan | 0 | Asian |
| S_Kinh-1.DG | Vietnam | 0 | Asian |
| S_Tujia-1.DG | China | 0 | Asian |
| S_Thai-1.DG | Thailand | 0 | Asian |
| S_Brahui-2.DG | Pakistan | 0 | Asian |
| S_Tu-1.DG | China | 0 | Asian |
| S_Pathan-1.DG | Pakistan | 0 | Asian |
| S_Sindhi-1.DG | Pakistan | 0 | Asian |
| S_Dai-3.DG | China | 0 | Asian |
| S_Han-1.DG | China | 0 | Asian |
| S_Japanese-2.DG | Japan | 0 | Asian |
| S_Makrani-2.DG | Pakistan | 0 | Asian |
| S_Balochi-1.DG | Pakistan | 0 | Asian |
| S_Brahui-1.DG | Pakistan | 0 | Asian |
| S_Miao-2.DG | China | 0 | Asian |
| BR_Mala-1.DG | India | 0 | Asian |
| S_Kinh-2.DG | Vietnam | 0 | Asian |
| S_Relli-2.DG | India | 0 | Asian |
| B_Han-3.DG | China | 0 | Asian |
| S_Mala-3.DG | India | 0 | Asian |

| | | | |
|---|---|---|---|
| S_Dusun-1.DG | Brunei | 0 | Asian |
| S_She-2.DG | China | 0 | Asian |
| S_Yemenite_Jew-1.DG | Yemen | 0 | Asian |
| S_Punjabi-3.DG | Pakistan | 0 | Asian |
| S_Iranian-2.DG | Iran | 0 | Asian |
| S_Jordanian-1.DG | Jordan | 0 | Asian |
| S_Yemenite_Jew-2.DG | Yemen | 0 | Asian |
| T_Tibetan-1.DG | Nepal | 0 | Asian |
| S_Miao-1.DG | China | 0 | Asian |
| S_Balochi-2.DG | Pakistan | 0 | Asian |
| S_Makrani-1.DG | Pakistan | 0 | Asian |
| S_Japanese-1.DG | Japan | 0 | Asian |
| S_Han-2.DG | China | 0 | Asian |
| S_Punjabi-1.DG | Pakistan | 0 | Asian |
| S_Palestinian-1.DG | Israel | 0 | Asian |
| S_Kyrgyz-2.DG | Kyrgyzstan | 0 | Asian |
| BR_Kashmiri_Pandit-1.DG | India | 0 | Asian |
| S_Tu-2.DG | China | 0 | Asian |
| S_Pathan-2.DG | Pakistan | 0 | Asian |
| S_Xibo-1.DG | China | 0 | Asian |
| S_Madiga-2.DG | India | 0 | Asian |
| S_Dai-1.DG | China | 0 | Asian |
| S_Samaritan-1.DG | Israel | 0 | Asian |
| S_Palestinian-2.DG | Israel | 0 | Asian |
| S_Hezhen-1.DG | China | 0 | Asian |
| S_Druze-2.DG | Israel | 0 | Asian |
| S_Cambodian-1.DG | Cambodia | 0 | Asian |
| S_Kapu-2.DG | India | 0 | Asian |
| S_Burmese-1.DG | Myanmar | 0 | Asian |

| | | | |
|---|---|---|---|
| S_Lahu-2.DG | China | 0 | Asian |
| S_Lahu-1.DG | China | 0 | Asian |
| S_Naxi-3.DG | China | 0 | Asian |
| S_Palestinian-3.DG | Israel | 0 | Asian |
| S_Khonda_Dora-1.DG | India | 0 | Asian |
| S_Mala-2.DG | India | 0 | Asian |
| S_Hezhen-2.DG | China | 0 | Asian |
| S_Naxi-1.DG | China | 0 | Asian |
| S_Japanese-3.DG | Japan | 0 | Asian |
| S_Brahmin-2.DG | India | 0 | Asian |
| S_Xibo-2.DG | China | 0 | Asian |
| S_Kyrgyz-1.DG | Kyrgyzstan | 0 | Asian |
| S_Korean-2.DG | South Korea | 0 | Asian |
| S_Bengali-2.DG | Bangladesh | 0 | Asian |
| S_Yi-2.DG | China | 0 | Asian |
| S_Kusunda-1.DG | Nepal | 0 | Asian |
| Manny.SG | India | 0 | Asian |
| Tadjik.SG | Tajikistan | 0 | Asian |
| M240.SG | Nepal | 2125 | Asian |
| M344.SG | Nepal | 2125 | Asian |
| S40.SG | Nepal | 1500 | Asian |
| C1.SG | Nepal | 2775 | Asian |
| M63.SG | Nepal | 2125 | Asian |
| S10.SG | Nepal | 1500 | Asian |
| S35.SG | Nepal | 1500 | Asian |
| S41.SG | Nepal | 1500 | Asian |
| AH1.SG | Iran | 9925 | Asian |
| AH2.SG | Iran | 9921 | Asian |
| AH4.SG | Iran | 9918 | Asian |

| | | | |
|---|---|---|---|
| F38.SG | Iran | 2833 | Asian |
| WC1.SG | Iran | 9214 | Asian |
| I1661 | Iran | 6539 | Asian |
| I2189_published | Israel | 3500 | Asian |
| I2190 | Israel | 3351 | Asian |
| I2195 | Israel | 3372 | Asian |
| I2198 | Israel | 3420 | Asian |
| I2200 | Israel | 3500 | Asian |
| I2201 | Israel | 2889 | Asian |
| I0563 | Kazakhstan | 2300 | Asian |
| I0562 | Kazakhstan | 2300 | Asian |
| I0507 | Kazakhstan | 3668 | Asian |
| I0644 | Israel | 5950 | Asian |
| I1180_published | Israel | 5950 | Asian |
| I1177_published | Israel | 5950 | Asian |
| I1155_published | Israel | 5950 | Asian |
| I1181_published | Israel | 5950 | Asian |
| I1172 | Israel | 5950 | Asian |
| I1187 | Israel | 5950 | Asian |
| I1171 | Israel | 5950 | Asian |
| I1164 | Israel | 5950 | Asian |
| I1184 | Israel | 5950 | Asian |
| I1179 | Israel | 5950 | Asian |
| I1182 | Israel | 5950 | Asian |
| I1170 | Israel | 5950 | Asian |
| I1168 | Israel | 5950 | Asian |
| I1152 | Israel | 5950 | Asian |
| I1178 | Israel | 5950 | Asian |
| I1290 | Iran | 9806 | Asian |

| | | | |
|---|---|---|---|
| I1705 | Jordan | 4029 | Asian |
| I1706 | Jordan | 4345 | Asian |
| I1679_published | Jordan | 8800 | Asian |
| I1709_published | Jordan | 10050 | Asian |
| I1727 | Jordan | 10050 | Asian |
| I1710 | Jordan | 9565 | Asian |
| I1707 | Jordan | 9569 | Asian |
| I1704 | Jordan | 9159 | Asian |
| I1730 | Jordan | 4356 | Asian |
| I1699 | Jordan | 8700 | Asian |
| I1670 | Iran | 6690 | Asian |
| I1662 | Iran | 6677 | Asian |
| I1674 | Iran | 5831 | Asian |
| I1665 | Iran | 5819 | Asian |
| I1671 | Iran | 7686 | Asian |
| RISE391.SG | Kazakhstan | 3924 | Asian |
| I0867 | Israel | 8700 | Asian |
| I1165 | Israel | 5950 | Asian |
| I0861 | Israel | 12700 | Asian |
| I1160 | Israel | 5950 | Asian |
| I1166 | Israel | 5950 | Asian |
| I1931 | Kazakhstan | 3343 | Asian |
| I2062 | Israel | 3170 | Asian |
| I1072_published | Israel | 12700 | Asian |
| IS2.SG | Kazakhstan | 2603 | Asian |
| Ze6b.SG | Kazakhstan | 2700 | Asian |
| I3345 | India | 124 | Asian |
| I3405 | India | 193 | Asian |
| I3406 | India | 1016 | Asian |

| | | | |
|---|---|---|---|
| I3451 | Kazakhstan | 2816 | Asian |
| I3769 | Kazakhstan | 3150 | Asian |
| I3788 | Kazakhstan | 3536 | Asian |
| I3860 | Kazakhstan | 3603 | Asian |
| I3861 | Kazakhstan | 3695 | Asian |
| I3864 | Kazakhstan | 3562 | Asian |
| I3352 | India | 1166 | Asian |
| I3342 | India | 1099 | Asian |
| I3350 | India | 131 | Asian |
| I3767 | Kazakhstan | 3684 | Asian |
| I3403 | India | 128 | Asian |
| I3404 | India | 129 | Asian |
| I3343 | India | 1109 | Asian |
| I3344 | India | 1107 | Asian |
| I3346 | India | 1129 | Asian |
| I3401_published | India | 150 | Asian |
| I3407 | India | 1070 | Asian |
| I3772 | Kazakhstan | 2911 | Asian |
| I3770 | Kazakhstan | 3968 | Asian |
| I3351 | India | 1118 | Asian |
| I3402 | India | 1118 | Asian |
| I3348 | India | 126 | Asian |
| I3349 | India | 1101 | Asian |
| I3611 | Taiwan | 1715 | Asian |
| I3612 | Taiwan | 1800 | Asian |
| I3753 | Kazakhstan | 2977 | Asian |
| I3763 | Kazakhstan | 3445 | Asian |
| I3965 | Israel | 3650 | Asian |
| I3966 | Israel | 3650 | Asian |

| | | | |
|---|---|---|---|
| I3976 | Kazakhstan | 3035 | Asian |
| I3977 | Kazakhstan | 3015 | Asian |
| I4085 | Turkmenistan | 5450 | Asian |
| I4086 | Turkmenistan | 5450 | Asian |
| I4087 | Turkmenistan | 5450 | Asian |
| I4153 | Uzbekistan | 3050 | Asian |
| I4156 | Uzbekistan | 3400 | Asian |
| I4159 | Uzbekistan | 3400 | Asian |
| I4161 | Uzbekistan | 3900 | Asian |
| I4255 | Uzbekistan | 3050 | Asian |
| I4257 | Tajikistan | 3423 | Asian |
| I4258 | Tajikistan | 3504 | Asian |
| I4259 | Turkmenistan | 5021 | Asian |
| I4262 | Kazakhstan | 3743 | Asian |
| I4263 | Kazakhstan | 3658 | Asian |
| I4264 | Kazakhstan | 3551 | Asian |
| I4265 | Kazakhstan | 3522 | Asian |
| I4267 | Kazakhstan | 3042 | Asian |
| I4285 | Uzbekistan | 3685 | Asian |
| I4286 | Uzbekistan | 3768 | Asian |
| I4288 | Uzbekistan | 3750 | Asian |
| I4289 | Uzbekistan | 3790 | Asian |
| I1784 | Turkmenistan | 4060 | Asian |
| I1934_published | Israel | 3200 | Asian |
| I1955 | Iran | 389 | Asian |
| I2513 | Iran | 4564 | Asian |
| I2514 | Iran | 4347 | Asian |
| I2923 | Iran | 4707 | Asian |
| I2924 | Iran | 4715 | Asian |

| | | | |
|---|---|---|---|
| I2925 | Iran | 4715 | Asian |
| I2954 | Pakistan | 2316 | Asian |
| I2959 | Pakistan | 822 | Asian |
| I3447 | Kazakhstan | 4577 | Asian |
| Tianyuan | China | 39565 | Asian |
| I4295 | Kazakhstan | 3090 | Asian |
| I4312 | Uzbekistan | 3609 | Asian |
| I4313 | Uzbekistan | 3421 | Asian |
| I4314 | Uzbekistan | 3756 | Asian |
| I4315 | Uzbekistan | 3461 | Asian |
| I4318 | Kazakhstan | 3656 | Asian |
| I4321 | Kazakhstan | 3522 | Asian |
| I4322 | Kazakhstan | 3484 | Asian |
| I4323 | Kazakhstan | 3622 | Asian |
| I4351 | Iran | 7920 | Asian |
| I4517 | Israel | 2951 | Asian |
| I1169 | Israel | 5950 | Asian |
| I4518 | Israel | 3375 | Asian |
| I4519 | Israel | 3425 | Asian |
| I4521 | Israel | 4194 | Asian |
| I4525 | Israel | 3500 | Asian |
| I4566 | Kazakhstan | 3573 | Asian |
| I4773 | Kazakhstan | 3508 | Asian |
| I4774 | Kazakhstan | 3503 | Asian |
| I4776 | Kazakhstan | 3656 | Asian |
| I4778 | Kazakhstan | 3562 | Asian |
| I4779 | Kazakhstan | 3631 | Asian |
| I4782 | Kazakhstan | 3609 | Asian |
| I4783 | Kazakhstan | 3456 | Asian |

| | | | |
|---|---|---|---|
| I4784 | Kazakhstan | 3508 | Asian |
| I4787 | Kazakhstan | 3216 | Asian |
| HG00403.SG | China | 0 | Asian |
| HG00404.SG | China | 0 | Asian |
| HG00406.SG | China | 0 | Asian |
| HG00407.SG | China | 0 | Asian |
| HG00409.SG | China | 0 | Asian |
| HG00410.SG | China | 0 | Asian |
| HG00419.SG | China | 0 | Asian |
| HG00421.SG | China | 0 | Asian |
| HG00422.SG | China | 0 | Asian |
| HG00428.SG | China | 0 | Asian |
| HG00436.SG | China | 0 | Asian |
| HG00437.SG | China | 0 | Asian |
| HG00442.SG | China | 0 | Asian |
| HG00443.SG | China | 0 | Asian |
| HG00445.SG | China | 0 | Asian |
| HG00446.SG | China | 0 | Asian |
| HG00448.SG | China | 0 | Asian |
| HG00449.SG | China | 0 | Asian |
| HG00451.SG | China | 0 | Asian |
| HG00452.SG | China | 0 | Asian |
| HG00457.SG | China | 0 | Asian |
| HG00458.SG | China | 0 | Asian |
| HG00463.SG | China | 0 | Asian |
| HG00464.SG | China | 0 | Asian |
| HG00472.SG | China | 0 | Asian |
| HG00473.SG | China | 0 | Asian |
| HG00475.SG | China | 0 | Asian |

| | | | |
|---|---|---|---|
| HG00476.SG | China | 0 | Asian |
| HG00478.SG | China | 0 | Asian |
| HG00479.SG | China | 0 | Asian |
| HG00500.SG | China | 0 | Asian |
| HG00501.SG | China | 0 | Asian |
| HG00513.SG | China | 0 | Asian |
| HG00524.SG | China | 0 | Asian |
| HG00525.SG | China | 0 | Asian |
| HG00530.SG | China | 0 | Asian |
| HG00531.SG | China | 0 | Asian |
| HG00533.SG | China | 0 | Asian |
| HG00534.SG | China | 0 | Asian |
| HG00536.SG | China | 0 | Asian |
| HG00537.SG | China | 0 | Asian |
| HG00542.SG | China | 0 | Asian |
| HG00543.SG | China | 0 | Asian |
| HG00556.SG | China | 0 | Asian |
| HG00557.SG | China | 0 | Asian |
| HG00559.SG | China | 0 | Asian |
| HG00560.SG | China | 0 | Asian |
| HG00565.SG | China | 0 | Asian |
| HG00566.SG | China | 0 | Asian |
| HG00580.SG | China | 0 | Asian |
| HG00581.SG | China | 0 | Asian |
| HG00583.SG | China | 0 | Asian |
| HG00584.SG | China | 0 | Asian |
| HG00589.SG | China | 0 | Asian |
| HG00590.SG | China | 0 | Asian |
| HG00592.SG | China | 0 | Asian |

| | | | |
|---|---|---|---|
| HG00593.SG | China | 0 | Asian |
| HG00595.SG | China | 0 | Asian |
| HG00596.SG | China | 0 | Asian |
| HG00598.SG | China | 0 | Asian |
| HG00599.SG | China | 0 | Asian |
| HG00607.SG | China | 0 | Asian |
| HG00608.SG | China | 0 | Asian |
| HG00610.SG | China | 0 | Asian |
| HG00611.SG | China | 0 | Asian |
| HG00613.SG | China | 0 | Asian |
| HG00614.SG | China | 0 | Asian |
| HG00619.SG | China | 0 | Asian |
| HG00620.SG | China | 0 | Asian |
| HG00622.SG | China | 0 | Asian |
| HG00623.SG | China | 0 | Asian |
| HG00625.SG | China | 0 | Asian |
| HG00626.SG | China | 0 | Asian |
| HG00628.SG | China | 0 | Asian |
| HG00629.SG | China | 0 | Asian |
| HG00631.SG | China | 0 | Asian |
| HG00632.SG | China | 0 | Asian |
| HG00634.SG | China | 0 | Asian |
| HG00635.SG | China | 0 | Asian |
| HG00650.SG | China | 0 | Asian |
| HG00651.SG | China | 0 | Asian |
| HG00653.SG | China | 0 | Asian |
| HG00654.SG | China | 0 | Asian |
| HG00656.SG | China | 0 | Asian |
| HG00657.SG | China | 0 | Asian |

| | | | |
|---|---|---|---|
| HG00662.SG | China | 0 | Asian |
| HG00663.SG | China | 0 | Asian |
| HG00671.SG | China | 0 | Asian |
| HG00672.SG | China | 0 | Asian |
| HG00674.SG | China | 0 | Asian |
| HG00675.SG | China | 0 | Asian |
| HG00683.SG | China | 0 | Asian |
| HG00684.SG | China | 0 | Asian |
| HG00689.SG | China | 0 | Asian |
| HG00690.SG | China | 0 | Asian |
| HG00692.SG | China | 0 | Asian |
| HG00693.SG | China | 0 | Asian |
| HG00698.SG | China | 0 | Asian |
| HG00699.SG | China | 0 | Asian |
| HG00701.SG | China | 0 | Asian |
| HG00702.SG | China | 0 | Asian |
| HG00704.SG | China | 0 | Asian |
| HG00705.SG | China | 0 | Asian |
| HG00707.SG | China | 0 | Asian |
| HG00708.SG | China | 0 | Asian |
| HG00717.SG | China | 0 | Asian |
| HG00728.SG | China | 0 | Asian |
| HG00729.SG | China | 0 | Asian |
| HG00759.SG | China | 0 | Asian |
| HG00766.SG | China | 0 | Asian |
| HG00844.SG | China | 0 | Asian |
| HG00851.SG | China | 0 | Asian |
| HG00864.SG | China | 0 | Asian |
| HG00867.SG | China | 0 | Asian |

| | | | |
|---|---|---|---|
| HG00879.SG | China | 0 | Asian |
| HG00881.SG | China | 0 | Asian |
| HG00956.SG | China | 0 | Asian |
| HG00978.SG | China | 0 | Asian |
| HG00982.SG | China | 0 | Asian |
| HG01028.SG | China | 0 | Asian |
| HG01029.SG | China | 0 | Asian |
| HG01031.SG | China | 0 | Asian |
| HG01046.SG | China | 0 | Asian |
| HG01583.SG | Pakistan | 0 | Asian |
| HG01586.SG | Pakistan | 0 | Asian |
| HG01589.SG | Pakistan | 0 | Asian |
| HG01593.SG | Pakistan | 0 | Asian |
| HG01595.SG | Vietnam | 0 | Asian |
| HG01596.SG | Vietnam | 0 | Asian |
| HG01597.SG | Vietnam | 0 | Asian |
| HG01598.SG | Vietnam | 0 | Asian |
| HG01599.SG | Vietnam | 0 | Asian |
| HG01600.SG | Vietnam | 0 | Asian |
| HG01794.SG | China | 0 | Asian |
| HG01795.SG | China | 0 | Asian |
| HG01796.SG | China | 0 | Asian |
| HG01797.SG | China | 0 | Asian |
| HG01798.SG | China | 0 | Asian |
| HG01799.SG | China | 0 | Asian |
| HG01800.SG | China | 0 | Asian |
| HG01801.SG | China | 0 | Asian |
| HG01802.SG | China | 0 | Asian |
| HGDP01319.SDG | China | 0 | Asian |

| | | | |
|---|---|---|---|
| HG01805.SG | China | 0 | Asian |
| HG01806.SG | China | 0 | Asian |
| HG01807.SG | China | 0 | Asian |
| HG01808.SG | China | 0 | Asian |
| HG01809.SG | China | 0 | Asian |
| HG01810.SG | China | 0 | Asian |
| HG01811.SG | China | 0 | Asian |
| HG01812.SG | China | 0 | Asian |
| HG01813.SG | China | 0 | Asian |
| HG01815.SG | China | 0 | Asian |
| HG01816.SG | China | 0 | Asian |
| HG01817.SG | China | 0 | Asian |
| HG01840.SG | Vietnam | 0 | Asian |
| HG01841.SG | Vietnam | 0 | Asian |
| HG01842.SG | Vietnam | 0 | Asian |
| HG01843.SG | Vietnam | 0 | Asian |
| HG01844.SG | Vietnam | 0 | Asian |
| HG01845.SG | Vietnam | 0 | Asian |
| HG01846.SG | Vietnam | 0 | Asian |
| HG01847.SG | Vietnam | 0 | Asian |
| HG01848.SG | Vietnam | 0 | Asian |
| HG01849.SG | Vietnam | 0 | Asian |
| HG01850.SG | Vietnam | 0 | Asian |
| HG01851.SG | Vietnam | 0 | Asian |
| HG01852.SG | Vietnam | 0 | Asian |
| HG01853.SG | Vietnam | 0 | Asian |
| HG01855.SG | Vietnam | 0 | Asian |
| HG01857.SG | Vietnam | 0 | Asian |
| HG01858.SG | Vietnam | 0 | Asian |

| | | | |
|---|---|---|---|
| HG01859.SG | Vietnam | 0 | Asian |
| HG01860.SG | Vietnam | 0 | Asian |
| HG01861.SG | Vietnam | 0 | Asian |
| HG01862.SG | Vietnam | 0 | Asian |
| HG01863.SG | Vietnam | 0 | Asian |
| HG01864.SG | Vietnam | 0 | Asian |
| HG01865.SG | Vietnam | 0 | Asian |
| HG01866.SG | Vietnam | 0 | Asian |
| HG01867.SG | Vietnam | 0 | Asian |
| HG01868.SG | Vietnam | 0 | Asian |
| HG01869.SG | Vietnam | 0 | Asian |
| HG01870.SG | Vietnam | 0 | Asian |
| HG01871.SG | Vietnam | 0 | Asian |
| HG01872.SG | Vietnam | 0 | Asian |
| HG01873.SG | Vietnam | 0 | Asian |
| HG01874.SG | Vietnam | 0 | Asian |
| HG01878.SG | Vietnam | 0 | Asian |
| HG02016.SG | Vietnam | 0 | Asian |
| HG02017.SG | Vietnam | 0 | Asian |
| HG02019.SG | Vietnam | 0 | Asian |
| HG02020.SG | Vietnam | 0 | Asian |
| HG02023.SG | Vietnam | 0 | Asian |
| HG02024.SG | Vietnam | 0 | Asian |
| HG02025.SG | Vietnam | 0 | Asian |
| HG02026.SG | Vietnam | 0 | Asian |
| HG02028.SG | Vietnam | 0 | Asian |
| HG02029.SG | Vietnam | 0 | Asian |
| HG02031.SG | Vietnam | 0 | Asian |
| HG02032.SG | Vietnam | 0 | Asian |

| | | | |
|---|---|---|---|
| HG02035.SG | Vietnam | 0 | Asian |
| HG02040.SG | Vietnam | 0 | Asian |
| HG02046.SG | Vietnam | 0 | Asian |
| HG02047.SG | Vietnam | 0 | Asian |
| HG02048.SG | Vietnam | 0 | Asian |
| HG02049.SG | Vietnam | 0 | Asian |
| HG02050.SG | Vietnam | 0 | Asian |
| HG02057.SG | Vietnam | 0 | Asian |
| HG02058.SG | Vietnam | 0 | Asian |
| HG02060.SG | Vietnam | 0 | Asian |
| HG02061.SG | Vietnam | 0 | Asian |
| HG02064.SG | Vietnam | 0 | Asian |
| HG02067.SG | Vietnam | 0 | Asian |
| HG02069.SG | Vietnam | 0 | Asian |
| HG02070.SG | Vietnam | 0 | Asian |
| HG02072.SG | Vietnam | 0 | Asian |
| HG02073.SG | Vietnam | 0 | Asian |
| HG02075.SG | Vietnam | 0 | Asian |
| HG02076.SG | Vietnam | 0 | Asian |
| HG02078.SG | Vietnam | 0 | Asian |
| HG02079.SG | Vietnam | 0 | Asian |
| HG02081.SG | Vietnam | 0 | Asian |
| HG02082.SG | Vietnam | 0 | Asian |
| HG02084.SG | Vietnam | 0 | Asian |
| HG02085.SG | Vietnam | 0 | Asian |
| HG02086.SG | Vietnam | 0 | Asian |
| HG02087.SG | Vietnam | 0 | Asian |
| HG02088.SG | Vietnam | 0 | Asian |
| HG02113.SG | Vietnam | 0 | Asian |

| | | | |
|---|---|---|---|
| HG02116.SG | Vietnam | 0 | Asian |
| HG02121.SG | Vietnam | 0 | Asian |
| HG02122.SG | Vietnam | 0 | Asian |
| HG02127.SG | Vietnam | 0 | Asian |
| HG02128.SG | Vietnam | 0 | Asian |
| HG02130.SG | Vietnam | 0 | Asian |
| HG02131.SG | Vietnam | 0 | Asian |
| HG02133.SG | Vietnam | 0 | Asian |
| HG02134.SG | Vietnam | 0 | Asian |
| HG02136.SG | Vietnam | 0 | Asian |
| HG02137.SG | Vietnam | 0 | Asian |
| HG02138.SG | Vietnam | 0 | Asian |
| HG02139.SG | Vietnam | 0 | Asian |
| HG02140.SG | Vietnam | 0 | Asian |
| HG02141.SG | Vietnam | 0 | Asian |
| HG02142.SG | Vietnam | 0 | Asian |
| HG02151.SG | China | 0 | Asian |
| HG02152.SG | China | 0 | Asian |
| HG02153.SG | China | 0 | Asian |
| HG02154.SG | China | 0 | Asian |
| HG02155.SG | China | 0 | Asian |
| HG02156.SG | China | 0 | Asian |
| HG02164.SG | China | 0 | Asian |
| HG02165.SG | China | 0 | Asian |
| HG02166.SG | China | 0 | Asian |
| HG02178.SG | China | 0 | Asian |
| HG02179.SG | China | 0 | Asian |
| HG02180.SG | China | 0 | Asian |
| HG02181.SG | China | 0 | Asian |

| | | | |
|---|---|---|---|
| HG02182.SG | China | 0 | Asian |
| HG02184.SG | China | 0 | Asian |
| HG02185.SG | China | 0 | Asian |
| HG02186.SG | China | 0 | Asian |
| HG02187.SG | China | 0 | Asian |
| HG02188.SG | China | 0 | Asian |
| HG02190.SG | China | 0 | Asian |
| HG02250.SG | China | 0 | Asian |
| HG02351.SG | China | 0 | Asian |
| HG02353.SG | China | 0 | Asian |
| HG02355.SG | China | 0 | Asian |
| HG02356.SG | China | 0 | Asian |
| HG02360.SG | China | 0 | Asian |
| HG02363.SG | China | 0 | Asian |
| HG02364.SG | China | 0 | Asian |
| HG02367.SG | China | 0 | Asian |
| HG02371.SG | China | 0 | Asian |
| HG02372.SG | China | 0 | Asian |
| HG02373.SG | China | 0 | Asian |
| HG02374.SG | China | 0 | Asian |
| HG02375.SG | China | 0 | Asian |
| HG02377.SG | China | 0 | Asian |
| HG02379.SG | China | 0 | Asian |
| HG02380.SG | China | 0 | Asian |
| HG02381.SG | China | 0 | Asian |
| HG02382.SG | China | 0 | Asian |
| HG02383.SG | China | 0 | Asian |
| HG02384.SG | China | 0 | Asian |
| HG02385.SG | China | 0 | Asian |

| | | | |
|---|---|---|---|
| HG02386.SG | China | 0 | Asian |
| HG02387.SG | China | 0 | Asian |
| HG02388.SG | China | 0 | Asian |
| HG02389.SG | China | 0 | Asian |
| HG02390.SG | China | 0 | Asian |
| HG02391.SG | China | 0 | Asian |
| HG02392.SG | China | 0 | Asian |
| HG02394.SG | China | 0 | Asian |
| HG02395.SG | China | 0 | Asian |
| HG02396.SG | China | 0 | Asian |
| HG02397.SG | China | 0 | Asian |
| HG02398.SG | China | 0 | Asian |
| HG02399.SG | China | 0 | Asian |
| HG02401.SG | China | 0 | Asian |
| HG02402.SG | China | 0 | Asian |
| HG02406.SG | China | 0 | Asian |
| HG02407.SG | China | 0 | Asian |
| HG02408.SG | China | 0 | Asian |
| HG02409.SG | China | 0 | Asian |
| HG02410.SG | China | 0 | Asian |
| HG02490.SG | Pakistan | 0 | Asian |
| HG02491.SG | Pakistan | 0 | Asian |
| HG02493.SG | Pakistan | 0 | Asian |
| HG02494.SG | Pakistan | 0 | Asian |
| HG02512.SG | Vietnam | 0 | Asian |
| HG02513.SG | Vietnam | 0 | Asian |
| HG02521.SG | Vietnam | 0 | Asian |
| HG02522.SG | Vietnam | 0 | Asian |
| HG02597.SG | Pakistan | 0 | Asian |

| | | | |
|---|---|---|---|
| HG02600.SG | Pakistan | 0 | Asian |
| HG02601.SG | Pakistan | 0 | Asian |
| HG02603.SG | Pakistan | 0 | Asian |
| HG02604.SG | Pakistan | 0 | Asian |
| HG02648.SG | Pakistan | 0 | Asian |
| HG02649.SG | Pakistan | 0 | Asian |
| HG02651.SG | Pakistan | 0 | Asian |
| HG02652.SG | Pakistan | 0 | Asian |
| HG02654.SG | Pakistan | 0 | Asian |
| HG02655.SG | Pakistan | 0 | Asian |
| HG02657.SG | Pakistan | 0 | Asian |
| HG02658.SG | Pakistan | 0 | Asian |
| HG02660.SG | Pakistan | 0 | Asian |
| HG02661.SG | Pakistan | 0 | Asian |
| HG02681.SG | Pakistan | 0 | Asian |
| HG02682.SG | Pakistan | 0 | Asian |
| HG02684.SG | Pakistan | 0 | Asian |
| HG02685.SG | Pakistan | 0 | Asian |
| HG02687.SG | Pakistan | 0 | Asian |
| HG02688.SG | Pakistan | 0 | Asian |
| HG02690.SG | Pakistan | 0 | Asian |
| HG02691.SG | Pakistan | 0 | Asian |
| HG02694.SG | Pakistan | 0 | Asian |
| HG02696.SG | Pakistan | 0 | Asian |
| HG02697.SG | Pakistan | 0 | Asian |
| HG02699.SG | Pakistan | 0 | Asian |
| HG02700.SG | Pakistan | 0 | Asian |
| HG02724.SG | Pakistan | 0 | Asian |
| HG02725.SG | Pakistan | 0 | Asian |

| | | | |
|---|---|---|---|
| HG02727.SG | Pakistan | 0 | Asian |
| HG02728.SG | Pakistan | 0 | Asian |
| HG02731.SG | Pakistan | 0 | Asian |
| HG02733.SG | Pakistan | 0 | Asian |
| HG02734.SG | Pakistan | 0 | Asian |
| HG02736.SG | Pakistan | 0 | Asian |
| HG02737.SG | Pakistan | 0 | Asian |
| HG02774.SG | Pakistan | 0 | Asian |
| HG02775.SG | Pakistan | 0 | Asian |
| HG02778.SG | Pakistan | 0 | Asian |
| HG02780.SG | Pakistan | 0 | Asian |
| HG02783.SG | Pakistan | 0 | Asian |
| HG02784.SG | Pakistan | 0 | Asian |
| HG02786.SG | Pakistan | 0 | Asian |
| HG02787.SG | Pakistan | 0 | Asian |
| HG02789.SG | Pakistan | 0 | Asian |
| HG02790.SG | Pakistan | 0 | Asian |
| HG02792.SG | Pakistan | 0 | Asian |
| HG02793.SG | Pakistan | 0 | Asian |
| HG03006.SG | Bangladesh | 0 | Asian |
| HG03007.SG | Bangladesh | 0 | Asian |
| HG03009.SG | Bangladesh | 0 | Asian |
| HG03012.SG | Bangladesh | 0 | Asian |
| HG03015.SG | Pakistan | 0 | Asian |
| HG03016.SG | Pakistan | 0 | Asian |
| HG03018.SG | Pakistan | 0 | Asian |
| HG03019.SG | Pakistan | 0 | Asian |
| HG03021.SG | Pakistan | 0 | Asian |
| HG03022.SG | Pakistan | 0 | Asian |

| | | | |
|---|---|---|---|
| HG03228.SG | Pakistan | 0 | Asian |
| HG03229.SG | Pakistan | 0 | Asian |
| HG03234.SG | Pakistan | 0 | Asian |
| HG03235.SG | Pakistan | 0 | Asian |
| HG03237.SG | Pakistan | 0 | Asian |
| HG03238.SG | Pakistan | 0 | Asian |
| HG03488.SG | Pakistan | 0 | Asian |
| HG03490.SG | Pakistan | 0 | Asian |
| HG03491.SG | Pakistan | 0 | Asian |
| HG03585.SG | Bangladesh | 0 | Asian |
| HG03589.SG | Bangladesh | 0 | Asian |
| HG03593.SG | Bangladesh | 0 | Asian |
| HG03594.SG | Bangladesh | 0 | Asian |
| HG03595.SG | Bangladesh | 0 | Asian |
| HG03598.SG | Bangladesh | 0 | Asian |
| HG03600.SG | Bangladesh | 0 | Asian |
| HG03603.SG | Bangladesh | 0 | Asian |
| HG03604.SG | Bangladesh | 0 | Asian |
| HG03607.SG | Bangladesh | 0 | Asian |
| HG03611.SG | Bangladesh | 0 | Asian |
| HG03615.SG | Bangladesh | 0 | Asian |
| HG03616.SG | Bangladesh | 0 | Asian |
| HG03619.SG | Pakistan | 0 | Asian |
| HG03624.SG | Pakistan | 0 | Asian |
| HG03625.SG | Pakistan | 0 | Asian |
| HG03629.SG | Pakistan | 0 | Asian |
| HG03631.SG | Pakistan | 0 | Asian |
| HG03634.SG | Pakistan | 0 | Asian |
| HG03636.SG | Pakistan | 0 | Asian |

| | | | |
|---|---|---|---|
| HG03640.SG | Pakistan | 0 | Asian |
| HG03642.SG | Sri Lanka | 0 | Asian |
| HG03643.SG | Sri Lanka | 0 | Asian |
| HG03644.SG | Sri Lanka | 0 | Asian |
| HG03645.SG | Sri Lanka | 0 | Asian |
| HG03646.SG | Sri Lanka | 0 | Asian |
| HG03649.SG | Pakistan | 0 | Asian |
| HG03652.SG | Pakistan | 0 | Asian |
| HG03653.SG | Pakistan | 0 | Asian |
| HG03660.SG | Pakistan | 0 | Asian |
| HG03663.SG | Pakistan | 0 | Asian |
| HG03667.SG | Pakistan | 0 | Asian |
| HG03668.SG | Pakistan | 0 | Asian |
| HG03672.SG | Sri Lanka | 0 | Asian |
| HG03673.SG | Sri Lanka | 0 | Asian |
| HG03679.SG | Sri Lanka | 0 | Asian |
| HG03680.SG | Sri Lanka | 0 | Asian |
| HG03681.SG | Sri Lanka | 0 | Asian |
| HG03684.SG | Sri Lanka | 0 | Asian |
| HG03685.SG | Sri Lanka | 0 | Asian |
| HG03686.SG | Sri Lanka | 0 | Asian |
| HG03687.SG | Sri Lanka | 0 | Asian |
| HG03689.SG | Sri Lanka | 0 | Asian |
| HG03690.SG | Sri Lanka | 0 | Asian |
| HG03691.SG | Sri Lanka | 0 | Asian |
| HG03692.SG | Sri Lanka | 0 | Asian |
| HG03693.SG | Sri Lanka | 0 | Asian |
| HG03694.SG | Sri Lanka | 0 | Asian |
| HG03695.SG | Sri Lanka | 0 | Asian |

| | | | |
|---|---|---|---|
| HG03696.SG | Sri Lanka | 0 | Asian |
| HG03697.SG | Sri Lanka | 0 | Asian |
| HG03698.SG | Sri Lanka | 0 | Asian |
| HG03702.SG | Pakistan | 0 | Asian |
| HG03703.SG | Pakistan | 0 | Asian |
| HG03705.SG | Pakistan | 0 | Asian |
| HG03706.SG | Pakistan | 0 | Asian |
| HG03708.SG | Pakistan | 0 | Asian |
| HG03709.SG | Pakistan | 0 | Asian |
| HG03711.SG | Sri Lanka | 0 | Asian |
| HG03713.SG | India | 0 | Asian |
| HG03714.SG | India | 0 | Asian |
| HG03715.SG | India | 0 | Asian |
| HG03716.SG | India | 0 | Asian |
| HG03717.SG | India | 0 | Asian |
| HG03718.SG | India | 0 | Asian |
| HG03720.SG | India | 0 | Asian |
| HG03722.SG | India | 0 | Asian |
| HG03727.SG | India | 0 | Asian |
| HG03729.SG | India | 0 | Asian |
| HG03730.SG | India | 0 | Asian |
| HG03731.SG | India | 0 | Asian |
| HG03733.SG | Sri Lanka | 0 | Asian |
| HG03736.SG | Sri Lanka | 0 | Asian |
| HG03738.SG | Sri Lanka | 0 | Asian |
| HG03740.SG | Sri Lanka | 0 | Asian |
| HG03741.SG | Sri Lanka | 0 | Asian |
| HG03742.SG | India | 0 | Asian |
| HG03743.SG | Sri Lanka | 0 | Asian |

| | | | |
|---|---|---|---|
| HG03744.SG | Sri Lanka | 0 | Asian |
| HG03745.SG | Sri Lanka | 0 | Asian |
| HG03746.SG | Sri Lanka | 0 | Asian |
| HG03750.SG | Sri Lanka | 0 | Asian |
| HG03752.SG | Sri Lanka | 0 | Asian |
| HG03753.SG | Sri Lanka | 0 | Asian |
| HG03754.SG | Sri Lanka | 0 | Asian |
| HG03755.SG | Sri Lanka | 0 | Asian |
| HG03756.SG | Sri Lanka | 0 | Asian |
| HG03757.SG | Sri Lanka | 0 | Asian |
| HG03760.SG | Sri Lanka | 0 | Asian |
| HG03762.SG | Pakistan | 0 | Asian |
| HG03765.SG | Pakistan | 0 | Asian |
| HG03767.SG | Pakistan | 0 | Asian |
| HG03770.SG | India | 0 | Asian |
| HG03771.SG | India | 0 | Asian |
| HG03772.SG | India | 0 | Asian |
| HG03773.SG | India | 0 | Asian |
| HG03774.SG | India | 0 | Asian |
| HG03775.SG | India | 0 | Asian |
| HG03777.SG | India | 0 | Asian |
| HG03778.SG | India | 0 | Asian |
| HG03779.SG | India | 0 | Asian |
| HG03780.SG | India | 0 | Asian |
| HG03781.SG | India | 0 | Asian |
| HG03782.SG | India | 0 | Asian |
| HG03784.SG | India | 0 | Asian |
| HG03785.SG | India | 0 | Asian |
| HG03786.SG | India | 0 | Asian |

| | | | |
|---|---|---|---|
| HG03787.SG | India | 0 | Asian |
| HG03788.SG | India | 0 | Asian |
| HG03789.SG | India | 0 | Asian |
| HG03790.SG | India | 0 | Asian |
| HG03792.SG | India | 0 | Asian |
| HG03793.SG | Bangladesh | 0 | Asian |
| HG03796.SG | Bangladesh | 0 | Asian |
| HG03800.SG | Bangladesh | 0 | Asian |
| HG03802.SG | Bangladesh | 0 | Asian |
| HG03803.SG | Bangladesh | 0 | Asian |
| HG03805.SG | Bangladesh | 0 | Asian |
| HG03808.SG | Bangladesh | 0 | Asian |
| HG03809.SG | Bangladesh | 0 | Asian |
| HG03812.SG | Bangladesh | 0 | Asian |
| HG03814.SG | Bangladesh | 0 | Asian |
| HG03815.SG | Bangladesh | 0 | Asian |
| HG03817.SG | Bangladesh | 0 | Asian |
| HG03821.SG | Bangladesh | 0 | Asian |
| HG03823.SG | Bangladesh | 0 | Asian |
| HG03824.SG | Bangladesh | 0 | Asian |
| HG03826.SG | Bangladesh | 0 | Asian |
| HG03829.SG | Bangladesh | 0 | Asian |
| HG03830.SG | Bangladesh | 0 | Asian |
| HG03832.SG | Bangladesh | 0 | Asian |
| HG03833.SG | Bangladesh | 0 | Asian |
| HG03836.SG | Sri Lanka | 0 | Asian |
| HG03837.SG | Sri Lanka | 0 | Asian |
| HG03838.SG | Sri Lanka | 0 | Asian |
| HG03844.SG | Sri Lanka | 0 | Asian |

| | | | |
|---|---|---|---|
| HG03846.SG | Sri Lanka | 0 | Asian |
| HG03848.SG | Sri Lanka | 0 | Asian |
| HG03849.SG | Sri Lanka | 0 | Asian |
| HG03850.SG | Sri Lanka | 0 | Asian |
| HG03851.SG | Sri Lanka | 0 | Asian |
| HG03854.SG | Sri Lanka | 0 | Asian |
| HG03856.SG | Sri Lanka | 0 | Asian |
| HG03857.SG | Sri Lanka | 0 | Asian |
| HG03858.SG | Sri Lanka | 0 | Asian |
| HG03861.SG | India | 0 | Asian |
| HG03862.SG | India | 0 | Asian |
| HG03863.SG | India | 0 | Asian |
| HG03864.SG | India | 0 | Asian |
| HG03866.SG | India | 0 | Asian |
| HG03867.SG | India | 0 | Asian |
| HG03868.SG | India | 0 | Asian |
| HG03869.SG | India | 0 | Asian |
| HG03870.SG | India | 0 | Asian |
| HG03871.SG | India | 0 | Asian |
| HG03872.SG | India | 0 | Asian |
| HG03873.SG | India | 0 | Asian |
| HG03874.SG | India | 0 | Asian |
| HG03875.SG | India | 0 | Asian |
| HG03882.SG | India | 0 | Asian |
| HG03884.SG | Sri Lanka | 0 | Asian |
| HG03885.SG | Sri Lanka | 0 | Asian |
| HG03886.SG | Sri Lanka | 0 | Asian |
| HG03887.SG | Sri Lanka | 0 | Asian |
| HG03888.SG | Sri Lanka | 0 | Asian |

| | | | |
|---|---|---|---|
| HG03890.SG | Sri Lanka | 0 | Asian |
| HG03894.SG | Sri Lanka | 0 | Asian |
| HG03895.SG | Sri Lanka | 0 | Asian |
| HG03896.SG | Sri Lanka | 0 | Asian |
| HG03897.SG | Sri Lanka | 0 | Asian |
| HG03898.SG | Sri Lanka | 0 | Asian |
| HG03899.SG | Sri Lanka | 0 | Asian |
| HG03900.SG | Sri Lanka | 0 | Asian |
| HG03902.SG | Bangladesh | 0 | Asian |
| HG03905.SG | Bangladesh | 0 | Asian |
| HG03907.SG | Bangladesh | 0 | Asian |
| HG03908.SG | Bangladesh | 0 | Asian |
| HG03910.SG | Bangladesh | 0 | Asian |
| HG03911.SG | Bangladesh | 0 | Asian |
| HG03913.SG | Bangladesh | 0 | Asian |
| HG03914.SG | Bangladesh | 0 | Asian |
| HG03916.SG | Bangladesh | 0 | Asian |
| HG03917.SG | Bangladesh | 0 | Asian |
| HG03919.SG | Bangladesh | 0 | Asian |
| HG03920.SG | Bangladesh | 0 | Asian |
| HG03922.SG | Bangladesh | 0 | Asian |
| HG03925.SG | Bangladesh | 0 | Asian |
| HG03926.SG | Bangladesh | 0 | Asian |
| HG03928.SG | Bangladesh | 0 | Asian |
| HG03931.SG | Bangladesh | 0 | Asian |
| HG03934.SG | Bangladesh | 0 | Asian |
| HG03937.SG | Bangladesh | 0 | Asian |
| HG03940.SG | Bangladesh | 0 | Asian |
| HG03941.SG | Bangladesh | 0 | Asian |

| | | | |
|---|---|---|---|
| HG03943.SG | Sri Lanka | 0 | Asian |
| HG03944.SG | Sri Lanka | 0 | Asian |
| HG03945.SG | Sri Lanka | 0 | Asian |
| HG03947.SG | Sri Lanka | 0 | Asian |
| HG03948.SG | Sri Lanka | 0 | Asian |
| HG03949.SG | Sri Lanka | 0 | Asian |
| HG03950.SG | Sri Lanka | 0 | Asian |
| HG03951.SG | Sri Lanka | 0 | Asian |
| HG03953.SG | Sri Lanka | 0 | Asian |
| HG03955.SG | Sri Lanka | 0 | Asian |
| HG03960.SG | India | 0 | Asian |
| HG03963.SG | India | 0 | Asian |
| HG03965.SG | India | 0 | Asian |
| HG03967.SG | India | 0 | Asian |
| HG03968.SG | India | 0 | Asian |
| HG03969.SG | India | 0 | Asian |
| HG03971.SG | India | 0 | Asian |
| HG03973.SG | India | 0 | Asian |
| HG03974.SG | India | 0 | Asian |
| HG03976.SG | India | 0 | Asian |
| HG03977.SG | India | 0 | Asian |
| HG03978.SG | India | 0 | Asian |
| HG03985.SG | Sri Lanka | 0 | Asian |
| HG03986.SG | Sri Lanka | 0 | Asian |
| HG03989.SG | Sri Lanka | 0 | Asian |
| HG03990.SG | Sri Lanka | 0 | Asian |
| HG03991.SG | Sri Lanka | 0 | Asian |
| HG03995.SG | Sri Lanka | 0 | Asian |
| HG03998.SG | Sri Lanka | 0 | Asian |

| | | | |
|---|---|---|---|
| HG03999.SG | Sri Lanka | 0 | Asian |
| HG04001.SG | India | 0 | Asian |
| HG04002.SG | India | 0 | Asian |
| HG04003.SG | Sri Lanka | 0 | Asian |
| HG04006.SG | Sri Lanka | 0 | Asian |
| HG04014.SG | India | 0 | Asian |
| HG04015.SG | India | 0 | Asian |
| HG04017.SG | India | 0 | Asian |
| HG04018.SG | India | 0 | Asian |
| HG04019.SG | India | 0 | Asian |
| HG04020.SG | India | 0 | Asian |
| HG04022.SG | India | 0 | Asian |
| HG04023.SG | India | 0 | Asian |
| HG04025.SG | India | 0 | Asian |
| HG04026.SG | India | 0 | Asian |
| HG04029.SG | Sri Lanka | 0 | Asian |
| HG04033.SG | Sri Lanka | 0 | Asian |
| HG04035.SG | Sri Lanka | 0 | Asian |
| HG04038.SG | Sri Lanka | 0 | Asian |
| HG04039.SG | Sri Lanka | 0 | Asian |
| HG04042.SG | Sri Lanka | 0 | Asian |
| HG04047.SG | Sri Lanka | 0 | Asian |
| HG04054.SG | India | 0 | Asian |
| HG04056.SG | India | 0 | Asian |
| HG04059.SG | India | 0 | Asian |
| HG04060.SG | India | 0 | Asian |
| HG04061.SG | India | 0 | Asian |
| HG04062.SG | India | 0 | Asian |
| HG04063.SG | India | 0 | Asian |

| | | | |
|---|---|---|---|
| HG04070.SG | India | 0 | Asian |
| HG04075.SG | Sri Lanka | 0 | Asian |
| HG04076.SG | India | 0 | Asian |
| HG04080.SG | India | 0 | Asian |
| HG04090.SG | India | 0 | Asian |
| HG04093.SG | India | 0 | Asian |
| HG04094.SG | India | 0 | Asian |
| HG04096.SG | India | 0 | Asian |
| HG04098.SG | India | 0 | Asian |
| HG04099.SG | Sri Lanka | 0 | Asian |
| HG04100.SG | Sri Lanka | 0 | Asian |
| HG04106.SG | Sri Lanka | 0 | Asian |
| HG04107.SG | Sri Lanka | 0 | Asian |
| HG04118.SG | India | 0 | Asian |
| HG04131.SG | Bangladesh | 0 | Asian |
| HG04134.SG | Bangladesh | 0 | Asian |
| HG04140.SG | Bangladesh | 0 | Asian |
| HG04141.SG | Bangladesh | 0 | Asian |
| HG04144.SG | Bangladesh | 0 | Asian |
| HG04146.SG | Bangladesh | 0 | Asian |
| HG04152.SG | Bangladesh | 0 | Asian |
| HG04153.SG | Bangladesh | 0 | Asian |
| HG04155.SG | Bangladesh | 0 | Asian |
| HG04156.SG | Bangladesh | 0 | Asian |
| HG04158.SG | Bangladesh | 0 | Asian |
| HG04159.SG | Bangladesh | 0 | Asian |
| HG04161.SG | Bangladesh | 0 | Asian |
| HG04162.SG | Bangladesh | 0 | Asian |
| HG04164.SG | Bangladesh | 0 | Asian |

| | | | |
|---|---|---|---|
| HG04171.SG | Bangladesh | 0 | Asian |
| HG04173.SG | Bangladesh | 0 | Asian |
| HG04176.SG | Bangladesh | 0 | Asian |
| HG04177.SG | Bangladesh | 0 | Asian |
| HG04180.SG | Bangladesh | 0 | Asian |
| HG04182.SG | Bangladesh | 0 | Asian |
| HG04183.SG | Bangladesh | 0 | Asian |
| HG04185.SG | Bangladesh | 0 | Asian |
| HG04186.SG | Bangladesh | 0 | Asian |
| HG04188.SG | Bangladesh | 0 | Asian |
| HG04189.SG | Bangladesh | 0 | Asian |
| HG04194.SG | Bangladesh | 0 | Asian |
| HG04195.SG | Bangladesh | 0 | Asian |
| HG04198.SG | India | 0 | Asian |
| HG04200.SG | India | 0 | Asian |
| HG04202.SG | India | 0 | Asian |
| HG04206.SG | India | 0 | Asian |
| HG04209.SG | India | 0 | Asian |
| HG04210.SG | Sri Lanka | 0 | Asian |
| HG04211.SG | India | 0 | Asian |
| HG04212.SG | India | 0 | Asian |
| HG04214.SG | India | 0 | Asian |
| HG04216.SG | India | 0 | Asian |
| HG04219.SG | India | 0 | Asian |
| HG04222.SG | India | 0 | Asian |
| HG04225.SG | India | 0 | Asian |
| HG04227.SG | Sri Lanka | 0 | Asian |
| HG04229.SG | Sri Lanka | 0 | Asian |
| HG04235.SG | India | 0 | Asian |

| | | | |
|---|---|---|---|
| HG04238.SG | India | 0 | Asian |
| HG04239.SG | India | 0 | Asian |
| NA18525.SG | China | 0 | Asian |
| NA18526.SG | China | 0 | Asian |
| NA18528.SG | China | 0 | Asian |
| NA18530.SG | China | 0 | Asian |
| NA18531.SG | China | 0 | Asian |
| NA18532.SG | China | 0 | Asian |
| NA18533.SG | China | 0 | Asian |
| NA18534.SG | China | 0 | Asian |
| NA18535.SG | China | 0 | Asian |
| NA18536.SG | China | 0 | Asian |
| NA18537.SG | China | 0 | Asian |
| NA18538.SG | China | 0 | Asian |
| NA18539.SG | China | 0 | Asian |
| NA18541.SG | China | 0 | Asian |
| NA18542.SG | China | 0 | Asian |
| NA18543.SG | China | 0 | Asian |
| NA18544.SG | China | 0 | Asian |
| NA18545.SG | China | 0 | Asian |
| NA18546.SG | China | 0 | Asian |
| NA18547.SG | China | 0 | Asian |
| NA18548.SG | China | 0 | Asian |
| NA18549.SG | China | 0 | Asian |
| NA18550.SG | China | 0 | Asian |
| NA18552.SG | China | 0 | Asian |
| NA18553.SG | China | 0 | Asian |
| NA18555.SG | China | 0 | Asian |
| NA18557.SG | China | 0 | Asian |

| | | | |
|---|---|---|---|
| NA18558.SG | China | 0 | Asian |
| NA18559.SG | China | 0 | Asian |
| NA18560.SG | China | 0 | Asian |
| NA18561.SG | China | 0 | Asian |
| NA18562.SG | China | 0 | Asian |
| NA18563.SG | China | 0 | Asian |
| NA18564.SG | China | 0 | Asian |
| NA18565.SG | China | 0 | Asian |
| NA18566.SG | China | 0 | Asian |
| NA18567.SG | China | 0 | Asian |
| NA18570.SG | China | 0 | Asian |
| NA18571.SG | China | 0 | Asian |
| NA18572.SG | China | 0 | Asian |
| NA18573.SG | China | 0 | Asian |
| NA18574.SG | China | 0 | Asian |
| NA18577.SG | China | 0 | Asian |
| NA18579.SG | China | 0 | Asian |
| NA18582.SG | China | 0 | Asian |
| NA18591.SG | China | 0 | Asian |
| NA18592.SG | China | 0 | Asian |
| NA18593.SG | China | 0 | Asian |
| NA18595.SG | China | 0 | Asian |
| NA18596.SG | China | 0 | Asian |
| NA18597.SG | China | 0 | Asian |
| NA18599.SG | China | 0 | Asian |
| NA18602.SG | China | 0 | Asian |
| NA18603.SG | China | 0 | Asian |
| NA18605.SG | China | 0 | Asian |
| NA18606.SG | China | 0 | Asian |

| | | | |
|---|---|---|---|
| NA18608.SG | China | 0 | Asian |
| NA18609.SG | China | 0 | Asian |
| NA18610.SG | China | 0 | Asian |
| NA18611.SG | China | 0 | Asian |
| NA18612.SG | China | 0 | Asian |
| NA18613.SG | China | 0 | Asian |
| NA18614.SG | China | 0 | Asian |
| NA18615.SG | China | 0 | Asian |
| NA18616.SG | China | 0 | Asian |
| NA18617.SG | China | 0 | Asian |
| NA18618.SG | China | 0 | Asian |
| NA18619.SG | China | 0 | Asian |
| NA18620.SG | China | 0 | Asian |
| NA18621.SG | China | 0 | Asian |
| NA18622.SG | China | 0 | Asian |
| NA18623.SG | China | 0 | Asian |
| NA18624.SG | China | 0 | Asian |
| NA18625.SG | China | 0 | Asian |
| NA18626.SG | China | 0 | Asian |
| NA18627.SG | China | 0 | Asian |
| NA18628.SG | China | 0 | Asian |
| NA18629.SG | China | 0 | Asian |
| NA18630.SG | China | 0 | Asian |
| NA18631.SG | China | 0 | Asian |
| NA18632.SG | China | 0 | Asian |
| NA18633.SG | China | 0 | Asian |
| NA18634.SG | China | 0 | Asian |
| NA18635.SG | China | 0 | Asian |
| NA18636.SG | China | 0 | Asian |

| | | | |
|---|---|---|---|
| NA18637.SG | China | 0 | Asian |
| NA18638.SG | China | 0 | Asian |
| NA18639.SG | China | 0 | Asian |
| NA18640.SG | China | 0 | Asian |
| NA18641.SG | China | 0 | Asian |
| NA18642.SG | China | 0 | Asian |
| NA18643.SG | China | 0 | Asian |
| NA18644.SG | China | 0 | Asian |
| NA18645.SG | China | 0 | Asian |
| NA18646.SG | China | 0 | Asian |
| NA18647.SG | China | 0 | Asian |
| NA18648.SG | China | 0 | Asian |
| NA18740.SG | China | 0 | Asian |
| NA18745.SG | China | 0 | Asian |
| NA18747.SG | China | 0 | Asian |
| NA18748.SG | China | 0 | Asian |
| NA18749.SG | China | 0 | Asian |
| NA18757.SG | China | 0 | Asian |
| NA18939.SG | Japan | 0 | Asian |
| NA18940.SG | Japan | 0 | Asian |
| NA18941.SG | Japan | 0 | Asian |
| NA18942.SG | Japan | 0 | Asian |
| NA18943.SG | Japan | 0 | Asian |
| NA18944.SG | Japan | 0 | Asian |
| NA18945.SG | Japan | 0 | Asian |
| NA18946.SG | Japan | 0 | Asian |
| NA18947.SG | Japan | 0 | Asian |
| NA18948.SG | Japan | 0 | Asian |
| NA18949.SG | Japan | 0 | Asian |

| | | | |
|---|---|---|---|
| NA18950.SG | Japan | 0 | Asian |
| NA18951.SG | Japan | 0 | Asian |
| NA18952.SG | Japan | 0 | Asian |
| NA18953.SG | Japan | 0 | Asian |
| NA18954.SG | Japan | 0 | Asian |
| NA18956.SG | Japan | 0 | Asian |
| NA18957.SG | Japan | 0 | Asian |
| NA18959.SG | Japan | 0 | Asian |
| NA18960.SG | Japan | 0 | Asian |
| NA18961.SG | Japan | 0 | Asian |
| NA18962.SG | Japan | 0 | Asian |
| NA18963.SG | Japan | 0 | Asian |
| NA18964.SG | Japan | 0 | Asian |
| NA18965.SG | Japan | 0 | Asian |
| NA18966.SG | Japan | 0 | Asian |
| NA18967.SG | Japan | 0 | Asian |
| NA18968.SG | Japan | 0 | Asian |
| NA18969.SG | Japan | 0 | Asian |
| NA18970.SG | Japan | 0 | Asian |
| NA18971.SG | Japan | 0 | Asian |
| NA18972.SG | Japan | 0 | Asian |
| NA18973.SG | Japan | 0 | Asian |
| NA18974.SG | Japan | 0 | Asian |
| NA18975.SG | Japan | 0 | Asian |
| NA18976.SG | Japan | 0 | Asian |
| NA18977.SG | Japan | 0 | Asian |
| NA18978.SG | Japan | 0 | Asian |
| NA18979.SG | Japan | 0 | Asian |
| NA18980.SG | Japan | 0 | Asian |

| | | | |
|---|---|---|---|
| NA18981.SG | Japan | 0 | Asian |
| NA18982.SG | Japan | 0 | Asian |
| NA18983.SG | Japan | 0 | Asian |
| NA18984.SG | Japan | 0 | Asian |
| NA18985.SG | Japan | 0 | Asian |
| NA18986.SG | Japan | 0 | Asian |
| NA18987.SG | Japan | 0 | Asian |
| NA18988.SG | Japan | 0 | Asian |
| NA18989.SG | Japan | 0 | Asian |
| NA18990.SG | Japan | 0 | Asian |
| NA18991.SG | Japan | 0 | Asian |
| NA18992.SG | Japan | 0 | Asian |
| NA18993.SG | Japan | 0 | Asian |
| NA18994.SG | Japan | 0 | Asian |
| NA18995.SG | Japan | 0 | Asian |
| NA18997.SG | Japan | 0 | Asian |
| NA18998.SG | Japan | 0 | Asian |
| NA18999.SG | Japan | 0 | Asian |
| NA19000.SG | Japan | 0 | Asian |
| NA19001.SG | Japan | 0 | Asian |
| NA19002.SG | Japan | 0 | Asian |
| NA19003.SG | Japan | 0 | Asian |
| NA19004.SG | Japan | 0 | Asian |
| NA19005.SG | Japan | 0 | Asian |
| NA19006.SG | Japan | 0 | Asian |
| NA19007.SG | Japan | 0 | Asian |
| NA19009.SG | Japan | 0 | Asian |
| NA19010.SG | Japan | 0 | Asian |
| NA19011.SG | Japan | 0 | Asian |

| | | | |
|---|---|---|---|
| NA19012.SG | Japan | 0 | Asian |
| NA19054.SG | Japan | 0 | Asian |
| NA19055.SG | Japan | 0 | Asian |
| NA19056.SG | Japan | 0 | Asian |
| NA19057.SG | Japan | 0 | Asian |
| NA19058.SG | Japan | 0 | Asian |
| NA19059.SG | Japan | 0 | Asian |
| NA19060.SG | Japan | 0 | Asian |
| NA19062.SG | Japan | 0 | Asian |
| NA19063.SG | Japan | 0 | Asian |
| NA19064.SG | Japan | 0 | Asian |
| NA19065.SG | Japan | 0 | Asian |
| NA19066.SG | Japan | 0 | Asian |
| NA19067.SG | Japan | 0 | Asian |
| NA19068.SG | Japan | 0 | Asian |
| NA19070.SG | Japan | 0 | Asian |
| NA19072.SG | Japan | 0 | Asian |
| NA19074.SG | Japan | 0 | Asian |
| NA19075.SG | Japan | 0 | Asian |
| NA19076.SG | Japan | 0 | Asian |
| NA19077.SG | Japan | 0 | Asian |
| NA19078.SG | Japan | 0 | Asian |
| NA19079.SG | Japan | 0 | Asian |
| NA19080.SG | Japan | 0 | Asian |
| NA19081.SG | Japan | 0 | Asian |
| NA19082.SG | Japan | 0 | Asian |
| NA19083.SG | Japan | 0 | Asian |
| NA19084.SG | Japan | 0 | Asian |
| NA19085.SG | Japan | 0 | Asian |

| | | | |
|---|---|---|---|
| NA19086.SG | Japan | 0 | Asian |
| NA19087.SG | Japan | 0 | Asian |
| NA19088.SG | Japan | 0 | Asian |
| NA19089.SG | Japan | 0 | Asian |
| NA19090.SG | Japan | 0 | Asian |
| NA19091.SG | Japan | 0 | Asian |
| I4899 | Uzbekistan | 3400 | Asian |
| I4901 | Uzbekistan | 3900 | Asian |
| I1183_published_d | Israel | 5950 | Asian |
| Darra.I.Kur_d | Afghanistan | 4466 | Asian |
| I4160 | Tajikistan | 3475 | Asian |
| I4163 | Uzbekistan | 3458 | Asian |
| I4634 | Turkmenistan | 4998 | Asian |
| I4635 | Turkmenistan | 5450 | Asian |
| I4910 | Tajikistan | 5526 | Asian |
| I4790 | Kazakhstan | 3425 | Asian |
| I4791 | Kazakhstan | 3449 | Asian |
| I4794 | Kazakhstan | 3450 | Asian |
| I5761 | Kazakhstan | 3796 | Asian |
| mondal_BIR-08.SG | India | 0 | Asian |
| mondal_BIR-11.SG | India | 0 | Asian |
| mondal_BIR-12.SG | India | 0 | Asian |
| mondal_BIR-13.SG | India | 0 | Asian |
| mondal_BIR-14.SG | India | 0 | Asian |
| mondal_BIR-15.SG | India | 0 | Asian |
| mondal_BIR-16.SG | India | 0 | Asian |
| mondal_BIR-18.SG | India | 0 | Asian |
| mondal_BIR-22.SG | India | 0 | Asian |
| mondal_BR1-10.SG | India | 0 | Asian |

| | | | |
|---|---|---|---|
| mondal_BR1-12.SG | India | 0 | Asian |
| mondal_BR1-13.SG | India | 0 | Asian |
| mondal_BR1-16.SG | India | 0 | Asian |
| mondal_BR1-18.SG | India | 0 | Asian |
| mondal_BR1-1.SG | India | 0 | Asian |
| mondal_BR1-3.SG | India | 0 | Asian |
| mondal_BR1-4.SG | India | 0 | Asian |
| mondal_BR1-5.SG | India | 0 | Asian |
| mondal_BR1-6.SG | India | 0 | Asian |
| mondal_IL-01.SG | India | 0 | Asian |
| mondal_IL-04.SG | India | 0 | Asian |
| mondal_IL-07.SG | India | 0 | Asian |
| mondal_IL-08.SG | India | 0 | Asian |
| mondal_IL-09.SG | India | 0 | Asian |
| mondal_IL-10.SG | India | 0 | Asian |
| mondal_IL-11.SG | India | 0 | Asian |
| mondal_IL-12.SG | India | 0 | Asian |
| mondal_IL-15.SG | India | 0 | Asian |
| mondal_IL-16.SG | India | 0 | Asian |
| mondal_PRBB-1.SG | India | 0 | Asian |
| mondal_PRBB-2.SG | India | 0 | Asian |
| mondal_RAI-11.SG | India | 0 | Asian |
| mondal_RAI-12.SG | India | 0 | Asian |
| mondal_RAI-13.SG | India | 0 | Asian |
| mondal_RAI-15.SG | India | 0 | Asian |
| mondal_RAI-2.SG | India | 0 | Asian |
| mondal_RAI-3.SG | India | 0 | Asian |
| mondal_RAI-4.SG | India | 0 | Asian |
| mondal_RAI-5.SG | India | 0 | Asian |

| | | | |
|---|---|---|---|
| mondal_RAI-8.SG | India | 0 | Asian |
| mondal_RAI-9.SG | India | 0 | Asian |
| mondal_RIA-12.SG | India | 0 | Asian |
| mondal_RIA-14.SG | India | 0 | Asian |
| mondal_RIA-19.SG | India | 0 | Asian |
| mondal_RIA-20.SG | India | 0 | Asian |
| mondal_RIA-23.SG | India | 0 | Asian |
| mondal_RIA-24.SG | India | 0 | Asian |
| mondal_RIA-41.SG | India | 0 | Asian |
| mondal_RIA-42.SG | India | 0 | Asian |
| mondal_RIA-45.SG | India | 0 | Asian |
| mondal_RIA-63.SG | India | 0 | Asian |
| mondal_VV1-01.SG | India | 0 | Asian |
| mondal_VV1-03.SG | India | 0 | Asian |
| mondal_VV1-04.SG | India | 0 | Asian |
| mondal_VV1-05.SG | India | 0 | Asian |
| mondal_VV1-06.SG | India | 0 | Asian |
| mondal_VV1-07.SG | India | 0 | Asian |
| mondal_VV1-14.SG | India | 0 | Asian |
| mondal_VV1-16.SG | India | 0 | Asian |
| mondal_VV1-20.SG | India | 0 | Asian |
| mondal_JAR-27.SG | India | 0 | Asian |
| mondal_JAR-32.SG | India | 0 | Asian |
| mondal_JAR-54.SG | India | 0 | Asian |
| mondal_JAR-61.SG | India | 0 | Asian |
| mondal_ONG-1.SG | India | 0 | Asian |
| mondal_ONG-12.SG | India | 0 | Asian |
| mondal_ONG-14.SG | India | 0 | Asian |
| mondal_ONG-4.SG | India | 0 | Asian |

| | | | |
|---|---|---|---|
| mondal_ONG-8.SG | India | 0 | Asian |
| mondal_ONG-9.SG | India | 0 | Asian |
| I2726_all | Vietnam | 3850 | Asian |
| I5396 | Pakistan | 2809 | Asian |
| I5397 | Pakistan | 2829 | Asian |
| I5398 | Pakistan | 2850 | Asian |
| I5400 | Pakistan | 2824 | Asian |
| I5604 | Uzbekistan | 3745 | Asian |
| I5605 | Uzbekistan | 3400 | Asian |
| I5608 | Uzbekistan | 3900 | Asian |
| I6218 | Turkmenistan | 4000 | Asian |
| I6221 | Mongolia | 4996 | Asian |
| I6222 | Mongolia | 5009 | Asian |
| I6224 | Mongolia | 2225 | Asian |
| I6225 | Mongolia | 2225 | Asian |
| I6226 | Mongolia | 2220 | Asian |
| I6228 | Mongolia | 1919 | Asian |
| I6230 | Mongolia | 2131 | Asian |
| I6231 | Mongolia | 2218 | Asian |
| I6232 | Mongolia | 2239 | Asian |
| I6233 | Mongolia | 2225 | Asian |
| I6262 | Mongolia | 3270 | Asian |
| I6263 | Mongolia | 2221 | Asian |
| I6264 | Mongolia | 3275 | Asian |
| I4349 | Iran | 7747 | Asian |
| I3453 | Kazakhstan | 3100 | Asian |
| I6117 | Turkmenistan | 4000 | Asian |
| I6118 | Turkmenistan | 4000 | Asian |
| I6119 | Turkmenistan | 3975 | Asian |

| | | | |
|---|---|---|---|
| I6120 | Turkmenistan | 4000 | Asian |
| I6122 | Turkmenistan | 4000 | Asian |
| I6124 | Turkmenistan | 4042 | Asian |
| I6125 | Turkmenistan | 4000 | Asian |
| I6126 | Turkmenistan | 4000 | Asian |
| I6127 | Turkmenistan | 4000 | Asian |
| I6195 | Pakistan | 2911 | Asian |
| I6217 | Turkmenistan | 4139 | Asian |
| I6310 | Turkmenistan | 4000 | Asian |
| I6312 | Turkmenistan | 4000 | Asian |
| I6318 | Turkmenistan | 4000 | Asian |
| I6463 | Jordan | 3300 | Asian |
| I6548 | Pakistan | 2850 | Asian |
| I6565 | Jordan | 3300 | Asian |
| I6461 | Jordan | 3300 | Asian |
| I6462 | Jordan | 3300 | Asian |
| I6464 | Jordan | 3300 | Asian |
| I6545 | Pakistan | 2821 | Asian |
| I6546 | Pakistan | 2841 | Asian |
| I6547 | Pakistan | 2000 | Asian |
| I6549 | Pakistan | 2041 | Asian |
| I6550 | Pakistan | 1926 | Asian |
| I6551 | Pakistan | 2000 | Asian |
| I6552 | Pakistan | 2051 | Asian |
| I6553 | Pakistan | 2834 | Asian |
| I6554 | Pakistan | 2758 | Asian |
| I6555 | Pakistan | 2811 | Asian |
| I6556 | Pakistan | 2776 | Asian |
| I6557 | Pakistan | 2850 | Asian |

| | | | |
|---|---|---|---|
| I6564 | Jordan | 3300 | Asian |
| I6566 | Jordan | 3300 | Asian |
| I6567 | Jordan | 3300 | Asian |
| I6569 | Jordan | 3300 | Asian |
| I6570 | Jordan | 3300 | Asian |
| I6571 | Jordan | 3385 | Asian |
| I6572 | Jordan | 3300 | Asian |
| I3728 | Taiwan | 1496 | Asian |
| I3736 | Taiwan | 1550 | Asian |
| I4241 | Iran | 7904 | Asian |
| I4243 | Iran | 4312 | Asian |
| I6292 | Pakistan | 2781 | Asian |
| I6349 | Mongolia | 2781 | Asian |
| I6351 | Mongolia | 2885 | Asian |
| I6352 | Mongolia | 2949 | Asian |
| I6353 | Mongolia | 2902 | Asian |
| I6356 | Mongolia | 2250 | Asian |
| I6357 | Mongolia | 2700 | Asian |
| I6358 | Mongolia | 7167 | Asian |
| I6359 | Mongolia | 2700 | Asian |
| I6361 | Mongolia | 4483 | Asian |
| I6362 | Mongolia | 2921 | Asian |
| I6364 | Mongolia | 2927 | Asian |
| I6459 | Jordan | 3234 | Asian |
| I6460 | Jordan | 3300 | Asian |
| I3614 | Taiwan | 1550 | Asian |
| I3615 | Taiwan | 1833 | Asian |
| I3617 | Taiwan | 1644 | Asian |
| I3618 | Taiwan | 1550 | Asian |

| | | | |
|---|---|---|---|
| I3619 | Taiwan | 1473 | Asian |
| I3620 | Taiwan | 1857 | Asian |
| I3707 | Jordan | 3286 | Asian |
| I3727 | Taiwan | 1550 | Asian |
| I3732 | Taiwan | 1235 | Asian |
| I3985 | Jordan | 3280 | Asian |
| I3986 | Jordan | 3300 | Asian |
| I3987 | Jordan | 3309 | Asian |
| I1701_published | Jordan | 9600 | Asian |
| I2870 | India | 128 | Asian |
| I3616 | Taiwan | 1641 | Asian |
| I3705 | Jordan | 3344 | Asian |
| I3706 | Jordan | 3304 | Asian |
| I3731 | Taiwan | 1550 | Asian |
| I3734 | Taiwan | 1575 | Asian |
| I3735 | Taiwan | 1532 | Asian |
| I1069_published | Israel | 12700 | Asian |
| I1414_published | Jordan | 10050 | Asian |
| I1415_published | Jordan | 9827 | Asian |
| I1685_published | Israel | 12700 | Asian |
| I1700_published | Jordan | 10050 | Asian |
| I1687_published | Israel | 13293 | Asian |
| I1690_published | Israel | 12700 | Asian |
| I2337 | Iran | 5522 | Asian |
| I2868 | India | 1011 | Asian |
| I2328 | Iran | 7900 | Asian |
| I2872 | India | 1099 | Asian |
| I2323 | Iran | 7910 | Asian |
| I2871 | India | 1099 | Asian |

| | | | |
|---|---|---|---|
| I2869 | India | 143 | Asian |
| I2327 | Iran | 3050 | Asian |
| I3832 | Israel | 3300 | Asian |
| I6669 | Turkmenistan | 4924 | Asian |
| I6671 | Turkmenistan | 4550 | Asian |
| I3365 | Turkmenistan | 4050 | Asian |
| I3703 | Jordan | 3300 | Asian |
| I6667 | Turkmenistan | 3400 | Asian |
| I6668 | Turkmenistan | 3250 | Asian |
| I6674 | Turkmenistan | 4148 | Asian |
| I6675 | Turkmenistan | 3250 | Asian |
| ERS1790729.SG | Lebanon | 3750 | Asian |
| ERS1790730.SG | Lebanon | 3650 | Asian |
| ERS1790731.SG | Lebanon | 3750 | Asian |
| ERS1790732.SG | Lebanon | 3650 | Asian |
| ERS1790733.SG | Lebanon | 3775 | Asian |
| I6932 | Israel | 3725 | Asian |
| I6925 | Israel | 3725 | Asian |
| I7026 | Mongolia | 2222 | Asian |
| I6924 | Israel | 3725 | Asian |
| I6928 | Israel | 3725 | Asian |
| I7002 | Israel | 3725 | Asian |
| I6922 | Israel | 3725 | Asian |
| I6947 | India | 131 | Asian |
| I7028 | Mongolia | 2232 | Asian |
| I6945 | India | 1178 | Asian |
| I6923 | Israel | 3725 | Asian |
| I7003 | Israel | 3725 | Asian |
| I6943 | India | 1228 | Asian |

| | | | |
|---|---|---|---|
| I6944 | India | 1125 | Asian |
| I6946 | India | 1109 | Asian |
| I7036 | India | 1023 | Asian |
| I6938 | India | 1152 | Asian |
| I6940 | India | 199 | Asian |
| I7035 | India | 1015 | Asian |
| I6941 | India | 1007 | Asian |
| I6942 | India | 1118 | Asian |
| I6939 | India | 128 | Asian |
| I6198 | Pakistan | 2950 | Asian |
| I6934 | India | 1109 | Asian |
| I6935 | India | 143 | Asian |
| I6937 | India | 188 | Asian |
| I6893 | Pakistan | 2225 | Asian |
| I7060 | Kazakhstan | 3439 | Asian |
| I6888 | Pakistan | 2747 | Asian |
| I6901 | Pakistan | 2950 | Asian |
| I6897 | Pakistan | 2950 | Asian |
| I6936 | India | 127 | Asian |
| I7029 | Mongolia | 2225 | Asian |
| I6900 | Pakistan | 3216 | Asian |
| I6896 | Pakistan | 2227 | Asian |
| I7023 | Mongolia | 2226 | Asian |
| I6890 | Pakistan | 853 | Asian |
| I6899 | Pakistan | 2811 | Asian |
| I7030 | Mongolia | 2244 | Asian |
| I6891 | Pakistan | 2242 | Asian |
| I7021 | Mongolia | 7050 | Asian |
| I7032 | Mongolia | 2849 | Asian |

| | | | |
|---|---|---|---|
| I7033 | Mongolia | 3066 | Asian |
| I7024 | Mongolia | 2244 | Asian |
| I7027 | Mongolia | 2278 | Asian |
| I6889 | Pakistan | 605 | Asian |
| I7022 | Mongolia | 2244 | Asian |
| I7039 | Mongolia | 3066 | Asian |
| I1805 | Pakistan | 382 | Asian |
| I1808 | Pakistan | 1121 | Asian |
| I1944 | Iran | 9800 | Asian |
| I1945 | Iran | 9800 | Asian |
| I1946 | Iran | 10000 | Asian |
| I1947 | Iran | 9992 | Asian |
| I1949 | Iran | 10042 | Asian |
| I1954 | Iran | 10162 | Asian |
| I1992 | Pakistan | 3025 | Asian |
| I1994 | Pakistan | 2904 | Asian |
| I2512 | Iran | 4838 | Asian |
| I2918 | Iran | 5566 | Asian |
| I1951 | Iran | 9848 | Asian |
| I1952 | Iran | 9952 | Asian |
| I4568 | Kazakhstan | 3631 | Asian |
| I4789 | Kazakhstan | 3487 | Asian |
| I6894 | Pakistan | 2250 | Asian |
| I1985 | Pakistan | 2939 | Asian |
| I3262 | Pakistan | 2844 | Asian |
| I1154 | Israel | 5950 | Asian |
| I7527 | Iran | 9900 | Asian |
| I8970 | Thailand | 2450 | Asian |
| I8974 | Thailand | 3050 | Asian |

| | | | |
|---|---|---|---|
| I8977 | Thailand | 3050 | Asian |
| I8978 | Thailand | 3300 | Asian |
| I2948_all | Vietnam | 1947 | Asian |
| I2947 | Vietnam | 3850 | Asian |
| I2948 | Vietnam | 1947 | Asian |
| I4458 | Thailand | 2603 | Asian |
| I4458_all | Thailand | 2603 | Asian |
| I10000 | Pakistan | 2850 | Asian |
| I10001 | Pakistan | 2850 | Asian |
| I10092 | Israel | 3750 | Asian |
| I10093 | Israel | 3750 | Asian |
| I10096 | Israel | 3500 | Asian |
| I10097 | Israel | 3500 | Asian |
| I10099 | Israel | 3500 | Asian |
| I10100 | Israel | 3548 | Asian |
| I10101 | Israel | 3500 | Asian |
| I10104 | Israel | 3825 | Asian |
| I10106 | Israel | 3550 | Asian |
| I10110 | Kazakhstan | 3100 | Asian |
| I10111 | Kazakhstan | 3396 | Asian |
| I10112 | Kazakhstan | 3497 | Asian |
| I10140 | Kazakhstan | 3575 | Asian |
| I10264 | Israel | 3750 | Asian |
| I10265 | Israel | 3825 | Asian |
| I10266 | Israel | 3462 | Asian |
| I10267 | Israel | 3527 | Asian |
| I10268 | Israel | 3853 | Asian |
| I10269 | Israel | 3600 | Asian |
| I10361 | Israel | 3500 | Asian |

| | | | |
|---|---|---|---|
| I10973 | Vietnam | 3850 | Asian |
| I10974 | Pakistan | 2903 | Asian |
| I11456_published | Iran | 4500 | Asian |
| I11459_published | Iran | 4699 | Asian |
| I11460_published | Iran | 4600 | Asian |
| I11461 | Iran | 4600 | Asian |
| I11462 | Iran | 4844 | Asian |
| I11466_published | Iran | 4200 | Asian |
| I11468 | Iran | 4900 | Asian |
| I11469_published | Iran | 5000 | Asian |
| I11471_published | Iran | 5115 | Asian |
| I11472 | Iran | 4750 | Asian |
| I11474 | Iran | 4600 | Asian |
| I11476 | Iran | 4600 | Asian |
| I11478 | Iran | 4500 | Asian |
| I11479 | Iran | 4796 | Asian |
| I11480_published | Iran | 4800 | Asian |
| I11483 | Iran | 4942 | Asian |
| I11488_published | Iran | 4600 | Asian |
| I4785 | Kazakhstan | 2675 | Asian |
| I5762 | Kazakhstan | 3500 | Asian |
| I6670 | Turkmenistan | 4450 | Asian |
| I7177 | Israel | 3725 | Asian |
| I7714 | Pakistan | 1944 | Asian |
| I7715 | Pakistan | 619 | Asian |
| I7716 | Pakistan | 728 | Asian |
| I7717 | Pakistan | 2283 | Asian |
| I7718 | Pakistan | 2250 | Asian |
| I7719 | Pakistan | 2250 | Asian |

| | | | |
|---|---|---|---|
| I7720 | Pakistan | 2250 | Asian |
| I7721 | Pakistan | 2250 | Asian |
| I7722 | Pakistan | 2250 | Asian |
| I7723 | Pakistan | 2250 | Asian |
| I8071 | Taiwan | 1550 | Asian |
| I8074 | Taiwan | 1550 | Asian |
| I8075 | Taiwan | 1459 | Asian |
| I8076 | Taiwan | 1550 | Asian |
| I8078 | Taiwan | 1575 | Asian |
| I8080 | Taiwan | 1550 | Asian |
| I8081 | Taiwan | 1409 | Asian |
| I8187 | Israel | 3675 | Asian |
| I8188 | Israel | 3675 | Asian |
| I8190 | Pakistan | 2847 | Asian |
| I8193 | Pakistan | 3128 | Asian |
| I8194 | Pakistan | 2950 | Asian |
| I8502 | Turkmenistan | 5013 | Asian |
| I8503 | Turkmenistan | 5050 | Asian |
| I8504 | Turkmenistan | 4963 | Asian |
| I8505 | Turkmenistan | 5050 | Asian |
| I8506 | Uzbekistan | 3614 | Asian |
| I8507 | Uzbekistan | 3950 | Asian |
| I8508 | Uzbekistan | 6356 | Asian |
| I8510 | Turkmenistan | 5163 | Asian |
| I8524 | Turkmenistan | 5050 | Asian |
| I8525 | Turkmenistan | 4943 | Asian |
| I8526 | Turkmenistan | 5050 | Asian |
| I8527 | Turkmenistan | 5050 | Asian |
| I8528 | Turkmenistan | 5050 | Asian |

| | | | |
|---|---|---|---|
| I8529 | Turkmenistan | 5050 | Asian |
| I8530 | Turkmenistan | 5050 | Asian |
| I8531 | Turkmenistan | 5050 | Asian |
| I8532 | Turkmenistan | 5050 | Asian |
| I8534 | Turkmenistan | 4862 | Asian |
| I8724 | Iran | 4900 | Asian |
| I8725 | Iran | 4862 | Asian |
| I8726 | Iran | 5000 | Asian |
| I8728_published | Iran | 4500 | Asian |
| I8997 | Pakistan | 2850 | Asian |
| I8998 | Pakistan | 2850 | Asian |
| I8999 | Pakistan | 2813 | Asian |
| I1799 | Pakistan | 3003 | Asian |
| I3260 | Pakistan | 2908 | Asian |
| I3261 | Pakistan | 2950 | Asian |
| I3621 | Taiwan | 1595 | Asian |
| I4290 | Tajikistan | 5450 | Asian |
| I4780 | Kazakhstan | 3649 | Asian |
| I6194 | Pakistan | 2950 | Asian |
| I6197 | Pakistan | 2950 | Asian |
| I6347 | Mongolia | 3295 | Asian |
| I6363 | Mongolia | 3095 | Asian |
| I6365 | Mongolia | 2744 | Asian |
| I6367 | Mongolia | 3116 | Asian |
| I6369 | Mongolia | 2554 | Asian |
| I7238 | Myanmar | 2950 | Asian |
| I8218 | Pakistan | 1949 | Asian |
| I8220 | Pakistan | 2566 | Asian |
| I8245 | Pakistan | 2197 | Asian |

| | | | |
|---|---|---|---|
| I8246_published | Pakistan | 2710 | Asian |
| DA100.SG | Kyrgyzstan | 1615 | Asian |
| DA101.SG | Kyrgyzstan | 1664 | Asian |
| DA104.SG | Kyrgyzstan | 1600 | Asian |
| DA105.SG | Kyrgyzstan | 1739 | Asian |
| DA106.SG | Kyrgyzstan | 735 | Asian |
| DA10.SG | Kazakhstan | 2550 | Asian |
| DA116.SG | Kyrgyzstan | 761 | Asian |
| DA117.SG | Kyrgyzstan | 139 | Asian |
| DA118.SG | Kyrgyzstan | 851 | Asian |
| DA11.SG | Kazakhstan | 2578 | Asian |
| DA121.SG | Kazakhstan | 1588 | Asian |
| DA123.SG | Kazakhstan | 1700 | Asian |
| DA124.SG | Kazakhstan | 1245 | Asian |
| DA125.SG | Kazakhstan | 1688 | Asian |
| DA126.SG | Kazakhstan | 991 | Asian |
| DA127.SG | Kazakhstan | 1616 | Asian |
| DA128.SG | Kazakhstan | 692 | Asian |
| DA129.SG | Kazakhstan | 2333 | Asian |
| DA130.SG | Kazakhstan | 2221 | Asian |
| DA13.SG | Kazakhstan | 2568 | Asian |
| DA14.SG | Kazakhstan | 2557 | Asian |
| DA15.SG | Kazakhstan | 2557 | Asian |
| DA16.SG | Kazakhstan | 2553 | Asian |
| DA177.SG | Kazakhstan | 1600 | Asian |
| DA179.SG | Kazakhstan | 850 | Asian |
| DA17.SG | Kazakhstan | 2704 | Asian |
| DA18.SG | Kazakhstan | 2549 | Asian |
| DA19.SG | Kazakhstan | 2631 | Asian |

| | | | |
|---|---|---|---|
| DA201.SG | Kazakhstan | 2069 | Asian |
| DA202.SG | Kazakhstan | 2476 | Asian |
| DA203.SG | Kazakhstan | 1000 | Asian |
| DA204.SG | Kazakhstan | 1017 | Asian |
| DA205.SG | Kazakhstan | 1000 | Asian |
| DA206.SG | Kazakhstan | 1697 | Asian |
| DA207.SG | Kazakhstan | 200 | Asian |
| DA208.SG | Kazakhstan | 200 | Asian |
| DA20.SG | Kazakhstan | 2224 | Asian |
| DA220.SG | Kazakhstan | 2201 | Asian |
| DA221.SG | Kazakhstan | 2563 | Asian |
| DA222.SG | Kazakhstan | 1100 | Asian |
| DA223.SG | Kazakhstan | 2311 | Asian |
| DA224.SG | Kazakhstan | 1627 | Asian |
| DA226.SG | Kazakhstan | 1700 | Asian |
| DA227.SG | Kazakhstan | 2150 | Asian |
| DA228.SG | Kazakhstan | 1350 | Asian |
| DA229.SG | Kazakhstan | 1765 | Asian |
| DA230.SG | Kazakhstan | 883 | Asian |
| DA231.SG | Kazakhstan | 3439 | Asian |
| DA23.SG | Kazakhstan | 840 | Asian |
| DA26.SG | Kazakhstan | 2300 | Asian |
| DA27.SG | Kazakhstan | 1508 | Asian |
| DA28.SG | Kazakhstan | 550 | Asian |
| DA29.SG | Kazakhstan | 550 | Asian |
| DA30.SG | Kazakhstan | 1948 | Asian |
| DA385.SG | Kyrgyzstan | 1458 | Asian |
| DA38.SG | Mongolia | 2110 | Asian |
| DA41.SG | Mongolia | 2148 | Asian |

| | | | |
|---|---|---|---|
| DA43.SG | Mongolia | 2100 | Asian |
| DA45.SG | Mongolia | 2046 | Asian |
| DA47.SG | Kyrgyzstan | 2063 | Asian |
| DA48.SG | Kyrgyzstan | 2200 | Asian |
| DA49.SG | Kyrgyzstan | 2209 | Asian |
| DA50.SG | Kyrgyzstan | 2300 | Asian |
| DA51.SG | Kyrgyzstan | 2229 | Asian |
| DA52.SG | Kyrgyzstan | 1472 | Asian |
| DA53.SG | Kyrgyzstan | 2372 | Asian |
| DA54.SG | Kyrgyzstan | 1594 | Asian |
| DA55.SG | Kyrgyzstan | 1976 | Asian |
| DA56.SG | Kyrgyzstan | 2043 | Asian |
| DA57.SG | Kyrgyzstan | 2065 | Asian |
| DA58.SG | Kyrgyzstan | 2060 | Asian |
| DA59.SG | Kyrgyzstan | 2149 | Asian |
| DA65.SG | Kyrgyzstan | 1494 | Asian |
| DA66.SG | Kyrgyzstan | 1435 | Asian |
| DA68.SG | Kyrgyzstan | 1354 | Asian |
| DA69.SG | Kyrgyzstan | 1509 | Asian |
| DA70.SG | Kyrgyzstan | 1802 | Asian |
| DA71.SG | Kyrgyzstan | 1627 | Asian |
| DA72.SG | Kyrgyzstan | 1575 | Asian |
| DA73.SG | Kyrgyzstan | 1618 | Asian |
| DA74.SG | Kyrgyzstan | 1492 | Asian |
| DA80.SG | Kyrgyzstan | 1600 | Asian |
| DA81.SG | Kyrgyzstan | 1656 | Asian |
| DA82.SG | Kyrgyzstan | 1779 | Asian |
| DA85.SG | Kyrgyzstan | 1666 | Asian |
| DA86.SG | Kyrgyzstan | 1461 | Asian |

| | | | |
|---|---|---|---|
| DA87.SG | Kazakhstan | 1259 | Asian |
| DA89.SG | Kazakhstan | 1231 | Asian |
| DA92.SG | Kazakhstan | 1899 | Asian |
| DA93.SG | Kazakhstan | 1126 | Asian |
| DA94.SG | Kazakhstan | 1000 | Asian |
| DA95.SG | Kazakhstan | 1596 | Asian |
| DA96.SG | Kyrgyzstan | 1608 | Asian |
| DA98.SG | Kyrgyzstan | 1802 | Asian |
| DA99.SG | Kyrgyzstan | 757 | Asian |
| BOT14.SG | Kazakhstan | 5337 | Asian |
| BOT15.SG | Kazakhstan | 5143 | Asian |
| BOT2016.SG | Kazakhstan | 5396 | Asian |
| DA379_final.SG | Turkmenistan | 5160 | Asian |
| DA380_final.SG | Turkmenistan | 5173 | Asian |
| DA381_final.SG | Turkmenistan | 5174 | Asian |
| DA382_final.SG | Turkmenistan | 2802 | Asian |
| DA383_final.SG | Turkmenistan | 5150 | Asian |
| Dungan1.SG | Kyrgyzstan | 0 | Asian |
| Dungan2.SG | Kyrgyzstan | 0 | Asian |
| EBA1.SG | Kazakhstan | 4108 | Asian |
| EBA2.SG | Kazakhstan | 4480 | Asian |
| Hazaras1.SG | Pakistan | 0 | Asian |
| Hazaras2.SG | Pakistan | 0 | Asian |
| Hazaras3.SG | Pakistan | 0 | Asian |
| Karakalpaks1.SG | Uzbekistan | 0 | Asian |
| Karakalpaks2.SG | Uzbekistan | 0 | Asian |
| Karakalpaks3.SG | Uzbekistan | 0 | Asian |
| Kazakhs2.SG | Kazakhstan | 0 | Asian |
| Kazakhs3.SG | Kazakhstan | 0 | Asian |

| | | | |
|---|---|---|---|
| Kazkahs1.SG | Kazakhstan | 0 | Asian |
| Kyrgyz1.SG | Kyrgyzstan | 0 | Asian |
| Kyrgyz2.SG | Kyrgyzstan | 0 | Asian |
| Tajiks1.SG | Tajikistan | 0 | Asian |
| Tajiks2.SG | Tajikistan | 0 | Asian |
| Tajiks3.SG | Tajikistan | 0 | Asian |
| Turkmens1.SG | Turkmenistan | 0 | Asian |
| Turkmens2.SG | Turkmenistan | 0 | Asian |
| Uyghurs1.SG | China | 0 | Asian |
| Uyghurs2.SG | China | 0 | Asian |
| Uyghurs3.SG | China | 0 | Asian |
| Uzbeks1.SG | Uzbekistan | 0 | Asian |
| Uzbeks2.SG | Uzbekistan | 0 | Asian |
| Uzbeks3.SG | Uzbekistan | 0 | Asian |
| Damgaard2018Yamnaya.SG | Kazakhstan | 4890 | Asian |
| HGDP01189.SDG | China | 0 | Asian |
| HGDP01216.SDG | China | 0 | Asian |
| HGDP01244.SDG | China | 0 | Asian |
| HGDP01243.SDG | China | 0 | Asian |
| HGDP01299.SDG | China | 0 | Asian |
| HGDP00135.SDG | Pakistan | 0 | Asian |
| HGDP00214.SDG | Pakistan | 0 | Asian |
| HGDP01096.SDG | China | 0 | Asian |
| HGDP01298.SDG | China | 0 | Asian |
| HGDP00715.SDG | Cambodia | 0 | Asian |
| HGDP00711.SDG | Cambodia | 0 | Asian |
| HGDP00103.SDG | Pakistan | 0 | Asian |
| HGDP00359.SDG | Pakistan | 0 | Asian |
| HGDP00136.SDG | Pakistan | 0 | Asian |

| | | | |
|---|---|---|---|
| HGDP00105.SDG | Pakistan | 0 | Asian |
| HGDP00127.SDG | Pakistan | 0 | Asian |
| HGDP00580.SDG | Israel | 0 | Asian |
| HGDP00130.SDG | Pakistan | 0 | Asian |
| HGDP00167.SDG | Pakistan | 0 | Asian |
| HGDP00576.SDG | Israel | 0 | Asian |
| HGDP00005.SDG | Pakistan | 0 | Asian |
| HGDP00163.SDG | Pakistan | 0 | Asian |
| HGDP00346.SDG | Pakistan | 0 | Asian |
| HGDP00609.SDG | Israel | 0 | Asian |
| HGDP00608.SDG | Israel | 0 | Asian |
| HGDP00714.SDG | Cambodia | 0 | Asian |
| HGDP00057.SDG | Pakistan | 0 | Asian |
| HGDP00213.SDG | Pakistan | 0 | Asian |
| HGDP00165.SDG | Pakistan | 0 | Asian |
| HGDP01300.SDG | China | 0 | Asian |
| HGDP00279.SDG | Pakistan | 0 | Asian |
| HGDP00056.SDG | Pakistan | 0 | Asian |
| HGDP00288.SDG | Pakistan | 0 | Asian |
| HGDP00562.SDG | Israel | 0 | Asian |
| HGDP00341.SDG | Pakistan | 0 | Asian |
| HGDP00723.SDG | Israel | 0 | Asian |
| HGDP00610.SDG | Israel | 0 | Asian |
| HGDP00001.SDG | Pakistan | 0 | Asian |
| HGDP00218.SDG | Pakistan | 0 | Asian |
| HGDP00062.SDG | Pakistan | 0 | Asian |
| HGDP01192.SDG | China | 0 | Asian |
| HGDP00009.SDG | Pakistan | 0 | Asian |
| HGDP01225.SDG | China | 0 | Asian |

| | | | |
|---|---|---|---|
| HGDP00724.SDG | Israel | 0 | Asian |
| HGDP00726.SDG | Israel | 0 | Asian |
| HGDP01190.SDG | China | 0 | Asian |
| HGDP00118.SDG | Pakistan | 0 | Asian |
| HGDP00781.SDG | China | 0 | Asian |
| HGDP00782.SDG | China | 0 | Asian |
| HGDP01245.SDG | China | 0 | Asian |
| HGDP00820.SDG | China | 0 | Asian |
| HGDP01214.SDG | China | 0 | Asian |
| HGDP00319.SDG | Pakistan | 0 | Asian |
| HGDP01186.SDG | China | 0 | Asian |
| HGDP01310.SDG | China | 0 | Asian |
| HGDP00821.SDG | China | 0 | Asian |
| HGDP00777.SDG | China | 0 | Asian |
| HGDP01327.SDG | China | 0 | Asian |
| HGDP01099.SDG | China | 0 | Asian |
| HGDP00972.SDG | China | 0 | Asian |
| HGDP01313.SDG | China | 0 | Asian |
| HGDP00813.SDG | China | 0 | Asian |
| HGDP00814.SDG | China | 0 | Asian |
| HGDP00971.SDG | China | 0 | Asian |
| HGDP01296.SDG | China | 0 | Asian |
| HGDP01330.SDG | China | 0 | Asian |
| HGDP00812.SDG | China | 0 | Asian |
| HGDP01213.SDG | China | 0 | Asian |
| HGDP01340.SDG | China | 0 | Asian |
| HGDP00811.SDG | China | 0 | Asian |
| HGDP01021.SDG | China | 0 | Asian |
| HGDP01289.SDG | China | 0 | Asian |

| | | | |
|---|---|---|---|
| HGDP01023.SDG | China | 0 | Asian |
| HGDP00776.SDG | China | 0 | Asian |
| HGDP00774.SDG | China | 0 | Asian |
| HGDP00815.SDG | China | 0 | Asian |
| HGDP00779.SDG | China | 0 | Asian |
| HGDP00818.SDG | China | 0 | Asian |
| HGDP00784.SDG | China | 0 | Asian |
| HGDP01318.SDG | China | 0 | Asian |
| HGDP01100.SDG | China | 0 | Asian |
| HGDP01290.SDG | China | 0 | Asian |
| HGDP00975.SDG | China | 0 | Asian |
| HGDP00974.SDG | China | 0 | Asian |
| HGDP01196.SDG | China | 0 | Asian |
| HGDP00786.SDG | China | 0 | Asian |
| HGDP01181.SDG | China | 0 | Asian |
| HGDP01354.SDG | China | 0 | Asian |
| HGDP01294.SDG | China | 0 | Asian |
| HGDP01352.SDG | China | 0 | Asian |
| HGDP01348.SDG | China | 0 | Asian |
| HGDP01322.SDG | China | 0 | Asian |
| HGDP00977.SDG | China | 0 | Asian |
| HGDP00780.SDG | China | 0 | Asian |
| HGDP01287.SDG | China | 0 | Asian |
| HGDP01292.SDG | China | 0 | Asian |
| HGDP00976.SDG | China | 0 | Asian |
| HGDP01224.SDG | China | 0 | Asian |
| HGDP01184.SDG | China | 0 | Asian |
| HGDP01334.SDG | China | 0 | Asian |
| HGDP01321.SDG | China | 0 | Asian |

| | | | |
|---|---|---|---|
| HGDP01101.SDG | China | 0 | Asian |
| HGDP01353.SDG | China | 0 | Asian |
| HGDP00817.SDG | China | 0 | Asian |
| HGDP01180.SDG | China | 0 | Asian |
| HGDP01195.SDG | China | 0 | Asian |
| HGDP01341.SDG | China | 0 | Asian |
| HG01804.SG | China | 0 | Asian |
| HGDP01311.SDG | China | 0 | Asian |
| HGDP01193.SDG | China | 0 | Asian |
| HGDP01104.SDG | China | 0 | Asian |
| HGDP01187.SDG | China | 0 | Asian |
| HGDP01347.SDG | China | 0 | Asian |
| HGDP01197.SDG | China | 0 | Asian |
| HGDP01349.SDG | China | 0 | Asian |
| HGDP01351.SDG | China | 0 | Asian |
| HGDP01328.SDG | China | 0 | Asian |
| HGDP01295.SDG | China | 0 | Asian |
| HGDP01288.SDG | China | 0 | Asian |
| HGDP01329.SDG | China | 0 | Asian |
| HGDP00747.SDG | Japan | 0 | Asian |
| HGDP01293.SDG | China | 0 | Asian |
| HGDP00973.SDG | China | 0 | Asian |
| HGDP00762.SDG | Japan | 0 | Asian |
| HGDP01185.SDG | China | 0 | Asian |
| HGDP01331.SDG | China | 0 | Asian |
| HGDP01103.SDG | China | 0 | Asian |
| HGDP01317.SDG | China | 0 | Asian |
| HGDP01346.SDG | China | 0 | Asian |
| HGDP01342.SDG | China | 0 | Asian |

| | | | |
|---|---|---|---|
| HGDP01238.SDG | China | 0 | Asian |
| HGDP01326.SDG | China | 0 | Asian |
| HGDP01356.SDG | China | 0 | Asian |
| HGDP01247.SDG | China | 0 | Asian |
| HGDP00766.SDG | Japan | 0 | Asian |
| HGDP01233.SDG | China | 0 | Asian |
| HGDP01207.SDG | China | 0 | Asian |
| HGDP01336.SDG | China | 0 | Asian |
| HGDP00819.SDG | China | 0 | Asian |
| HGDP00382.SDG | Pakistan | 0 | Asian |
| HGDP01332.SDG | China | 0 | Asian |
| HGDP01337.SDG | China | 0 | Asian |
| HGDP01222.SDG | China | 0 | Asian |
| HGDP01291.SDG | China | 0 | Asian |
| HGDP01183.SDG | China | 0 | Asian |
| HGDP00822.SDG | China | 0 | Asian |
| HGDP00755.SDG | Japan | 0 | Asian |
| HGDP01304.SDG | China | 0 | Asian |
| HGDP00716.SDG | Cambodia | 0 | Asian |
| HGDP01182.SDG | China | 0 | Asian |
| HGDP01230.SDG | China | 0 | Asian |
| HGDP01194.SDG | China | 0 | Asian |
| HGDP00376.SDG | Pakistan | 0 | Asian |
| HGDP00364.SDG | Pakistan | 0 | Asian |
| HGDP01234.SDG | China | 0 | Asian |
| HGDP00712.SDG | Cambodia | 0 | Asian |
| HGDP00754.SDG | Japan | 0 | Asian |
| HGDP01232.SDG | China | 0 | Asian |
| HGDP01102.SDG | China | 0 | Asian |

| | | | |
|---|---|---|---|
| HGDP01204.SDG | China | 0 | Asian |
| HGDP00100.SDG | Pakistan | 0 | Asian |
| HGDP01303.SDG | China | 0 | Asian |
| HGDP01206.SDG | China | 0 | Asian |
| HGDP00772.SDG | Japan | 0 | Asian |
| HGDP00099.SDG | Pakistan | 0 | Asian |
| HGDP00371.SDG | Pakistan | 0 | Asian |
| HGDP00758.SDG | Japan | 0 | Asian |
| HGDP00765.SDG | Japan | 0 | Asian |
| HGDP00750.SDG | Japan | 0 | Asian |
| HGDP01229.SDG | China | 0 | Asian |
| HGDP01237.SDG | China | 0 | Asian |
| HGDP00756.SDG | Japan | 0 | Asian |
| HGDP00751.SDG | Japan | 0 | Asian |
| HGDP00828.SDG | Japan | 0 | Asian |
| HGDP00108.SDG | Pakistan | 0 | Asian |
| HGDP00115.SDG | Pakistan | 0 | Asian |
| HGDP00790.SDG | Japan | 0 | Asian |
| HGDP00109.SDG | Pakistan | 0 | Asian |
| HGDP00356.SDG | Pakistan | 0 | Asian |
| HGDP00392.SDG | Pakistan | 0 | Asian |
| HGDP01221.SDG | China | 0 | Asian |
| HGDP01209.SDG | China | 0 | Asian |
| HGDP00719.SDG | Cambodia | 0 | Asian |
| HGDP00104.SDG | Pakistan | 0 | Asian |
| HGDP01249.SDG | China | 0 | Asian |
| HGDP01218.SDG | China | 0 | Asian |
| HGDP01251.SDG | China | 0 | Asian |
| HGDP00771.SDG | Japan | 0 | Asian |

| | | | |
|---|---|---|---|
| HGDP01220.SDG | China | 0 | Asian |
| HGDP01212.SDG | China | 0 | Asian |
| HGDP00761.SDG | Japan | 0 | Asian |
| HGDP01205.SDG | China | 0 | Asian |
| HGDP00351.SDG | Pakistan | 0 | Asian |
| HGDP00372.SDG | Pakistan | 0 | Asian |
| HGDP01241.SDG | China | 0 | Asian |
| HGDP00760.SDG | Japan | 0 | Asian |
| HGDP00769.SDG | Japan | 0 | Asian |
| HGDP00757.SDG | Japan | 0 | Asian |
| HGDP00721.SDG | Cambodia | 0 | Asian |
| HGDP00021.SDG | Pakistan | 0 | Asian |
| HGDP01231.SDG | China | 0 | Asian |
| HGDP00039.SDG | Pakistan | 0 | Asian |
| HGDP00764.SDG | Japan | 0 | Asian |
| HGDP01305.SDG | China | 0 | Asian |
| HGDP00060.SDG | Pakistan | 0 | Asian |
| HGDP00035.SDG | Pakistan | 0 | Asian |
| HGDP00791.SDG | Japan | 0 | Asian |
| HGDP00234.SDG | Pakistan | 0 | Asian |
| HGDP01227.SDG | China | 0 | Asian |
| HGDP01236.SDG | China | 0 | Asian |
| HGDP00110.SDG | Pakistan | 0 | Asian |
| HGDP00173.SDG | Pakistan | 0 | Asian |
| HGDP00106.SDG | Pakistan | 0 | Asian |
| HGDP00285.SDG | Pakistan | 0 | Asian |
| HGDP00047.SDG | Pakistan | 0 | Asian |
| HGDP00049.SDG | Pakistan | 0 | Asian |
| HGDP00031.SDG | Pakistan | 0 | Asian |

| | | | |
|---|---|---|---|
| HGDP00438.SDG | Pakistan | 0 | Asian |
| HGDP00199.SDG | Pakistan | 0 | Asian |
| HGDP00313.SDG | Pakistan | 0 | Asian |
| HGDP00066.SDG | Pakistan | 0 | Asian |
| HGDP00239.SDG | Pakistan | 0 | Asian |
| HGDP00013.SDG | Pakistan | 0 | Asian |
| HGDP00323.SDG | Pakistan | 0 | Asian |
| HGDP00070.SDG | Pakistan | 0 | Asian |
| HGDP00262.SDG | Pakistan | 0 | Asian |
| HGDP00224.SDG | Pakistan | 0 | Asian |
| HGDP00304.SDG | Pakistan | 0 | Asian |
| HGDP00098.SDG | Pakistan | 0 | Asian |
| HGDP00043.SDG | Pakistan | 0 | Asian |
| HGDP00181.SDG | Pakistan | 0 | Asian |
| HGDP00247.SDG | Pakistan | 0 | Asian |
| HGDP00445.SDG | Pakistan | 0 | Asian |
| HGDP00423.SDG | Pakistan | 0 | Asian |
| HGDP00189.SDG | Pakistan | 0 | Asian |
| HGDP00243.SDG | Pakistan | 0 | Asian |
| HGDP00033.SDG | Pakistan | 0 | Asian |
| HGDP00011.SDG | Pakistan | 0 | Asian |
| HGDP00210.SDG | Pakistan | 0 | Asian |
| HGDP00082.SDG | Pakistan | 0 | Asian |
| HGDP00258.SDG | Pakistan | 0 | Asian |
| HGDP00315.SDG | Pakistan | 0 | Asian |
| HGDP00052.SDG | Pakistan | 0 | Asian |
| HGDP00074.SDG | Pakistan | 0 | Asian |
| HGDP00185.SDG | Pakistan | 0 | Asian |
| HGDP00072.SDG | Pakistan | 0 | Asian |

| | | | |
|---|---|---|---|
| HGDP00192.SDG | Pakistan | 0 | Asian |
| HGDP00244.SDG | Pakistan | 0 | Asian |
| HGDP00277.SDG | Pakistan | 0 | Asian |
| HGDP00412.SDG | Pakistan | 0 | Asian |
| HGDP00092.SDG | Pakistan | 0 | Asian |
| HGDP00228.SDG | Pakistan | 0 | Asian |
| HGDP00264.SDG | Pakistan | 0 | Asian |
| HGDP00290.SDG | Pakistan | 0 | Asian |
| HGDP00330.SDG | Pakistan | 0 | Asian |
| HGDP00003.SDG | Pakistan | 0 | Asian |
| HGDP00417.SDG | Pakistan | 0 | Asian |
| HGDP00025.SDG | Pakistan | 0 | Asian |
| HGDP00139.SDG | Pakistan | 0 | Asian |
| HGDP00205.SDG | Pakistan | 0 | Asian |
| HGDP00183.SDG | Pakistan | 0 | Asian |
| HGDP00402.SDG | Pakistan | 0 | Asian |
| HGDP00169.SDG | Pakistan | 0 | Asian |
| HGDP00080.SDG | Pakistan | 0 | Asian |
| HGDP00096.SDG | Pakistan | 0 | Asian |
| HGDP00015.SDG | Pakistan | 0 | Asian |
| HGDP00141.SDG | Pakistan | 0 | Asian |
| HGDP00086.SDG | Pakistan | 0 | Asian |
| HGDP00298.SDG | Pakistan | 0 | Asian |
| HGDP00146.SDG | Pakistan | 0 | Asian |
| HGDP00251.SDG | Pakistan | 0 | Asian |
| HGDP00150.SDG | Pakistan | 0 | Asian |
| HGDP00226.SDG | Pakistan | 0 | Asian |
| HGDP00309.SDG | Pakistan | 0 | Asian |
| HGDP00153.SDG | Pakistan | 0 | Asian |

| | | | |
|---|---|---|---|
| HGDP00088.SDG | Pakistan | 0 | Asian |
| HGDP00145.SDG | Pakistan | 0 | Asian |
| HGDP00134.SDG | Pakistan | 0 | Asian |
| HGDP00137.SDG | Pakistan | 0 | Asian |
| HGDP00201.SDG | Pakistan | 0 | Asian |
| HGDP00154.SDG | Pakistan | 0 | Asian |
| HGDP00143.SDG | Pakistan | 0 | Asian |
| HGDP00177.SDG | Pakistan | 0 | Asian |
| HGDP00078.SDG | Pakistan | 0 | Asian |
| HGDP00149.SDG | Pakistan | 0 | Asian |
| HGDP00158.SDG | Pakistan | 0 | Asian |
| HGDP00094.SDG | Pakistan | 0 | Asian |
| HGDP00151.SDG | Pakistan | 0 | Asian |
| HGDP00133.SDG | Pakistan | 0 | Asian |
| HGDP00076.SDG | Pakistan | 0 | Asian |
| HGDP00140.SDG | Pakistan | 0 | Asian |
| HGDP00148.SDG | Pakistan | 0 | Asian |
| HGDP00155.SDG | Pakistan | 0 | Asian |
| HGDP00131.SDG | Pakistan | 0 | Asian |
| HGDP00144.SDG | Pakistan | 0 | Asian |
| HGDP00161.SDG | Pakistan | 0 | Asian |
| HGDP00561.SDG | Israel | 0 | Asian |
| HGDP00563.SDG | Israel | 0 | Asian |
| HGDP00572.SDG | Israel | 0 | Asian |
| HGDP00573.SDG | Israel | 0 | Asian |
| HGDP00583.SDG | Israel | 0 | Asian |
| HGDP00582.SDG | Israel | 0 | Asian |
| HGDP00579.SDG | Israel | 0 | Asian |
| HGDP00649.SDG | Israel | 0 | Asian |

| | | | |
|---|---|---|---|
| HGDP00651.SDG | Israel | 0 | Asian |
| HGDP00691.SDG | Israel | 0 | Asian |
| HGDP00700.SDG | Israel | 0 | Asian |
| HGDP00590.SDG | Israel | 0 | Asian |
| HGDP00632.SDG | Israel | 0 | Asian |
| HGDP00675.SDG | Israel | 0 | Asian |
| HGDP00575.SDG | Israel | 0 | Asian |
| HGDP00565.SDG | Israel | 0 | Asian |
| HGDP00741.SDG | Israel | 0 | Asian |
| HGDP00646.SDG | Israel | 0 | Asian |
| HGDP00122.SDG | Pakistan | 0 | Asian |
| HGDP00248.SDG | Pakistan | 0 | Asian |
| HGDP00254.SDG | Pakistan | 0 | Asian |
| HGDP00179.SDG | Pakistan | 0 | Asian |
| HGDP00241.SDG | Pakistan | 0 | Asian |
| HGDP00581.SDG | Israel | 0 | Asian |
| HGDP00045.SDG | Pakistan | 0 | Asian |
| HGDP00578.SDG | Israel | 0 | Asian |
| HGDP00631.SDG | Israel | 0 | Asian |
| HGDP00682.SDG | Israel | 0 | Asian |
| HGDP00653.SDG | Israel | 0 | Asian |
| HGDP00129.SDG | Pakistan | 0 | Asian |
| HGDP00604.SDG | Israel | 0 | Asian |
| HGDP00684.SDG | Israel | 0 | Asian |
| HGDP00639.SDG | Israel | 0 | Asian |
| HGDP00620.SDG | Israel | 0 | Asian |
| HGDP00566.SDG | Israel | 0 | Asian |
| HGDP00626.SDG | Israel | 0 | Asian |
| HGDP00102.SDG | Pakistan | 0 | Asian |

| | | | |
|---|---|---|---|
| HGDP00767.SDG | Japan | 0 | Asian |
| HGDP00326.SDG | Pakistan | 0 | Asian |
| HGDP00120.SDG | Pakistan | 0 | Asian |
| HGDP00175.SDG | Pakistan | 0 | Asian |
| HGDP00333.SDG | Pakistan | 0 | Asian |
| HGDP00588.SDG | Israel | 0 | Asian |
| HGDP00595.SDG | Israel | 0 | Asian |
| HGDP00621.SDG | Israel | 0 | Asian |
| HGDP00623.SDG | Israel | 0 | Asian |
| HGDP00685.SDG | Israel | 0 | Asian |
| HGDP00696.SDG | Israel | 0 | Asian |
| HGDP00687.SDG | Israel | 0 | Asian |
| HGDP00121.SDG | Pakistan | 0 | Asian |
| HGDP00388.SDG | Pakistan | 0 | Asian |
| HGDP01248.SDG | China | 0 | Asian |
| HGDP00698.SDG | Israel | 0 | Asian |
| HGDP00037.SDG | Pakistan | 0 | Asian |
| HGDP00307.SDG | Pakistan | 0 | Asian |
| HGDP00407.SDG | Pakistan | 0 | Asian |
| HGDP00259.SDG | Pakistan | 0 | Asian |
| HGDP00571.SDG | Israel | 0 | Asian |
| HGDP00641.SDG | Israel | 0 | Asian |
| HGDP00591.SDG | Israel | 0 | Asian |
| HGDP00643.SDG | Israel | 0 | Asian |
| HGDP00676.SDG | Israel | 0 | Asian |
| HGDP00640.SDG | Israel | 0 | Asian |
| HGDP00577.SDG | Israel | 0 | Asian |
| HGDP00686.SDG | Israel | 0 | Asian |
| HGDP00614.SDG | Israel | 0 | Asian |

| | | | |
|---|---|---|---|
| HGDP00732.SDG | Israel | 0 | Asian |
| HGDP00054.SDG | Pakistan | 0 | Asian |
| HGDP00191.SDG | Pakistan | 0 | Asian |
| HGDP00311.SDG | Pakistan | 0 | Asian |
| HGDP00752.SDG | Japan | 0 | Asian |
| HGDP00753.SDG | Japan | 0 | Asian |
| HGDP00727.SDG | Israel | 0 | Asian |
| HGDP00679.SDG | Israel | 0 | Asian |
| HGDP00688.SDG | Israel | 0 | Asian |
| HGDP00635.SDG | Israel | 0 | Asian |
| HGDP00557.SDG | Israel | 0 | Asian |
| HGDP00678.SDG | Israel | 0 | Asian |
| HGDP01217.SDG | China | 0 | Asian |
| HGDP00638.SDG | Israel | 0 | Asian |
| HGDP00618.SDG | Israel | 0 | Asian |
| HGDP00759.SDG | Japan | 0 | Asian |
| HGDP00230.SDG | Pakistan | 0 | Asian |
| HGDP00068.SDG | Pakistan | 0 | Asian |
| HGDP00029.SDG | Pakistan | 0 | Asian |
| HGDP00615.SDG | Israel | 0 | Asian |
| HGDP01309.SDG | China | 0 | Asian |
| HGDP00628.SDG | Israel | 0 | Asian |
| HGDP00594.SDG | Israel | 0 | Asian |
| HGDP00624.SDG | Israel | 0 | Asian |
| HGDP00598.SDG | Israel | 0 | Asian |
| HGDP00647.SDG | Israel | 0 | Asian |
| HGDP00735.SDG | Israel | 0 | Asian |
| HGDP00699.SDG | Israel | 0 | Asian |
| HGDP00602.SDG | Israel | 0 | Asian |

| | | | |
|---|---|---|---|
| HGDP00586.SDG | Israel | 0 | Asian |
| HGDP00648.SDG | Israel | 0 | Asian |
| HGDP01239.SDG | China | 0 | Asian |
| HGDP00690.SDG | Israel | 0 | Asian |
| HGDP00017.SDG | Pakistan | 0 | Asian |
| HGDP00444.SDG | Pakistan | 0 | Asian |
| HGDP00197.SDG | Pakistan | 0 | Asian |
| HGDP00560.SDG | Israel | 0 | Asian |
| HGDP00625.SDG | Israel | 0 | Asian |
| HGDP00607.SDG | Israel | 0 | Asian |
| HGDP00683.SDG | Israel | 0 | Asian |
| HGDP00738.SDG | Israel | 0 | Asian |
| HGDP00739.SDG | Israel | 0 | Asian |
| HGDP00629.SDG | Israel | 0 | Asian |
| HGDP00746.SDG | Israel | 0 | Asian |
| HGDP00619.SDG | Israel | 0 | Asian |
| HGDP00636.SDG | Israel | 0 | Asian |
| HGDP00574.SDG | Israel | 0 | Asian |
| HGDP00587.SDG | Israel | 0 | Asian |
| HGDP00599.SDG | Israel | 0 | Asian |
| HGDP00637.SDG | Israel | 0 | Asian |
| HGDP00677.SDG | Israel | 0 | Asian |
| HGDP00748.SDG | Japan | 0 | Asian |
| HGDP00171.SDG | Pakistan | 0 | Asian |
| HGDP00041.SDG | Pakistan | 0 | Asian |
| HGDP00023.SDG | Pakistan | 0 | Asian |
| HGDP00558.SDG | Israel | 0 | Asian |
| HGDP00611.SDG | Israel | 0 | Asian |
| HGDP00730.SDG | Israel | 0 | Asian |

| | | | |
|---|---|---|---|
| HGDP00745.SDG | Israel | 0 | Asian |
| HGDP00564.SDG | Israel | 0 | Asian |
| HGDP00433.SDG | Pakistan | 0 | Asian |
| HGDP00397.SDG | Pakistan | 0 | Asian |
| HGDP00237.SDG | Pakistan | 0 | Asian |
| HGDP01302.SDG | China | 0 | Asian |
| HGDP00729.SDG | Israel | 0 | Asian |
| HGDP00634.SDG | Israel | 0 | Asian |
| HGDP00693.SDG | Israel | 0 | Asian |
| HGDP00644.SDG | Israel | 0 | Asian |
| HGDP00613.SDG | Israel | 0 | Asian |
| HGDP00654.SDG | Israel | 0 | Asian |
| HGDP00734.SDG | Israel | 0 | Asian |
| HGDP00645.SDG | Israel | 0 | Asian |
| HGDP00701.SDG | Israel | 0 | Asian |
| HGDP00733.SDG | Israel | 0 | Asian |
| HGDP00744.SDG | Israel | 0 | Asian |
| HGDP01208.SDG | China | 0 | Asian |
| HGDP00302.SDG | Pakistan | 0 | Asian |
| HGDP01339.SDG | China | 0 | Asian |
| HGDP00568.SDG | Israel | 0 | Asian |
| HGDP00622.SDG | Israel | 0 | Asian |
| HGDP00692.SDG | Israel | 0 | Asian |
| HGDP00736.SDG | Israel | 0 | Asian |
| HGDP00612.SDG | Israel | 0 | Asian |
| HGDP00740.SDG | Israel | 0 | Asian |
| HGDP00584.SDG | Israel | 0 | Asian |
| HGDP00600.SDG | Israel | 0 | Asian |
| HGDP00627.SDG | Israel | 0 | Asian |

| | | | |
|---|---|---|---|
| HGDP00559.SDG | Israel | 0 | Asian |
| HGDP01301.SDG | China | 0 | Asian |
| HGDP00642.SDG | Israel | 0 | Asian |
| HGDP00694.SDG | Israel | 0 | Asian |
| HGDP00187.SDG | Pakistan | 0 | Asian |
| HGDP00274.SDG | Pakistan | 0 | Asian |
| HGDP00281.SDG | Pakistan | 0 | Asian |
| HGDP00606.SDG | Israel | 0 | Asian |
| HGDP00601.SDG | Israel | 0 | Asian |
| HGDP00689.SDG | Israel | 0 | Asian |
| HGDP00697.SDG | Israel | 0 | Asian |
| HGDP00064.SDG | Pakistan | 0 | Asian |
| HGDP00222.SDG | Pakistan | 0 | Asian |
| HGDP00007.SDG | Pakistan | 0 | Asian |
| HGDP00567.SDG | Israel | 0 | Asian |
| HGDP00206.SDG | Pakistan | 0 | Asian |
| HGDP00731.SDG | Israel | 0 | Asian |
| HGDP00630.SDG | Israel | 0 | Asian |
| HGDP00680.SDG | Israel | 0 | Asian |
| HGDP00778.SDG | China | 0 | Asian |
| HGDP01307.SDG | China | 0 | Asian |
| HGDP01323.SDG | China | 0 | Asian |
| HGDP00616.SDG | Israel | 0 | Asian |
| HGDP00725.SDG | Israel | 0 | Asian |
| HGDP01338.SDG | China | 0 | Asian |
| HGDP01333.SDG | China | 0 | Asian |
| HGDP00027.SDG | Pakistan | 0 | Asian |
| HGDP01345.SDG | China | 0 | Asian |
| HGDP00737.SDG | Israel | 0 | Asian |

| | | | |
|---|---|---|---|
| HGDP00232.SDG | Pakistan | 0 | Asian |
| HGDP01246.SDG | China | 0 | Asian |
| HGDP00773.SDG | Japan | 0 | Asian |
| HGDP00749.SDG | Japan | 0 | Asian |
| HGDP00597.SDG | Israel | 0 | Asian |
| HGDP01198.SDG | China | 0 | Asian |
| HGDP00058.SDG | Pakistan | 0 | Asian |
| HGDP00785.SDG | China | 0 | Asian |
| HGDP01098.SDG | China | 0 | Asian |
| HGDP00569.SDG | Israel | 0 | Asian |
| HGDP01240.SDG | China | 0 | Asian |
| HGDP01315.SDG | China | 0 | Asian |
| HGDP00160.SDG | Pakistan | 0 | Asian |
| HGDP00338.SDG | Pakistan | 0 | Asian |
| HGDP00216.SDG | Pakistan | 0 | Asian |
| HGDP01242.SDG | China | 0 | Asian |
| HGDP01320.SDG | China | 0 | Asian |
| HGDP01179.SDG | China | 0 | Asian |
| HGDP00124.SDG | Pakistan | 0 | Asian |
| HGDP01308.SDG | China | 0 | Asian |
| HGDP00090.SDG | Pakistan | 0 | Asian |
| HGDP00195.SDG | Pakistan | 0 | Asian |
| HGDP01188.SDG | China | 0 | Asian |
| HGDP01250.SDG | China | 0 | Asian |
| HGDP00650.SDG | Israel | 0 | Asian |
| HGDP00157.SDG | Pakistan | 0 | Asian |
| HGDP00286.SDG | Pakistan | 0 | Asian |
| HGDP00717.SDG | Cambodia | 0 | Asian |
| HGDP00783.SDG | China | 0 | Asian |

| | | | |
|---|---|---|---|
| HGDP00722.SDG | Israel | 0 | Asian |
| HGDP01335.SDG | China | 0 | Asian |
| HGDP01306.SDG | China | 0 | Asian |
| HGDP00713.SDG | Cambodia | 0 | Asian |
| HGDP00775.SDG | China | 0 | Asian |
| HGDP01215.SDG | China | 0 | Asian |
| HGDP00428.SDG | Pakistan | 0 | Asian |
| HGDP01350.SDG | China | 0 | Asian |
| HGDP01314.SDG | China | 0 | Asian |
| HGDP00208.SDG | Pakistan | 0 | Asian |
| HGDP01095.SDG | China | 0 | Asian |
| HGDP01223.SDG | China | 0 | Asian |
| HGDP01228.SDG | China | 0 | Asian |
| HGDP00125.SDG | Pakistan | 0 | Asian |
| HGDP01297.SDG | China | 0 | Asian |
| HGDP01191.SDG | China | 0 | Asian |
| HGDP00328.SDG | Pakistan | 0 | Asian |
| HGDP00019.SDG | Pakistan | 0 | Asian |
| HGDP01312.SDG | China | 0 | Asian |
| HGDP01355.SDG | China | 0 | Asian |
| HGDP01211.SDG | China | 0 | Asian |
| HGDP01203.SDG | China | 0 | Asian |
| I10771 | Israel | 3550 | Asian |
| I10769 | Israel | 3450 | Asian |
| I10770 | Israel | 3450 | Asian |
| I10768 | Israel | 3500 | Asian |
| I10411 | Turkmenistan | 4200 | Asian |
| I10410 | Turkmenistan | 3750 | Asian |
| I10409_published | Turkmenistan | 4113 | Asian |

| | | | |
|---|---|---|---|
| I11041_published | Turkmenistan | 4015 | Asian |
| I11026 | Uzbekistan | 3400 | Asian |
| I11027 | Uzbekistan | 3526 | Asian |
| I11042 | Turkmenistan | 4225 | Asian |
| I11028 | Uzbekistan | 5099 | Asian |
| I11040 | Turkmenistan | 3850 | Asian |
| I11025 | Uzbekistan | 3400 | Asian |
| I11039 | Turkmenistan | 4188 | Asian |
| I10551 | Tajikistan | 4450 | Asian |
| I12147 | Pakistan | 3052 | Asian |
| I11501 | Kazakhstan | 5164 | Asian |
| I11519 | Uzbekistan | 3400 | Asian |
| I11527 | Kyrgyzstan | 3943 | Asian |
| I11526 | Kyrgyzstan | 4079 | Asian |
| I11531 | Kazakhstan | 4937 | Asian |
| I11521 | Uzbekistan | 3400 | Asian |
| I11520 | Uzbekistan | 3497 | Asian |
| I10523 | Pakistan | 2850 | Asian |
| I11732 | Kazakhstan | 4921 | Asian |
| I11735 | Kazakhstan | 4328 | Asian |
| I11736 | Kazakhstan | 5021 | Asian |
| I11734 | Kazakhstan | 4918 | Asian |
| I11737 | Kazakhstan | 4046 | Asian |
| I11565 | Pakistan | 1450 | Asian |
| I12980 | Pakistan | 2850 | Asian |
| I12988 | Pakistan | 2850 | Asian |
| I12976 | Mongolia | 3332 | Asian |
| I12982 | Pakistan | 2850 | Asian |
| I12981 | Pakistan | 2850 | Asian |

| | | | |
|---|---|---|---|
| I12968 | Pakistan | 2850 | Asian |
| I12969 | Mongolia | 3001 | Asian |
| I12975 | Mongolia | 3110 | Asian |
| I12955 | Mongolia | 3824 | Asian |
| I12987 | Pakistan | 2850 | Asian |
| I12960 | Mongolia | 3332 | Asian |
| I12970 | Mongolia | 2286 | Asian |
| I12971 | Mongolia | 2849 | Asian |
| I12957 | Mongolia | 4462 | Asian |
| I12977 | Mongolia | 4822 | Asian |
| I12972 | Mongolia | 3347 | Asian |
| I12956 | Mongolia | 2326 | Asian |
| I12978 | Mongolia | 4472 | Asian |
| I12973 | Mongolia | 3226 | Asian |
| I12958 | Mongolia | 3483 | Asian |
| I12984 | Pakistan | 2850 | Asian |
| I12985 | Pakistan | 2850 | Asian |
| I12979 | Pakistan | 2850 | Asian |
| I12470 | Pakistan | 2850 | Asian |
| I12448 | Pakistan | 2850 | Asian |
| I12485 | Turkmenistan | 5050 | Asian |
| I12451 | Pakistan | 2850 | Asian |
| I12456 | Pakistan | 2850 | Asian |
| I12471 | Pakistan | 2850 | Asian |
| I12463 | Pakistan | 2850 | Asian |
| I12455 | Pakistan | 2850 | Asian |
| I12464 | Pakistan | 2850 | Asian |
| I12499 | Uzbekistan | 3369 | Asian |
| I12477 | Pakistan | 2850 | Asian |

| | | | |
|---|---|---|---|
| I12462 | Pakistan | 2850 | Asian |
| I12457 | Pakistan | 2928 | Asian |
| I12483 | Turkmenistan | 5050 | Asian |
| I12450 | Pakistan | 2751 | Asian |
| I12478 | Turkmenistan | 5050 | Asian |
| I12487 | Turkmenistan | 5050 | Asian |
| I12475 | Pakistan | 2850 | Asian |
| I12465 | Pakistan | 2850 | Asian |
| I12466 | Pakistan | 2850 | Asian |
| I12482 | Turkmenistan | 5050 | Asian |
| I12446 | Pakistan | 2850 | Asian |
| I12481 | Turkmenistan | 5050 | Asian |
| I12445 | Pakistan | 2850 | Asian |
| I12480 | Turkmenistan | 5050 | Asian |
| I12460 | Pakistan | 2850 | Asian |
| I12453 | Pakistan | 2850 | Asian |
| I12486 | Turkmenistan | 5050 | Asian |
| I12447 | Pakistan | 2850 | Asian |
| I12461 | Pakistan | 2850 | Asian |
| I12454 | Pakistan | 2850 | Asian |
| I12458 | Pakistan | 2850 | Asian |
| I12459 | Pakistan | 2850 | Asian |
| I12444 | Pakistan | 2850 | Asian |
| I12479 | Turkmenistan | 5050 | Asian |
| I12473 | Pakistan | 2850 | Asian |
| I13174 | Mongolia | 3410 | Asian |
| I13224 | Pakistan | 2850 | Asian |
| I13173 | Mongolia | 3722 | Asian |
| I13175 | Mongolia | 1483 | Asian |

| | | | |
|---|---|---|---|
| I13222 | Pakistan | 2850 | Asian |
| I13180 | Mongolia | 4862 | Asian |
| I13223 | Pakistan | 2850 | Asian |
| I13227 | Pakistan | 2850 | Asian |
| I13226 | Pakistan | 2911 | Asian |
| I13221 | Pakistan | 2850 | Asian |
| I13220 | Pakistan | 2850 | Asian |
| I13219 | Pakistan | 2850 | Asian |
| I13178 | Mongolia | 2923 | Asian |
| I13228 | Pakistan | 2850 | Asian |
| I12294 | Tajikistan | 1958 | Asian |
| I12257 | Tajikistan | 2000 | Asian |
| I12260 | Tajikistan | 2051 | Asian |
| I12292 | Tajikistan | 2036 | Asian |
| I12293 | Tajikistan | 2000 | Asian |
| I11697 | Mongolia | 7500 | Asian |
| I11696 | Mongolia | 7528 | Asian |
| I13504 | Mongolia | 2368 | Asian |
| I13505 | Mongolia | 3001 | Asian |
| I13766 | Mongolia | 3208 | Asian |
| I13767 | Mongolia | 3198 | Asian |
| IK002.SG | Japan | 2782 | Asian |
| In661.WGC | Indonesia | 1827 | Asian |
| In662.SG | Indonesia | 2161 | Asian |
| JHF05.SG | Malaysia | 0 | Asian |
| JHM06.SG | Malaysia | 0 | Asian |
| La364.SG | Laos | 2987 | Asian |
| La368.SG | Laos | 7872 | Asian |
| La727.WGC | Laos | 2328 | Asian |

| | | | |
|---|---|---|---|
| La898.SG | Laos | 5951 | Asian |
| Ma554.WGC | Malaysia | 432 | Asian |
| Ma555.SG | Malaysia | 378 | Asian |
| Ma911.WGC | Malaysia | 4303 | Asian |
| Ma912.SG | Malaysia | 2459 | Asian |
| Phl534.WGC | Philippines | 1783 | Asian |
| Th519.SG | Thailand | 1659 | Asian |
| Th521.SG | Thailand | 1666 | Asian |
| Th530.SG | Thailand | 1646 | Asian |
| Th531.WGC | Thailand | 1585 | Asian |
| Th703.WGC | Thailand | 1659 | Asian |
| Vt719.SG | Vietnam | 219 | Asian |
| Vt777.SG | Vietnam | 2271 | Asian |
| Vt778.SG | Vietnam | 2648 | Asian |
| Vt779.SG | Vietnam | 2237 | Asian |
| Vt781.SG | Vietnam | 2242 | Asian |
| Vt796.SG | Vietnam | 2138 | Asian |
| Vt808.SG | Vietnam | 2248 | Asian |
| Vt833.SG | Vietnam | 4168 | Asian |
| Vt880.SG | Vietnam | 3950 | Asian |
| I14037 | Mongolia | 3306 | Asian |
| I14194 | Mongolia | 2834 | Asian |
| I13692 | Taiwan | 1550 | Asian |
| I13695 | Taiwan | 1459 | Asian |
| I13694 | Taiwan | 1550 | Asian |
| Andaman.SG | India | 90 | Asian |
| I6348 | Mongolia | 3632 | Asian |
| I11698 | Mongolia | 7520 | Asian |
| I13959 | Mongolia | 3339 | Asian |

| | | | |
|---|---|---|---|
| I13883 | Japan | 2859 | Asian |
| I13884 | Japan | 4351 | Asian |
| I13885 | Japan | 3216 | Asian |
| I13882 | Japan | 3234 | Asian |
| I13886 | Japan | 4003 | Asian |
| I13957 | Mongolia | 4625 | Asian |
| I13964 | Mongolia | 3350 | Asian |
| I13965 | Mongolia | 2270 | Asian |
| I13887 | Japan | 4032 | Asian |
| I13963 | Mongolia | 2829 | Asian |
| I14926 | Taiwan | 1550 | Asian |
| I14925 | Taiwan | 1550 | Asian |
| I14933 | Taiwan | 1550 | Asian |
| I14928 | Taiwan | 1550 | Asian |
| I14931 | Taiwan | 1550 | Asian |
| I14930 | Taiwan | 1550 | Asian |
| I14929 | Taiwan | 1550 | Asian |
| I14934 | Taiwan | 1550 | Asian |
| I14927 | Taiwan | 1550 | Asian |
| I14937 | Taiwan | 1550 | Asian |
| I13721 | Taiwan | 3169 | Asian |
| I14000 | Mongolia | 7480 | Asian |
| ARS001.A0101 | Mongolia | 2880 | Asian |
| ARS002.B0101 | Mongolia | 3055 | Asian |
| ARS003.B0101 | Mongolia | 3247 | Asian |
| ARS004.A0101 | Mongolia | 2983 | Asian |
| ARS005.A0101 | Mongolia | 3038 | Asian |
| ARS006.A0101 | Mongolia | 3003 | Asian |
| ARS007.B0101 | Mongolia | 3172 | Asian |

| | | | |
|---|---|---|---|
| ARS008.A0101 | Mongolia | 3173 | Asian |
| ARS009.A0101 | Mongolia | 2900 | Asian |
| ARS011.A0101 | Mongolia | 3081 | Asian |
| ARS012.A0101 | Mongolia | 3229 | Asian |
| ARS013.A0101 | Mongolia | 3279 | Asian |
| ARS014.B0101 | Mongolia | 3234 | Asian |
| ARS015.A0101 | Mongolia | 3255 | Asian |
| ARS016.A0101 | Mongolia | 3389 | Asian |
| ARS017.A0101 | Mongolia | 3256 | Asian |
| ARS018.A0101 | Mongolia | 2951 | Asian |
| ARS024.A0101 | Mongolia | 3171 | Asian |
| ARS025.A0101 | Mongolia | 3263 | Asian |
| ARS026.A0101 | Mongolia | 2957 | Asian |
| BAJ001_BAJ001.A0101 | Jordan | 8850 | Asian |
| KFH2_KFH002.A0101 | Israel | 9591 | Asian |
| QED-12.SG | Lebanon | 1525 | Asian |
| QED-2.SG | Lebanon | 1610 | Asian |
| QED-4.SG | Lebanon | 1406 | Asian |
| QED-7.SG | Lebanon | 1627 | Asian |
| SI-38.SG | Lebanon | 800 | Asian |
| SI-39.SG | Lebanon | 702 | Asian |
| SI-40.SG | Lebanon | 800 | Asian |
| SI-41.SG | Lebanon | 710 | Asian |
| SI-42.SG | Lebanon | 725 | Asian |
| SI-44.SG | Lebanon | 800 | Asian |
| SI-45.SG | Lebanon | 698 | Asian |
| SI-47.SG | Lebanon | 800 | Asian |
| SI-53.SG | Lebanon | 851 | Asian |
| BOT2016 | Kazakhstan | 5396 | Asian |

| | | | |
|---|---|---|---|
| I13698 | Mongolia | 7504 | Asian |
| I15162 | Taiwan | 1550 | Asian |
| I15156 | Taiwan | 1550 | Asian |
| I15164 | Taiwan | 1550 | Asian |
| I15158 | Taiwan | 1550 | Asian |
| I15166 | Taiwan | 1550 | Asian |
| I15160 | Taiwan | 1550 | Asian |
| I15159 | Taiwan | 1550 | Asian |
| I6113_d | India | 4300 | Asian |
| I4319_published | Kazakhstan | 3632 | Asian |
| I5399_published | Pakistan | 2850 | Asian |
| I2921_published | Iran | 5533 | Asian |
| I8219_published | Pakistan | 2841 | Asian |
| I11537_published | Kazakhstan | 2554 | Asian |
| I11540_published | Kazakhstan | 1723 | Asian |
| I11538_published | Kazakhstan | 3614 | Asian |
| I11542_published | Kazakhstan | 3250 | Asian |
| I11541_published | Kazakhstan | 3684 | Asian |
| I11458_published | Iran | 4600 | Asian |
| I12146_published | Pakistan | 2850 | Asian |
| I12138_published | Pakistan | 2949 | Asian |
| I12144_published | Pakistan | 2850 | Asian |
| I12136_published | Pakistan | 2850 | Asian |
| I12142_published | Pakistan | 2850 | Asian |
| I12149_published | Pakistan | 2850 | Asian |
| I12134_published | Pakistan | 2850 | Asian |
| I12137_published | Pakistan | 2850 | Asian |
| I12143_published | Pakistan | 2850 | Asian |
| I12141_published | Pakistan | 2850 | Asian |

| | | | |
|---|---|---|---|
| I12140_published | Pakistan | 2850 | Asian |
| I12983_published | Pakistan | 2850 | Asian |
| I12449_published | Pakistan | 2850 | Asian |
| F004.SG | China | 2230 | Asian |
| M010.SG | China | 2292 | Asian |
| M012.SG | China | 2224 | Asian |
| M15-1.SG | China | 2250 | Asian |
| M15-2.SG | China | 2250 | Asian |
| M4.SG | China | 2227 | Asian |
| M8R1.SG | China | 2231 | Asian |
| X3.SG | China | 2250 | Asian |
| M819.SG | China | 2250 | Asian |
| M820.SG | China | 2213 | Asian |
| M827.SG | China | 2300 | Asian |
| MJ-51.SG | Kazakhstan | 2587 | Asian |
| MJ-52.SG | Kazakhstan | 2621 | Asian |
| MJ-53.SG | Kazakhstan | 2246 | Asian |
| MJ-54.SG | Kazakhstan | 2551 | Asian |
| ASH008 | Israel | 3095 | Asian |
| ASH029 | Israel | 3513 | Asian |
| ASH033 | Israel | 3639 | Asian |
| ASH034 | Israel | 3550 | Asian |
| ASH066 | Israel | 3184 | Asian |
| ASH067 | Israel | 3206 | Asian |
| ASH068 | Israel | 3167 | Asian |
| ASH087 | Israel | 3100 | Asian |
| ASH135 | Israel | 3100 | Asian |
| ASH2-3 | Israel | 3213 | Asian |
| Sanganji131464.SG | Japan | 3280 | Asian |

| | | | |
|---|---|---|---|
| BOT14 | Kazakhstan | 5337 | Asian |
| I8072 | Taiwan | 1550 | Asian |
| I1293_all_d | Iran | 10800 | Asian |
| I10263_published | Israel | 3500 | Asian |
| I12255 | Turkmenistan | 3800 | Asian |
| I4567_published | Kazakhstan | 3626 | Asian |
| I10270_published | Israel | 3500 | Asian |
| I10359_published | Israel | 3512 | Asian |
| I2312_all_published_d | Iran | 11950 | Asian |
| I11562 | Pakistan | 1450 | Asian |
| I12472 | Pakistan | 2850 | Asian |
| I12484_published | Turkmenistan | 5100 | Asian |
| I6341 | Japan | 3200 | Asian |
| ETM001 | Syria | 3800 | Asian |
| ETM003 | Syria | 2818 | Asian |
| ETM004 | Syria | 3850 | Asian |
| ETM005 | Syria | 3850 | Asian |
| ETM006 | Syria | 3850 | Asian |
| ETM010 | Syria | 4550 | Asian |
| ETM012 | Syria | 4470 | Asian |
| ETM014 | Syria | 3850 | Asian |
| ETM015 | Syria | 3850 | Asian |
| ETM016 | Syria | 3914 | Asian |
| ETM017 | Syria | 3850 | Asian |
| ETM018 | Syria | 4004 | Asian |
| ETM021 | Syria | 3800 | Asian |
| ETM023 | Syria | 4275 | Asian |
| ETM025 | Syria | 3800 | Asian |
| ETM026 | Syria | 3850 | Asian |

| | | | |
|---|---|---|---|
| 18R21262.EC | China | 5050 | Asian |
| 18R21265.EC | China | 5050 | Asian |
| 18R21266.EC | China | 4785 | Asian |
| S118.EC | China | 5050 | Asian |
| S120.EC | China | 5050 | Asian |
| S123.EC | China | 5050 | Asian |
| S91.EC | China | 5050 | Asian |
| S94.EC | China | 5050 | Asian |
| S95.EC | China | 5050 | Asian |
| S96.EC | China | 5050 | Asian |
| S97.EC | China | 5050 | Asian |
| SFI-11.SG | Lebanon | 1984 | Asian |
| SFI-12.SG | Lebanon | 2123 | Asian |
| SFI-15.SG | Lebanon | 2027 | Asian |
| SFI-20.SG | Lebanon | 2043 | Asian |
| SFI-24.SG | Lebanon | 1946 | Asian |
| SFI-33.SG | Lebanon | 1804 | Asian |
| SFI-34.SG | Lebanon | 2385 | Asian |
| SFI-35.SG | Lebanon | 2385 | Asian |
| SFI-36.SG | Lebanon | 2385 | Asian |
| SFI-39.SG | Lebanon | 2385 | Asian |
| SFI-42.SG | Lebanon | 2415 | Asian |
| SFI-43.SG | Lebanon | 2487 | Asian |
| SFI-44.SG | Lebanon | 2385 | Asian |
| SFI-45.SG | Lebanon | 2385 | Asian |
| SFI-47.SG | Lebanon | 2385 | Asian |
| SFI-5.SG | Lebanon | 2159 | Asian |
| SFI-50.SG | Lebanon | 2385 | Asian |
| SFI-55.SG | Lebanon | 2720 | Asian |

| | | | |
|---|---|---|---|
| SFI-56.SG | Lebanon | 2720 | Asian |
| 91KLH11 | China | 2650 | Asian |
| 91KLH18 | China | 2812 | Asian |
| 91KLM2 | China | 2650 | Asian |
| BLSM27S | China | 5250 | Asian |
| BLSM41 | China | 5250 | Asian |
| BLSM45 | China | 5162 | Asian |
| DCZM17IV | China | 1850 | Asian |
| DCZ-M21II | China | 1763 | Asian |
| DCZ-M22IV | China | 1850 | Asian |
| DCZ-M6 | China | 1850 | Asian |
| EDM124 | China | 3544 | Asian |
| EDM139 | China | 3750 | Asian |
| EDM176 | China | 3750 | Asian |
| HJTM107 | China | 3993 | Asian |
| HJTM109 | China | 3908 | Asian |
| HJTM115 | China | 2225 | Asian |
| HJTW13 | China | 2131 | Asian |
| HMF32 | China | 5594 | Asian |
| JCKM1-1 | China | 4296 | Asian |
| JXNTM23 | China | 3332 | Asian |
| JXNTM2 | China | 3250 | Asian |
| LGM41 | China | 2100 | Asian |
| LGM79 | China | 2250 | Asian |
| LJM14 | China | 3900 | Asian |
| LJM25 | China | 3900 | Asian |
| LJM2 | China | 3900 | Asian |
| LJM3 | China | 4479 | Asian |
| LJM4 | China | 3900 | Asian |

| | | | |
|---|---|---|---|
| LJM5 | China | 3900 | Asian |
| MGS-M6 | China | 1800 | Asian |
| MGS-M7L | China | 1800 | Asian |
| MGS-M7R | China | 1800 | Asian |
| MZGM10-1 | China | 5250 | Asian |
| MZGM16 | China | 5250 | Asian |
| MZGM25-2 | China | 5250 | Asian |
| PLTM310 | China | 4088 | Asian |
| PLTM311 | China | 4049 | Asian |
| PLTM312 | China | 3977 | Asian |
| PLTM313 | China | 3934 | Asian |
| SM-SGDLM27 | China | 4050 | Asian |
| SM-SGDLM6 | China | 4589 | Asian |
| SM-SGDLM7X | China | 4050 | Asian |
| WD-WT1H16 | China | 4536 | Asian |
| WD-WT5M2 | China | 3900 | Asian |
| WGH35-1 | China | 5250 | Asian |
| WGM20 | China | 5250 | Asian |
| WGM35 | China | 5950 | Asian |
| WGM43 | China | 5250 | Asian |
| WGM70 | China | 5250 | Asian |
| WGM76S | China | 5250 | Asian |
| WGM94 | China | 5250 | Asian |
| WQM4 | China | 7361 | Asian |
| XW-M1R18 | China | 6927 | Asian |
| ZLNR-1 | China | 1786 | Asian |
| ZLNR-2 | China | 7389 | Asian |
| L5692_d | China | 4477 | Asian |
| L5706_d | China | 4459 | Asian |

| | | | |
|---|---|---|---|
| SuogangB1_d | China | 4550 | Asian |
| SuogangB3_d | China | 4550 | Asian |
| L5696_d | China | 4350 | Asian |
| L5698_d | China | 4287 | Asian |
| L5701_d | China | 4325 | Asian |
| L5703_d | China | 4571 | Asian |
| L5704_d | China | 4505 | Asian |
| L7417_d | China | 4472 | Asian |
| L5694 | China | 338 | Asian |
| L5700 | China | 4477 | Asian |
| L5705 | China | 4333 | Asian |
| L7415 | China | 4333 | Asian |
| Qihe2_d | China | 8392 | Asian |
| Xiaogao | China | 7827 | Asian |
| XJS1309_M4 | China | 7827 | Asian |
| XJS1309_M7 | China | 7814 | Asian |
| XJS1311_M16 | China | 7850 | Asian |
| Yumin | China | 8375 | Asian |
| BS | China | 9125 | Asian |
| LD1 | China | 8190 | Asian |
| LD2 | China | 7575 | Asian |
| Bianbian | China | 9709 | Asian |
| I13179 | Mongolia | 7515 | Asian |
| ARG001 | Mongolia | 700 | Asian |
| ARG002 | Mongolia | 700 | Asian |
| ARG003 | Mongolia | 700 | Asian |
| AST001 | Mongolia | 2056 | Asian |
| ATS001 | Mongolia | 1878 | Asian |
| BAM001 | Mongolia | 2000 | Asian |

| | | | |
|---|---|---|---|
| BAU001 | Mongolia | 600 | Asian |
| BAY001 | Mongolia | 700 | Asian |
| BAZ001 | Mongolia | 700 | Asian |
| BER002 | Mongolia | 3168 | Asian |
| BIL001 | Mongolia | 3297 | Asian |
| BOR001 | Mongolia | 2858 | Asian |
| BRG001 | Mongolia | 700 | Asian |
| BRG002 | Mongolia | 700 | Asian |
| BRG004 | Mongolia | 700 | Asian |
| BRG005 | Mongolia | 700 | Asian |
| BRL001 | Mongolia | 601 | Asian |
| BRL002 | Mongolia | 600 | Asian |
| BRU001 | Mongolia | 2000 | Asian |
| BTO001 | Mongolia | 2000 | Asian |
| BUL001 | Mongolia | 3306 | Asian |
| BUL002 | Mongolia | 3300 | Asian |
| BUR001 | Mongolia | 1993 | Asian |
| BUR002 | Mongolia | 1800 | Asian |
| BUR003 | Mongolia | 1800 | Asian |
| BUR004 | Mongolia | 1919 | Asian |
| CHD001 | Mongolia | 700 | Asian |
| CHN001 | Mongolia | 2226 | Asian |
| CHN003 | Mongolia | 2232 | Asian |
| CHN004 | Mongolia | 1400 | Asian |
| CHN006 | Mongolia | 1400 | Asian |
| CHN007 | Mongolia | 1400 | Asian |
| CHN008 | Mongolia | 1400 | Asian |
| CHN010 | Mongolia | 1961 | Asian |
| CHN012 | Mongolia | 1400 | Asian |

| | | | |
|---|---|---|---|
| CHN014 | Mongolia | 574 | Asian |
| CHN015 | Mongolia | 1400 | Asian |
| CHN016 | Mongolia | 1400 | Asian |
| DAR001 | Mongolia | 2503 | Asian |
| DAR002 | Mongolia | 2525 | Asian |
| DAS001 | Mongolia | 700 | Asian |
| DEE001 | Mongolia | 540 | Asian |
| DEL001 | Mongolia | 1978 | Asian |
| DOL001 | Mongolia | 2000 | Asian |
| DUU001 | Mongolia | 1941 | Asian |
| DUU002 | Mongolia | 704 | Asian |
| EME002 | Mongolia | 2000 | Asian |
| ERD001 | Mongolia | 700 | Asian |
| ERM001 | Mongolia | 5653 | Asian |
| GAN002 | Mongolia | 700 | Asian |
| GTO001 | Mongolia | 700 | Asian |
| GUN002 | Mongolia | 700 | Asian |
| HUD001 | Mongolia | 2000 | Asian |
| IAG001 | Mongolia | 4466 | Asian |
| IKU001 | Mongolia | 700 | Asian |
| JAA001 | Mongolia | 2051 | Asian |
| JAG001 | Mongolia | 2082 | Asian |
| KGK001 | Mongolia | 700 | Asian |
| KHI001 | Mongolia | 2930 | Asian |
| KHL001 | Mongolia | 700 | Asian |
| KHN001 | Mongolia | 700 | Asian |
| KHO001 | Mongolia | 746 | Asian |
| KHO006 | Mongolia | 1875 | Asian |
| KHO007 | Mongolia | 1864 | Asian |

| | | | |
|---|---|---|---|
| KHU001 | Mongolia | 3300 | Asian |
| KHV002 | Mongolia | 700 | Asian |
| KNN001 | Mongolia | 700 | Asian |
| KNU001 | Mongolia | 700 | Asian |
| KRN001 | Mongolia | 700 | Asian |
| KRN002 | Mongolia | 700 | Asian |
| KUM001 | Mongolia | 4862 | Asian |
| KUR001 | Mongolia | 4482 | Asian |
| MIT001 | Mongolia | 3010 | Asian |
| MRI001 | Mongolia | 700 | Asian |
| NAI001 | Mongolia | 2000 | Asian |
| NAI002 | Mongolia | 2000 | Asian |
| NOM001 | Mongolia | 1250 | Asian |
| NRC001 | Mongolia | 700 | Asian |
| OLN001 | Mongolia | 1075 | Asian |
| OLN002 | Mongolia | 1075 | Asian |
| OLN003 | Mongolia | 1088 | Asian |
| OLN004 | Mongolia | 1075 | Asian |
| OLN005 | Mongolia | 1075 | Asian |
| OLN007 | Mongolia | 1075 | Asian |
| OLN008 | Mongolia | 1075 | Asian |
| OLN009 | Mongolia | 1075 | Asian |
| OLN010 | Mongolia | 1075 | Asian |
| OLN011 | Mongolia | 1075 | Asian |
| OLN012 | Mongolia | 1075 | Asian |
| RAH001 | Mongolia | 700 | Asian |
| SAN001 | Mongolia | 1890 | Asian |
| SBG001 | Mongolia | 3314 | Asian |
| SHA001 | Mongolia | 700 | Asian |

| | | | |
|---|---|---|---|
| SHG001 | Mongolia | 700 | Asian |
| SHG002 | Mongolia | 700 | Asian |
| SHG003 | Mongolia | 700 | Asian |
| SHR001 | Mongolia | 700 | Asian |
| SHT001 | Mongolia | 5009 | Asian |
| SHT002 | Mongolia | 4996 | Asian |
| SHU001 | Mongolia | 2929 | Asian |
| SHU002 | Mongolia | 726 | Asian |
| SKT001 | Mongolia | 2051 | Asian |
| SKT002 | Mongolia | 2103 | Asian |
| SKT003 | Mongolia | 2051 | Asian |
| SKT004 | Mongolia | 2051 | Asian |
| SKT005 | Mongolia | 2051 | Asian |
| SKT006 | Mongolia | 2048 | Asian |
| SKT007 | Mongolia | 2051 | Asian |
| SKT008 | Mongolia | 2051 | Asian |
| SKT009 | Mongolia | 2051 | Asian |
| SKT010 | Mongolia | 2176 | Asian |
| SKT012 | Mongolia | 2051 | Asian |
| SOL001 | Mongolia | 2000 | Asian |
| SON001 | Mongolia | 2000 | Asian |
| SOU001 | Mongolia | 6529 | Asian |
| TAH002 | Mongolia | 700 | Asian |
| TAK001 | Mongolia | 2000 | Asian |
| TAV001 | Mongolia | 1300 | Asian |
| TAV005 | Mongolia | 1300 | Asian |
| TAV006 | Mongolia | 1300 | Asian |
| TAV011 | Mongolia | 1300 | Asian |
| TEV002 | Mongolia | 2000 | Asian |

| | | | |
|---|---|---|---|
| TEV003 | Mongolia | 2000 | Asian |
| TMI001 | Mongolia | 1919 | Asian |
| TSA001 | Mongolia | 700 | Asian |
| TSA002 | Mongolia | 700 | Asian |
| TSA003 | Mongolia | 700 | Asian |
| TSA004 | Mongolia | 700 | Asian |
| TSA005 | Mongolia | 700 | Asian |
| TSA006 | Mongolia | 700 | Asian |
| TSA007 | Mongolia | 700 | Asian |
| TSB001 | Mongolia | 1231 | Asian |
| TSI001 | Mongolia | 2797 | Asian |
| TUH001 | Mongolia | 1800 | Asian |
| TUH002 | Mongolia | 1800 | Asian |
| TUK001 | Mongolia | 1623 | Asian |
| TUK002 | Mongolia | 1800 | Asian |
| TUK003 | Mongolia | 1800 | Asian |
| TUM001 | Mongolia | 1250 | Asian |
| UAA001 | Mongolia | 3722 | Asian |
| UGO001 | Mongolia | 700 | Asian |
| UGO002 | Mongolia | 700 | Asian |
| UGU001 | Mongolia | 1225 | Asian |
| UGU002 | Mongolia | 1927 | Asian |
| UGU003 | Mongolia | 1550 | Asian |
| UGU004 | Mongolia | 1927 | Asian |
| UGU005 | Mongolia | 1885 | Asian |
| UGU006 | Mongolia | 1550 | Asian |
| UGU010 | Mongolia | 1939 | Asian |
| UGU011 | Mongolia | 1550 | Asian |
| ULA001 | Mongolia | 846 | Asian |

| | | | |
|---|---|---|---|
| ULI001 | Mongolia | 3131 | Asian |
| ULI002 | Mongolia | 1248 | Asian |
| ULI003 | Mongolia | 3046 | Asian |
| ULN001 | Mongolia | 3239 | Asian |
| ULN002 | Mongolia | 2100 | Asian |
| ULN003 | Mongolia | 3289 | Asian |
| ULN004 | Mongolia | 1952 | Asian |
| ULN005 | Mongolia | 3429 | Asian |
| ULN006 | Mongolia | 2100 | Asian |
| ULN007 | Mongolia | 2100 | Asian |
| ULN009 | Mongolia | 2100 | Asian |
| ULN010 | Mongolia | 2100 | Asian |
| ULN011 | Mongolia | 719 | Asian |
| ULN015 | Mongolia | 3317 | Asian |
| ULZ001 | Mongolia | 2921 | Asian |
| UUS001 | Mongolia | 600 | Asian |
| UUS002 | Mongolia | 604 | Asian |
| UVG001 | Mongolia | 2000 | Asian |
| YAG001 | Mongolia | 4462 | Asian |
| YUR001 | Mongolia | 1958 | Asian |
| ZAA001 | Mongolia | 1075 | Asian |
| ZAA002 | Mongolia | 1324 | Asian |
| ZAA003 | Mongolia | 867 | Asian |
| ZAA004 | Mongolia | 1075 | Asian |
| ZAA005 | Mongolia | 850 | Asian |
| ZAA007 | Mongolia | 1227 | Asian |
| ZAM001 | Mongolia | 700 | Asian |
| ZAM002 | Mongolia | 700 | Asian |
| ZAR002 | Mongolia | 700 | Asian |

| | | | |
|---|---|---|---|
| ZAY001 | Mongolia | 700 | Asian |
| salkhit1.SG | Mongolia | 34873 | Asian |
| I1072_published_d | Israel | 12700 | Asian |
| I13768_d | Mongolia | 3074 | Asian |
| tai001.SG | Taiwan | 8195 | Asian |
| tai002.SG | Taiwan | 7550 | Asian |
| AIG001 | Kazakhstan | 2350 | Asian |
| AIG002 | Kazakhstan | 2350 | Asian |
| AIG003 | Kazakhstan | 2234 | Asian |
| AIG005 | Kazakhstan | 1790 | Asian |
| AIG006 | Kazakhstan | 2350 | Asian |
| AKB001 | Kazakhstan | 2677 | Asian |
| ALN001 | Kyrgyzstan | 1632 | Asian |
| ALN002 | Kyrgyzstan | 1719 | Asian |
| ALN003 | Kyrgyzstan | 1700 | Asian |
| ALN004 | Kyrgyzstan | 1621 | Asian |
| ALN005 | Kyrgyzstan | 1592 | Asian |
| ALN006 | Kyrgyzstan | 2507 | Asian |
| ALN007 | Kyrgyzstan | 2346 | Asian |
| ALN008 | Kyrgyzstan | 1700 | Asian |
| ALN009 | Kyrgyzstan | 1700 | Asian |
| BDY001 | Kazakhstan | 2225 | Asian |
| BDY002 | Kazakhstan | 2255 | Asian |
| BDY003 | Kazakhstan | 2123 | Asian |
| BIR010 | Kazakhstan | 2550 | Asian |
| BIR012 | Kazakhstan | 2606 | Asian |
| BIR013 | Kazakhstan | 2585 | Asian |
| BKT001 | Kazakhstan | 2587 | Asian |
| BRE001 | Kazakhstan | 2224 | Asian |

| | | | |
|---|---|---|---|
| BRE002 | Kazakhstan | 2223 | Asian |
| BRE003 | Kazakhstan | 2100 | Asian |
| BRE004 | Kazakhstan | 1627 | Asian |
| BRE005 | Kazakhstan | 1905 | Asian |
| BRE006 | Kazakhstan | 2100 | Asian |
| BRE007 | Kazakhstan | 1667 | Asian |
| BRE008 | Kazakhstan | 1629 | Asian |
| BRE009 | Kazakhstan | 1886 | Asian |
| BRE010 | Kazakhstan | 2100 | Asian |
| BRE011 | Kazakhstan | 1615 | Asian |
| BRE012 | Kazakhstan | 1886 | Asian |
| BRE013 | Kazakhstan | 1716 | Asian |
| BRE014 | Kazakhstan | 1630 | Asian |
| BRV001 | Kazakhstan | 2000 | Asian |
| BSB001 | Kazakhstan | 2592 | Asian |
| BSB002 | Kazakhstan | 2300 | Asian |
| BSB003 | Kazakhstan | 2300 | Asian |
| BSB004 | Kazakhstan | 2300 | Asian |
| CHK001 | Kyrgyzstan | 2373 | Asian |
| CHK002 | Kyrgyzstan | 2522 | Asian |
| CHK003 | Kyrgyzstan | 2539 | Asian |
| CHK004 | Kyrgyzstan | 1129 | Asian |
| CHK005 | Kyrgyzstan | 2050 | Asian |
| CLK001 | Kazakhstan | 2400 | Asian |
| CSP001 | Kazakhstan | 2241 | Asian |
| CSP002 | Kazakhstan | 2350 | Asian |
| CSP003 | Kazakhstan | 2654 | Asian |
| CSP004 | Kazakhstan | 2350 | Asian |
| CSP005 | Kazakhstan | 2739 | Asian |

| | | | |
|---|---|---|---|
| ESZ001 | Kazakhstan | 2612 | Asian |
| ESZ002 | Kazakhstan | 2583 | Asian |
| ESZ003 | Kazakhstan | 2600 | Asian |
| KBO001 | Kazakhstan | 2297 | Asian |
| KBU001 | Kazakhstan | 2300 | Asian |
| KBU002 | Kazakhstan | 2300 | Asian |
| KBU003 | Kazakhstan | 2300 | Asian |
| KEN001 | Kyrgyzstan | 2202 | Asian |
| KEN002 | Kyrgyzstan | 2406 | Asian |
| KEN003 | Kyrgyzstan | 2151 | Asian |
| KKB001 | Kazakhstan | 1109 | Asian |
| KKB002 | Kazakhstan | 1250 | Asian |
| KKM001 | Kazakhstan | 2550 | Asian |
| KLK002 | Kazakhstan | 1050 | Asian |
| KNT001 | Kazakhstan | 1641 | Asian |
| KNT002 | Kazakhstan | 1650 | Asian |
| KNT003 | Kazakhstan | 1650 | Asian |
| KNT004 | Kazakhstan | 1653 | Asian |
| KNT005 | Kazakhstan | 1622 | Asian |
| KRY001 | Kazakhstan | 1552 | Asian |
| KSH001 | Kazakhstan | 2603 | Asian |
| KSH002 | Kazakhstan | 2769 | Asian |
| KSH003 | Kazakhstan | 2550 | Asian |
| KSH004 | Kazakhstan | 2550 | Asian |
| KSK002 | Kazakhstan | 2300 | Asian |
| KYZ001 | Kazakhstan | 2475 | Asian |
| KYZ002 | Kazakhstan | 2616 | Asian |
| KZL001 | Kazakhstan | 2585 | Asian |
| KZL003 | Kazakhstan | 2500 | Asian |

| | | | |
|---|---|---|---|
| KZL004 | Kazakhstan | 3318 | Asian |
| MMR001 | Kazakhstan | 2500 | Asian |
| NUL001 | Kazakhstan | 2584 | Asian |
| NUR002 | Kazakhstan | 2596 | Asian |
| SBL001 | Kazakhstan | 2350 | Asian |
| SGZ001 | Kazakhstan | 2226 | Asian |
| SGZ002 | Kazakhstan | 2350 | Asian |
| SRK001 | Kazakhstan | 2400 | Asian |
| TAL003 | Kazakhstan | 2550 | Asian |
| TAL004 | Kazakhstan | 2619 | Asian |
| TAL005 | Kazakhstan | 2618 | Asian |
| WAR001 | Kazakhstan | 2537 | Asian |
| NE20 | China | 33592 | Asian |
| NE56 | China | 19427 | Asian |
| NE34 | China | 14521 | Asian |
| NE5 | China | 14174 | Asian |
| NE1 | China | 12650 | Asian |
| NE8 | China | 11263 | Asian |
| NE57 | China | 11026 | Asian |
| NE36 | China | 10673 | Asian |
| NE3 | China | 10628 | Asian |
| NE4 | China | 10557 | Asian |
| NE30 | China | 9896 | Asian |
| NE45 | China | 9241 | Asian |
| NE44 | China | 9232 | Asian |
| NE35 | China | 8990 | Asian |
| NE16 | China | 8564 | Asian |
| NE49 | China | 8368 | Asian |
| NE39 | China | 8196 | Asian |

| | | | |
|---|---|---|---|
| NE58 | China | 7308 | Asian |
| NE18 | China | 7062 | Asian |
| NE19 | China | 7039 | Asian |
| NE29 | China | 6877 | Asian |
| NE22 | China | 6838 | Asian |
| NE9 | China | 6331 | Asian |
| NE2 | China | 6321 | Asian |
| NE61 | China | 3421 | Asian |
| A_Mbuti-5.DG | Congo | 0 | Sub-Saharan African |
| A_Yoruba-4.DG | Nigeria | 0 | Sub-Saharan African |
| A_Dinka-4.DG | Sudan | 0 | Sub-Saharan African |
| A_Mandenka-4.DG | Senegal | 0 | Sub-Saharan African |
| A_Ju_hoan_North-5.DG | Namibia | 0 | Sub-Saharan African |
| S_Luo-2.DG | Kenya | 0 | Sub-Saharan African |
| S_Igbo-2.DG | Nigeria | 0 | Sub-Saharan African |
| S_Mbuti-3.DG | Congo | 0 | Sub-Saharan African |
| S_Biaka-2.DG | Central African Republic | 0 | Sub-Saharan African |
| S_Mandenka-1.DG | Senegal | 0 | Sub-Saharan African |
| S_Ju_hoan_North-2.DG | Namibia | 0 | Sub-Saharan African |

| | | | |
|---|---|---|---|
| S_Yoruba-2.DG | Nigeria | 0 | Sub-Saharan African |
| S_Mandenka-2.DG | Senegal | 0 | Sub-Saharan African |
| S_Kongo-2.DG | Congo | 0 | Sub-Saharan African |
| T_Mada-1.DG | Cameroon | 0 | Sub-Saharan African |
| T_Bakola-1.DG | Cameroon | 0 | Sub-Saharan African |
| T_Mursi-1.DG | Ethiopia | 0 | Sub-Saharan African |
| T_Kaba-2.DG | Central African Republic | 0 | Sub-Saharan African |
| T_Aari-2.DG | Ethiopia | 0 | Sub-Saharan African |
| S_Igbo-1.DG | Nigeria | 0 | Sub-Saharan African |
| T_Hadza-2.DG | Tanzania | 0 | Sub-Saharan African |
| T_Tikar_South-1.DG | Cameroon | 0 | Sub-Saharan African |
| T_Ogiek-1.DG | Kenya | 0 | Sub-Saharan African |
| S_Dinka-2.DG | Sudan | 0 | Sub-Saharan African |
| S_Esan-1.DG | Nigeria | 0 | Sub-Saharan African |
| T_Kikuyu-2.DG | Kenya | 0 | Sub-Saharan African |
| T_Sengwer-2.DG | Kenya | 0 | Sub-Saharan African |

| | | | |
|---|---|---|---|
| T_Sandawe-1.DG | Tanzania | 0 | Sub-Saharan African |
| T_Amhara-1.DG | Ethiopia | 0 | Sub-Saharan African |
| T_Laka-1.DG | Chad | 0 | Sub-Saharan African |
| B_Yoruba-3.DG | Nigeria | 0 | Sub-Saharan African |
| B_Mandenka-3.DG | Senegal | 0 | Sub-Saharan African |
| B_Mbuti-4.DG | Congo | 0 | Sub-Saharan African |
| B_Ju_hoan_North-4.DG | Namibia | 0 | Sub-Saharan African |
| S_Ju_hoan_North-1.DG | Namibia | 0 | Sub-Saharan African |
| S_Luhya-2.DG | Kenya | 0 | Sub-Saharan African |
| T_Bedzan-1.DG | Cameroon | 0 | Sub-Saharan African |
| T_Aari-1.DG | Ethiopia | 0 | Sub-Saharan African |
| T_Kaba-1.DG | Central African Republic | 0 | Sub-Saharan African |
| S_Dinka-1.DG | Sudan | 0 | Sub-Saharan African |
| T_Elmolo-2.DG | Kenya | 0 | Sub-Saharan African |
| T_Baka-1.DG | Cameroon | 0 | Sub-Saharan African |
| T_Ogiek-2.DG | Kenya | 0 | Sub-Saharan African |

| | | | |
|---|---|---|---|
| T_Tikar_South-2.DG | Cameroon | 0 | Sub-Saharan African |
| T_Mursi-2.DG | Ethiopia | 0 | Sub-Saharan African |
| T_Bulala-2.DG | Chad | 0 | Sub-Saharan African |
| T_Agaw-2.DG | Ethiopia | 0 | Sub-Saharan African |
| S_Masai-1.DG | Kenya | 0 | Sub-Saharan African |
| T_Bakola-2.DG | Cameroon | 0 | Sub-Saharan African |
| S_Luo-1.DG | Kenya | 0 | Sub-Saharan African |
| T_Fulani-1.DG | Cameroon | 0 | Sub-Saharan African |
| S_Esan-2.DG | Nigeria | 0 | Sub-Saharan African |
| T_Ngumba-1.DG | Cameroon | 0 | Sub-Saharan African |
| T_Rendille-1.DG | Kenya | 0 | Sub-Saharan African |
| T_Iraqw-2.DG | Tanzania | 0 | Sub-Saharan African |
| S_Lemande-1.DG | Cameroon | 0 | Sub-Saharan African |
| S_Lemande-2.DG | Cameroon | 0 | Sub-Saharan African |
| S_BantuKenya-1.DG | Kenya | 0 | Sub-Saharan African |
| S_Khomani_San-1.DG | South Africa | 0 | Sub-Saharan African |

| | | | |
|---|---|---|---|
| S_Mende-1.DG | Sierra Leone | 0 | Sub-Saharan African |
| S_Gambian-1.DG | Gambia | 0 | Sub-Saharan African |
| S_Saharawi-1.DG | Western Sahara (Morocco) | 0 | Sub-Saharan African |
| S_Ju_hoan_North-3.DG | Namibia | 0 | Sub-Saharan African |
| S_Somali-1.DG | Kenya | 0 | Sub-Saharan African |
| S_Gambian-2.DG | Gambia | 0 | Sub-Saharan African |
| S_Mende-2.DG | Sierra Leone | 0 | Sub-Saharan African |
| S_Luhya-1.DG | Kenya | 0 | Sub-Saharan African |
| S_Mozabite-1.DG | Algeria | 0 | Sub-Saharan African |
| S_Biaka-1.DG | Central African Republic | 0 | Sub-Saharan African |
| T_Mada-2.DG | Cameroon | 0 | Sub-Saharan African |
| T_Iraqw-1.DG | Tanzania | 0 | Sub-Saharan African |
| T_Rendille-2.DG | Kenya | 0 | Sub-Saharan African |
| T_Ngumba-2.DG | Cameroon | 0 | Sub-Saharan African |
| S_BantuKenya-2.DG | Kenya | 0 | Sub-Saharan African |
| S_Mbuti-2.DG | Congo | 0 | Sub-Saharan African |

| | | | |
|---|---|---|---|
| T_Bedzan-2.DG | Cameroon | 0 | Sub-Saharan African |
| T_Laka-2.DG | Chad | 0 | Sub-Saharan African |
| T_Baka-2.DG | Cameroon | 0 | Sub-Saharan African |
| S_Mozabite-2.DG | Algeria | 0 | Sub-Saharan African |
| S_Yoruba-1.DG | Nigeria | 0 | Sub-Saharan African |
| T_Kikuyu-1.DG | Kenya | 0 | Sub-Saharan African |
| T_Agaw-1.DG | Ethiopia | 0 | Sub-Saharan African |
| T_Bulala-1.DG | Chad | 0 | Sub-Saharan African |
| S_Masai-2.DG | Kenya | 0 | Sub-Saharan African |
| S_Khomani_San-2.DG | South Africa | 0 | Sub-Saharan African |
| S_Mbuti-1.DG | Congo | 0 | Sub-Saharan African |
| T_Elmolo-1.DG | Kenya | 0 | Sub-Saharan African |
| T_Sengwer-1.DG | Kenya | 0 | Sub-Saharan African |
| T_Amhara-2.DG | Ethiopia | 0 | Sub-Saharan African |
| S_Saharawi-2.DG | Western Sahara (Morocco) | 0 | Sub-Saharan African |
| T_Fulani-2.DG | Cameroon | 0 | Sub-Saharan African |

| | | | |
|---|---|---|---|
| B_Dinka-3.DG | Sudan | 0 | Sub-Saharan African |
| I0589.SG | Tanzania | 1331 | Sub-Saharan African |
| I1048.SG | Tanzania | 1357 | Sub-Saharan African |
| I2966.SG | Malawi | 8895 | Sub-Saharan African |
| I2967.SG | Malawi | 8018 | Sub-Saharan African |
| I2298_published | Tanzania | 590 | Sub-Saharan African |
| I1048_published | Tanzania | 1357 | Sub-Saharan African |
| mota.SG | Ethiopia | 4470 | Sub-Saharan African |
| I9133.SG | South Africa | 1910 | Sub-Saharan African |
| I9134.SG | South Africa | 1183 | Sub-Saharan African |
| I9028.SG | South Africa | 2245 | Sub-Saharan African |
| I0595 | Kenya | 434 | Sub-Saharan African |
| HG02461.SG | Gambia | 0 | Sub-Saharan African |
| HG02462.SG | Gambia | 0 | Sub-Saharan African |
| HG02464.SG | Gambia | 0 | Sub-Saharan African |
| HG02465.SG | Gambia | 0 | Sub-Saharan African |

| | | | |
|---|---|---|---|
| HG02561.SG | Gambia | 0 | Sub-Saharan African |
| HG02562.SG | Gambia | 0 | Sub-Saharan African |
| HG02568.SG | Gambia | 0 | Sub-Saharan African |
| HG02570.SG | Gambia | 0 | Sub-Saharan African |
| HG02571.SG | Gambia | 0 | Sub-Saharan African |
| HG02573.SG | Gambia | 0 | Sub-Saharan African |
| HG02574.SG | Gambia | 0 | Sub-Saharan African |
| HG02582.SG | Gambia | 0 | Sub-Saharan African |
| HG02583.SG | Gambia | 0 | Sub-Saharan African |
| HG02585.SG | Gambia | 0 | Sub-Saharan African |
| HG02586.SG | Gambia | 0 | Sub-Saharan African |
| HG02588.SG | Gambia | 0 | Sub-Saharan African |
| HG02589.SG | Gambia | 0 | Sub-Saharan African |
| HG02594.SG | Gambia | 0 | Sub-Saharan African |
| HG02595.SG | Gambia | 0 | Sub-Saharan African |
| HG02610.SG | Gambia | 0 | Sub-Saharan African |

| | | | |
|---|---|---|---|
| HG02611.SG | Gambia | 0 | Sub-Saharan African |
| HG02613.SG | Gambia | 0 | Sub-Saharan African |
| HG02614.SG | Gambia | 0 | Sub-Saharan African |
| HG02620.SG | Gambia | 0 | Sub-Saharan African |
| HG02621.SG | Gambia | 0 | Sub-Saharan African |
| HG02623.SG | Gambia | 0 | Sub-Saharan African |
| HG02624.SG | Gambia | 0 | Sub-Saharan African |
| HG02628.SG | Gambia | 0 | Sub-Saharan African |
| HG02629.SG | Gambia | 0 | Sub-Saharan African |
| HG02634.SG | Gambia | 0 | Sub-Saharan African |
| HG02635.SG | Gambia | 0 | Sub-Saharan African |
| HG02642.SG | Gambia | 0 | Sub-Saharan African |
| HG02643.SG | Gambia | 0 | Sub-Saharan African |
| HG02645.SG | Gambia | 0 | Sub-Saharan African |
| HG02646.SG | Gambia | 0 | Sub-Saharan African |
| HG02666.SG | Gambia | 0 | Sub-Saharan African |

| | | | |
|---|---|---|---|
| HG02667.SG | Gambia | 0 | Sub-Saharan African |
| HG02675.SG | Gambia | 0 | Sub-Saharan African |
| HG02676.SG | Gambia | 0 | Sub-Saharan African |
| HG02678.SG | Gambia | 0 | Sub-Saharan African |
| HG02679.SG | Gambia | 0 | Sub-Saharan African |
| HG02702.SG | Gambia | 0 | Sub-Saharan African |
| HG02703.SG | Gambia | 0 | Sub-Saharan African |
| HG02715.SG | Gambia | 0 | Sub-Saharan African |
| HG02716.SG | Gambia | 0 | Sub-Saharan African |
| HG02721.SG | Gambia | 0 | Sub-Saharan African |
| HG02722.SG | Gambia | 0 | Sub-Saharan African |
| HG02756.SG | Gambia | 0 | Sub-Saharan African |
| HG02757.SG | Gambia | 0 | Sub-Saharan African |
| HG02759.SG | Gambia | 0 | Sub-Saharan African |
| HG02760.SG | Gambia | 0 | Sub-Saharan African |
| HG02763.SG | Gambia | 0 | Sub-Saharan African |

| | | | |
|---|---|---|---|
| HG02768.SG | Gambia | 0 | Sub-Saharan African |
| HG02769.SG | Gambia | 0 | Sub-Saharan African |
| HG02771.SG | Gambia | 0 | Sub-Saharan African |
| HG02772.SG | Gambia | 0 | Sub-Saharan African |
| HG02798.SG | Gambia | 0 | Sub-Saharan African |
| HG02799.SG | Gambia | 0 | Sub-Saharan African |
| HG02804.SG | Gambia | 0 | Sub-Saharan African |
| HG02805.SG | Gambia | 0 | Sub-Saharan African |
| HG02807.SG | Gambia | 0 | Sub-Saharan African |
| HG02808.SG | Gambia | 0 | Sub-Saharan African |
| HG02810.SG | Gambia | 0 | Sub-Saharan African |
| HG02811.SG | Gambia | 0 | Sub-Saharan African |
| HG02813.SG | Gambia | 0 | Sub-Saharan African |
| HG02814.SG | Gambia | 0 | Sub-Saharan African |
| HG02816.SG | Gambia | 0 | Sub-Saharan African |
| HG02817.SG | Gambia | 0 | Sub-Saharan African |

| | | | |
|---|---|---|---|
| HG02819.SG | Gambia | 0 | Sub-Saharan African |
| HG02820.SG | Gambia | 0 | Sub-Saharan African |
| HG02836.SG | Gambia | 0 | Sub-Saharan African |
| HG02837.SG | Gambia | 0 | Sub-Saharan African |
| HG02839.SG | Gambia | 0 | Sub-Saharan African |
| HG02840.SG | Gambia | 0 | Sub-Saharan African |
| HG02851.SG | Gambia | 0 | Sub-Saharan African |
| HG02852.SG | Gambia | 0 | Sub-Saharan African |
| HG02854.SG | Gambia | 0 | Sub-Saharan African |
| HG02855.SG | Gambia | 0 | Sub-Saharan African |
| HG02860.SG | Gambia | 0 | Sub-Saharan African |
| HG02861.SG | Gambia | 0 | Sub-Saharan African |
| HG02870.SG | Gambia | 0 | Sub-Saharan African |
| HG02878.SG | Gambia | 0 | Sub-Saharan African |
| HG02879.SG | Gambia | 0 | Sub-Saharan African |
| HG02881.SG | Gambia | 0 | Sub-Saharan African |

| | | | |
|---|---|---|---|
| HG02882.SG | Gambia | 0 | Sub-Saharan African |
| HG02884.SG | Gambia | 0 | Sub-Saharan African |
| HG02885.SG | Gambia | 0 | Sub-Saharan African |
| HG02887.SG | Gambia | 0 | Sub-Saharan African |
| HG02888.SG | Gambia | 0 | Sub-Saharan African |
| HG02890.SG | Gambia | 0 | Sub-Saharan African |
| HG02891.SG | Gambia | 0 | Sub-Saharan African |
| HG02895.SG | Gambia | 0 | Sub-Saharan African |
| HG02896.SG | Gambia | 0 | Sub-Saharan African |
| HG02922.SG | Nigeria | 0 | Sub-Saharan African |
| HG02923.SG | Nigeria | 0 | Sub-Saharan African |
| HG02938.SG | Nigeria | 0 | Sub-Saharan African |
| HG02941.SG | Nigeria | 0 | Sub-Saharan African |
| HG02943.SG | Nigeria | 0 | Sub-Saharan African |
| HG02944.SG | Nigeria | 0 | Sub-Saharan African |
| HG02946.SG | Nigeria | 0 | Sub-Saharan African |

| | | | |
|---|---|---|---|
| HG02947.SG | Nigeria | 0 | Sub-Saharan African |
| HG02952.SG | Nigeria | 0 | Sub-Saharan African |
| HG02953.SG | Nigeria | 0 | Sub-Saharan African |
| HG02968.SG | Nigeria | 0 | Sub-Saharan African |
| HG02970.SG | Nigeria | 0 | Sub-Saharan African |
| HG02971.SG | Nigeria | 0 | Sub-Saharan African |
| HG02973.SG | Nigeria | 0 | Sub-Saharan African |
| HG02974.SG | Nigeria | 0 | Sub-Saharan African |
| HG02976.SG | Nigeria | 0 | Sub-Saharan African |
| HG02977.SG | Nigeria | 0 | Sub-Saharan African |
| HG02979.SG | Nigeria | 0 | Sub-Saharan African |
| HG02981.SG | Nigeria | 0 | Sub-Saharan African |
| HG02982.SG | Gambia | 0 | Sub-Saharan African |
| HG02983.SG | Gambia | 0 | Sub-Saharan African |
| HG03024.SG | Gambia | 0 | Sub-Saharan African |
| HG03025.SG | Gambia | 0 | Sub-Saharan African |

| | | | |
|---|---|---|---|
| HG03027.SG | Gambia | 0 | Sub-Saharan African |
| HG03028.SG | Gambia | 0 | Sub-Saharan African |
| HG03039.SG | Gambia | 0 | Sub-Saharan African |
| HG03040.SG | Gambia | 0 | Sub-Saharan African |
| HG03045.SG | Gambia | 0 | Sub-Saharan African |
| HG03046.SG | Gambia | 0 | Sub-Saharan African |
| HG03048.SG | Gambia | 0 | Sub-Saharan African |
| HG03049.SG | Gambia | 0 | Sub-Saharan African |
| HG03052.SG | Sierra Leone | 0 | Sub-Saharan African |
| HG03054.SG | Sierra Leone | 0 | Sub-Saharan African |
| HG03055.SG | Sierra Leone | 0 | Sub-Saharan African |
| HG03057.SG | Sierra Leone | 0 | Sub-Saharan African |
| HG03058.SG | Sierra Leone | 0 | Sub-Saharan African |
| HG03060.SG | Sierra Leone | 0 | Sub-Saharan African |
| HG03061.SG | Sierra Leone | 0 | Sub-Saharan African |
| HG03063.SG | Sierra Leone | 0 | Sub-Saharan African |

| | | | |
|---|---|---|---|
| HG03064.SG | Sierra Leone | 0 | Sub-Saharan African |
| HG03066.SG | Sierra Leone | 0 | Sub-Saharan African |
| HG03069.SG | Sierra Leone | 0 | Sub-Saharan African |
| HG03072.SG | Sierra Leone | 0 | Sub-Saharan African |
| HG03073.SG | Sierra Leone | 0 | Sub-Saharan African |
| HG03074.SG | Sierra Leone | 0 | Sub-Saharan African |
| HG03077.SG | Sierra Leone | 0 | Sub-Saharan African |
| HG03078.SG | Sierra Leone | 0 | Sub-Saharan African |
| HG03079.SG | Sierra Leone | 0 | Sub-Saharan African |
| HG03081.SG | Sierra Leone | 0 | Sub-Saharan African |
| HG03082.SG | Sierra Leone | 0 | Sub-Saharan African |
| HG03084.SG | Sierra Leone | 0 | Sub-Saharan African |
| HG03085.SG | Sierra Leone | 0 | Sub-Saharan African |
| HG03086.SG | Sierra Leone | 0 | Sub-Saharan African |
| HG03088.SG | Sierra Leone | 0 | Sub-Saharan African |
| HG03091.SG | Sierra Leone | 0 | Sub-Saharan African |

| | | | |
|---|---|---|---|
| HG03095.SG | Sierra Leone | 0 | Sub-Saharan African |
| HG03096.SG | Sierra Leone | 0 | Sub-Saharan African |
| HG03097.SG | Sierra Leone | 0 | Sub-Saharan African |
| HG03099.SG | Nigeria | 0 | Sub-Saharan African |
| HG03100.SG | Nigeria | 0 | Sub-Saharan African |
| HG03103.SG | Nigeria | 0 | Sub-Saharan African |
| HG03105.SG | Nigeria | 0 | Sub-Saharan African |
| HG03108.SG | Nigeria | 0 | Sub-Saharan African |
| HG03109.SG | Nigeria | 0 | Sub-Saharan African |
| HG03111.SG | Nigeria | 0 | Sub-Saharan African |
| HG03112.SG | Nigeria | 0 | Sub-Saharan African |
| HG03114.SG | Nigeria | 0 | Sub-Saharan African |
| HG03115.SG | Nigeria | 0 | Sub-Saharan African |
| HG03117.SG | Nigeria | 0 | Sub-Saharan African |
| HG03118.SG | Nigeria | 0 | Sub-Saharan African |
| HG03120.SG | Nigeria | 0 | Sub-Saharan African |

| | | | |
|---|---|---|---|
| HG03121.SG | Nigeria | 0 | Sub-Saharan African |
| HG03123.SG | Nigeria | 0 | Sub-Saharan African |
| HG03124.SG | Nigeria | 0 | Sub-Saharan African |
| HG03126.SG | Nigeria | 0 | Sub-Saharan African |
| HG03127.SG | Nigeria | 0 | Sub-Saharan African |
| HG03129.SG | Nigeria | 0 | Sub-Saharan African |
| HG03130.SG | Nigeria | 0 | Sub-Saharan African |
| HG03132.SG | Nigeria | 0 | Sub-Saharan African |
| HG03133.SG | Nigeria | 0 | Sub-Saharan African |
| HG03135.SG | Nigeria | 0 | Sub-Saharan African |
| HG03136.SG | Nigeria | 0 | Sub-Saharan African |
| HG03139.SG | Nigeria | 0 | Sub-Saharan African |
| HG03157.SG | Nigeria | 0 | Sub-Saharan African |
| HG03159.SG | Nigeria | 0 | Sub-Saharan African |
| HG03160.SG | Nigeria | 0 | Sub-Saharan African |
| HG03162.SG | Nigeria | 0 | Sub-Saharan African |

| | | | |
|---|---|---|---|
| HG03163.SG | Nigeria | 0 | Sub-Saharan African |
| HG03166.SG | Nigeria | 0 | Sub-Saharan African |
| HG03168.SG | Nigeria | 0 | Sub-Saharan African |
| HG03169.SG | Nigeria | 0 | Sub-Saharan African |
| HG03172.SG | Nigeria | 0 | Sub-Saharan African |
| HG03175.SG | Nigeria | 0 | Sub-Saharan African |
| HG03189.SG | Nigeria | 0 | Sub-Saharan African |
| HG03190.SG | Nigeria | 0 | Sub-Saharan African |
| HG03193.SG | Nigeria | 0 | Sub-Saharan African |
| HG03195.SG | Nigeria | 0 | Sub-Saharan African |
| HG03196.SG | Nigeria | 0 | Sub-Saharan African |
| HG03198.SG | Nigeria | 0 | Sub-Saharan African |
| HG03199.SG | Nigeria | 0 | Sub-Saharan African |
| HG03202.SG | Nigeria | 0 | Sub-Saharan African |
| HG03209.SG | Sierra Leone | 0 | Sub-Saharan African |
| HG03212.SG | Sierra Leone | 0 | Sub-Saharan African |

| | | | |
|---|---|---|---|
| HG03224.SG | Sierra Leone | 0 | Sub-Saharan African |
| HG03225.SG | Sierra Leone | 0 | Sub-Saharan African |
| HG03240.SG | Gambia | 0 | Sub-Saharan African |
| HG03241.SG | Gambia | 0 | Sub-Saharan African |
| HG03246.SG | Gambia | 0 | Sub-Saharan African |
| HG03247.SG | Gambia | 0 | Sub-Saharan African |
| HG03258.SG | Gambia | 0 | Sub-Saharan African |
| HG03259.SG | Gambia | 0 | Sub-Saharan African |
| HG03265.SG | Nigeria | 0 | Sub-Saharan African |
| HG03267.SG | Nigeria | 0 | Sub-Saharan African |
| HG03268.SG | Nigeria | 0 | Sub-Saharan African |
| HG03270.SG | Nigeria | 0 | Sub-Saharan African |
| HG03271.SG | Nigeria | 0 | Sub-Saharan African |
| HG03279.SG | Nigeria | 0 | Sub-Saharan African |
| HG03280.SG | Nigeria | 0 | Sub-Saharan African |
| HG03291.SG | Nigeria | 0 | Sub-Saharan African |

| | | | |
|---|---|---|---|
| HG03294.SG | Nigeria | 0 | Sub-Saharan African |
| HG03295.SG | Nigeria | 0 | Sub-Saharan African |
| HG03297.SG | Nigeria | 0 | Sub-Saharan African |
| HG03298.SG | Nigeria | 0 | Sub-Saharan African |
| HG03300.SG | Nigeria | 0 | Sub-Saharan African |
| HG03301.SG | Nigeria | 0 | Sub-Saharan African |
| HG03303.SG | Nigeria | 0 | Sub-Saharan African |
| HG03304.SG | Nigeria | 0 | Sub-Saharan African |
| HG03311.SG | Nigeria | 0 | Sub-Saharan African |
| HG03313.SG | Nigeria | 0 | Sub-Saharan African |
| HG03342.SG | Nigeria | 0 | Sub-Saharan African |
| HG03343.SG | Nigeria | 0 | Sub-Saharan African |
| HG03351.SG | Nigeria | 0 | Sub-Saharan African |
| HG03352.SG | Nigeria | 0 | Sub-Saharan African |
| HG03354.SG | Nigeria | 0 | Sub-Saharan African |
| HG03363.SG | Nigeria | 0 | Sub-Saharan African |

| | | | |
|---|---|---|---|
| HG03366.SG | Nigeria | 0 | Sub-Saharan African |
| HG03367.SG | Nigeria | 0 | Sub-Saharan African |
| HG03369.SG | Nigeria | 0 | Sub-Saharan African |
| HG03370.SG | Nigeria | 0 | Sub-Saharan African |
| HG03372.SG | Nigeria | 0 | Sub-Saharan African |
| HG03376.SG | Sierra Leone | 0 | Sub-Saharan African |
| HG03378.SG | Sierra Leone | 0 | Sub-Saharan African |
| HG03380.SG | Sierra Leone | 0 | Sub-Saharan African |
| HG03382.SG | Sierra Leone | 0 | Sub-Saharan African |
| HG03385.SG | Sierra Leone | 0 | Sub-Saharan African |
| HG03388.SG | Sierra Leone | 0 | Sub-Saharan African |
| HG03391.SG | Sierra Leone | 0 | Sub-Saharan African |
| HG03394.SG | Sierra Leone | 0 | Sub-Saharan African |
| HG03397.SG | Sierra Leone | 0 | Sub-Saharan African |
| HG03401.SG | Sierra Leone | 0 | Sub-Saharan African |
| HG03410.SG | Sierra Leone | 0 | Sub-Saharan African |

| | | | |
|---|---|---|---|
| HG03419.SG | Sierra Leone | 0 | Sub-Saharan African |
| HG03428.SG | Sierra Leone | 0 | Sub-Saharan African |
| HG03432.SG | Sierra Leone | 0 | Sub-Saharan African |
| HG03433.SG | Sierra Leone | 0 | Sub-Saharan African |
| HG03436.SG | Sierra Leone | 0 | Sub-Saharan African |
| HG03437.SG | Sierra Leone | 0 | Sub-Saharan African |
| HG03439.SG | Sierra Leone | 0 | Sub-Saharan African |
| HG03442.SG | Sierra Leone | 0 | Sub-Saharan African |
| HG03445.SG | Sierra Leone | 0 | Sub-Saharan African |
| HG03446.SG | Sierra Leone | 0 | Sub-Saharan African |
| HG03449.SG | Sierra Leone | 0 | Sub-Saharan African |
| HG03451.SG | Sierra Leone | 0 | Sub-Saharan African |
| HG03452.SG | Sierra Leone | 0 | Sub-Saharan African |
| HG03455.SG | Sierra Leone | 0 | Sub-Saharan African |
| HG03457.SG | Sierra Leone | 0 | Sub-Saharan African |
| HG03458.SG | Sierra Leone | 0 | Sub-Saharan African |

| | | | |
|---|---|---|---|
| HG03460.SG | Sierra Leone | 0 | Sub-Saharan African |
| HG03461.SG | Sierra Leone | 0 | Sub-Saharan African |
| HG03464.SG | Sierra Leone | 0 | Sub-Saharan African |
| HG03469.SG | Sierra Leone | 0 | Sub-Saharan African |
| HG03470.SG | Sierra Leone | 0 | Sub-Saharan African |
| HG03472.SG | Sierra Leone | 0 | Sub-Saharan African |
| HG03473.SG | Sierra Leone | 0 | Sub-Saharan African |
| HG03476.SG | Sierra Leone | 0 | Sub-Saharan African |
| HG03478.SG | Sierra Leone | 0 | Sub-Saharan African |
| HG03479.SG | Sierra Leone | 0 | Sub-Saharan African |
| HG03484.SG | Sierra Leone | 0 | Sub-Saharan African |
| HG03485.SG | Sierra Leone | 0 | Sub-Saharan African |
| HG03499.SG | Nigeria | 0 | Sub-Saharan African |
| HG03511.SG | Nigeria | 0 | Sub-Saharan African |
| HG03514.SG | Nigeria | 0 | Sub-Saharan African |
| HG03515.SG | Nigeria | 0 | Sub-Saharan African |

| | | | |
|---|---|---|---|
| HG03517.SG | Nigeria | 0 | Sub-Saharan African |
| HG03518.SG | Nigeria | 0 | Sub-Saharan African |
| HG03520.SG | Nigeria | 0 | Sub-Saharan African |
| HG03521.SG | Nigeria | 0 | Sub-Saharan African |
| HG03538.SG | Gambia | 0 | Sub-Saharan African |
| HG03539.SG | Gambia | 0 | Sub-Saharan African |
| HG03547.SG | Sierra Leone | 0 | Sub-Saharan African |
| HG03548.SG | Sierra Leone | 0 | Sub-Saharan African |
| HG03556.SG | Sierra Leone | 0 | Sub-Saharan African |
| HG03557.SG | Sierra Leone | 0 | Sub-Saharan African |
| HG03558.SG | Sierra Leone | 0 | Sub-Saharan African |
| HG03559.SG | Sierra Leone | 0 | Sub-Saharan African |
| HG03563.SG | Sierra Leone | 0 | Sub-Saharan African |
| HG03565.SG | Sierra Leone | 0 | Sub-Saharan African |
| HG03567.SG | Sierra Leone | 0 | Sub-Saharan African |
| HG03571.SG | Sierra Leone | 0 | Sub-Saharan African |

| | | | |
|---|---|---|---|
| HG03572.SG | Sierra Leone | 0 | Sub-Saharan African |
| HG03575.SG | Sierra Leone | 0 | Sub-Saharan African |
| HG03577.SG | Sierra Leone | 0 | Sub-Saharan African |
| HG03578.SG | Sierra Leone | 0 | Sub-Saharan African |
| HG03583.SG | Sierra Leone | 0 | Sub-Saharan African |
| NA18486.SG | Nigeria | 0 | Sub-Saharan African |
| NA18488.SG | Nigeria | 0 | Sub-Saharan African |
| NA18489.SG | Nigeria | 0 | Sub-Saharan African |
| NA18498.SG | Nigeria | 0 | Sub-Saharan African |
| NA18499.SG | Nigeria | 0 | Sub-Saharan African |
| NA18501.SG | Nigeria | 0 | Sub-Saharan African |
| NA18502.SG | Nigeria | 0 | Sub-Saharan African |
| NA18504.SG | Nigeria | 0 | Sub-Saharan African |
| NA18505.SG | Nigeria | 0 | Sub-Saharan African |
| NA18507.SG | Nigeria | 0 | Sub-Saharan African |
| NA18508.SG | Nigeria | 0 | Sub-Saharan African |

| | | | |
|---|---|---|---|
| NA18510.SG | Nigeria | 0 | Sub-Saharan African |
| NA18511.SG | Nigeria | 0 | Sub-Saharan African |
| NA18516.SG | Nigeria | 0 | Sub-Saharan African |
| NA18517.SG | Nigeria | 0 | Sub-Saharan African |
| NA18519.SG | Nigeria | 0 | Sub-Saharan African |
| NA18520.SG | Nigeria | 0 | Sub-Saharan African |
| NA18522.SG | Nigeria | 0 | Sub-Saharan African |
| NA18523.SG | Nigeria | 0 | Sub-Saharan African |
| NA18853.SG | Nigeria | 0 | Sub-Saharan African |
| NA18856.SG | Nigeria | 0 | Sub-Saharan African |
| NA18858.SG | Nigeria | 0 | Sub-Saharan African |
| NA18861.SG | Nigeria | 0 | Sub-Saharan African |
| NA18864.SG | Nigeria | 0 | Sub-Saharan African |
| NA18865.SG | Nigeria | 0 | Sub-Saharan African |
| NA18867.SG | Nigeria | 0 | Sub-Saharan African |
| NA18868.SG | Nigeria | 0 | Sub-Saharan African |

| | | | |
|---|---|---|---|
| NA18870.SG | Nigeria | 0 | Sub-Saharan African |
| NA18871.SG | Nigeria | 0 | Sub-Saharan African |
| NA18873.SG | Nigeria | 0 | Sub-Saharan African |
| NA18874.SG | Nigeria | 0 | Sub-Saharan African |
| NA18876.SG | Nigeria | 0 | Sub-Saharan African |
| NA18877.SG | Nigeria | 0 | Sub-Saharan African |
| NA18878.SG | Nigeria | 0 | Sub-Saharan African |
| NA18879.SG | Nigeria | 0 | Sub-Saharan African |
| NA18881.SG | Nigeria | 0 | Sub-Saharan African |
| NA18907.SG | Nigeria | 0 | Sub-Saharan African |
| NA18908.SG | Nigeria | 0 | Sub-Saharan African |
| NA18909.SG | Nigeria | 0 | Sub-Saharan African |
| NA18910.SG | Nigeria | 0 | Sub-Saharan African |
| NA18912.SG | Nigeria | 0 | Sub-Saharan African |
| NA18915.SG | Nigeria | 0 | Sub-Saharan African |
| NA18916.SG | Nigeria | 0 | Sub-Saharan African |

| | | | |
|---|---|---|---|
| NA18917.SG | Nigeria | 0 | Sub-Saharan African |
| NA18923.SG | Nigeria | 0 | Sub-Saharan African |
| NA18924.SG | Nigeria | 0 | Sub-Saharan African |
| NA18933.SG | Nigeria | 0 | Sub-Saharan African |
| NA18934.SG | Nigeria | 0 | Sub-Saharan African |
| NA19017.SG | Kenya | 0 | Sub-Saharan African |
| NA19019.SG | Kenya | 0 | Sub-Saharan African |
| NA19020.SG | Kenya | 0 | Sub-Saharan African |
| NA19023.SG | Kenya | 0 | Sub-Saharan African |
| NA19024.SG | Kenya | 0 | Sub-Saharan African |
| NA19025.SG | Kenya | 0 | Sub-Saharan African |
| NA19026.SG | Kenya | 0 | Sub-Saharan African |
| NA19027.SG | Kenya | 0 | Sub-Saharan African |
| NA19028.SG | Kenya | 0 | Sub-Saharan African |
| NA19030.SG | Kenya | 0 | Sub-Saharan African |
| NA19031.SG | Kenya | 0 | Sub-Saharan African |

| | | | |
|---|---|---|---|
| NA19035.SG | Kenya | 0 | Sub-Saharan African |
| NA19036.SG | Kenya | 0 | Sub-Saharan African |
| NA19037.SG | Kenya | 0 | Sub-Saharan African |
| NA19038.SG | Kenya | 0 | Sub-Saharan African |
| NA19041.SG | Kenya | 0 | Sub-Saharan African |
| NA19042.SG | Kenya | 0 | Sub-Saharan African |
| NA19043.SG | Kenya | 0 | Sub-Saharan African |
| NA19092.SG | Nigeria | 0 | Sub-Saharan African |
| NA19093.SG | Nigeria | 0 | Sub-Saharan African |
| NA19095.SG | Nigeria | 0 | Sub-Saharan African |
| NA19096.SG | Nigeria | 0 | Sub-Saharan African |
| NA19098.SG | Nigeria | 0 | Sub-Saharan African |
| NA19099.SG | Nigeria | 0 | Sub-Saharan African |
| NA19102.SG | Nigeria | 0 | Sub-Saharan African |
| NA19107.SG | Nigeria | 0 | Sub-Saharan African |
| NA19108.SG | Nigeria | 0 | Sub-Saharan African |

| | | | |
|---|---|---|---|
| NA19113.SG | Nigeria | 0 | Sub-Saharan African |
| NA19114.SG | Nigeria | 0 | Sub-Saharan African |
| NA19116.SG | Nigeria | 0 | Sub-Saharan African |
| NA19117.SG | Nigeria | 0 | Sub-Saharan African |
| NA19118.SG | Nigeria | 0 | Sub-Saharan African |
| NA19119.SG | Nigeria | 0 | Sub-Saharan African |
| NA19121.SG | Nigeria | 0 | Sub-Saharan African |
| NA19129.SG | Nigeria | 0 | Sub-Saharan African |
| NA19130.SG | Nigeria | 0 | Sub-Saharan African |
| NA19131.SG | Nigeria | 0 | Sub-Saharan African |
| NA19137.SG | Nigeria | 0 | Sub-Saharan African |
| NA19138.SG | Nigeria | 0 | Sub-Saharan African |
| NA19141.SG | Nigeria | 0 | Sub-Saharan African |
| NA19143.SG | Nigeria | 0 | Sub-Saharan African |
| NA19144.SG | Nigeria | 0 | Sub-Saharan African |
| NA19146.SG | Nigeria | 0 | Sub-Saharan African |

| | | | |
|---|---|---|---|
| NA19147.SG | Nigeria | 0 | Sub-Saharan African |
| NA19149.SG | Nigeria | 0 | Sub-Saharan African |
| NA19152.SG | Nigeria | 0 | Sub-Saharan African |
| NA19153.SG | Nigeria | 0 | Sub-Saharan African |
| NA19159.SG | Nigeria | 0 | Sub-Saharan African |
| NA19160.SG | Nigeria | 0 | Sub-Saharan African |
| NA19171.SG | Nigeria | 0 | Sub-Saharan African |
| NA19172.SG | Nigeria | 0 | Sub-Saharan African |
| NA19175.SG | Nigeria | 0 | Sub-Saharan African |
| NA19184.SG | Nigeria | 0 | Sub-Saharan African |
| NA19185.SG | Nigeria | 0 | Sub-Saharan African |
| NA19189.SG | Nigeria | 0 | Sub-Saharan African |
| NA19190.SG | Nigeria | 0 | Sub-Saharan African |
| NA19197.SG | Nigeria | 0 | Sub-Saharan African |
| NA19198.SG | Nigeria | 0 | Sub-Saharan African |
| NA19200.SG | Nigeria | 0 | Sub-Saharan African |

| | | | |
|---|---|---|---|
| NA19201.SG | Nigeria | 0 | Sub-Saharan African |
| NA19204.SG | Nigeria | 0 | Sub-Saharan African |
| NA19206.SG | Nigeria | 0 | Sub-Saharan African |
| NA19207.SG | Nigeria | 0 | Sub-Saharan African |
| NA19209.SG | Nigeria | 0 | Sub-Saharan African |
| NA19210.SG | Nigeria | 0 | Sub-Saharan African |
| NA19213.SG | Nigeria | 0 | Sub-Saharan African |
| NA19214.SG | Nigeria | 0 | Sub-Saharan African |
| NA19222.SG | Nigeria | 0 | Sub-Saharan African |
| NA19223.SG | Nigeria | 0 | Sub-Saharan African |
| NA19225.SG | Nigeria | 0 | Sub-Saharan African |
| NA19235.SG | Nigeria | 0 | Sub-Saharan African |
| NA19236.SG | Nigeria | 0 | Sub-Saharan African |
| NA19238.SG | Nigeria | 0 | Sub-Saharan African |
| NA19239.SG | Nigeria | 0 | Sub-Saharan African |
| NA19240.SG | Nigeria | 0 | Sub-Saharan African |

| | | | |
|---|---|---|---|
| NA19247.SG | Nigeria | 0 | Sub-Saharan African |
| NA19248.SG | Nigeria | 0 | Sub-Saharan African |
| NA19256.SG | Nigeria | 0 | Sub-Saharan African |
| NA19257.SG | Nigeria | 0 | Sub-Saharan African |
| NA19307.SG | Kenya | 0 | Sub-Saharan African |
| NA19308.SG | Kenya | 0 | Sub-Saharan African |
| NA19309.SG | Kenya | 0 | Sub-Saharan African |
| NA19310.SG | Kenya | 0 | Sub-Saharan African |
| NA19311.SG | Kenya | 0 | Sub-Saharan African |
| NA19312.SG | Kenya | 0 | Sub-Saharan African |
| NA19313.SG | Kenya | 0 | Sub-Saharan African |
| NA19314.SG | Kenya | 0 | Sub-Saharan African |
| NA19315.SG | Kenya | 0 | Sub-Saharan African |
| NA19316.SG | Kenya | 0 | Sub-Saharan African |
| NA19317.SG | Kenya | 0 | Sub-Saharan African |
| NA19318.SG | Kenya | 0 | Sub-Saharan African |

| | | | |
|---|---|---|---|
| NA19319.SG | Kenya | 0 | Sub-Saharan African |
| NA19320.SG | Kenya | 0 | Sub-Saharan African |
| NA19321.SG | Kenya | 0 | Sub-Saharan African |
| NA19323.SG | Kenya | 0 | Sub-Saharan African |
| NA19324.SG | Kenya | 0 | Sub-Saharan African |
| NA19327.SG | Kenya | 0 | Sub-Saharan African |
| NA19328.SG | Kenya | 0 | Sub-Saharan African |
| NA19331.SG | Kenya | 0 | Sub-Saharan African |
| NA19332.SG | Kenya | 0 | Sub-Saharan African |
| NA19334.SG | Kenya | 0 | Sub-Saharan African |
| NA19338.SG | Kenya | 0 | Sub-Saharan African |
| NA19346.SG | Kenya | 0 | Sub-Saharan African |
| NA19347.SG | Kenya | 0 | Sub-Saharan African |
| NA19350.SG | Kenya | 0 | Sub-Saharan African |
| NA19351.SG | Kenya | 0 | Sub-Saharan African |
| NA19355.SG | Kenya | 0 | Sub-Saharan African |

| | | | |
|---|---|---|---|
| NA19360.SG | Kenya | 0 | Sub-Saharan African |
| NA19372.SG | Kenya | 0 | Sub-Saharan African |
| NA19374.SG | Kenya | 0 | Sub-Saharan African |
| NA19375.SG | Kenya | 0 | Sub-Saharan African |
| NA19376.SG | Kenya | 0 | Sub-Saharan African |
| NA19377.SG | Kenya | 0 | Sub-Saharan African |
| NA19378.SG | Kenya | 0 | Sub-Saharan African |
| NA19379.SG | Kenya | 0 | Sub-Saharan African |
| NA19380.SG | Kenya | 0 | Sub-Saharan African |
| NA19383.SG | Kenya | 0 | Sub-Saharan African |
| NA19384.SG | Kenya | 0 | Sub-Saharan African |
| NA19385.SG | Kenya | 0 | Sub-Saharan African |
| NA19390.SG | Kenya | 0 | Sub-Saharan African |
| NA19391.SG | Kenya | 0 | Sub-Saharan African |
| NA19393.SG | Kenya | 0 | Sub-Saharan African |
| NA19394.SG | Kenya | 0 | Sub-Saharan African |

| | | | |
|---|---|---|---|
| NA19395.SG | Kenya | 0 | Sub-Saharan African |
| NA19397.SG | Kenya | 0 | Sub-Saharan African |
| NA19399.SG | Kenya | 0 | Sub-Saharan African |
| NA19401.SG | Kenya | 0 | Sub-Saharan African |
| NA19403.SG | Kenya | 0 | Sub-Saharan African |
| NA19404.SG | Kenya | 0 | Sub-Saharan African |
| NA19428.SG | Kenya | 0 | Sub-Saharan African |
| NA19429.SG | Kenya | 0 | Sub-Saharan African |
| NA19430.SG | Kenya | 0 | Sub-Saharan African |
| NA19431.SG | Kenya | 0 | Sub-Saharan African |
| NA19434.SG | Kenya | 0 | Sub-Saharan African |
| NA19435.SG | Kenya | 0 | Sub-Saharan African |
| NA19436.SG | Kenya | 0 | Sub-Saharan African |
| NA19437.SG | Kenya | 0 | Sub-Saharan African |
| NA19438.SG | Kenya | 0 | Sub-Saharan African |
| NA19439.SG | Kenya | 0 | Sub-Saharan African |

| | | | |
|---|---|---|---|
| NA19440.SG | Kenya | 0 | Sub-Saharan African |
| NA19443.SG | Kenya | 0 | Sub-Saharan African |
| NA19445.SG | Kenya | 0 | Sub-Saharan African |
| NA19446.SG | Kenya | 0 | Sub-Saharan African |
| NA19448.SG | Kenya | 0 | Sub-Saharan African |
| NA19449.SG | Kenya | 0 | Sub-Saharan African |
| NA19451.SG | Kenya | 0 | Sub-Saharan African |
| NA19452.SG | Kenya | 0 | Sub-Saharan African |
| NA19454.SG | Kenya | 0 | Sub-Saharan African |
| NA19455.SG | Kenya | 0 | Sub-Saharan African |
| NA19456.SG | Kenya | 0 | Sub-Saharan African |
| NA19457.SG | Kenya | 0 | Sub-Saharan African |
| NA19461.SG | Kenya | 0 | Sub-Saharan African |
| NA19462.SG | Kenya | 0 | Sub-Saharan African |
| NA19463.SG | Kenya | 0 | Sub-Saharan African |
| NA19466.SG | Kenya | 0 | Sub-Saharan African |

| | | | |
|---|---|---|---|
| NA19467.SG | Kenya | 0 | Sub-Saharan African |
| NA19468.SG | Kenya | 0 | Sub-Saharan African |
| NA19471.SG | Kenya | 0 | Sub-Saharan African |
| NA19472.SG | Kenya | 0 | Sub-Saharan African |
| NA19473.SG | Kenya | 0 | Sub-Saharan African |
| NA19474.SG | Kenya | 0 | Sub-Saharan African |
| NA19475.SG | Kenya | 0 | Sub-Saharan African |
| I0589_all | Tanzania | 1331 | Sub-Saharan African |
| I3726_all | Tanzania | 3023 | Sub-Saharan African |
| JK2134 | Egypt | 2609 | Sub-Saharan African |
| JK2888 | Egypt | 1997 | Sub-Saharan African |
| JK2911 | Egypt | 2602 | Sub-Saharan African |
| I2967_published | Malawi | 8018 | Sub-Saharan African |
| I3726 | Tanzania | 3023 | Sub-Saharan African |
| baa001.SG | South Africa | 1888 | Sub-Saharan African |
| bab001.SG | South Africa | 2037 | Sub-Saharan African |

| | | | |
|---|---|---|---|
| cha001.SG | South Africa | 355 | Sub-Saharan African |
| doo001.SG | South Africa | 2039 | Sub-Saharan African |
| ela001.SG | South Africa | 491 | Sub-Saharan African |
| mfo001.SG | South Africa | 390 | Sub-Saharan African |
| new001.SG | South Africa | 435 | Sub-Saharan African |
| gun002.SG | Canary Islands | 851 | Sub-Saharan African |
| gun005.SG | Canary Islands | 992 | Sub-Saharan African |
| gun008.SG | Canary Islands | 1014 | Sub-Saharan African |
| gun011.SG | Canary Islands | 1137 | Sub-Saharan African |
| gun012.SG | Canary Islands | 1324 | Sub-Saharan African |
| I2966_all | Malawi | 8895 | Sub-Saharan African |
| I0589 | Tanzania | 1331 | Sub-Saharan African |
| I2966 | Malawi | 8895 | Sub-Saharan African |
| TAF009 | Morocco | 14221 | Sub-Saharan African |
| TAF010 | Morocco | 14649 | Sub-Saharan African |
| TAF011 | Morocco | 14450 | Sub-Saharan African |

| | | | |
|---|---|---|---|
| TAF012 | Morocco | 14455 | Sub-Saharan African |
| TAF013 | Morocco | 14500 | Sub-Saharan African |
| TAF014 | Morocco | 14500 | Sub-Saharan African |
| TAF015 | Morocco | 14500 | Sub-Saharan African |
| I10719 | Kenya | 1969 | Sub-Saharan African |
| I8758 | Kenya | 2591 | Sub-Saharan African |
| I8802 | Kenya | 1072 | Sub-Saharan African |
| I8804 | Kenya | 2576 | Sub-Saharan African |
| I8805 | Kenya | 1783 | Sub-Saharan African |
| I8808 | Kenya | 1793 | Sub-Saharan African |
| I8809 | Kenya | 2967 | Sub-Saharan African |
| I8814 | Kenya | 2419 | Sub-Saharan African |
| I8820_published | Kenya | 2781 | Sub-Saharan African |
| I8830_published | Kenya | 2340 | Sub-Saharan African |
| I8874 | Kenya | 3287 | Sub-Saharan African |
| I8892_published | Kenya | 1096 | Sub-Saharan African |

| | | | |
|---|---|---|---|
| I8918 | Kenya | 2232 | Sub-Saharan African |
| I8919 | Kenya | 2250 | Sub-Saharan African |
| I8920_published | Kenya | 2338 | Sub-Saharan African |
| I8922 | Kenya | 2050 | Sub-Saharan African |
| I2967_all | Malawi | 8018 | Sub-Saharan African |
| HGDP01201.SDG | Senegal | 0 | Sub-Saharan African |
| HGDP00479.SDG | Central African Republic | 0 | Sub-Saharan African |
| HGDP01408.SDG | Kenya | 0 | Sub-Saharan African |
| HGDP00471.SDG | Congo | 0 | Sub-Saharan African |
| HGDP01411.SDG | Kenya | 0 | Sub-Saharan African |
| HGDP00937.SDG | Nigeria | 0 | Sub-Saharan African |
| HGDP01094.SDG | Central African Republic | 0 | Sub-Saharan African |
| HGDP00469.SDG | Central African Republic | 0 | Sub-Saharan African |
| HGDP00475.SDG | Central African Republic | 0 | Sub-Saharan African |
| HGDP00473.SDG | Central African Republic | 0 | Sub-Saharan African |
| HGDP01405.SDG | Kenya | 0 | Sub-Saharan African |

| | | | |
|---|---|---|---|
| HGDP00470.SDG | Central African Republic | 0 | Sub-Saharan African |
| HGDP00458.SDG | Central African Republic | 0 | Sub-Saharan African |
| HGDP00904.SDG | Senegal | 0 | Sub-Saharan African |
| HGDP01200.SDG | Senegal | 0 | Sub-Saharan African |
| HGDP00452.SDG | Central African Republic | 0 | Sub-Saharan African |
| HGDP00459.SDG | Central African Republic | 0 | Sub-Saharan African |
| HGDP00914.SDG | Senegal | 0 | Sub-Saharan African |
| HGDP00942.SDG | Nigeria | 0 | Sub-Saharan African |
| HGDP01031.SDG | Angola | 0 | Sub-Saharan African |
| HGDP00944.SDG | Nigeria | 0 | Sub-Saharan African |
| HGDP01406.SDG | Kenya | 0 | Sub-Saharan African |
| HGDP00907.SDG | Senegal | 0 | Sub-Saharan African |
| HGDP01202.SDG | Senegal | 0 | Sub-Saharan African |
| HGDP00908.SDG | Senegal | 0 | Sub-Saharan African |
| HGDP00940.SDG | Nigeria | 0 | Sub-Saharan African |
| HGDP00911.SDG | Senegal | 0 | Sub-Saharan African |

| | | | |
|---|---|---|---|
| HGDP00464.SDG | Central African Republic | 0 | Sub-Saharan African |
| HGDP00929.SDG | Nigeria | 0 | Sub-Saharan African |
| HGDP00986.SDG | Central African Republic | 0 | Sub-Saharan African |
| HGDP00941.SDG | Nigeria | 0 | Sub-Saharan African |
| HGDP00934.SDG | Nigeria | 0 | Sub-Saharan African |
| HGDP00462.SDG | Congo | 0 | Sub-Saharan African |
| HGDP00453.SDG | Central African Republic | 0 | Sub-Saharan African |
| HGDP00935.SDG | Nigeria | 0 | Sub-Saharan African |
| HGDP00943.SDG | Nigeria | 0 | Sub-Saharan African |
| HGDP01412.SDG | Kenya | 0 | Sub-Saharan African |
| HGDP00994.SDG | Lesotho | 0 | Sub-Saharan African |
| HGDP01283.SDG | Senegal | 0 | Sub-Saharan African |
| HGDP00993.SDG | South Africa | 0 | Sub-Saharan African |
| HGDP00918.SDG | Senegal | 0 | Sub-Saharan African |
| HGDP00985.SDG | Central African Republic | 0 | Sub-Saharan African |
| HGDP00910.SDG | Senegal | 0 | Sub-Saharan African |

| | | | |
|---|---|---|---|
| HGDP00472.SDG | Central African Republic | 0 | Sub-Saharan African |
| HGDP00450.SDG | Congo | 0 | Sub-Saharan African |
| HGDP00905.SDG | Senegal | 0 | Sub-Saharan African |
| HGDP01285.SDG | Senegal | 0 | Sub-Saharan African |
| HGDP00463.SDG | Congo | 0 | Sub-Saharan African |
| HGDP01257.SDG | Algeria | 0 | Sub-Saharan African |
| HGDP00931.SDG | Nigeria | 0 | Sub-Saharan African |
| HGDP00466.SDG | Central African Republic | 0 | Sub-Saharan African |
| HGDP00467.SDG | Congo | 0 | Sub-Saharan African |
| HGDP00933.SDG | Nigeria | 0 | Sub-Saharan African |
| HGDP01086.SDG | Central African Republic | 0 | Sub-Saharan African |
| HGDP00924.SDG | Nigeria | 0 | Sub-Saharan African |
| HGDP00920.SDG | Nigeria | 0 | Sub-Saharan African |
| HGDP00925.SDG | Nigeria | 0 | Sub-Saharan African |
| HGDP00938.SDG | Nigeria | 0 | Sub-Saharan African |
| HGDP00478.SDG | Congo | 0 | Sub-Saharan African |

| | | | |
|---|---|---|---|
| HGDP00939.SDG | Nigeria | 0 | Sub-Saharan African |
| HGDP01256.SDG | Algeria | 0 | Sub-Saharan African |
| HGDP01029.SDG | Namibia | 0 | Sub-Saharan African |
| HGDP01081.SDG | Congo | 0 | Sub-Saharan African |
| HGDP01415.SDG | Kenya | 0 | Sub-Saharan African |
| HGDP01418.SDG | Kenya | 0 | Sub-Saharan African |
| HGDP01419.SDG | Kenya | 0 | Sub-Saharan African |
| HGDP00992.SDG | Namibia | 0 | Sub-Saharan African |
| HGDP00926.SDG | Nigeria | 0 | Sub-Saharan African |
| HGDP01255.SDG | Algeria | 0 | Sub-Saharan African |
| HGDP00460.SDG | Central African Republic | 0 | Sub-Saharan African |
| HGDP00930.SDG | Nigeria | 0 | Sub-Saharan African |
| HGDP00984.SDG | Congo | 0 | Sub-Saharan African |
| HGDP01416.SDG | Kenya | 0 | Sub-Saharan African |
| HGDP01033.SDG | South Africa | 0 | Sub-Saharan African |
| HGDP01090.SDG | Central African Republic | 0 | Sub-Saharan African |

| | | | |
|---|---|---|---|
| HGDP00917.SDG | Senegal | 0 | Sub-Saharan African |
| HGDP00912.SDG | Senegal | 0 | Sub-Saharan African |
| HGDP01269.SDG | Algeria | 0 | Sub-Saharan African |
| HGDP01277.SDG | Algeria | 0 | Sub-Saharan African |
| HGDP00909.SDG | Senegal | 0 | Sub-Saharan African |
| HGDP01268.SDG | Algeria | 0 | Sub-Saharan African |
| HGDP01260.SDG | Algeria | 0 | Sub-Saharan African |
| HGDP01254.SDG | Algeria | 0 | Sub-Saharan African |
| HGDP01270.SDG | Algeria | 0 | Sub-Saharan African |
| HGDP01276.SDG | Algeria | 0 | Sub-Saharan African |
| HGDP01262.SDG | Algeria | 0 | Sub-Saharan African |
| HGDP00906.SDG | Senegal | 0 | Sub-Saharan African |
| HGDP01282.SDG | Algeria | 0 | Sub-Saharan African |
| HGDP01275.SDG | Algeria | 0 | Sub-Saharan African |
| HGDP00454.SDG | Central African Republic | 0 | Sub-Saharan African |
| HGDP00455.SDG | Central African Republic | 0 | Sub-Saharan African |

| | | | |
|---|---|---|---|
| HGDP01271.SDG | Algeria | 0 | Sub-Saharan African |
| HGDP01267.SDG | Algeria | 0 | Sub-Saharan African |
| HGDP00913.SDG | Senegal | 0 | Sub-Saharan African |
| HGDP01258.SDG | Algeria | 0 | Sub-Saharan African |
| HGDP01280.SDG | Algeria | 0 | Sub-Saharan African |
| HGDP01261.SDG | Algeria | 0 | Sub-Saharan African |
| HGDP01264.SDG | Algeria | 0 | Sub-Saharan African |
| HGDP01259.SDG | Algeria | 0 | Sub-Saharan African |
| HGDP01263.SDG | Algeria | 0 | Sub-Saharan African |
| HGDP01266.SDG | Algeria | 0 | Sub-Saharan African |
| HGDP01265.SDG | Algeria | 0 | Sub-Saharan African |
| HGDP01272.SDG | Algeria | 0 | Sub-Saharan African |
| HGDP01279.SDG | Algeria | 0 | Sub-Saharan African |
| HGDP01284.SDG | Senegal | 0 | Sub-Saharan African |
| HGDP00456.SDG | Congo | 0 | Sub-Saharan African |
| HGDP00927.SDG | Nigeria | 0 | Sub-Saharan African |

| | | | |
|---|---|---|---|
| HGDP00449.SDG | Congo | 0 | Sub-Saharan African |
| HGDP00476.SDG | Congo | 0 | Sub-Saharan African |
| HGDP01253.SDG | Algeria | 0 | Sub-Saharan African |
| HGDP01032.SDG | Namibia | 0 | Sub-Saharan African |
| HGDP00932.SDG | Nigeria | 0 | Sub-Saharan African |
| HGDP00991.SDG | Namibia | 0 | Sub-Saharan African |
| HGDP00936.SDG | Nigeria | 0 | Sub-Saharan African |
| HGDP01274.SDG | Algeria | 0 | Sub-Saharan African |
| HGDP01286.SDG | Senegal | 0 | Sub-Saharan African |
| HGDP01417.SDG | Kenya | 0 | Sub-Saharan African |
| HGDP01414.SDG | Kenya | 0 | Sub-Saharan African |
| HGDP00474.SDG | Congo | 0 | Sub-Saharan African |
| HGDP01199.SDG | Senegal | 0 | Sub-Saharan African |
| HGDP01036.SDG | Namibia | 0 | Sub-Saharan African |
| HGDP00982.SDG | Congo | 0 | Sub-Saharan African |
| HGDP00987.SDG | Namibia | 0 | Sub-Saharan African |

| | | | |
|---|---|---|---|
| HGDP00461.SDG | Central African Republic | 0 | Sub-Saharan African |
| HGDP00915.SDG | Senegal | 0 | Sub-Saharan African |
| HGDP00928.SDG | Nigeria | 0 | Sub-Saharan African |
| HGDP00457.SDG | Central African Republic | 0 | Sub-Saharan African |
| I10759 | Curacao | 650 | Sub-Saharan African |
| BOT.1.SG | Morocco | 7950 | Sub-Saharan African |
| IAM.3.SG | Morocco | 7210 | Sub-Saharan African |
| IAM.6.SG | Morocco | 7098 | Sub-Saharan African |
| IAM.7.SG | Morocco | 6871 | Sub-Saharan African |
| KEB.4.SG | Morocco | 5650 | Sub-Saharan African |
| KEB.6.SG | Morocco | 5650 | Sub-Saharan African |
| I12967 | Curacao | 660 | Sub-Saharan African |
| I12398 | Kenya | 2589 | Sub-Saharan African |
| I12394 | Kenya | 1466 | Sub-Saharan African |
| I12391 | Kenya | 1009 | Sub-Saharan African |
| I12381 | Kenya | 604 | Sub-Saharan African |

| | | | |
|---|---|---|---|
| I12379 | Kenya | 324 | Sub-Saharan African |
| I12384 | Kenya | 1680 | Sub-Saharan African |
| I13472 | Curacao | 602 | Sub-Saharan African |
| I11892 | Curacao | 603 | Sub-Saharan African |
| I13763_published | Tanzania | 1930 | Sub-Saharan African |
| I12533 | Kenya | 4011 | Sub-Saharan African |
| I8759_published | Kenya | 2693 | Sub-Saharan African |
| I8904 | Kenya | 4200 | Sub-Saharan African |
| I8930_published | Kenya | 4450 | Sub-Saharan African |
| I10758 | Curacao | 666 | Sub-Saharan African |
| I12534 | Kenya | 3958 | Sub-Saharan African |
| I8901 | Kenya | 1009 | Sub-Saharan African |
| I8923 | Kenya | 2508 | Sub-Saharan African |
| I8931_published | Kenya | 4450 | Sub-Saharan African |
| I13762 | Tanzania | 2051 | Sub-Saharan African |
| I13982_published | Tanzania | 2150 | Sub-Saharan African |

| | | | |
|---|---|---|---|
| I13977 | Tanzania | 1924 | Sub-Saharan African |
| I13983_published | Tanzania | 2150 | Sub-Saharan African |
| I13970 | Tanzania | 1950 | Sub-Saharan African |
| I13979 | Tanzania | 2409 | Sub-Saharan African |
| I13978 | Tanzania | 2273 | Sub-Saharan African |
| I13972 | Tanzania | 1647 | Sub-Saharan African |
| I13980 | Tanzania | 2587 | Sub-Saharan African |
| I13981 | Tanzania | 2566 | Sub-Saharan African |
| I10871.DG | Cameroon | 7913 | Sub-Saharan African |
| I10871.SG | Cameroon | 7913 | Sub-Saharan African |
| I10873.SG | Cameroon | 3097 | Sub-Saharan African |
| IAM.4_IAM.5.SG | Morocco | 7063 | Sub-Saharan African |
| KEB.1_KEB.8.SG | Morocco | 5657 | Sub-Saharan African |
| I1048_published_all | Tanzania | 1357 | Sub-Saharan African |
| I4421_published | Malawi | 5150 | Sub-Saharan African |
| I4422_published | Malawi | 5156 | Sub-Saharan African |

| | | | |
|---|---|---|---|
| I4426_published | Malawi | 2427 | Sub-Saharan African |
| I4427_published | Malawi | 6009 | Sub-Saharan African |
| I4468_published | Malawi | 6039 | Sub-Saharan African |
| I2967_published_all | Malawi | 8018 | Sub-Saharan African |
| I10871_published | Cameroon | 7913 | Sub-Saharan African |
| I10872_published | Cameroon | 7820 | Sub-Saharan African |
| I10873_published | Cameroon | 3097 | Sub-Saharan African |
| I10874_published | Cameroon | 3135 | Sub-Saharan African |
| HYR002 | Kenya | 2366 | Sub-Saharan African |
| KIN002 | DR Congo | 211 | Sub-Saharan African |
| KIN003 | DR Congo | 163 | Sub-Saharan African |
| KIN004 | DR Congo | 255 | Sub-Saharan African |
| KPL001 | Kenya | 3887 | Sub-Saharan African |
| KPL002 | Kenya | 213 | Sub-Saharan African |
| KPL003 | Kenya | 808 | Sub-Saharan African |
| LUK001 | Kenya | 3526 | Sub-Saharan African |

| | | | |
|---|---|---|---|
| LUK003 | Kenya | 3591 | Sub-Saharan African |
| MOL001 | Kenya | 1404 | Sub-Saharan African |
| MOL003 | Kenya | 2062 | Sub-Saharan African |
| MTN001 | DR Congo | 766 | Sub-Saharan African |
| MUN001 | Uganda | 450 | Sub-Saharan African |
| NGO001 | DR Congo | 202 | Sub-Saharan African |
| NQO002 | Botswana | 1055 | Sub-Saharan African |
| NYA002 | Kenya | 3462 | Sub-Saharan African |
| NYA003 | Kenya | 3475 | Sub-Saharan African |
| TAU001 | Botswana | 1000 | Sub-Saharan African |
| XAR001 | Botswana | 1100 | Sub-Saharan African |
| XAR002 | Botswana | 1100 | Sub-Saharan African |
| I8904_d | Kenya | 4200 | Sub-Saharan African |
| I10871.v50.0.SG | Cameroon | 7913 | Sub-Saharan African |
| I10871.v50.0_d.SG | Cameroon | 7913 | Sub-Saharan African |
| I0719.SG | USA | 505 | American |
| A_Karitiana-4.DG | Brazil | 0 | American |

| | | | |
|---|---|---|---|
| S_Piapoco-1.DG | Colombia | 0 | American |
| S_Surui-2.DG | Brazil | 0 | American |
| S_Quechua-1.DG | Peru | 0 | American |
| S_Cree-2.DG | Canada | 0 | American |
| S_Chane-1.DG | Argentina | 0 | American |
| S_Quechua-3.DG | Peru | 0 | American |
| B_Mixe-1.DG | Mexico | 0 | American |
| B_Karitiana-3.DG | Brazil | 0 | American |
| S_Pima-2.DG | Mexico | 0 | American |
| S_Chipewyan-2.DG | Canada | 0 | American |
| S_Hawaiian-1.DG | USA | 0 | American |
| S_Zapotec-2.DG | Mexico | 0 | American |
| S_Mixe-2.DG | Mexico | 0 | American |
| S_Mixtec-2.DG | Mexico | 0 | American |
| S_Cree-1.DG | Canada | 0 | American |
| S_Nahua-2.DG | Mexico | 0 | American |
| S_Surui-1.DG | Brazil | 0 | American |
| S_Zapotec-1.DG | Mexico | 0 | American |
| S_Mayan-1.DG | Mexico | 0 | American |
| S_Karitiana-1.DG | Brazil | 0 | American |
| S_Pima-1.DG | Mexico | 0 | American |
| S_Mixe-3.DG | Mexico | 0 | American |
| S_Piapoco-2.DG | Colombia | 0 | American |
| S_Quechua-2.DG | Peru | 0 | American |
| S_Karitiana-2.DG | Brazil | 0 | American |
| S_Mayan-2.DG | Mexico | 0 | American |
| S_Mixtec-1.DG | Mexico | 0 | American |
| S_Chipewyan-1.DG | Canada | 0 | American |
| BI16.SG | Brazil | 0 | American |

| | | | |
|---|---|---|---|
| HGDP00877.SG | Mexico | 0 | American |
| HUI03.SG | Mexico | 0 | American |
| TA6.SG | Peru | 0 | American |
| Tsimshian.SG | Canada | 0 | American |
| Y2040.SG | Venezuela | 0 | American |
| XVII-B-939.SG | Canada | 6502 | American |
| Chinchorroi15.SG | Chile | 6242 | American |
| Enoque65.SG | Brazil | 3542 | American |
| Yaghan890.SG | Argentina | 100 | American |
| Yaghan894.SG | Argentina | 100 | American |
| Yaghan895.SG | Argentina | 100 | American |
| AM71.SG | Chile | 100 | American |
| BC23.SG | Mexico | 500 | American |
| BC25.SG | Mexico | 500 | American |
| BC27.SG | Mexico | 500 | American |
| BC28.SG | Mexico | 500 | American |
| BC29.SG | Mexico | 500 | American |
| BC30.SG | Mexico | 500 | American |
| F9.SG | Mexico | 3200 | American |
| MA572.SG | Argentina | 100 | American |
| MA575.SG | Argentina | 100 | American |
| MA577.SG | Argentina | 100 | American |
| MOM6.SG | Mexico | 3200 | American |
| AM66.SG | Chile | 100 | American |
| AM72.SG | Chile | 100 | American |
| AM73.SG | Chile | 100 | American |
| AM74.SG | Chile | 100 | American |
| MARC1492.SG | Canada | 676 | American |
| I0974 | Peru | 1500 | American |

| | | | |
|---|---|---|---|
| I0975 | Peru | 1220 | American |
| I0041 | Peru | 8505 | American |
| I0039 | Peru | 3530 | American |
| I0040 | Peru | 5845 | American |
| I0045 | Peru | 1575 | American |
| I0979 | Bolivia | 1125 | American |
| I0977 | Bolivia | 990 | American |
| I0976 | Bolivia | 900 | American |
| I0978 | Bolivia | 865 | American |
| I0971 | Peru | 1250 | American |
| I0969 | Peru | 685 | American |
| I0968 | Peru | 865 | American |
| I0972 | Peru | 765 | American |
| I0964 | Peru | 540 | American |
| I0965 | Peru | 555 | American |
| I0967 | Peru | 765 | American |
| I0966 | Peru | 765 | American |
| I1748 | Argentina | 7180 | American |
| I0309 | Argentina | 7650 | American |
| I1357 | Peru | 925 | American |
| I1484 | Peru | 815 | American |
| I1743 | Peru | 495 | American |
| I1744 | Peru | 605 | American |
| I1400 | Peru | 605 | American |
| I0238 | Peru | 8450 | American |
| I1479 | Peru | 550 | American |
| I1396 | Peru | 590 | American |
| I1358 | Peru | 735 | American |
| I1485 | Peru | 1060 | American |

| | | | |
|---|---|---|---|
| I1742 | Peru | 1125 | American |
| I1356 | Peru | 600 | American |
| I1123_published | USA | 315 | American |
| I1124_published | USA | 230 | American |
| I1128 | USA | 405 | American |
| I1129 | USA | 395 | American |
| I1125 | USA | 575 | American |
| I1127 | USA | 435 | American |
| I1118 | USA | 315 | American |
| I0721_published | USA | 2070 | American |
| I0712 | USA | 1065 | American |
| I0719 | USA | 505 | American |
| LateDorset-XIV-H_126.SG | Canada | 800 | American |
| MARC1491.SG | Canada | 1550 | American |
| MARC481.SG | Canada | 1550 | American |
| XIV_C_748.SG | Canada | 600 | American |
| Botocudo15.SG | Brazil | 309 | American |
| Botocudo17.SG | Brazil | 281 | American |
| I0237 | Peru | 860 | American |
| I0038 | Peru | 8625 | American |
| I0044 | Peru | 615 | American |
| kennewick.SG | USA | 8752 | American |
| I0308 | Argentina | 7435 | American |
| I2230 | Argentina | 8670 | American |
| I2232 | Argentina | 8360 | American |
| I2233 | Peru | 1915 | American |
| I2236 | Peru | 650 | American |
| I2237 | Peru | 1335 | American |
| I2238 | Peru | 1550 | American |

| | | | |
|---|---|---|---|
| I2241 | Peru | 925 | American |
| I2242 | Peru | 685 | American |
| I2243 | Peru | 925 | American |
| I2244 | Peru | 925 | American |
| I2250 | Peru | 700 | American |
| I2251 | Peru | 700 | American |
| I2252 | Peru | 1025 | American |
| I2253 | Peru | 1025 | American |
| I2261 | Peru | 4105 | American |
| I2262 | Peru | 1550 | American |
| I2263 | Peru | 1095 | American |
| I2264 | Peru | 700 | American |
| I2268 | Peru | 4150 | American |
| I2537 | Chile | 645 | American |
| I2539 | Chile | 990 | American |
| I2540 | Chile | 1000 | American |
| I2543 | Peru | 945 | American |
| I2544 | Peru | 675 | American |
| I2545 | Peru | 470 | American |
| I2549 | Peru | 540 | American |
| I2550 | Peru | 935 | American |
| I2551 | Peru | 825 | American |
| I1752 | Chile | 595 | American |
| I1753 | Chile | 5095 | American |
| I1754 | Chile | 825 | American |
| I2557 | Peru | 1395 | American |
| I2558 | Peru | 1320 | American |
| I2560 | Peru | 950 | American |
| I2563 | Peru | 650 | American |

| | | | |
|---|---|---|---|
| I2258_d | Peru | 600 | American |
| CP18 | Brazil | 9510 | American |
| CP19 | Brazil | 9880 | American |
| CP23 | Brazil | 9650 | American |
| CP25 | Brazil | 9605 | American |
| CP26 | Brazil | 9650 | American |
| HG00551.SG | Puerto Rico | 0 | American |
| HG00553.SG | Puerto Rico | 0 | American |
| HG00554.SG | Puerto Rico | 0 | American |
| HG00637.SG | Puerto Rico | 0 | American |
| HG00638.SG | Puerto Rico | 0 | American |
| HG00640.SG | Puerto Rico | 0 | American |
| HG00641.SG | Puerto Rico | 0 | American |
| HG00731.SG | Puerto Rico | 0 | American |
| HG00732.SG | Puerto Rico | 0 | American |
| HG00733.SG | Puerto Rico | 0 | American |
| HG00734.SG | Puerto Rico | 0 | American |
| HG00736.SG | Puerto Rico | 0 | American |
| HG00737.SG | Puerto Rico | 0 | American |
| HG00739.SG | Puerto Rico | 0 | American |
| HG00740.SG | Puerto Rico | 0 | American |
| HG00742.SG | Puerto Rico | 0 | American |
| HG00743.SG | Puerto Rico | 0 | American |
| HG01047.SG | Puerto Rico | 0 | American |
| HG01048.SG | Puerto Rico | 0 | American |
| HG01049.SG | Puerto Rico | 0 | American |
| HG01051.SG | Puerto Rico | 0 | American |
| HG01052.SG | Puerto Rico | 0 | American |
| HG01054.SG | Puerto Rico | 0 | American |

| | | | |
|---|---|---|---|
| HG01055.SG | Puerto Rico | 0 | American |
| HG01058.SG | Puerto Rico | 0 | American |
| HG01060.SG | Puerto Rico | 0 | American |
| HG01061.SG | Puerto Rico | 0 | American |
| HG01063.SG | Puerto Rico | 0 | American |
| HG01064.SG | Puerto Rico | 0 | American |
| HG01066.SG | Puerto Rico | 0 | American |
| HG01067.SG | Puerto Rico | 0 | American |
| HG01069.SG | Puerto Rico | 0 | American |
| HG01070.SG | Puerto Rico | 0 | American |
| HG01072.SG | Puerto Rico | 0 | American |
| HG01073.SG | Puerto Rico | 0 | American |
| HG01075.SG | Puerto Rico | 0 | American |
| HG01077.SG | Puerto Rico | 0 | American |
| HG01079.SG | Puerto Rico | 0 | American |
| HG01080.SG | Puerto Rico | 0 | American |
| HG01082.SG | Puerto Rico | 0 | American |
| HG01083.SG | Puerto Rico | 0 | American |
| HG01085.SG | Puerto Rico | 0 | American |
| HG01086.SG | Puerto Rico | 0 | American |
| HG01088.SG | Puerto Rico | 0 | American |
| HG01089.SG | Puerto Rico | 0 | American |
| HG01092.SG | Puerto Rico | 0 | American |
| HG01094.SG | Puerto Rico | 0 | American |
| HG01095.SG | Puerto Rico | 0 | American |
| HG01097.SG | Puerto Rico | 0 | American |
| HG01098.SG | Puerto Rico | 0 | American |
| HG01101.SG | Puerto Rico | 0 | American |
| HG01102.SG | Puerto Rico | 0 | American |

| | | | |
|---|---|---|---|
| HG01104.SG | Puerto Rico | 0 | American |
| HG01105.SG | Puerto Rico | 0 | American |
| HG01107.SG | Puerto Rico | 0 | American |
| HG01108.SG | Puerto Rico | 0 | American |
| HG01110.SG | Puerto Rico | 0 | American |
| HG01111.SG | Puerto Rico | 0 | American |
| HG01112.SG | Colombia | 0 | American |
| HG01113.SG | Colombia | 0 | American |
| HG01119.SG | Colombia | 0 | American |
| HG01121.SG | Colombia | 0 | American |
| HG01122.SG | Colombia | 0 | American |
| HG01124.SG | Colombia | 0 | American |
| HG01125.SG | Colombia | 0 | American |
| HG01130.SG | Colombia | 0 | American |
| HG01131.SG | Colombia | 0 | American |
| HG01133.SG | Colombia | 0 | American |
| HG01134.SG | Colombia | 0 | American |
| HG01136.SG | Colombia | 0 | American |
| HG01137.SG | Colombia | 0 | American |
| HG01139.SG | Colombia | 0 | American |
| HG01140.SG | Colombia | 0 | American |
| HG01142.SG | Colombia | 0 | American |
| HG01148.SG | Colombia | 0 | American |
| HG01149.SG | Colombia | 0 | American |
| HG01161.SG | Puerto Rico | 0 | American |
| HG01162.SG | Puerto Rico | 0 | American |
| HG01164.SG | Puerto Rico | 0 | American |
| HG01167.SG | Puerto Rico | 0 | American |
| HG01168.SG | Puerto Rico | 0 | American |

| | | | |
|---|---|---|---|
| HG01170.SG | Puerto Rico | 0 | American |
| HG01171.SG | Puerto Rico | 0 | American |
| HG01173.SG | Puerto Rico | 0 | American |
| HG01174.SG | Puerto Rico | 0 | American |
| HG01176.SG | Puerto Rico | 0 | American |
| HG01177.SG | Puerto Rico | 0 | American |
| HG01182.SG | Puerto Rico | 0 | American |
| HG01183.SG | Puerto Rico | 0 | American |
| HG01187.SG | Puerto Rico | 0 | American |
| HG01188.SG | Puerto Rico | 0 | American |
| HG01190.SG | Puerto Rico | 0 | American |
| HG01191.SG | Puerto Rico | 0 | American |
| HG01197.SG | Puerto Rico | 0 | American |
| HG01198.SG | Puerto Rico | 0 | American |
| HG01200.SG | Puerto Rico | 0 | American |
| HG01204.SG | Puerto Rico | 0 | American |
| HG01205.SG | Puerto Rico | 0 | American |
| HG01241.SG | Puerto Rico | 0 | American |
| HG01242.SG | Puerto Rico | 0 | American |
| HG01247.SG | Puerto Rico | 0 | American |
| HG01248.SG | Puerto Rico | 0 | American |
| HG01250.SG | Colombia | 0 | American |
| HG01251.SG | Colombia | 0 | American |
| HG01253.SG | Colombia | 0 | American |
| HG01254.SG | Colombia | 0 | American |
| HG01256.SG | Colombia | 0 | American |
| HG01257.SG | Colombia | 0 | American |
| HG01259.SG | Colombia | 0 | American |
| HG01260.SG | Colombia | 0 | American |

| | | | |
|---|---|---|---|
| HG01269.SG | Colombia | 0 | American |
| HG01271.SG | Colombia | 0 | American |
| HG01272.SG | Colombia | 0 | American |
| HG01275.SG | Colombia | 0 | American |
| HG01277.SG | Colombia | 0 | American |
| HG01280.SG | Colombia | 0 | American |
| HG01281.SG | Colombia | 0 | American |
| HG01284.SG | Colombia | 0 | American |
| HG01286.SG | Puerto Rico | 0 | American |
| HG01302.SG | Puerto Rico | 0 | American |
| HG01303.SG | Puerto Rico | 0 | American |
| HG01305.SG | Puerto Rico | 0 | American |
| HG01308.SG | Puerto Rico | 0 | American |
| HG01311.SG | Puerto Rico | 0 | American |
| HG01312.SG | Puerto Rico | 0 | American |
| HG01323.SG | Puerto Rico | 0 | American |
| HG01325.SG | Puerto Rico | 0 | American |
| HG01326.SG | Puerto Rico | 0 | American |
| HG01341.SG | Colombia | 0 | American |
| HG01342.SG | Colombia | 0 | American |
| HG01344.SG | Colombia | 0 | American |
| HG01345.SG | Colombia | 0 | American |
| HG01348.SG | Colombia | 0 | American |
| HG01350.SG | Colombia | 0 | American |
| HG01351.SG | Colombia | 0 | American |
| HG01353.SG | Colombia | 0 | American |
| HG01354.SG | Colombia | 0 | American |
| HG01356.SG | Colombia | 0 | American |
| HG01357.SG | Colombia | 0 | American |

| | | | |
|---|---|---|---|
| HG01359.SG | Colombia | 0 | American |
| HG01360.SG | Colombia | 0 | American |
| HG01362.SG | Colombia | 0 | American |
| HG01363.SG | Colombia | 0 | American |
| HG01365.SG | Colombia | 0 | American |
| HG01366.SG | Colombia | 0 | American |
| HG01369.SG | Colombia | 0 | American |
| HG01372.SG | Colombia | 0 | American |
| HG01374.SG | Colombia | 0 | American |
| HG01375.SG | Colombia | 0 | American |
| HG01377.SG | Colombia | 0 | American |
| HG01378.SG | Colombia | 0 | American |
| HG01383.SG | Colombia | 0 | American |
| HG01384.SG | Colombia | 0 | American |
| HG01389.SG | Colombia | 0 | American |
| HG01390.SG | Colombia | 0 | American |
| HG01392.SG | Puerto Rico | 0 | American |
| HG01393.SG | Puerto Rico | 0 | American |
| HG01395.SG | Puerto Rico | 0 | American |
| HG01396.SG | Puerto Rico | 0 | American |
| HG01398.SG | Puerto Rico | 0 | American |
| HG01402.SG | Puerto Rico | 0 | American |
| HG01403.SG | Puerto Rico | 0 | American |
| HG01405.SG | Puerto Rico | 0 | American |
| HG01412.SG | Puerto Rico | 0 | American |
| HG01413.SG | Puerto Rico | 0 | American |
| HG01414.SG | Puerto Rico | 0 | American |
| HG01431.SG | Colombia | 0 | American |
| HG01432.SG | Colombia | 0 | American |

| | | | |
|---|---|---|---|
| HG01435.SG | Colombia | 0 | American |
| HG01437.SG | Colombia | 0 | American |
| HG01438.SG | Colombia | 0 | American |
| HG01440.SG | Colombia | 0 | American |
| HG01441.SG | Colombia | 0 | American |
| HG01443.SG | Colombia | 0 | American |
| HG01444.SG | Colombia | 0 | American |
| HG01447.SG | Colombia | 0 | American |
| HG01455.SG | Colombia | 0 | American |
| HG01456.SG | Colombia | 0 | American |
| HG01459.SG | Colombia | 0 | American |
| HG01461.SG | Colombia | 0 | American |
| HG01462.SG | Colombia | 0 | American |
| HG01464.SG | Colombia | 0 | American |
| HG01465.SG | Colombia | 0 | American |
| HG01468.SG | Colombia | 0 | American |
| HG01474.SG | Colombia | 0 | American |
| HG01479.SG | Colombia | 0 | American |
| HG01485.SG | Colombia | 0 | American |
| HG01486.SG | Colombia | 0 | American |
| HG01488.SG | Colombia | 0 | American |
| HG01489.SG | Colombia | 0 | American |
| HG01491.SG | Colombia | 0 | American |
| HG01492.SG | Colombia | 0 | American |
| HG01494.SG | Colombia | 0 | American |
| HG01495.SG | Colombia | 0 | American |
| HG01497.SG | Colombia | 0 | American |
| HG01498.SG | Colombia | 0 | American |
| HG01550.SG | Colombia | 0 | American |

| | | | |
|---|---|---|---|
| HG01551.SG | Colombia | 0 | American |
| HG01556.SG | Colombia | 0 | American |
| HG01565.SG | Peru | 0 | American |
| HG01566.SG | Peru | 0 | American |
| HG01571.SG | Peru | 0 | American |
| HG01572.SG | Peru | 0 | American |
| HG01577.SG | Peru | 0 | American |
| HG01578.SG | Peru | 0 | American |
| HG01879.SG | Barbados | 0 | American |
| HG01880.SG | Barbados | 0 | American |
| HG01882.SG | Barbados | 0 | American |
| HG01883.SG | Barbados | 0 | American |
| HG01885.SG | Barbados | 0 | American |
| HG01886.SG | Barbados | 0 | American |
| HG01889.SG | Barbados | 0 | American |
| HG01890.SG | Barbados | 0 | American |
| HG01892.SG | Peru | 0 | American |
| HG01893.SG | Peru | 0 | American |
| HG01894.SG | Barbados | 0 | American |
| HG01896.SG | Barbados | 0 | American |
| HG01912.SG | Barbados | 0 | American |
| HG01914.SG | Barbados | 0 | American |
| HG01915.SG | Barbados | 0 | American |
| HG01917.SG | Peru | 0 | American |
| HG01918.SG | Peru | 0 | American |
| HG01920.SG | Peru | 0 | American |
| HG01921.SG | Peru | 0 | American |
| HG01923.SG | Peru | 0 | American |
| HG01924.SG | Peru | 0 | American |

| | | | |
|---|---|---|---|
| HG01926.SG | Peru | 0 | American |
| HG01927.SG | Peru | 0 | American |
| HG01932.SG | Peru | 0 | American |
| HG01933.SG | Peru | 0 | American |
| HG01935.SG | Peru | 0 | American |
| HG01936.SG | Peru | 0 | American |
| HG01938.SG | Peru | 0 | American |
| HG01939.SG | Peru | 0 | American |
| HG01941.SG | Peru | 0 | American |
| HG01942.SG | Peru | 0 | American |
| HG01944.SG | Peru | 0 | American |
| HG01945.SG | Peru | 0 | American |
| HG01947.SG | Peru | 0 | American |
| HG01948.SG | Peru | 0 | American |
| HG01950.SG | Peru | 0 | American |
| HG01951.SG | Peru | 0 | American |
| HG01953.SG | Peru | 0 | American |
| HG01954.SG | Peru | 0 | American |
| HG01956.SG | Barbados | 0 | American |
| HG01958.SG | Barbados | 0 | American |
| HG01961.SG | Peru | 0 | American |
| HG01965.SG | Peru | 0 | American |
| HG01967.SG | Peru | 0 | American |
| HG01968.SG | Peru | 0 | American |
| HG01970.SG | Peru | 0 | American |
| HG01971.SG | Peru | 0 | American |
| HG01973.SG | Peru | 0 | American |
| HG01974.SG | Peru | 0 | American |
| HG01976.SG | Peru | 0 | American |

| | | | |
|---|---|---|---|
| HG01977.SG | Peru | 0 | American |
| HG01979.SG | Peru | 0 | American |
| HG01980.SG | Peru | 0 | American |
| HG01982.SG | Peru | 0 | American |
| HG01983.SG | Peru | 0 | American |
| HG01985.SG | Barbados | 0 | American |
| HG01986.SG | Barbados | 0 | American |
| HG01988.SG | Barbados | 0 | American |
| HG01989.SG | Barbados | 0 | American |
| HG01990.SG | Barbados | 0 | American |
| HG01991.SG | Peru | 0 | American |
| HG01992.SG | Peru | 0 | American |
| HG01997.SG | Peru | 0 | American |
| HG02002.SG | Peru | 0 | American |
| HG02003.SG | Peru | 0 | American |
| HG02006.SG | Peru | 0 | American |
| HG02008.SG | Peru | 0 | American |
| HG02009.SG | Barbados | 0 | American |
| HG02010.SG | Barbados | 0 | American |
| HG02012.SG | Barbados | 0 | American |
| HG02013.SG | Barbados | 0 | American |
| HG02014.SG | Barbados | 0 | American |
| HG02051.SG | Barbados | 0 | American |
| HG02052.SG | Barbados | 0 | American |
| HG02053.SG | Barbados | 0 | American |
| HG02054.SG | Barbados | 0 | American |
| HG02089.SG | Peru | 0 | American |
| HG02090.SG | Peru | 0 | American |
| HG02095.SG | Barbados | 0 | American |

| | | | |
|---|---|---|---|
| HG02102.SG | Peru | 0 | American |
| HG02104.SG | Peru | 0 | American |
| HG02105.SG | Peru | 0 | American |
| HG02107.SG | Barbados | 0 | American |
| HG02108.SG | Barbados | 0 | American |
| HG02111.SG | Barbados | 0 | American |
| HG02143.SG | Barbados | 0 | American |
| HG02144.SG | Barbados | 0 | American |
| HG02146.SG | Peru | 0 | American |
| HG02147.SG | Peru | 0 | American |
| HG02150.SG | Peru | 0 | American |
| HG02252.SG | Peru | 0 | American |
| HG02253.SG | Peru | 0 | American |
| HG02255.SG | Barbados | 0 | American |
| HG02256.SG | Barbados | 0 | American |
| HG02259.SG | Peru | 0 | American |
| HG02260.SG | Peru | 0 | American |
| HG02262.SG | Peru | 0 | American |
| HG02265.SG | Peru | 0 | American |
| HG02266.SG | Peru | 0 | American |
| HG02271.SG | Peru | 0 | American |
| HG02272.SG | Peru | 0 | American |
| HG02274.SG | Peru | 0 | American |
| HG02275.SG | Peru | 0 | American |
| HG02277.SG | Peru | 0 | American |
| HG02278.SG | Peru | 0 | American |
| HG02281.SG | Barbados | 0 | American |
| HG02282.SG | Barbados | 0 | American |
| HG02283.SG | Barbados | 0 | American |

| | | | |
|---|---|---|---|
| HG02284.SG | Barbados | 0 | American |
| HG02285.SG | Peru | 0 | American |
| HG02286.SG | Peru | 0 | American |
| HG02291.SG | Peru | 0 | American |
| HG02292.SG | Peru | 0 | American |
| HG02298.SG | Peru | 0 | American |
| HG02299.SG | Peru | 0 | American |
| HG02301.SG | Peru | 0 | American |
| HG02304.SG | Peru | 0 | American |
| HG02307.SG | Barbados | 0 | American |
| HG02308.SG | Barbados | 0 | American |
| HG02309.SG | Barbados | 0 | American |
| HG02312.SG | Peru | 0 | American |
| HG02314.SG | Barbados | 0 | American |
| HG02315.SG | Barbados | 0 | American |
| HG02317.SG | Barbados | 0 | American |
| HG02318.SG | Barbados | 0 | American |
| HG02322.SG | Barbados | 0 | American |
| HG02323.SG | Barbados | 0 | American |
| HG02325.SG | Barbados | 0 | American |
| HG02330.SG | Barbados | 0 | American |
| HG02332.SG | Barbados | 0 | American |
| HG02334.SG | Barbados | 0 | American |
| HG02337.SG | Barbados | 0 | American |
| HG02339.SG | Barbados | 0 | American |
| HG02343.SG | Barbados | 0 | American |
| HG02345.SG | Peru | 0 | American |
| HG02348.SG | Peru | 0 | American |
| HG02419.SG | Barbados | 0 | American |

| | | | |
|---|---|---|---|
| HG02420.SG | Barbados | 0 | American |
| HG02425.SG | Peru | 0 | American |
| HG02427.SG | Barbados | 0 | American |
| HG02429.SG | Barbados | 0 | American |
| HG02433.SG | Barbados | 0 | American |
| HG02439.SG | Barbados | 0 | American |
| HG02442.SG | Barbados | 0 | American |
| HG02445.SG | Barbados | 0 | American |
| HG02449.SG | Barbados | 0 | American |
| HG02450.SG | Barbados | 0 | American |
| HG02455.SG | Barbados | 0 | American |
| HG02470.SG | Barbados | 0 | American |
| HG02471.SG | Barbados | 0 | American |
| HG02476.SG | Barbados | 0 | American |
| HG02477.SG | Barbados | 0 | American |
| HG02479.SG | Barbados | 0 | American |
| HG02481.SG | Barbados | 0 | American |
| HG02484.SG | Barbados | 0 | American |
| HG02485.SG | Barbados | 0 | American |
| HG02489.SG | Barbados | 0 | American |
| HG02496.SG | Barbados | 0 | American |
| HG02497.SG | Barbados | 0 | American |
| HG02501.SG | Barbados | 0 | American |
| HG02502.SG | Barbados | 0 | American |
| HG02505.SG | Barbados | 0 | American |
| HG02508.SG | Barbados | 0 | American |
| HG02511.SG | Barbados | 0 | American |
| HG02536.SG | Barbados | 0 | American |
| HG02537.SG | Barbados | 0 | American |

| | | | |
|---|---|---|---|
| HG02541.SG | Barbados | 0 | American |
| HG02545.SG | Barbados | 0 | American |
| HG02546.SG | Barbados | 0 | American |
| HG02549.SG | Barbados | 0 | American |
| HG02554.SG | Barbados | 0 | American |
| HG02555.SG | Barbados | 0 | American |
| HG02557.SG | Barbados | 0 | American |
| HG02558.SG | Barbados | 0 | American |
| HG02577.SG | Barbados | 0 | American |
| HG02580.SG | Barbados | 0 | American |
| NA06984.SG | USA | 0 | American |
| NA06985.SG | USA | 0 | American |
| NA06986.SG | USA | 0 | American |
| NA06989.SG | USA | 0 | American |
| NA06994.SG | USA | 0 | American |
| NA07000.SG | USA | 0 | American |
| NA07037.SG | USA | 0 | American |
| NA07048.SG | USA | 0 | American |
| NA07051.SG | USA | 0 | American |
| NA07056.SG | USA | 0 | American |
| NA07347.SG | USA | 0 | American |
| NA07357.SG | USA | 0 | American |
| NA10847.SG | USA | 0 | American |
| NA10851.SG | USA | 0 | American |
| NA11829.SG | USA | 0 | American |
| NA11830.SG | USA | 0 | American |
| NA11831.SG | USA | 0 | American |
| NA11832.SG | USA | 0 | American |
| NA11840.SG | USA | 0 | American |

| | | | |
|---|---|---|---|
| NA11843.SG | USA | 0 | American |
| NA11881.SG | USA | 0 | American |
| NA11892.SG | USA | 0 | American |
| NA11893.SG | USA | 0 | American |
| NA11894.SG | USA | 0 | American |
| NA11918.SG | USA | 0 | American |
| NA11919.SG | USA | 0 | American |
| NA11920.SG | USA | 0 | American |
| NA11930.SG | USA | 0 | American |
| NA11931.SG | USA | 0 | American |
| NA11932.SG | USA | 0 | American |
| NA11933.SG | USA | 0 | American |
| NA11992.SG | USA | 0 | American |
| NA11994.SG | USA | 0 | American |
| NA11995.SG | USA | 0 | American |
| NA12003.SG | USA | 0 | American |
| NA12004.SG | USA | 0 | American |
| NA12005.SG | USA | 0 | American |
| NA12006.SG | USA | 0 | American |
| NA12043.SG | USA | 0 | American |
| NA12044.SG | USA | 0 | American |
| NA12045.SG | USA | 0 | American |
| NA12046.SG | USA | 0 | American |
| NA12058.SG | USA | 0 | American |
| NA12144.SG | USA | 0 | American |
| NA12154.SG | USA | 0 | American |
| NA12155.SG | USA | 0 | American |
| NA12156.SG | USA | 0 | American |
| NA12234.SG | USA | 0 | American |

| | | | |
|---|---|---|---|
| NA12249.SG | USA | 0 | American |
| NA12272.SG | USA | 0 | American |
| NA12273.SG | USA | 0 | American |
| NA12275.SG | USA | 0 | American |
| NA12282.SG | USA | 0 | American |
| NA12283.SG | USA | 0 | American |
| NA12286.SG | USA | 0 | American |
| NA12287.SG | USA | 0 | American |
| NA12340.SG | USA | 0 | American |
| NA12341.SG | USA | 0 | American |
| NA12342.SG | USA | 0 | American |
| NA12347.SG | USA | 0 | American |
| NA12348.SG | USA | 0 | American |
| NA12383.SG | USA | 0 | American |
| NA12399.SG | USA | 0 | American |
| NA12400.SG | USA | 0 | American |
| NA12413.SG | USA | 0 | American |
| NA12414.SG | USA | 0 | American |
| NA12489.SG | USA | 0 | American |
| NA12546.SG | USA | 0 | American |
| NA12716.SG | USA | 0 | American |
| NA12717.SG | USA | 0 | American |
| NA12718.SG | USA | 0 | American |
| NA12748.SG | USA | 0 | American |
| NA12749.SG | USA | 0 | American |
| NA12750.SG | USA | 0 | American |
| NA12751.SG | USA | 0 | American |
| NA12760.SG | USA | 0 | American |
| NA12761.SG | USA | 0 | American |

| | | | |
|---|---|---|---|
| NA12762.SG | USA | 0 | American |
| NA12763.SG | USA | 0 | American |
| NA12775.SG | USA | 0 | American |
| NA12776.SG | USA | 0 | American |
| NA12777.SG | USA | 0 | American |
| NA12778.SG | USA | 0 | American |
| NA12812.SG | USA | 0 | American |
| NA12813.SG | USA | 0 | American |
| NA12814.SG | USA | 0 | American |
| NA12815.SG | USA | 0 | American |
| NA12827.SG | USA | 0 | American |
| NA12828.SG | USA | 0 | American |
| NA12829.SG | USA | 0 | American |
| NA12830.SG | USA | 0 | American |
| NA12842.SG | USA | 0 | American |
| NA12843.SG | USA | 0 | American |
| NA12872.SG | USA | 0 | American |
| NA12873.SG | USA | 0 | American |
| NA12874.SG | USA | 0 | American |
| NA12878.SG | USA | 0 | American |
| NA12889.SG | USA | 0 | American |
| NA12890.SG | USA | 0 | American |
| NA19625.SG | USA | 0 | American |
| NA19648.SG | USA | 0 | American |
| NA19649.SG | USA | 0 | American |
| NA19651.SG | USA | 0 | American |
| NA19652.SG | USA | 0 | American |
| NA19654.SG | USA | 0 | American |
| NA19655.SG | USA | 0 | American |

| | | | |
|---|---|---|---|
| NA19657.SG | USA | 0 | American |
| NA19658.SG | USA | 0 | American |
| NA19660.SG | USA | 0 | American |
| NA19661.SG | USA | 0 | American |
| NA19663.SG | USA | 0 | American |
| NA19664.SG | USA | 0 | American |
| NA19669.SG | USA | 0 | American |
| NA19670.SG | USA | 0 | American |
| NA19675.SG | USA | 0 | American |
| NA19676.SG | USA | 0 | American |
| NA19678.SG | USA | 0 | American |
| NA19679.SG | USA | 0 | American |
| NA19681.SG | USA | 0 | American |
| NA19682.SG | USA | 0 | American |
| NA19684.SG | USA | 0 | American |
| NA19685.SG | USA | 0 | American |
| NA19700.SG | USA | 0 | American |
| NA19701.SG | USA | 0 | American |
| NA19703.SG | USA | 0 | American |
| NA19704.SG | USA | 0 | American |
| NA19707.SG | USA | 0 | American |
| NA19711.SG | USA | 0 | American |
| NA19712.SG | USA | 0 | American |
| NA19713.SG | USA | 0 | American |
| NA19716.SG | USA | 0 | American |
| NA19717.SG | USA | 0 | American |
| NA19719.SG | USA | 0 | American |
| NA19720.SG | USA | 0 | American |
| NA19722.SG | USA | 0 | American |

| | | | |
|---|---|---|---|
| NA19723.SG | USA | 0 | American |
| NA19725.SG | USA | 0 | American |
| NA19726.SG | USA | 0 | American |
| NA19728.SG | USA | 0 | American |
| NA19729.SG | USA | 0 | American |
| NA19731.SG | USA | 0 | American |
| NA19732.SG | USA | 0 | American |
| NA19734.SG | USA | 0 | American |
| NA19735.SG | USA | 0 | American |
| NA19740.SG | USA | 0 | American |
| NA19741.SG | USA | 0 | American |
| NA19746.SG | USA | 0 | American |
| NA19747.SG | USA | 0 | American |
| NA19749.SG | USA | 0 | American |
| NA19750.SG | USA | 0 | American |
| NA19752.SG | USA | 0 | American |
| NA19755.SG | USA | 0 | American |
| NA19756.SG | USA | 0 | American |
| NA19758.SG | USA | 0 | American |
| NA19759.SG | USA | 0 | American |
| NA19761.SG | USA | 0 | American |
| NA19762.SG | USA | 0 | American |
| NA19764.SG | USA | 0 | American |
| NA19770.SG | USA | 0 | American |
| NA19771.SG | USA | 0 | American |
| NA19773.SG | USA | 0 | American |
| NA19774.SG | USA | 0 | American |
| NA19776.SG | USA | 0 | American |
| NA19777.SG | USA | 0 | American |

| | | | |
|---|---|---|---|
| NA19779.SG | USA | 0 | American |
| NA19780.SG | USA | 0 | American |
| NA19782.SG | USA | 0 | American |
| NA19783.SG | USA | 0 | American |
| NA19785.SG | USA | 0 | American |
| NA19786.SG | USA | 0 | American |
| NA19788.SG | USA | 0 | American |
| NA19789.SG | USA | 0 | American |
| NA19792.SG | USA | 0 | American |
| NA19794.SG | USA | 0 | American |
| NA19795.SG | USA | 0 | American |
| NA19818.SG | USA | 0 | American |
| NA19819.SG | USA | 0 | American |
| NA19834.SG | USA | 0 | American |
| NA19835.SG | USA | 0 | American |
| NA19900.SG | USA | 0 | American |
| NA19901.SG | USA | 0 | American |
| NA19904.SG | USA | 0 | American |
| NA19908.SG | USA | 0 | American |
| NA19909.SG | USA | 0 | American |
| NA19913.SG | USA | 0 | American |
| NA19914.SG | USA | 0 | American |
| NA19916.SG | USA | 0 | American |
| NA19917.SG | USA | 0 | American |
| NA19920.SG | USA | 0 | American |
| NA19921.SG | USA | 0 | American |
| NA19922.SG | USA | 0 | American |
| NA19923.SG | USA | 0 | American |
| NA19982.SG | USA | 0 | American |

| | | | |
|---|---|---|---|
| NA19984.SG | USA | 0 | American |
| NA19985.SG | USA | 0 | American |
| NA20126.SG | USA | 0 | American |
| NA20127.SG | USA | 0 | American |
| NA20274.SG | USA | 0 | American |
| NA20276.SG | USA | 0 | American |
| NA20278.SG | USA | 0 | American |
| NA20281.SG | USA | 0 | American |
| NA20282.SG | USA | 0 | American |
| NA20287.SG | USA | 0 | American |
| NA20289.SG | USA | 0 | American |
| NA20291.SG | USA | 0 | American |
| NA20294.SG | USA | 0 | American |
| NA20296.SG | USA | 0 | American |
| NA20298.SG | USA | 0 | American |
| NA20299.SG | USA | 0 | American |
| NA20314.SG | USA | 0 | American |
| NA20317.SG | USA | 0 | American |
| NA20318.SG | USA | 0 | American |
| NA20320.SG | USA | 0 | American |
| NA20321.SG | USA | 0 | American |
| NA20322.SG | USA | 0 | American |
| NA20332.SG | USA | 0 | American |
| NA20334.SG | USA | 0 | American |
| NA20336.SG | USA | 0 | American |
| NA20339.SG | USA | 0 | American |
| NA20340.SG | USA | 0 | American |
| NA20341.SG | USA | 0 | American |
| NA20342.SG | USA | 0 | American |

| | | | |
|---|---|---|---|
| NA20344.SG | USA | 0 | American |
| NA20346.SG | USA | 0 | American |
| NA20348.SG | USA | 0 | American |
| NA20351.SG | USA | 0 | American |
| NA20355.SG | USA | 0 | American |
| NA20356.SG | USA | 0 | American |
| NA20357.SG | USA | 0 | American |
| NA20359.SG | USA | 0 | American |
| NA20362.SG | USA | 0 | American |
| NA20412.SG | USA | 0 | American |
| NA20845.SG | USA | 0 | American |
| NA20846.SG | USA | 0 | American |
| NA20847.SG | USA | 0 | American |
| NA20849.SG | USA | 0 | American |
| NA20850.SG | USA | 0 | American |
| NA20851.SG | USA | 0 | American |
| NA20852.SG | USA | 0 | American |
| NA20853.SG | USA | 0 | American |
| NA20854.SG | USA | 0 | American |
| NA20856.SG | USA | 0 | American |
| NA20858.SG | USA | 0 | American |
| NA20859.SG | USA | 0 | American |
| NA20861.SG | USA | 0 | American |
| NA20862.SG | USA | 0 | American |
| NA20863.SG | USA | 0 | American |
| NA20864.SG | USA | 0 | American |
| NA20866.SG | USA | 0 | American |
| NA20867.SG | USA | 0 | American |
| NA20868.SG | USA | 0 | American |

| | | | |
|---|---|---|---|
| NA20869.SG | USA | 0 | American |
| NA20870.SG | USA | 0 | American |
| NA20871.SG | USA | 0 | American |
| NA20872.SG | USA | 0 | American |
| NA20874.SG | USA | 0 | American |
| NA20875.SG | USA | 0 | American |
| NA20876.SG | USA | 0 | American |
| NA20877.SG | USA | 0 | American |
| NA20878.SG | USA | 0 | American |
| NA20881.SG | USA | 0 | American |
| NA20882.SG | USA | 0 | American |
| NA20884.SG | USA | 0 | American |
| NA20885.SG | USA | 0 | American |
| NA20886.SG | USA | 0 | American |
| NA20887.SG | USA | 0 | American |
| NA20888.SG | USA | 0 | American |
| NA20889.SG | USA | 0 | American |
| NA20890.SG | USA | 0 | American |
| NA20891.SG | USA | 0 | American |
| NA20892.SG | USA | 0 | American |
| NA20893.SG | USA | 0 | American |
| NA20894.SG | USA | 0 | American |
| NA20895.SG | USA | 0 | American |
| NA20896.SG | USA | 0 | American |
| NA20897.SG | USA | 0 | American |
| NA20898.SG | USA | 0 | American |
| NA20899.SG | USA | 0 | American |
| NA20900.SG | USA | 0 | American |
| NA20901.SG | USA | 0 | American |

| | | | |
|---|---|---|---|
| NA20902.SG | USA | 0 | American |
| NA20903.SG | USA | 0 | American |
| NA20904.SG | USA | 0 | American |
| NA20905.SG | USA | 0 | American |
| NA20906.SG | USA | 0 | American |
| NA20908.SG | USA | 0 | American |
| NA20910.SG | USA | 0 | American |
| NA20911.SG | USA | 0 | American |
| NA21086.SG | USA | 0 | American |
| NA21087.SG | USA | 0 | American |
| NA21088.SG | USA | 0 | American |
| NA21089.SG | USA | 0 | American |
| NA21090.SG | USA | 0 | American |
| NA21091.SG | USA | 0 | American |
| NA21092.SG | USA | 0 | American |
| NA21093.SG | USA | 0 | American |
| NA21094.SG | USA | 0 | American |
| NA21095.SG | USA | 0 | American |
| NA21097.SG | USA | 0 | American |
| NA21098.SG | USA | 0 | American |
| NA21099.SG | USA | 0 | American |
| NA21100.SG | USA | 0 | American |
| NA21101.SG | USA | 0 | American |
| NA21102.SG | USA | 0 | American |
| NA21103.SG | USA | 0 | American |
| NA21104.SG | USA | 0 | American |
| NA21105.SG | USA | 0 | American |
| NA21106.SG | USA | 0 | American |
| NA21107.SG | USA | 0 | American |

| | | | |
|---|---|---|---|
| NA21108.SG | USA | 0 | American |
| NA21109.SG | USA | 0 | American |
| NA21110.SG | USA | 0 | American |
| NA21111.SG | USA | 0 | American |
| NA21112.SG | USA | 0 | American |
| NA21113.SG | USA | 0 | American |
| NA21114.SG | USA | 0 | American |
| NA21115.SG | USA | 0 | American |
| NA21116.SG | USA | 0 | American |
| NA21117.SG | USA | 0 | American |
| NA21118.SG | USA | 0 | American |
| NA21119.SG | USA | 0 | American |
| NA21120.SG | USA | 0 | American |
| NA21122.SG | USA | 0 | American |
| NA21123.SG | USA | 0 | American |
| NA21124.SG | USA | 0 | American |
| NA21125.SG | USA | 0 | American |
| NA21126.SG | USA | 0 | American |
| NA21127.SG | USA | 0 | American |
| NA21128.SG | USA | 0 | American |
| NA21129.SG | USA | 0 | American |
| NA21130.SG | USA | 0 | American |
| NA21133.SG | USA | 0 | American |
| NA21135.SG | USA | 0 | American |
| NA21137.SG | USA | 0 | American |
| NA21141.SG | USA | 0 | American |
| NA21142.SG | USA | 0 | American |
| NA21143.SG | USA | 0 | American |
| NA21144.SG | USA | 0 | American |

| | | | |
|---|---|---|---|
| I9058 | Brazil | 2260 | American |
| I9056_d | Brazil | 1175 | American |
| I9057_d | Brazil | 2220 | American |
| I9054_d | Brazil | 1935 | American |
| I9055_d | Brazil | 1870 | American |
| CP21_published | Brazil | 9250 | American |
| CP22_published | Brazil | 9580 | American |
| CP29 | Peru | 9005 | American |
| CP8 | Peru | 4115 | American |
| I5456 | Belize | 7375 | American |
| I5457 | Belize | 7376 | American |
| I1126_published | USA | 965 | American |
| I3443_published | Belize | 9301 | American |
| I2718 | USA | 1018 | American |
| I2715 | USA | 943 | American |
| I2719 | USA | 937 | American |
| I2716 | USA | 934 | American |
| I2717 | USA | 859 | American |
| I7088 | Argentina | 7428 | American |
| I7086 | Argentina | 7789 | American |
| I5319 | USA | 725 | American |
| I5320 | USA | 725 | American |
| I5321 | USA | 725 | American |
| I0324 | Peru | 1095 | American |
| I7090 | Argentina | 7162 | American |
| USR1.SG | USA | 11425 | American |
| USR2.SG | USA | 11425 | American |
| I10126 | Dominican Republic | 3022 | American |
| I7965 | Dominican Republic | 1230 | American |

| | | | |
|---|---|---|---|
| I7966 | Dominican Republic | 1249 | American |
| I7967 | Dominican Republic | 1228 | American |
| I7968 | Dominican Republic | 1015 | American |
| I7969 | Dominican Republic | 604 | American |
| I7970 | Dominican Republic | 600 | American |
| I7971 | Dominican Republic | 624 | American |
| I7973 | Dominican Republic | 849 | American |
| I7974 | Dominican Republic | 1002 | American |
| I7976 | Dominican Republic | 894 | American |
| I8118 | Dominican Republic | 641 | American |
| I8121_published | Dominican Republic | 1150 | American |
| I8122_published | Dominican Republic | 1150 | American |
| I8348 | Argentina | 6875 | American |
| I8349 | Argentina | 6715 | American |
| I8547 | Dominican Republic | 849 | American |
| I8548 | Dominican Republic | 1688 | American |
| I8549 | Dominican Republic | 688 | American |
| I8550 | Dominican Republic | 1272 | American |
| I8575 | Chile | 459 | American |
| I8576 | Chile | 100 | American |
| PC537.SG | Bahamas | 993 | American |
| HGDP00845.SDG | Brazil | 0 | American |
| HGDP00703.SDG | Colombia | 0 | American |
| HGDP01050.SDG | Mexico | 0 | American |
| HGDP01009.SDG | Brazil | 0 | American |
| HGDP01013.SDG | Brazil | 0 | American |
| HGDP00856.SDG | Mexico | 0 | American |
| HGDP00710.SDG | Colombia | 0 | American |
| HGDP00837.SDG | Brazil | 0 | American |

| | | | |
|---|---|---|---|
| HGDP01043.SDG | Mexico | 0 | American |
| HGDP00843.SDG | Brazil | 0 | American |
| HGDP00877.SDG | Mexico | 0 | American |
| HGDP01037.SDG | Mexico | 0 | American |
| HGDP00868.SDG | Mexico | 0 | American |
| HGDP00970.SDG | Colombia | 0 | American |
| HGDP00870.SDG | Mexico | 0 | American |
| HGDP00863.SDG | Mexico | 0 | American |
| HGDP00858.SDG | Mexico | 0 | American |
| HGDP01001.SDG | Brazil | 0 | American |
| HGDP00873.SDG | Mexico | 0 | American |
| HGDP01041.SDG | Mexico | 0 | American |
| HGDP00999.SDG | Brazil | 0 | American |
| HGDP00704.SDG | Colombia | 0 | American |
| HGDP00708.SDG | Colombia | 0 | American |
| HGDP00869.SDG | Mexico | 0 | American |
| HGDP00995.SDG | Brazil | 0 | American |
| HGDP00876.SDG | Mexico | 0 | American |
| HGDP00854.SDG | Mexico | 0 | American |
| HGDP00875.SDG | Mexico | 0 | American |
| HGDP01056.SDG | Mexico | 0 | American |
| HGDP01058.SDG | Mexico | 0 | American |
| HGDP00832.SDG | Brazil | 0 | American |
| HGDP01060.SDG | Mexico | 0 | American |
| HGDP00859.SDG | Mexico | 0 | American |
| HGDP01053.SDG | Mexico | 0 | American |
| HGDP00872.SDG | Mexico | 0 | American |
| HGDP01059.SDG | Mexico | 0 | American |
| HGDP00849.SDG | Brazil | 0 | American |

| | | | |
|---|---|---|---|
| HGDP00860.SDG | Mexico | 0 | American |
| HGDP00861.SDG | Mexico | 0 | American |
| HGDP01019.SDG | Brazil | 0 | American |
| HGDP01055.SDG | Mexico | 0 | American |
| HGDP01010.SDG | Brazil | 0 | American |
| HGDP01014.SDG | Brazil | 0 | American |
| HGDP00871.SDG | Mexico | 0 | American |
| HGDP00862.SDG | Mexico | 0 | American |
| HGDP00864.SDG | Mexico | 0 | American |
| HGDP00838.SDG | Brazil | 0 | American |
| HGDP01057.SDG | Mexico | 0 | American |
| HGDP00865.SDG | Mexico | 0 | American |
| HGDP00998.SDG | Brazil | 0 | American |
| HGDP00706.SDG | Colombia | 0 | American |
| HGDP01047.SDG | Mexico | 0 | American |
| HGDP00846.SDG | Brazil | 0 | American |
| HGDP01044.SDG | Mexico | 0 | American |
| HGDP01018.SDG | Brazil | 0 | American |
| HGDP00855.SDG | Mexico | 0 | American |
| HGDP01015.SDG | Brazil | 0 | American |
| HGDP00852.SDG | Brazil | 0 | American |
| HGDP00702.SDG | Colombia | 0 | American |
| HGDP01012.SDG | Brazil | 0 | American |
| HGDP00857.SDG | Mexico | 0 | American |
| I5319.DG | USA | 725 | American |
| I5319.SG | USA | 725 | American |
| I8351 | Argentina | 1630 | American |
| I10427_published | Canada | 1755 | American |
| CUN008 | Peru | 3316 | American |

| | | | |
|---|---|---|---|
| LAR001 | Brazil | 6555 | American |
| LAR002 | Brazil | 6790 | American |
| MOS001 | Brazil | 5809 | American |
| 523a.SG | USA | 100 | American |
| Ala1.SG | USA | 300 | American |
| B-03.SG | Mexico | 2700 | American |
| B-04.SG | Mexico | 2700 | American |
| CH-01.SG | USA | 1469 | American |
| CK-03.SG | Canada | 667 | American |
| CK-09.SG | Canada | 400 | American |
| CK-10.SG | Canada | 625 | American |
| CK-13.SG | Canada | 4842 | American |
| CR-01.SG | USA | 1017 | American |
| CT-01.SG | USA | 424 | American |
| CT-02.SG | USA | 400 | American |
| LU-01.SG | Canada | 625 | American |
| LU-02.SG | Canada | 625 | American |
| LU-03.SG | Canada | 625 | American |
| LU-05.SG | Canada | 625 | American |
| LU-06.SG | Canada | 625 | American |
| MX-01.SG | Mexico | 2700 | American |
| NC.SG | USA | 1336 | American |
| PS-02.SG | USA | 1550 | American |
| PS-03.SG | USA | 1456 | American |
| PS-04.SG | USA | 1550 | American |
| PS-06.SG | USA | 1454 | American |
| PS-07.SG | USA | 1447 | American |
| PS-09.SG | USA | 1550 | American |
| PS-17.SG | USA | 1550 | American |

| | | | |
|---|---|---|---|
| PS-18.SG | USA | 1550 | American |
| PS-23.SG | USA | 1550 | American |
| PS-26.SG | USA | 1550 | American |
| PS-34.SG | USA | 1550 | American |
| SC-01.SG | USA | 950 | American |
| SC-02.SG | USA | 950 | American |
| SC-03.SG | USA | 968 | American |
| SC-04.SG | USA | 799 | American |
| SC-05.SG | USA | 1011 | American |
| SC-06.SG | USA | 950 | American |
| SC-07.SG | USA | 950 | American |
| SM-01.SG | USA | 606 | American |
| SM-02.SG | USA | 722 | American |
| SN-01.SG | USA | 3150 | American |
| SN-03.SG | USA | 1588 | American |
| SN-04.SG | USA | 4842 | American |
| SN-09.SG | USA | 3150 | American |
| SN-10.SG | USA | 3150 | American |
| SN-11.SG | USA | 1087 | American |
| SN-12.SG | USA | 1209 | American |
| SN-13.SG | USA | 796 | American |
| SN-15.SG | USA | 1762 | American |
| SN-16.SG | USA | 3150 | American |
| SN-17.SG | USA | 5164 | American |
| SN-20.SG | USA | 4875 | American |
| SN-25.SG | USA | 4378 | American |
| SN-31.SG | USA | 4875 | American |
| SN-32.SG | USA | 5109 | American |
| SN-37.SG | USA | 4875 | American |

| | | | |
|---|---|---|---|
| SN-38.SG | USA | 3150 | American |
| SN-40.SG | USA | 4875 | American |
| SN-41.SG | USA | 4875 | American |
| SN-43.SG | USA | 2635 | American |
| SN-44.SG | USA | 5390 | American |
| SN-45.SG | USA | 4875 | American |
| SN-48.SG | USA | 4875 | American |
| SN-50.SG | USA | 1467 | American |
| SN-51.SG | USA | 1478 | American |
| SN-52.SG | USA | 919 | American |
| SN-53.SG | USA | 737 | American |
| SN-54.SG | USA | 4409 | American |
| SN-55.SG | USA | 4875 | American |
| SN-56.SG | USA | 4875 | American |
| SN-57.SG | USA | 4875 | American |
| SN-58.SG | USA | 4875 | American |
| SN-59.SG | USA | 5053 | American |
| SN-60.SG | USA | 4409 | American |
| US-14.SG | USA | 334 | American |
| IPK12.SG | Chile | 858 | American |
| IPK13a.SG | Chile | 1211 | American |
| IPY08b.SG | Chile | 1000 | American |
| I11974_published | Chile | 11885 | American |
| I12364 | Argentina | 770 | American |
| I12366 | Argentina | 100 | American |
| I12357 | Argentina | 330 | American |
| I12365 | Argentina | 740 | American |
| I12359 | Argentina | 590 | American |
| I12361 | Argentina | 420 | American |

| | | | |
|---|---|---|---|
| I12355 | Argentina | 1490 | American |
| I12376 | Argentina | 2370 | American |
| I12360 | Argentina | 400 | American |
| I12363 | Argentina | 380 | American |
| I12367 | Argentina | 700 | American |
| I12941 | Argentina | 160 | American |
| I12943 | Argentina | 180 | American |
| I12942 | Argentina | 60 | American |
| I13208 | Dominican Republic | 660 | American |
| I13190 | Dominican Republic | 711 | American |
| I13195 | Dominican Republic | 601 | American |
| I13203 | Dominican Republic | 750 | American |
| I13201 | Dominican Republic | 598 | American |
| I13196 | Dominican Republic | 679 | American |
| I13189 | Dominican Republic | 753 | American |
| I13199 | Dominican Republic | 534 | American |
| I13192 | Dominican Republic | 688 | American |
| I13206 | Dominican Republic | 516 | American |
| I13204 | Dominican Republic | 525 | American |
| I13207 | Dominican Republic | 525 | American |
| I13198 | Dominican Republic | 483 | American |
| I13559_published | Bahamas | 750 | American |
| I13560_published | Bahamas | 663 | American |
| I13558 | Bahamas | 599 | American |
| I12356 | Argentina | 280 | American |
| I12576 | Haiti | 950 | American |
| Anzick_realigned.SG | USA | 12712 | American |
| I12341 | Dominican Republic | 1150 | American |
| I12344 | Dominican Republic | 1150 | American |

| | | | |
|---|---|---|---|
| I12347 | Dominican Republic | 1150 | American |
| I12350 | Dominican Republic | 1150 | American |
| I13738_published | Bahamas | 611 | American |
| I13441 | Puerto Rico | 950 | American |
| I13737 | Bahamas | 752 | American |
| I13739_published | Bahamas | 614 | American |
| I13323 | Puerto Rico | 701 | American |
| I13539 | Puerto Rico | 1315 | American |
| I13538 | Puerto Rico | 850 | American |
| I13321 | Bahamas | 750 | American |
| I13540 | Puerto Rico | 1014 | American |
| I13322 | Bahamas | 1017 | American |
| I13320 | Bahamas | 850 | American |
| I13541 | Puerto Rico | 1000 | American |
| I13318 | Bahamas | 750 | American |
| I13319 | Bahamas | 750 | American |
| I13326 | Puerto Rico | 1000 | American |
| I13324 | Puerto Rico | 850 | American |
| IL2.SG | Peru | 1625 | American |
| IL3.SG | Peru | 1697 | American |
| IL4.SG | Peru | 1475 | American |
| IL5.SG | Peru | 1658 | American |
| IL7.SG | Peru | 1778 | American |
| K1.SG | Peru | 3950 | American |
| SMP5.SG | Peru | 6831 | American |
| 19651.SG | Canada | 5666 | American |
| SA5832.SG | Chile | 7293 | American |
| A460.SG | Chile | 5130 | American |
| Aconcagua.SG | Argentina | 500 | American |

| | | | |
|---|---|---|---|
| AHUR_2064.SG | USA | 10962 | American |
| AHUR770c.SG | USA | 10988 | American |
| Lovelock1.SG | USA | 1935 | American |
| Lovelock2.SG | USA | 1839 | American |
| Lovelock3.SG | USA | 648 | American |
| Lovelock4.SG | USA | 1743 | American |
| Sumidouro4.SG | Brazil | 10174 | American |
| Sumidouro5.SG | Brazil | 10403 | American |
| Sumidouro6.SG | Brazil | 10124 | American |
| Sumidouro7.SG | Brazil | 10232 | American |
| Sumidouro8.SG | Brazil | 9816 | American |
| TrailCreek.SG | USA | 9005 | American |
| I12575 | Haiti | 950 | American |
| I14922 | Bahamas | 1003 | American |
| I14921 | Bahamas | 679 | American |
| I14920 | Bahamas | 938 | American |
| I14923 | Bahamas | 603 | American |
| I14875 | Bahamas | 1073 | American |
| I14879 | Bahamas | 651 | American |
| I14880 | Bahamas | 589 | American |
| I14878 | Bahamas | 795 | American |
| I15587 | Dominican Republic | 825 | American |
| I14881_published | Bahamas | 597 | American |
| I14876_published | Bahamas | 624 | American |
| I14882 | Bahamas | 635 | American |
| I14883 | Bahamas | 588 | American |
| I15682 | Dominican Republic | 1125 | American |
| I14009 | Chile | 750 | American |
| I16178_published | Dominican Republic | 702 | American |

| | | | |
|---|---|---|---|
| I16173 | Dominican Republic | 700 | American |
| I16175 | Dominican Republic | 675 | American |
| I16176 | Dominican Republic | 694 | American |
| I16181_published | Dominican Republic | 675 | American |
| I16172 | Dominican Republic | 875 | American |
| I16174 | Dominican Republic | 894 | American |
| I16180 | Dominican Republic | 862 | American |
| I16171 | Dominican Republic | 1125 | American |
| I15109 | Dominican Republic | 510 | American |
| I15051 | Dominican Republic | 852 | American |
| I15108 | Dominican Republic | 512 | American |
| I15112 | Dominican Republic | 702 | American |
| I15106 | Dominican Republic | 702 | American |
| I15082 | Dominican Republic | 673 | American |
| I15111 | Dominican Republic | 607 | American |
| I15107 | Dominican Republic | 604 | American |
| I15081 | Dominican Republic | 859 | American |
| I15105 | Dominican Republic | 706 | American |
| I14990 | Dominican Republic | 697 | American |
| I15050 | Dominican Republic | 849 | American |
| I15667 | Dominican Republic | 516 | American |
| I15674 | Dominican Republic | 625 | American |
| I15594 | Dominican Republic | 875 | American |
| I15597 | Dominican Republic | 1175 | American |
| I15596 | Dominican Republic | 1125 | American |
| I15600_published | Dominican Republic | 1217 | American |
| I15679 | Dominican Republic | 974 | American |
| I15599_published | Dominican Republic | 1125 | American |
| I15666 | Dominican Republic | 825 | American |

| | | | |
|---|---|---|---|
| I15593 | Dominican Republic | 974 | American |
| I15590 | Dominican Republic | 875 | American |
| I15678 | Dominican Republic | 970 | American |
| I15591 | Dominican Republic | 615 | American |
| I15676 | Dominican Republic | 464 | American |
| I15668 | Dominican Republic | 597 | American |
| I15677 | Dominican Republic | 855 | American |
| I15602 | Dominican Republic | 1175 | American |
| I15675 | Dominican Republic | 1125 | American |
| I15665 | Dominican Republic | 723 | American |
| I15671_published | Dominican Republic | 1125 | American |
| I15601 | Dominican Republic | 1228 | American |
| I15604 | Dominican Republic | 875 | American |
| I15592 | Dominican Republic | 859 | American |
| I15598 | Dominican Republic | 875 | American |
| I15595 | Dominican Republic | 1286 | American |
| I15971 | Dominican Republic | 679 | American |
| I15976 | Dominican Republic | 1112 | American |
| I15964 | Dominican Republic | 1125 | American |
| I15977 | Dominican Republic | 1002 | American |
| I15965 | Dominican Republic | 1125 | American |
| I15968 | Dominican Republic | 930 | American |
| I15962 | Dominican Republic | 517 | American |
| I15972 | Dominican Republic | 536 | American |
| I15970 | Dominican Republic | 875 | American |
| I15969 | Dominican Republic | 995 | American |
| I15966 | Dominican Republic | 768 | American |
| I15978 | Dominican Republic | 959 | American |
| I15975 | Dominican Republic | 850 | American |

| | | | |
|---|---|---|---|
| I15963 | Dominican Republic | 1218 | American |
| I11974.SG | Chile | 11885 | American |
| I14991 | Dominican Republic | 614 | American |
| I14994 | Dominican Republic | 703 | American |
| I16687 | Dominican Republic | 935 | American |
| IO2 | Bolivia | 985 | American |
| MIS3 | Bolivia | 1120 | American |
| MIS5 | Bolivia | 1090 | American |
| MIS7 | Bolivia | 1100 | American |
| shukaKaa.SG | USA | 10366 | American |
| I13736 | Bahamas | 750 | American |
| I15672 | Dominican Republic | 867 | American |
| I15973 | Dominican Republic | 635 | American |
| I16540 | Dominican Republic | 875 | American |
| I16556 | Dominican Republic | 1125 | American |
| I18437 | Dominican Republic | 956 | American |
| I10333 | Dominican Republic | 2500 | American |
| I12362 | Argentina | 5820 | American |
| I17889 | Venezuela | 2345 | American |
| I17891 | Venezuela | 2230 | American |
| I17905_published | Dominican Republic | 604 | American |
| I17896 | Venezuela | 2250 | American |
| I17906 | Dominican Republic | 1130 | American |
| I17899 | Dominican Republic | 471 | American |
| I17890 | Venezuela | 2235 | American |
| I17894 | Venezuela | 2263 | American |
| I17908 | Dominican Republic | 524 | American |
| I17900 | Dominican Republic | 529 | American |
| I17902 | Dominican Republic | 661 | American |

| | | | |
|---|---|---|---|
| I17893 | Venezuela | 2226 | American |
| I17907 | Dominican Republic | 587 | American |
| I17903 | Dominican Republic | 1175 | American |
| I17892 | Venezuela | 2337 | American |
| I17895 | Venezuela | 2250 | American |
| I17901 | Dominican Republic | 517 | American |
| I18134 | Dominican Republic | 1119 | American |
| I18301 | Dominican Republic | 848 | American |
| I17497 | Chile | 603 | American |
| I18300 | Dominican Republic | 800 | American |
| I7972_published | Dominican Republic | 925 | American |
| I14992 | Dominican Republic | 543 | American |
| I12354 | Argentina | 450 | American |
| I16539 | Dominican Republic | 858 | American |
| I12358 | Argentina | 440 | American |
| I16519 | Dominican Republic | 930 | American |
| I16520 | Dominican Republic | 875 | American |
| I2538 | Chile | 1084 | American |
| I8350_published | Argentina | 7700 | American |
| IPY10.SG | Chile | 775 | American |
| UC8-8168 | Peru | 500 | American |
| UC8-8173 | Peru | 500 | American |
| UC12-12 | Peru | 500 | American |
| UC12-20 | Peru | 500 | American |
| UC12-24 | Peru | 500 | American |
| UC12-25 | Peru | 500 | American |
| I20527 | Dominican Republic | 998 | American |
| PI-420A.WGC | Puerto Rico | 672 | American |
| PI-51.WGC | Puerto Rico | 862 | American |

| | | | |
|---|---|---|---|
| CAA002 | Cuba | 1561 | American |
| ALG001 | Guadeloupe | 935 | American |
| ALG002 | Guadeloupe | 1024 | American |
| ALG003 | Guadeloupe | 637 | American |
| ALG004 | Guadeloupe | 663 | American |
| CAA001003 | Cuba | 1736 | American |
| CAO001 | Cuba | 849 | American |
| CAO002008 | Cuba | 1326 | American |
| CAO004 | Cuba | 1214 | American |
| CAO006 | Cuba | 2000 | American |
| CAO007 | Cuba | 1104 | American |
| CAO009013 | Cuba | 1329 | American |
| CAO010 | Cuba | 2000 | American |
| CAO011 | Cuba | 905 | American |
| CAO012 | Cuba | 1328 | American |
| CAO014 | Cuba | 1542 | American |
| CAO015 | Cuba | 3131 | American |
| CAO016 | Cuba | 2000 | American |
| CAO017 | Cuba | 2000 | American |
| CAO018 | Cuba | 3150 | American |
| CAO019 | Cuba | 3188 | American |
| CAO020 | Cuba | 3112 | American |
| CAO021 | Cuba | 750 | American |
| CAO022026 | Cuba | 1313 | American |
| CAO023025 | Cuba | 932 | American |
| CAO024 | Cuba | 1319 | American |
| CAO027 | Cuba | 1299 | American |
| CAO028 | Cuba | 709 | American |
| CAO029 | Cuba | 1323 | American |

| | | | |
|---|---|---|---|
| CAO030 | Cuba | 1300 | American |
| CAO031 | Cuba | 1320 | American |
| CAO032 | Cuba | 1214 | American |
| CDE001 | Cuba | 393 | American |
| CDE002 | Cuba | 488 | American |
| CDE003 | Cuba | 515 | American |
| CDE004 | Cuba | 517 | American |
| CDE005 | Cuba | 425 | American |
| CIP001 | Cuba | 1455 | American |
| CIP002 | Cuba | 1414 | American |
| CIP003 | Cuba | 1950 | American |
| CIP007012 | Cuba | 1345 | American |
| CIP008 | Cuba | 1232 | American |
| CIP009 | Cuba | 2734 | American |
| CIP010 | Cuba | 1397 | American |
| CIP011 | Cuba | 1257 | American |
| CUC001 | Cuba | 2000 | American |
| CUC002 | Cuba | 2000 | American |
| CUC003 | Cuba | 2000 | American |
| CUC004 | Cuba | 2000 | American |
| ELM001 | Cuba | 530 | American |
| GUY001 | Cuba | 2150 | American |
| GUY002 | Cuba | 2510 | American |
| GUY003 | Cuba | 1657 | American |
| LAV001 | St. Lucia | 830 | American |
| LAV002 | St. Lucia | 685 | American |
| LAV003 | St. Lucia | 672 | American |
| LAV004 | St. Lucia | 725 | American |
| LAV005 | St. Lucia | 855 | American |

| | | | |
|---|---|---|---|
| LAV006 | St. Lucia | 725 | American |
| LAV007 | St. Lucia | 659 | American |
| LAV008 | St. Lucia | 675 | American |
| LAV009 | St. Lucia | 646 | American |
| LAV010 | St. Lucia | 659 | American |
| LAV011 | St. Lucia | 698 | American |
| LAV012 | St. Lucia | 602 | American |
| LOI001 | Puerto Rico | 612 | American |
| MTO001 | Cuba | 1231 | American |
| PCA001 | Puerto Rico | 1367 | American |
| PCA002 | Puerto Rico | 1175 | American |
| PCA006 | Puerto Rico | 1026 | American |
| PCA009 | Puerto Rico | 1132 | American |
| PCA010 | Puerto Rico | 1175 | American |
| PCV001 | Bahamas | 875 | American |
| PCV002 | Bahamas | 795 | American |
| PCV003 | Bahamas | 883 | American |
| PCV004 | Bahamas | 875 | American |
| PDI003 | Puerto Rico | 925 | American |
| PDI006 | Puerto Rico | 1081 | American |
| PDI008 | Puerto Rico | 672 | American |
| PDI009 | Puerto Rico | 925 | American |
| PDI010 | Puerto Rico | 862 | American |
| PDI011 | Puerto Rico | 801 | American |
| PDI012013 | Puerto Rico | 900 | American |
| PDM001 | Cuba | 1826 | American |
| PDM002 | Cuba | 1900 | American |
| PDM003 | Cuba | 1960 | American |
| PDM004 | Cuba | 1824 | American |

| | | | |
|---|---|---|---|
| PDM006 | Cuba | 1992 | American |
| PDM008 | Cuba | 1783 | American |
| PDM009 | Cuba | 1900 | American |
| PDM010 | Cuba | 1877 | American |
| TIB001 | Puerto Rico | 1333 | American |
| TIB002 | Puerto Rico | 1068 | American |
| I10334 | Dominican Republic | 2500 | American |
| I7977 | Dominican Republic | 1887 | American |
| UC12_12.SG | Peru | 598 | American |
| UC12_20.SG | Peru | 575 | American |
| UC12_24.SG | Peru | 506 | American |
| UC12_25.SG | Peru | 575 | American |
| UC8_8168.SG | Peru | 467 | American |
| UC8_8173.SG | Peru | 533 | American |
| PAPV_114.SG | Panama | 1316 | American |
| PAPV_117.SG | Panama | 944 | American |
| PAPV_118.SG | Panama | 610 | American |
| PAPV_137.SG | Panama | 990 | American |
| PAPV_146.SG | Panama | 1061 | American |
| PAPV_167.SG | Panama | 1023 | American |
| PAPV_172.SG | Panama | 763 | American |
| PAPV_173.SG | Panama | 547 | American |
| PAPV_174.SG | Panama | 603 | American |
| PAPV_175.SG | Panama | 1237 | American |
| PAPV_26.SG | Panama | 425 | American |
| PAPV_27.SG | Panama | 425 | American |
| PAPV_52.SG | Panama | 300 | American |
| PAPV_53.SG | Panama | 300 | American |
| PAPV_61.SG | Panama | 300 | American |

| | | | |
|---|---|---|---|
| PAPV_93.SG | Panama | 300 | American |
| I14879.v50.0.SG | Bahamas | 651 | American |
| I14880.v50.0.SG | Bahamas | 589 | American |
| I14882.v50.0.SG | Bahamas | 635 | American |
| I14883.v50.0.SG | Bahamas | 588 | American |
| I15595.v50.0.SG | Dominican Republic | 1286 | American |
| I15667.v50.0.SG | Dominican Republic | 516 | American |
| I15962.v50.0.SG | Dominican Republic | 517 | American |
| I15968.v50.0.SG | Dominican Republic | 930 | American |
| I20527.v50.0.SG | Dominican Republic | 998 | American |
| I11974.v50.0.SG | Chile | 11885 | American |
| I14879.v50.0_d.SG | Bahamas | 651 | American |
| I14880.v50.0_d.SG | Bahamas | 589 | American |
| I14882.v50.0_d.SG | Bahamas | 635 | American |
| I14883.v50.0_d.SG | Bahamas | 588 | American |
| I15595.v50.0_d.SG | Dominican Republic | 1286 | American |
| I15667.v50.0_d.SG | Dominican Republic | 516 | American |
| I15962.v50.0_d.SG | Dominican Republic | 517 | American |
| I15968.v50.0_d.SG | Dominican Republic | 930 | American |
| I20527.v50.0_d.SG | Dominican Republic | 998 | American |
| I11974.v50.0_d.SG | Chile | 11885 | American |
